# Automated Disease Activity Assessment in Systemic Lupus Erythematosus Using Privacy-Preserving Large Language Models

**DOI:** 10.64898/2026.07.09.26357586

**Authors:** Danting Zhang, Renee L. Leung, Chun-Ka Wong, Shirley Chiu Wai Chan, Yi Li, Eric H. M. Tang, Tingting Wu, Tak Mao Chan, Chak-Sing Lau, Carlos King Ho Wong, Kathy Sze Man Leung, Zoie Shui-Yee Wong, Joseph Tsz-Kei Wu, Desmond Yat-Hin Yap

## Abstract

The Systemic Lupus Erythematosus Disease Activity Index 2000 (SLEDAI-2K) is a crucial but labor-intensive tool for managing SLE. We developed a privacy-preserving, model-agnostic large language model (LLM) framework to automate SLEDAI-2K assessment from real-world electronic health records. The framework was developed on a specialist-verified ground truth of 658 clinical notes and externally validated on 56 MIMIC-IV discharge summaries. Seven open-source LLMs were evaluated using advanced prompting and ensemble strategies. The top-performing model, a two-layered GPT-OSS-120B + verifier, achieved a micro-F1 of 94.2% for descriptor classification and an 86% exact match for SLEDAI-2K scores on the internal set, with corresponding external validation performance of 87.7% and 64%, respectively. To demonstrate clinical utility, the LLMs were deployed on 2,576 serial notes from 108 SLE patients. Patients identified by the LLMs as achieving sustained low disease activity had a significantly lower incidence of stage 3 chronic kidney disease (log-rank p = 0.0053), the need for kidney replacement therapy (p = 0.044), and hospitalization (p = 0.021) over 18.3 years of follow-up. These findings demonstrate that privacy-preserving LLMs, when guided by a well-designed framework, can assist in specialist-level reasoning in autoimmune diseases, offering a scalable solution for clinical decision support and patient management.

## Introduction

Systemic lupus erythematosus (SLE) is a complex autoimmune disease characterized by multi-systemic involvement. Recurrent flares and accumulated organ damage translate into patient mortality, renal failure, impaired quality of life, as well as increased socio-economic and healthcare burdens ^1–3^.

Monitoring disease activity is crucial in SLE management, involving cross-sectional assessment at individual time points and longitudinal monitoring over time. The SLE Activity Index (SLEDAI) and its refined versions, including SELENA-SLEDAI and SLEDAI-2K ^4^, have been widely accepted as disease activity scoring systems used for clinical trial recruitment, outcome assessment, and defining treatment targets ^5–7^. Accurate SLEDAI assessment requires evaluation by highly skilled specialists, which demands good clinical reasoning and considers the multisystem differential diagnosis, timing of disease manifestations, paraclinical test results, as well as treatment actions and drug-related effects. Given this labor-intensive process and the worldwide scarcity of rheumatologists ^8^, the SLEDAI is often not properly documented in routine clinical practice and research settings.

With growing evidence of their diagnostic capacities, large language models (LLMs) represent a promising solution for automating SLEDAI assessment. Early LLM advances in clinical NLP were driven by BERT (Bidirectional Encoder Representations from Transformers) ^9,10^ and its domain-specific variant ClinicalBERT ^11^, which established state-of-the-art performance in clinical named entity recognition (NER). The emergence of generative LLMs (e.g., GPT-3.5) marked a major shift in the field, enabling more complex clinical decision-support tasks that leverage advanced reasoning ^12,13^, extended context windows ^14^, and fine-tuning ^15,16^. So far, the mainstream application of LLMs in the medical field has relied on proprietary models like ChatGPT and GPT-4 ^17^. However, stringent healthcare data regulations, which mandate on-premises storage and strict access controls, limit the application of proprietary LLM in core clinical tasks involving sensitive data, such as electronic health records (EHRs). Therefore, the medical community is advocating for open-source LLMs that prioritize data security, privacy, and transparency, allowing for local deployment ^18,19^. Current adoption of LLMs in rheumatic diseases is predominantly confined to simple information extraction tasks and to diseases primarily characterized by articular involvement, such as rheumatoid arthritis, gout, and axial spondyloarthritis ^20,21^. Due to the complex disease manifestations and differential diagnosis, the application of LLMs in SLE remains under-investigated. Prior work has focused on automating SLE information extraction and basic classification tasks from EHRs using traditional NLP, including Bag-of-Words ^22^ and MetaMap ^23^. Research on proprietary generative LLMs (e.g., GPT-4, Claude, Gemini) has primarily assessed their medical education capabilities ^24–26^.

Studies extending LLMs’ application to clinical practice have yielded fair performance in automating SLE diagnosis and cutaneous scores, but the cloud-based nature of some models introduces data security concerns ^27,28^. To date, no published work has used LLMs to automate SLEDAI scoring.

Acknowledging the huge potential of LLMs in SLE management, this study set out to establish a new model-agnostic workflow in automating SLEDAI-2K scoring from real-world narrative EHR clinical notes using privacy-preserving LLMs. We also simulated a clinical implementation to demonstrate the clinical and research utility of our proposed framework in SLE and lupus nephritis (LN).

## Results

### Characteristics of patients and clinical notes

The characteristics of clinical notes are presented in Table 1. The average note length was 71 words for the real-world outpatient notes, 135 for pseudo notes, and 570 for MIMIC-IV notes. The dataset presents a right-skewed distribution of SLEDAI-2K, with mean scores of 3.8 for outpatient notes, 9.3 for pseudo notes, and 6.0 for MIMIC-IV notes. The most prevalent positive descriptor in the internal dataset was low complement (26.6%), followed by increased anti-dsDNA binding (19.0%) and proteinuria (12.2%) (***Figures S1 & S2, Tables S1-S4***).

**Table 1.** Characteristics of the patients and clinical notes from the internal comprehensive SLE clinical dataset (training and testing) and the external MIMIC-IV validation set.

| Characteristics | The comprehensive SLE clinical dataset | Training set |  |  | Testing set |  |  | Validation set (MIMIC-IV) |
| --- | --- | --- | --- | --- | --- | --- | --- | --- |
| Patients | Total (n=250) | Total (n=125) | Real (n=55) | Pseudo (n=70) | Total (n=125) | Real (n=51) | Pseudo (n=74) | Total (n=51) |
| Female, n (%) | 217 (86.8) | 112 (89.0) | 50 (90.0) | 62 (88.2) | 105 (84.8) | 43 (86.0) | 62 (84.0) | 50 (98.0) |
| Notes | Total (n=658) | Total (n=300) | Real (n=221) | Pseudo (n=79) | Total (n=358) | Real (n=268) | Pseudo (n=90) | Total (n=56) |
| Age at visit, mean (SD) years | 50.81 (13.07) | 51.07 (12.74) | 53.62 (8.77) | 43.94 (18.23) | 50.59 (13.33) | 53.53 (9.91) | 41.84 (17.67) | 35.02 (11.61) |
| Female, n (%) | 583 (88.6) | 273 (90.1) | 202 (90.1) | 71 (90.0) | 310 (86.6) | 232 (86.7) | 78 (86.4) | 55 (98.2) |
| SLEDAI-2K score, mean (SD) | 3.83 (5.73) | 3.80 (5.86) | 1.77 (2.29) | 9.47 (8.50) | 3.79 (5.60) | 1.96 (2.55) | 9.24 (8.10) | 5.88 (5.28) |
| SLEDAI-2K score categories, n (%) |  |  |  |  |  |  |  |  |
| 0 | 271 (41.2) | 127 (42.3) | 111 (50.2) | 16 (20.3) | 144 (40.2) | 129 (48.1) | 15 (16.7) | 11 (19.6) |
| 1-5 | 246 (37.4) | 110 (36.7) | 93 (42.1) | 17 (21.5) | 136 (38.0) | 114 (42.5) | 22 (24.4) | 17 (30.4) |
| 6-10 | 67 (10.2) | 29 (9.7) | 16 (7.2) | 13 (16.5) | 38 (10.6) | 22 (8.2) | 16 (17.8) | 18 (32.1) |
| 11-19 | 58 (8.8) | 26 (8.7) | 1 (0.5) | 25 (31.6) | 32 (8.9) | 3 (1.1) | 29 (32.2) | 9 (16.1) |
| >=20 | 16 (2.4) | 8 (2.7) | 0 (0.0) | 8 (10.1) | 8 (2.2) | 0 (0.0) | 8 (8.9) | 1 (1.8) |
| Notes length, mean (SD) words | 88.49 (58.46) | 90.51 (61.42) | 73.28 (49.82) | 138.71 (65.03) | 86.80 (55.80) | 70.71 (43.99) | 134.70 (59.65) | 569.74 (402.91) |

SLE, systemic lupus erythematosus; SLEDAI-2K, Systemic Lupus Erythematosus Disease Activity Index 2000; SD, standard deviation

### Performance of LLMs with different ensemble strategies

***Figure 2B*** summarizes the performances of LLMs against ground-truth on the testing and validation sets. Comparisons between the individual and grouped prompting revealed that individual-descriptor prompts led to better performance for both Qwen3-32B and GPT-OSS-120B on all metrics. Therefore, we selected this prompt strategy for further between-model comparisons.

**Figure 1.**
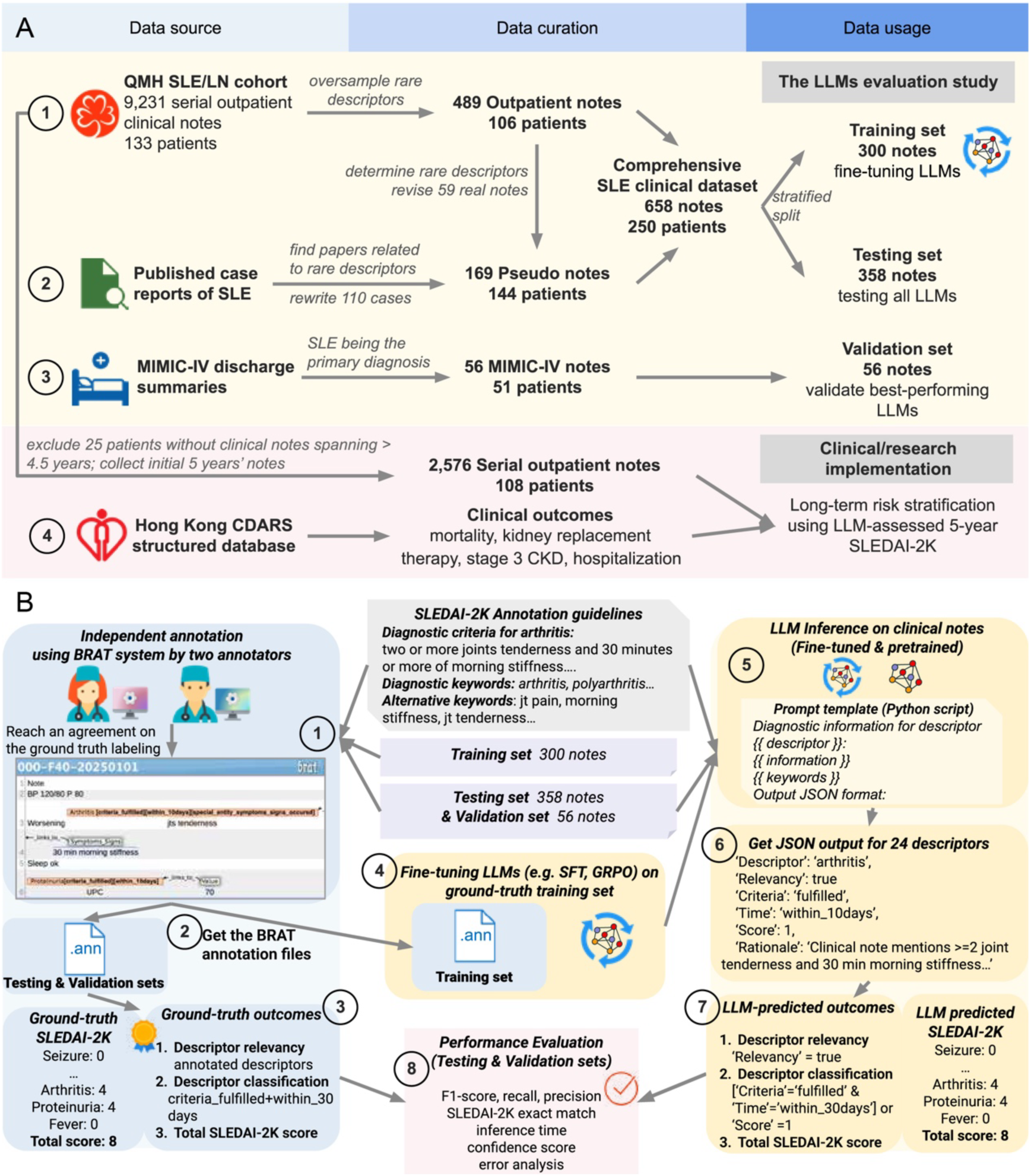
(A) Overview of the data used for LLM evaluation and clinical/research implementation study. (B) The named entity framework (annotation guidelines, ground-truth labeling, prompt engineering), fine-tuning strategies, and evaluation metrics of LLMs. Blue boxes indicate human-driven steps; yellow boxes indicate machine-driven steps. Graphical elements were sourced from Icons8 (https://icons8.com/), used under the free license terms.

**Figure 2.**
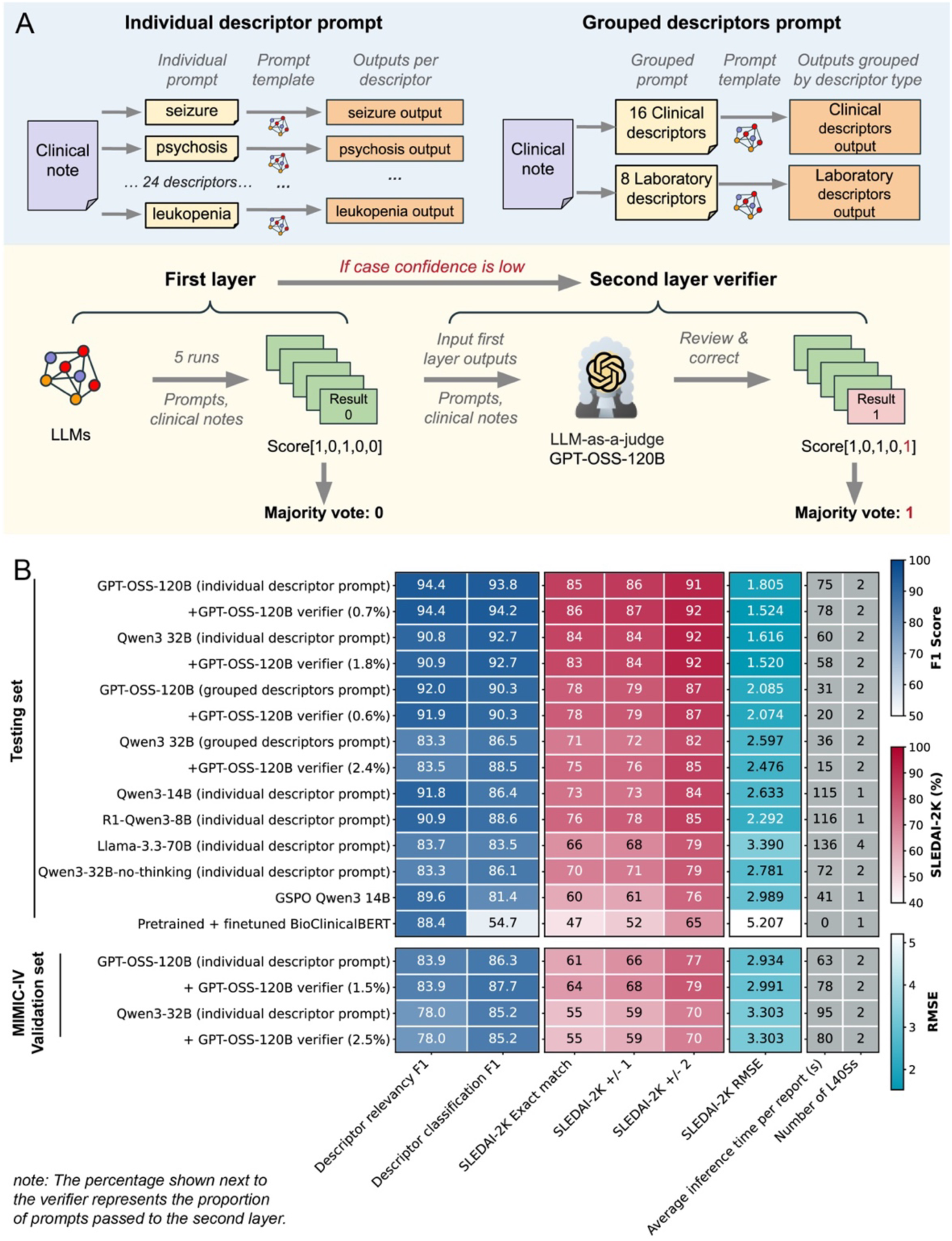
(A) Illustration of prompting strategies, majority vote, and second-layer LLM-as-a-judge. (B) Performance of different LLMs and ensemble strategies on the testing set (358 cases) and validation set (56 MIMIC-IV cases). Descriptor relevancy F1 measures whether the model correctly identifies which entities are relevant or irrelevant within the input clinical note. Descriptor classification F1 measures whether the model assigns the correct binary score to each entity. The percentage shown next to the verifier represents the proportion of prompts passed to the second layer. Color scales across all figures are standardized to the same range. The reported inference time is for a single run. Graphical elements were sourced from Icons8 (https://icons8.com/), used under the free license terms.

A two-layered GPT-OSS-120B + verifier emerged as the best model on both sets. On the testing set, performance was as follows: a micro-F1 of 94.4% for descriptor relevancy, 94.2% for descriptor classification, an 86% exact match for SLEDAI-2K scores, and 91% within a ±2-point tolerance. The corresponding performance in the validation set was 83.9%, 87.7%, 64% and 77%, respectively (***Figure 2C***). The performance was closely followed by one-layer GPT-OSS-120B, Qwen3-32B, and its two-layer variants.

The deterministic configuration, while offering better reproducibility, underperformed the non-deterministic one (***Figure S3***). Larger models (Qwen3-32B) within the same family outperformed the smaller ones (Qwen3-14B and Deepseek-R1-Qwen3-8B). Performance of non-reasoning models (Qwen3-32B no-thinking and Llama3.3-70B) is weaker than that of reasoning models. Similar findings are observed in the full dataset for non-finetuned LLMs (***Figure S4***). Reinforcement learning finetuning with GSPO did not improve the overall performance of Qwen3-14B. With drastically faster inference, the finetuned BioClinicalBERT achieved an 88.4% F1-score in descriptor relevancy detection, even outperforming the non-reasoning models, but the F1-score declined sharply in descriptor classification.

### Overall and by-descriptor performance of the best model

On testing and validation sets, most predictions by the best-performing two-layered GPT-OSS-120B + verifier cluster tightly around the ideal prediction line across a wide score range, with few outliers near the ±8 threshold (***Figures 3A & 3B***). Generally, most yellowish points with higher confidence (∼9.5) align closely with the ideal line, while bluish points (∼7) are more dispersed. The best model’s AUCs for predicting four binary SLEDAI-2K classifications were >95% on the internal set and >90% on the external set, with consistently high sensitivity and specificity (***Figures 3A & 3B, Table S5***). By-descriptor analysis on the testing set showed that the GPT-OSS-120B + verifier yielded varied performance across descriptors (***Figure 3C, Figure S5***). The model performance excelled in scoring Laboratory descriptors (micro-F1: 96.6%), declined for non-8-point Clinical descriptors (90%), and was even lower for 8-point Clinical descriptors (88.2%). Specifically, the model achieved 100% F1-scores for seizure, CVA, urinary casts, and thrombocytopenia, and >97% F1s for hematuria, alopecia, low complement, and leukopenia. In contrast, the F1-score was the weakest for pericarditis (60%), which was attributed to low recall (higher false negatives). On the validation set, the model continued to excel with Laboratory descriptors (micro-F1: 94.9%) but performed poorly for 8-point Clinical descriptors (40%) (***Figure 3D***). Similar performance of the two-layered GPT-OSS-120B + verifier is observed in the full dataset (***Figures S6 & S7***).

**Figure 3.**
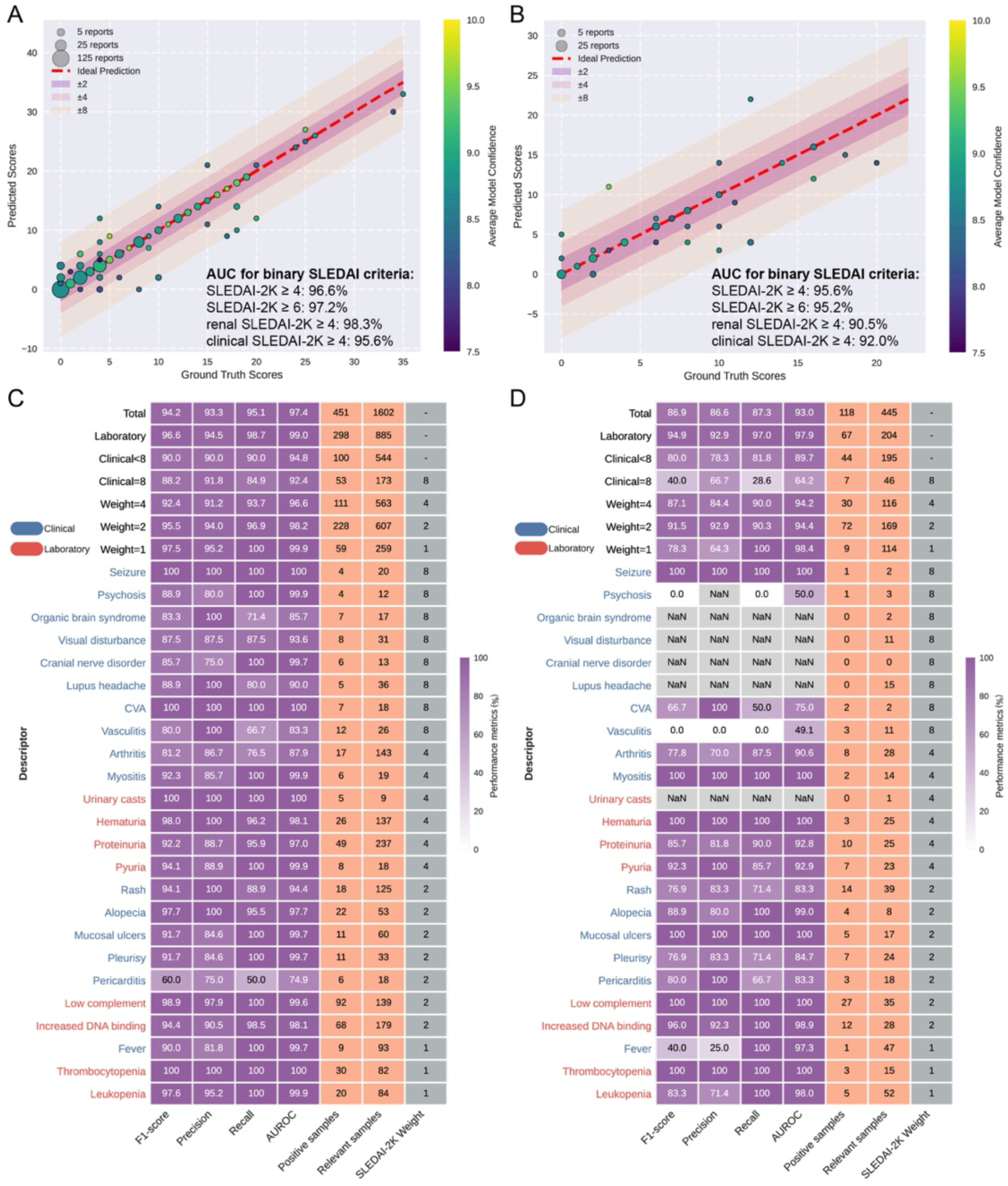
Performance of the best LLM model (GPT-OSS-120B + verifier) showing predicted versus ground truth SLEDAI-2K scores on the (A) testing set (358 cases) and (B) validation set (56 MIMIC-IV cases), with AUC values for predicting binary SLEDAI-2K criteria. By-descriptor performance of GPT-OSS-120B + verifier on the (C) testing set (358 cases) and (D) validation set (56 cases). Scatter plots illustrate the agreement between model-predicted scores and the actual ground truth scores. Each bubble denotes predictions, with the bubble size proportional to the number of overlapping cases it represents. The dashed red line denotes the line of ideal prediction, where the prediction equals the ground truth. The shades indicate deviation intervals of ±2, ±4, and ±8 points from the ideal. The color of each bubble represents the model’s average case confidence for that prediction, ranging from around 7.5 (purple) to 10 (yellow). Clinical SLEDAI-2K excludes increased DNA binding and low complement. Renal SLEDAI-2K includes four renal descriptors: hematuria, pyuria, proteinuria, and urinary casts.

Other LLMs showed similar trends, with performance of Laboratory descriptors consistently achieving over 90% for reasoning models (over 80% for non-reasoning), non-8-point Clinical over 80% (except Qwen3-14B), and 8-point Clinical over 70% (***Figure S8***). Those challenging Clinical descriptors, which had lower F1 scores, also required a greater average human annotation time (***Figure S9***). GSPO-based reinforcement learning fine-tuning improved Qwen-3-14B’s performance across 14 predicted descriptors, including 12 clinical descriptors (***Figure S10***). ***Table S6*** demonstrates the reasoning trace where the inference of GPT-OSS-120B led to correct predictions for all descriptors.

### Error analysis of the reasoning processes

We pooled error records from the three sets to compare the reasoning flaws of the first-layer GPT-OSS-120B (143 records) and Qwen3-32B (172 records). Clinical descriptors accounted for more than half of all errors, despite their lower frequency in clinical notes (***Figure 4A***). While both models generated an equal number of errors from Clinical descriptors, Qwen3-32B generated more errors from Laboratory descriptors. In-depth analysis identified the cause: Qwen3-32B is more likely to “forget” to double ELISA method references and incorrectly score proteinuria from tests other than the urine protein/creatinine ratio or 24-hour urine protein.

**Figure 4.**
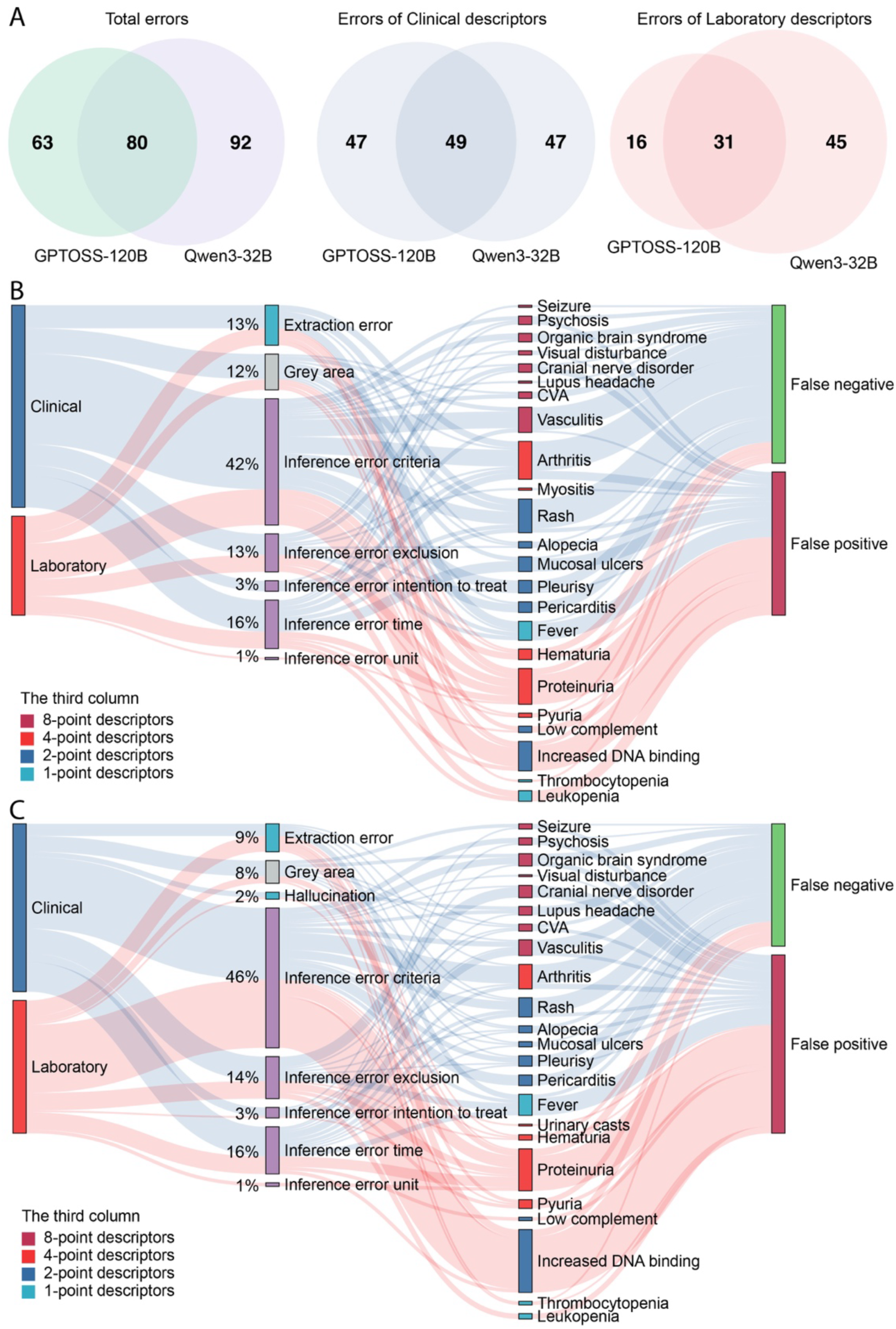
Error analysis of all error records at the descriptor level (714 cases pooled together). **(A) Comparison of error records between GPT-OSS-120B and Qwen3-32B. (B) A Sankey plot showing error records generated by first-layer GPT-OSS-120B. (C) A Sankey plot showing error records generated by first-layer Qwen3-32B.** In the Sankey plot, each line represents an error at the descriptor level, with blue lines indicating Clinical descriptors and red lines representing Laboratory descriptors.

Inference errors concerning criteria, time, and exclusions were the primary challenges, while hallucinations were rare (***Figures 4B & 4C***, the second column). Both models leaned toward false-negative errors when inferring Clinical descriptors (***Figures 4B & 4C***, last column; ***Figure S11***). This was not due to a failure to recognize relevancy but an excessively rigid application of criteria. For instance, the models demanded fulfillment of all criteria for neuropsychiatric SLE despite a clear diagnosis being stated. Similarly, for pain-based diagnosis (e.g., pericarditis), models required explicitly "typical" pain, whereas clinicians could credit atypical pain. Temporal reasoning errors involved difficulties in identifying the ongoing status. For instance, given a duration of "disease for 5 weeks", a model might erroneously conclude, "duration is 35 days exceeding 30-day threshold for scoring", failing to realize that a presently continuing condition should be scored.

We also observed the "lost in the middle" effect leading to extraction errors, especially in lengthy text from MIMIC-IV, where LLMs reach a conclusion based on a descriptor’s initial mention but overlook later serial changes. This significantly impaired LLMs’ performance in descriptors like fever in the MIMIC-IV notes, where body temperature changed quickly from ER to admission. Extraction errors also occurred when symbols like parentheses and angle brackets caused interference. (***Table S7***). The second-layer verifier corrected 25 errors from the first-layer GPT-OSS-120B output but introduced 13 new errors. For the Qwen3-32B model, the verifier corrected 11 errors while introducing 2 new ones.

### The clinical/research implementation study

108 SLE patients were included in the clinical/research implementation study [median age at baseline: 45 (39-52) years; 87% had a history of LN; duration of SLE: 14.55 (9.44-21.07) years]. After assessing serial notes for the first 5 years, the LLM classified 48 (44.4%) patients into the SSLDA group. The baseline clinical characteristics were similar between the SSLDA and non-SSLDA groups (***Table S8***). After follow-up for 18.34 (IQR 12.78, 18.91) years, the LLM-determined SSLDA group showed a significantly lower cumulative incidence of death-censored stage 3 CKD, KRT, and hospitalization compared to the non-SSLDA group (log-rank p < 0.0053, 0.044, and 0.021, respectively, ***Figures 5E & 5D, Figure S12***), with no significant difference in all-cause mortality (log-rank p=0.53, ***Figure 5C***). The LLM-determined non-SSLDA is an independent risk factor for the development of stage 3 CKD and hospitalization following adjustment for age, sex, and LN duration (adjusted hazard ratio, 5.66; 95% CI, 1.72-18.6 and 1.88 (1.06-3.33), respectively) (***Table S9***).

**Figure 5.**
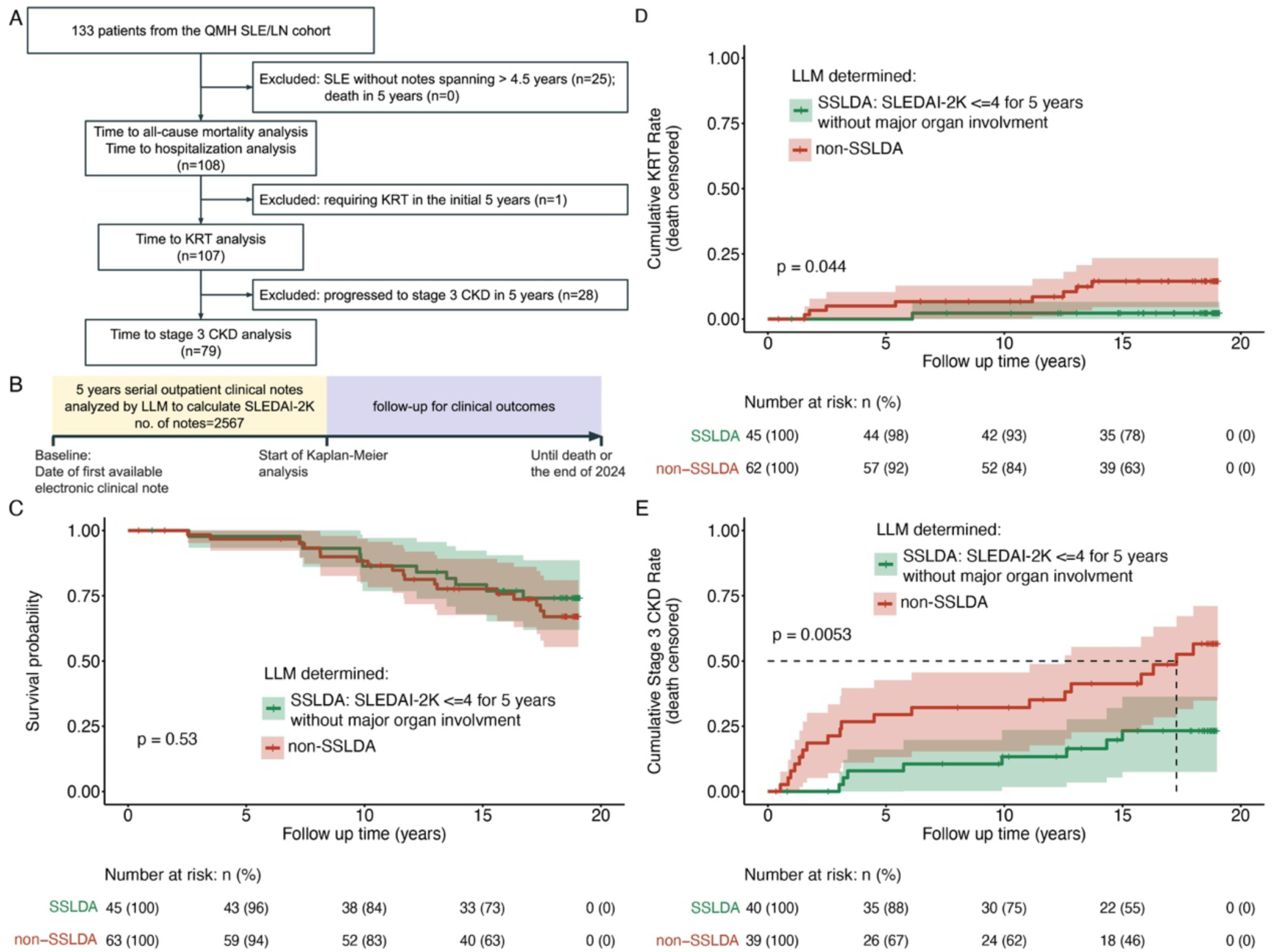
The clinical/research implementation study using large language models determined sustained SLEDAI-2K low disease activity (SSLDA) as a risk stratification tool for long-term outcomes in an LN-dominant SLE cohort. **(A) Flowchart for patient screening. (B) Study design for the clinical/research implementation study. Kaplan-Meier curve for time to (C) all-cause mortality, (D) death-censored kidney replacement therapy (KRT), and (E) death-censored stage 3 chronic kidney disease (CKD).** SSLDA is defined as SLEDAI-2K ≤4 for five consecutive years without major organ involvement (renal, central nervous system, cardiopulmonary, or vasculitis). Stage 3 CKD was defined when a patient’s estimated glomerular filtration rate (eGFR) was first documented to be persistently below 60 ml/min/1.73m² for at least 3 months. KRT is the initiation of hemodialysis, peritoneal dialysis, or kidney transplantation. The LLM used is the best-performing two-layered GPT-OSS + verifier.

## Discussion

We developed a model-agnostic and annotation-guided workflow that enables lightweight, open-source LLMs to assess SLEDAI-2K from narrative clinical notes in a secure environment. Our contribution spans the entire research workflow: (1) a representative ground truth, (2) a tailored NE framework, (3) advanced ensemble and fine-tuning strategies for LLM, and (4) rigorous evaluation and error analysis. Finally, we further validate its clinical utility through a simulated clinical/research implementation study.

We demonstrate that modern open-source reasoning LLMs can safely and efficiently perform medical concept identification and complex clinical reasoning from narrative EHR notes. Importantly, our clinical implementation successfully validates that maintaining a low SLEDAI can prevent damage accrual ^29^, substantiating LLMs’ potential for real-world deployment for clinical and research purposes. While the SLEDAI-2K may be considered a relatively easy-to-score index for specialists, it is also important to appreciate that clinics are often attended by trainees who are less familiar with the scoring systems. In real-life situations, the documentation for SLEDAI-2K may not be that comprehensive for specialists who are less research-oriented. Previous studies reported that SLEDAI documentation rates in real-world settings are as low as 10% without active interventions ^30^, and clinician-recorded SLEDAI scores are inconsistently available in real-world datasets ^31^. Instead of building a complex, resource-intensive framework to predict clinical outcomes, the primary objective of this LLM is to develop a fast, scalable framework for automated retrieval of serial SLEDAI-2K scores from electronic health records (EHR), which is useful for clinical audit and research purposes ^32^. This framework could be further adapted to other clinical instruments (e.g., SLE-DAS, BILAG), enabling highly efficient and scalable data retrieval and predictive model development. Its high performance in concept extraction also positions it as a foundation for clinical decision support systems, enabling diagnostic assistance and evidence-based practices. Results from our simulated clinical/research implementation study also indicated that SLEDAI-2K scores extracted by our LLM could risk-stratify patients and predict important clinical outcomes (e.g., renal outcomes and hospitalization). These findings lent further support to the robustness and clinical utility of our LLM. To enhance the generalizability and clinical applications, we have built an online SLEDAI-2K prompt customizer (https://reneeleung.github.io/sledai-llm/) for users to tailor-make their prompts for different clinical settings, should they want to modify the clinical criteria according to their institutional practices. Overall, this framework provides a blueprint for developing, validating, and implementing LLM-powered tools for SLE and diseases characterized by complex, multisystem clinical features.

Our LLM showed better performance in laboratory data-based evaluation than those based on clinical descriptors. However, it is important to appreciate that our model deals with clinical scenarios that are most commonly seen in real-world practice, where derangement in laboratory values is highly prevalent ^33^. Our findings align with the emerging consensus that AI should serve as a collaborative tool rather than a total replacement. Topol’s landmark review described this paradigm as "high-performance medicine": the convergence of human and artificial intelligence (AI), where AI augments clinical decision-making rather than supplanting clinical expertise ^34^. Our results reinforce the value of a Human-in-the-Loop (HITL) framework, in which AI provides scalable, standardized preliminary evaluation while the clinician retains ultimate authority over high-stakes cases, a path now advocated as the responsible use of medical AI ^35^. The best model correctly identified descriptor relevance in more than 90% of error records, which enables reliable flagging of uncertain descriptors for further review. Importantly, our current results are promising as the performance gap will have room for further improvement. The proposed framework is model-agnostic and can readily accommodate emerging open-source LLMs as they become available, enabling benchmarking and iterative improvement over time.

While Claude-3 ^27^ showed potential to reduce the diagnostic burden of SLE using EHR data, large-scale clinical implementation of such proprietary-based LLMs faces many challenges. The dependence on closed-source APIs poses limitations for clinical integration, including concerns over data privacy, institutional control, and output consistency. Furthermore, any local deployment workflow must be cost-effective to function within the limited GPU resources available in most medical institutions ^36^. Our work addresses these constraints by pivoting to a secure, affordable, verifiable, and scalable framework. The inference only requires 1-2 minutes per clinical note in a single run. Moreover, our results demonstrate that the latest open-source reasoning models, when guided by strategic ensemble and fine-tuning, can effectively achieve competitive outcomes, reinforcing prior evidence on the capacity of open models to narrow the gap with leading commercial models ^37^.

Building on the latest evidence that well-crafted prompts with domain-relevant semantic anchors enhance NER performance ^38^, our methodology leveraged comprehensive annotation guidelines to provide disease-specific clinician-curated targeted indicators to guide LLM analysis. Additionally, we incorporated error-feedback workflows to refine prompts, minimizing hallucinations and omissions ^39,40^. To inspire clinical reasoning, our prompting incorporated strategies of “intuitive reasoning” (diagnostic deduction from symptoms, signs, and paraclinical tests) and “differential diagnosis” (exclusions), which help mitigate the “black box” limitation in LLMs while maintaining diagnostic accuracy ^41^. This emphasis on prompt refinement may also explain the suboptimal accuracy (72%) in a prior study using Claude-3 for SLE diagnosis, which had limited prompt engineering ^27^. Moreover, both GPT-OSS and Qwen seem to prefer individual prompting (“single-task”) over grouped prompting (“multi-task”). This tendency, consistent with findings in a previous publication ^42^, suggests that single-task prompting allows more focused analysis and reduces the risk of being “lost in the middle” or overlooking details, though larger models may be less affected. While our LLM-as-a-judge design improves the performance, it has the risk of introducing new errors. This aligns with prior findings that LLM judge performance is closely correlated with the difficulty of reasoning tasks ^43^. Therefore, the confidence threshold may be model-dependent and requires experimentation to establish the optimal value. Future directions can consider escalating challenge cases to more powerful verifier models, akin to a “Hierarchical Referral System”.

Although our GSPO fine-tuning approaches for Qwen3 did not yield significant overall gains in contrast to findings reported in other biomedical LLM studies ^44^, they improved performance for 14 descriptors, most of which were Clinical descriptors. This suggests that symptom-based criteria, which rely on clinical judgement and reasoning, benefit from finetuning, whereas laboratory-based criteria, based on factual numeric data, already achieve strong performance without finetuning. Future work could leverage separate reward functions for different descriptor types and encourage deeper reasoning by incorporating text-matching methods to assess whether reasoning traces are consistent with relevant evidence in the clinical note.

Finally, we also find a complementary division of labor between domain-specific representation models and general-purpose generative LLMs. While fine-tuned BioClinicalBERT offers rapid, reliable NER ideal for large-scale data screening, reasoning LLMs (e.g., GPT-OSS, Qwen3) should be prioritized for advanced inference. This suggests that the previously proposed BERT-based framework for inferring lupus cutaneous activity scores from clinical notes ^28^ may benefit from integrating a reasoning LLM. Implementing an LLM-based structuring layer could be a more automated path for handling format variability. However, utilizing AI for pre-processing can introduce extraction errors and hallucinations, which require careful examination and scoping. Future work could employ similar BERT-based models as a preprocessing layer for standardizing input structure.

There are limitations to this work. Despite rigorous efforts to create high-quality annotations, real-world clinical data often involves ambiguity and atypical presentations where a single definitively "correct" answer does not exist. Our LLM’s accuracy depends on the clinical text provided, and this can be challenging for some conditions, such as distinguishing “constitutional arthralgia” from “arthritis”, when discriminating features (e.g., morning stiffness) are not explicitly documented. Reassuringly, over 70% of potential arthritis cases had additional clinical features documented to determine their inflammatory status. Nevertheless, to circumvent these issues, we shall require a multimodal, conversational framework in a prospective setting, where an LLM interacts directly with patients or clinicians. Regarding external validation, the lower exact-match of SLEDAI-2K is partly attributable to the masking of date information in MIMIC-IV, which increases ambiguity. One may argue that the external MIMIC-IV dataset lacks rare clinical descriptors such as neuropsychiatric SLE and vasculitis; this phenomenon reflects the real-world situation where these manifestations only occur in less than 3% of patient visits ^33^. Moreover, MIMIC-IV notes span the entire episode from emergency presentation to inpatient, a context in which disease activity can change rapidly. Adapting fully to this setting would require reinserting pseudo-dates and multiple rounds of prompt refinement; however, we intentionally limited such modifications to maintain methodological transparency and consistency. Finally, this study used a refined version of the SLEDAI-2K criteria rather than the original version. However, such modifications were necessary because the inherent ambiguity and outdated concepts in the original SLEDAI-2K could lead to scoring inconsistencies for annotators and LLMs.

In conclusion, this study introduces a novel, model-agnostic LLM workflow that is promising for automating SLEDAI-2K measurement from narrative clinical notes. Our simulated clinical/research implementation demonstrated the clinical and research utility of our framework in SLE/LN. Our framework can serve as a blueprint for the development, validation, and implementation of open-source LLM-powered clinical tools for SLE and other rheumatic diseases.

## Methods

This study is divided into two parts: 1) development and validation of a named entity framework to facilitate the automation of SLEDAI-2K scoring from free-text clinical notes leveraging LLMs, and 2) simulating a clinical implementation of the developed framework. ***Figure 1*** depicts the data curation process and the named entity framework.

### Ethics statement

This study received approval from the Institutional Review Board of The University of Hong Kong/Hospital Authority Hong Kong West Cluster for use of clinical notes (UW 24-255) and clinical data (UW 25-243). Consent from the patient was waived for de-identified data.

### Data sources

As shown in ***Figure 1A***, three different sources of clinical narrative notes/texts were used for the comprehensive evaluation of the named entity framework. The outpatient notes were obtained from a SLE/LN cohort of 133 patients from Queen Mary Hospital (QMH) who were clinically diagnosed with SLE according to the ACR 1997 classification criteria, excluding those with overlap syndrome. To capture a richer SLE phenotype, we retrieved all available serial outpatient notes (9,231 notes) from the Clinical Management System (2000 to mid-2024) **(*Figure S13*)**. All real-world clinical notes underwent data cleaning and de-identification procedures, including filtering of incomplete metadata and masking of identifiers **(*Figure S14*)**. An automated history removal step was then applied to exclude text unrelated to SLEDAI-2K scoring within the preceding 30 days **(*Figure S15*)**. To increase rare descriptor cases, our study incorporated doctor-synthesized pseudo notes modified from the outpatient notes and published case reports (described below). Together, these two data sources constituted an SLEDAI-representative internal set. An external test set was also included, comprising 56 MIMIC-IV discharge summaries ^45^ corresponding to 51 patients with SLE as the primary discharge diagnosis. The study used MIMIC-IV data under PhysioNet credentialed access granted in March 2025. Analyses were conducted on de-identified clinical notes. No patient-level records or source-note excerpts are included in this manuscript. All notes were written in English. Structured data used in the simulated clinical implementation were retrieved from the Clinical Data Analysis and Reporting System (CDARS).

### Data curation for the LLM evaluation study

We curated three sets of clinical notes for the LLM evaluation study (***Figure 1A***). Given the numerous rare descriptors (e.g., neuropsychiatric SLE), we first oversampled 489 notes likely to contain such rare cases from QMH outpatient notes with rule-based keyword filtering and LLM assistance **(*Supplementary Note 1, Figure S16 & Table S10***). Human annotation of these notes revealed that several descriptors remained under-represented; hence, 169 pseudo-cases were synthesized by modifying real-world outpatient notes and rewriting published case reports (***Supplementary Note 2, Table S11***). These 658 notes from 250 patients formed the internal comprehensive SLE clinical dataset, which provides comprehensive coverage and balanced representation across all SLEDAI-2K descriptors. The dataset was further divided into training (for fine-tuning only) and testing sets (for performance evaluation) in a 1:1 ratio, ensuring each descriptor was represented in both. Each patient was assigned exclusively to one subset to avoid patient-specific bias **(*Supplementary Note 3*)**. The external MIMIC-IV validation set was used to validate the best-performing LLMs. Cases were selected based on SLE-related ICD codes, availability of laboratory and physical exam data, and ‘lupus’ in the first discharge diagnosis item **(*Figure S17*)**. Among 21 sections of MIMIC discharge summaries, 10 sections related to admission status were included for assessing SLEDAI-2K at admission **(*Table S12*)**.

### Named entity framework development

The NE framework included annotation guidelines, ground-truth labeling, and prompts for local LLMs to execute (***Figure 1B***). Both the human annotations (***Figure 1B, steps 1 to 3***) and the LLM predictions (***Figure 1B, steps 4 to 7***) sought to determine three outcomes: 1) descriptor relevancy (relevant vs. irrelevant), 2) descriptor classification (scored vs. unscored), and 3) total SLEDAI-2K score. A relevancy is the presence of any clinical concept (e.g., diagnosis, symptoms, or tests) relevant to a descriptor. To be scored, a relevant descriptor must fulfill the diagnostic criteria within the 30-day assessment window.

The annotation guidelines refined the original SLEDAI-2K criteria ^4^, in which the 24 descriptors are categorized into two types with distinct annotation schema: 16 Clinical descriptors require a comprehensive clinical appraisal; 8 Laboratory descriptors rely on laboratory findings (***see annotation guidelines Tables 2 & 8 & 9***). Annotations were performed independently by two clinicians experienced in SLE research (D.Z. & Y.L.), and discrepancies were escalated to senior experts (E.W., S.C., and D.Y.) to reach consensus **(*Supplementary Note 4*)**.

We evaluated seven state-of-the-art LLMs covering different model sizes using NVIDIA L40S GPUs. These include five prompt-based LLMs **(*Table S13*)** and two different fine-tuning methods. A universal inference prompt directing LLMs to identify all descriptors from each clinical note was designed to incorporate three components: task instructions tailored for each descriptor, a clinical note, and the pre-defined output format (***Supplementary Prompts).*** We evaluated LLMs with reasoning enabled versus non-reasoning, and deterministic configurations versus non-deterministic (via a 3-out-of-5 majority vote). We employed two prompting strategies: 1) *individual-descriptor* prompting, in which each prompt was restricted to a single descriptor for focused analysis, and 2) *grouped-descriptors* prompting, where descriptors of the same type were combined within a single prompt to leverage contextual relationships. We also tried an LLM-as-a-judge system, where we used a second-layer GPT-OSS-120B verifier to correct potential errors when the first layer’s confidence is low (***Figure 2A, Supplementary Note 5***). The two fine-tuning strategies include supervised fine-tuning (SFT) ^46^ for BioClinicalBERT and Group Sequence Policy Optimization (GSPO) ^15^ for the Qwen3-14B model, chosen based on the computing power we had available. BioClinicalBERT was fine-tuned on our NER framework to identify descriptor entities and attributes from clinical notes. For GSPO, we used a reward function with format, correctness, and reasoning components (***Supplementary Note 6, Figures S18 & S19***). These models were fine-tuned on the internal training set and further evaluated on the testing set along with other pretrained LLMs.

To measure the certainty of LLMs, we calculated case confidence based on the model-generated confidence scores (***Supplementary Note 7***). The predicted SLEDAI-2K score was calculated using the assigned weights (***Supplementary Note 8***). Model performance was evaluated using F1, precision, recall, and AUC (Area Under the Curve) per descriptor, micro-F1 for cross-descriptors, and root mean squared error (RMSE) for SLEDAI-2K scores (***Supplementary Note 9***). AUC, accuracy, sensitivity, specificity, positive predictive value, and negative predictive value were computed to evaluate four binary SLEDAI-2K classifications: SLEDAI-2K ≥ 4, SLEDAI-2K ≥ 6, renal SLEDAI-2K ≥ 4 (four renal descriptors), and clinical SLEDAI-2K ≥ 4 (excluding increased DNA binding and low complement). Errors in LLM rationales for misclassified descriptors were classified into four types: inference errors, extraction errors, hallucinations, and the grey area (***Supplementary Note 10***).

### The clinical/research implementation study

To validate the utility of our framework, we simulated a clinical/research implementation by extracting LLM-based SLEDAI-2K scores and examining their relationships with long-term clinical outcomes (***Figure 1A***). The LLM assessed five-year serial SLEDAI-2K scores in 108 SLE patients with ≥5 years of outpatient records (2,576 outpatient notes) (***Figures 5A & 5B***). Patients were stratified into sustained SLEDAI-2K low disease activity (SSLDA) group [defined as SLEDAI-2K score ≤ 4 in the absence of major organ involvement throughout five years ^29^] and non-SSLDA group. The cumulative incidence of four clinical outcomes [all-cause mortality, chronic kidney disease (CKD), kidney replacement therapy (KRT), and hospitalization] at last follow-up was compared between the SSLDA and non-SSLDA groups using Kaplan-Meier analysis and the log-rank test. A two-sided p-value < 0.05 was statistically significant (***Supplementary Note 11***).

## Supporting information

Annotation Guidelines

## Data Availability

Due to privacy regulations in Hong Kong, clinical notes cannot be made publicly available. Only a few anonymised notes are referenced where essential for annotation guidance or reviewing the reasoning behind the LLMs’ responses. An online SLEDAI-2K prompt customizer for tailoring prompts according to different clinical settings is available at https://reneeleung.github.io/sledai-llm/. The annotated pseudo-clinical notes used in this study are available on Zenodo (https://doi.org/10.5281/zenodo.20042443). The MIMIC-IV data used in this study are available through PhysioNet, subject to credentialed access and completion of required training. In accordance with the PhysioNet Credentialed Health Data License, the authors are not permitted to redistribute MIMIC-IV data in any form, including discharge summaries, patient-level labels, prompts containing source text, or derived datasets. Aggregate results or code base may be shared upon request to the corresponding author.

## Code Availability

All code is publicly available at https://github.com/reneeleung/sledai-llm.

## Acknowledgements

This research was supported by the Health and Medical Research Fund (reference no.: CID-HKU2) and the AIR@InnoHK, administered by the Innovation and Technology Commission of the Government of the Hong Kong Special Administrative Region. These funders played no role in study design, data collection, analysis and interpretation of data, or the writing of this manuscript. We gratefully acknowledge Icons8 (https://icons8.com/) for providing the free graphical elements used in Figures 1 and 2.

## Author Contributions

Conceptualization: D.Z., C.W., S.C., C.K.W., K.L., Z.W., J.W., D.Y. Methodology: D.Z., R.L., C.W., E.T., T.W., C.K.W., K.L., Z.W., J.W., D.Y. Software: R.L., E.T., T.W. Validation: D.Z., R.L., S.C., T.C., C.L., D.Y. Formal Analysis: D.Z., R.L. Investigation: D.Z., R.L., Y.L. Resources: S.C., T.C., C.L., C.K.W., K.L., J.W., D.Y. Data Curation: D.Z., R.L., C.W., S.C., Y.L., Z.W., D.Y. Writing-Original Draft: D.Z., R.L. Writing-Review & Editing: C.W., K.L., Z.W., J.W., D.Y. Visualization: D.Z., R.L. Supervision: K.L., Z.W., J.W., D.Y. Project Administration: K.L., Z.W., J.W., D.Y. Funding Acquisition: J.W.

