## Supplementary File for "Automated Disease Activity Assessment in Systemic Lupus Erythematosus Using Privacy-Preserving Large Language Models"

### Supplementary Information

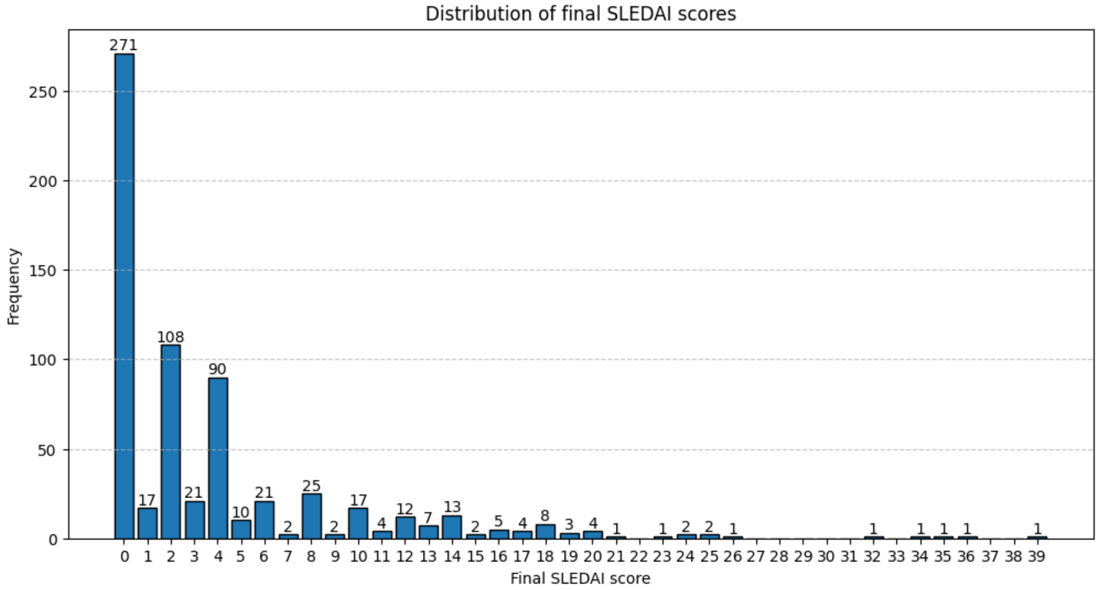

**Figure S1. Distribution of SLEDAI-2K scores in the Comprehensive SLE Clinical Dataset.** A bar chart illustrating the number of cases corresponding to each SLEDAI-2K score in the ground-truth annotated dataset of 658 cases. In summary, the distribution of SLEDAI-2K was right-skewed, with 41.2% of cases scoring 0, 37.4% falling into the mild activity range (1-5), 10.2% in moderate (6-10), 8.8% in high (11-19), and 2.4% in very high (≥20) disease activity levels.

**Table S1. Inter-annotator agreement scores for span-level named entity annotation.** Metrics include span F1, entity Cohen's κ, and attribute-level agreement for SLEDAI criteria, time, and special entity annotations.

| Inter-annotator agreement scores | |
|---|---|
| Span F1 | 0.80 |
| Entity κ | 0.87 |
| Attribute agreement scores | |
| SLEDAI criteria | 0.81 |
| Time | 0.92 |
| Special entity | 0.58 |

**Table S2. Descriptor distributions of the Comprehensive SLE Clinical Dataset.** A table summarizing the frequency of positive and negative descriptors in the ground-truth annotated dataset of 658 cases.

| Descriptor | Positive | Relevant & Negative | Irrelevant |
|---|---|---|---|
| seizure | 12 (1.8%) | 28 (4.3%) | 618 (93.9%) |
| psychosis | 10 (1.5%) | 21 (3.2%) | 627 (95.3%) |
| organic_brain_syndrome | 13 (2.0%) | 18 (2.7%) | 627 (95.3%) |
| visual_disturbance | 12 (1.8%) | 43 (6.5%) | 603 (91.6%) |
| cranial_nerve_disorder | 11 (1.7%) | 16 (2.4%) | 631 (95.9%) |
| lupus_headache | 10 (1.5%) | 67 (10.2%) | 581 (88.3%) |
| CVA | 12 (1.8%) | 26 (4.0%) | 620 (94.2%) |
| vasculitis | 16 (2.4%) | 19 (2.9%) | 623 (94.7%) |
| arthritis | 38 (5.8%) | 246 (37.4%) | 374 (56.8%) |
| myositis | 11 (1.7%) | 27 (4.1%) | 620 (94.2%) |
| urinary_casts | 11 (1.7%) | 11 (1.7%) | 636 (96.7%) |
| hematuria | 41 (6.2%) | 197 (29.9%) | 420 (63.8%) |
| proteinuria | 84 (12.8%) | 333 (50.6%) | 241 (36.6%) |
| pyuria | 14 (2.1%) | 18 (2.7%) | 626 (95.1%) |
| rash | 43 (6.5%) | 198 (30.1%) | 417 (63.4%) |
| alopecia | 39 (5.9%) | 70 (10.6%) | 549 (83.4%) |
| mucosal_ulcers | 21 (3.2%) | 91 (13.8%) | 546 (83.0%) |
| pleurisy | 15 (2.3%) | 46 (7.0%) | 597 (90.7%) |
| pericarditis | 13 (2.0%) | 19 (2.9%) | 626 (95.1%) |
| low_complement | 175 (26.6%) | 107 (16.3%) | 376 (57.1%) |
| increased_DNA_binding | 123 (18.7%) | 202 (30.7%) | 333 (50.6%) |
| fever | 19 (2.9%) | 145 (22.0%) | 494 (75.1%) |
| thrombocytopenia | 44 (6.7%) | 92 (14.0%) | 522 (79.3%) |
| leukopenia | 38 (5.8%) | 104 (15.8%) | 516 (78.4%) |

If it is scored for SLEDAI, the case is classified as positive for a certain descriptor; otherwise, it is considered negative. Relevant but negative cases are those where the descriptor-related clinical concept is mentioned but cannot be scored for SLEDAI. After incorporating pseudo cases into the real cases, we have at least ten positive and ten relevant and negative cases for evaluating model performance.

**Table S3. Demographics of the selected MIMIC-IV data.**

| Characteristics | Total |
|---|---|
| | Patients (n=51) |
| Female, n (%) | 50 (98.0) |
| Race, n (%) | |
| Asian | 13 (25.5) |
| White | 12 (23.5) |
| Black or African | 15 (29.4) |
| Other | 11 (21.6) |

Note: The selected MIMIC-IV data cohort includes 51 unique patients, of whom only one is male. The racial distribution is as follows: Asian (25.5%), White (23.5%), Black or African American (29.4%), and Other (21.6%). Across the 56 discharge notes, the mean age at admission was 35.02 years, and the average note length was 890.52 words.

**Table S4. Descriptor distributions of the selected MIMIC-IV data.** A table summarizing the frequency of positive and negative descriptors in the ground-truth annotated dataset of 56 cases.

| Descriptor | Positive | Relevant & Negative | Irrelevant |
|---|---|---|---|
| seizure | 1 (1.8%) | 1 (1.8%) | 54 (96.4%) |
| psychosis | 1 (1.8%) | 2 (3.6%) | 53 (94.6%) |
| organic_brain_syndrome | 0 (0.0%) | 2 (3.6%) | 54 (96.4%) |
| visual_disturbance | 0 (0.0%) | 11 (19.6%) | 45 (80.4%) |
| cranial_nerve_disorder | 0 (0.0%) | 0 (0.0%) | 56 (100.0%) |
| lupus_headache | 0 (0.0%) | 15 (26.8%) | 41 (73.2%) |
| CVA | 2 (3.6%) | 0 (0.0%) | 54 (96.4%) |
| vasculitis | 3 (5.4%) | 8 (14.3%) | 45 (80.4%) |
| arthritis | 8 (14.3%) | 20 (35.7%) | 28 (50.0%) |
| myositis | 2 (3.6%) | 12 (21.4%) | 42 (75.0%) |
| urinary_casts | 0 (0.0%) | 1 (1.8%) | 55 (98.2%) |
| hematuria | 3 (5.4%) | 22 (39.3%) | 31 (55.4%) |
| proteinuria | 10 (17.9%) | 15 (26.8%) | 31 (55.4%) |
| pyuria | 7 (12.5%) | 16 (28.6%) | 33 (58.9%) |
| rash | 14 (25.0%) | 25 (44.6%) | 17 (30.4%) |
| alopecia | 4 (7.1%) | 4 (7.1%) | 48 (85.7%) |
| mucosal_ulcers | 5 (8.9%) | 12 (21.4%) | 39 (69.6%) |
| pleurisy | 7 (12.5%) | 17 (30.4%) | 32 (57.1%) |
| pericarditis | 3 (5.4%) | 15 (26.8%) | 38 (67.9%) |
| low_complement | 27 (48.2%) | 8 (14.3%) | 21 (37.5%) |
| increased_DNA_binding | 12 (21.4%) | 16 (28.6%) | 28 (50.0%) |
| fever | 1 (1.8%) | 46 (82.1%) | 9 (16.1%) |
| thrombocytopenia | 3 (5.4%) | 12 (21.4%) | 41 (73.2%) |
| leukopenia | 5 (8.9%) | 47 (83.9%) | 4 (7.1%) |

Note: Relevant cases are those in which the descriptor is mentioned. If both the diagnostic and time criteria are fulfilled, the case is classified as positive for a certain descriptor; otherwise, it is considered negative. The distribution of positive samples was highly imbalanced across descriptors. Several descriptors, including psychosis, visual disturbance, cranial nerve disorder, lupus headache, and urinary casts, contained no positive cases in the dataset. In addition, seizure, organic brain syndrome, myositis, and fever were represented by only a single positive case each.

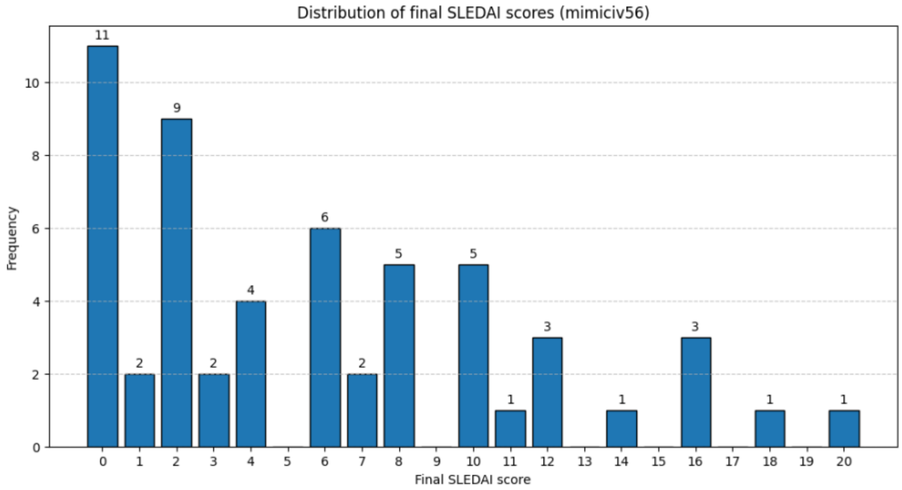

**Figure S2. Distribution of SLEDAI-2K scores in the selected MIMIC-IV data.** A bar chart illustrating the number of cases corresponding to each SLEDAI-2K score in the ground-truth annotated dataset of 56 cases.

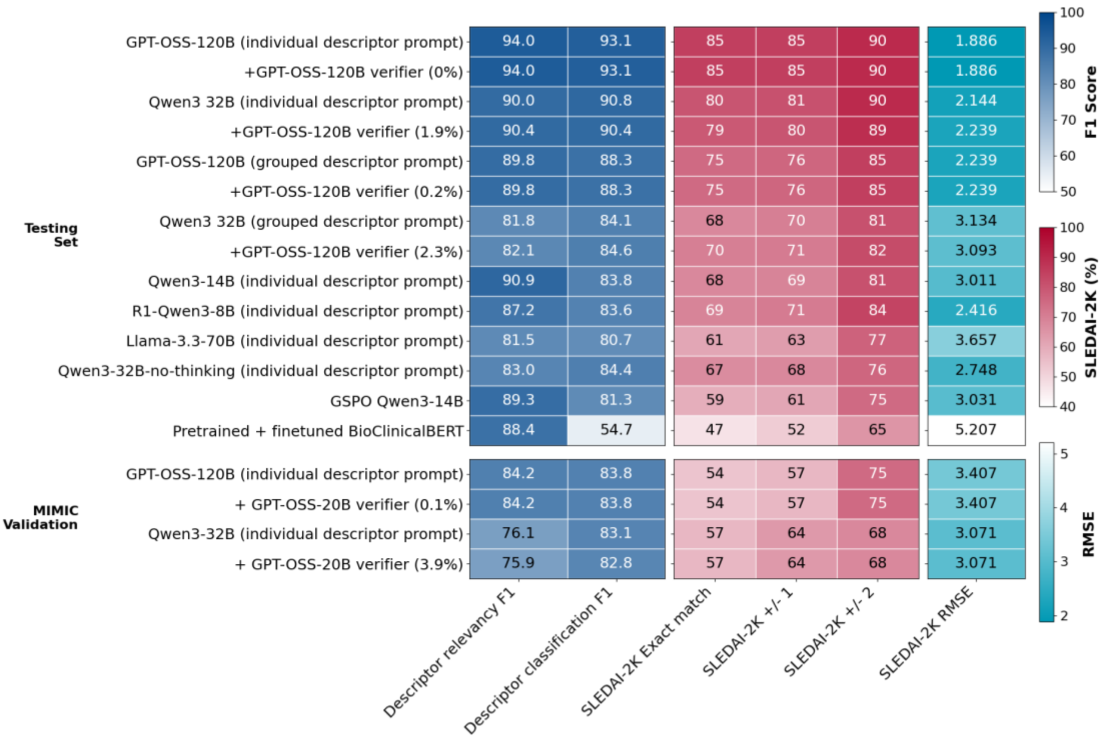

**Figure S3. Heatmap of deterministic performance on the testing set and MIMIC-IV validation set.** A heatmap comparing deterministic performances (temperature of 0) of different LLMs and prompting strategies on the testing set and MIMIC-IV validation set, including descriptor relevancy F1, descriptor classification F1, SLEDAI-2K exact match, within tolerance of 1 and 2, and RMSE. Deterministic performance is slightly poorer than the majority voting approach under recommended temperature settings, suggesting that creativity may be required for complex diagnostic reasoning tasks. For the MIMIC-IV data, no outputs had model-generated confidence of less than the threshold. Thus, only performance of the one-layer GPT-OSS-120B is reported.

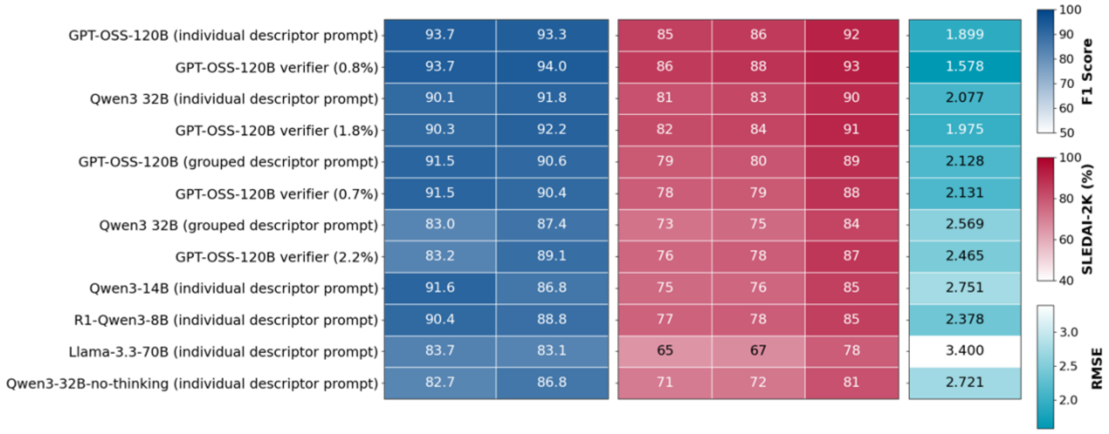

**Figure S4. Heatmap of performance on the full dataset for models without finetuning.** A heatmap presenting the F1-scores for descriptor relevancy identification and descriptor classification, SLEDAI-2K exact match, +/- 1, +/- 2, and RMSE, evaluated on the full dataset of 658 cases. Descriptor relevancy identification measures whether the model correctly identifies which entities are relevant or irrelevant within the input clinical note, while descriptor classification measures whether the model assigns the correct binary score to each entity. The percentage shown next to the verifier represents the proportion of prompts passed to the second layer. Consistent with the testing set performance reported in the primary study, the GPT-OSS-120B model achieved the highest accuracy of 0.933 descriptor classification F1, with 85% exact match, 92% within ±2 points, and root mean squared errors (RMSE) of 1.899.

**Table S5. Model performance for binary SLEDAI-2K criteria across the internal and external datasets (GPT-OSS-120B + GPT-OSS-120B verifier).**

| Criterion | Dataset | True positive cases | AUC (%) | Accuracy (%) | Sensitivity (%) | Specificity (%) | PPV (%) | NPV (%) |
|---|---|---|---|---|---|---|---|---|
| SLEDAI-2K >= 4 | Internal full (N=658) | 243 | 96.8 | 94.8 | 93.8 | 95.4 | 92.3 | 96.4 |
| | Internal test (N=358) | 134 | 96.6 | 94.7 | 94.0 | 95.1 | 92.0 | 96.4 |
| | External (N=56) | 32 | 95.6 | 94.6 | 96.9 | 91.7 | 93.9 | 95.7 |
| SLEDAI-2K >= 6 | Internal full (N=658) | 142 | 96.8 | 97.0 | 93.7 | 97.9 | 92.4 | 98.2 |
| | Internal test (N=358) | 79 | 97.2 | 97.2 | 94.9 | 97.8 | 92.6 | 98.6 |
| | External (N=56) | 28 | 95.2 | 89.3 | 82.1 | 96.4 | 95.8 | 84.4 |
| Renal SLEDAI-2K >= 4 | Internal full (N=658) | 100 | 98.7 | 98.0 | 99.0 | 97.8 | 89.2 | 99.8 |
| | Internal test (N=358) | 60 | 98.3 | 97.8 | 98.3 | 97.7 | 89.4 | 99.7 |
| | External (N=56) | 12 | 90.5 | 92.9 | 83.3 | 95.5 | 83.3 | 95.5 |
| Clinical SLEDAI-2K >= 4 | Internal full (N=658) | 196 | 96.1 | 95.9 | 93.9 | 96.8 | 92.5 | 97.4 |
| | Internal test (N=358) | 105 | 95.6 | 95.8 | 93.3 | 96.8 | 92.5 | 97.2 |
| | External (N=56) | 28 | 92.0 | 91.1 | 89.3 | 92.9 | 92.6 | 89.7 |

All metrics are reported as percentages. PPV = positive predictive value; NPV = negative predictive value; AUC = area under ROC curve. Clinical SLEDAI-2K excludes increased DNA binding and low complement. Renal SLEDAI-2K includes four renal descriptors: hematuria, pyuria, proteinuria, and urinary casts.

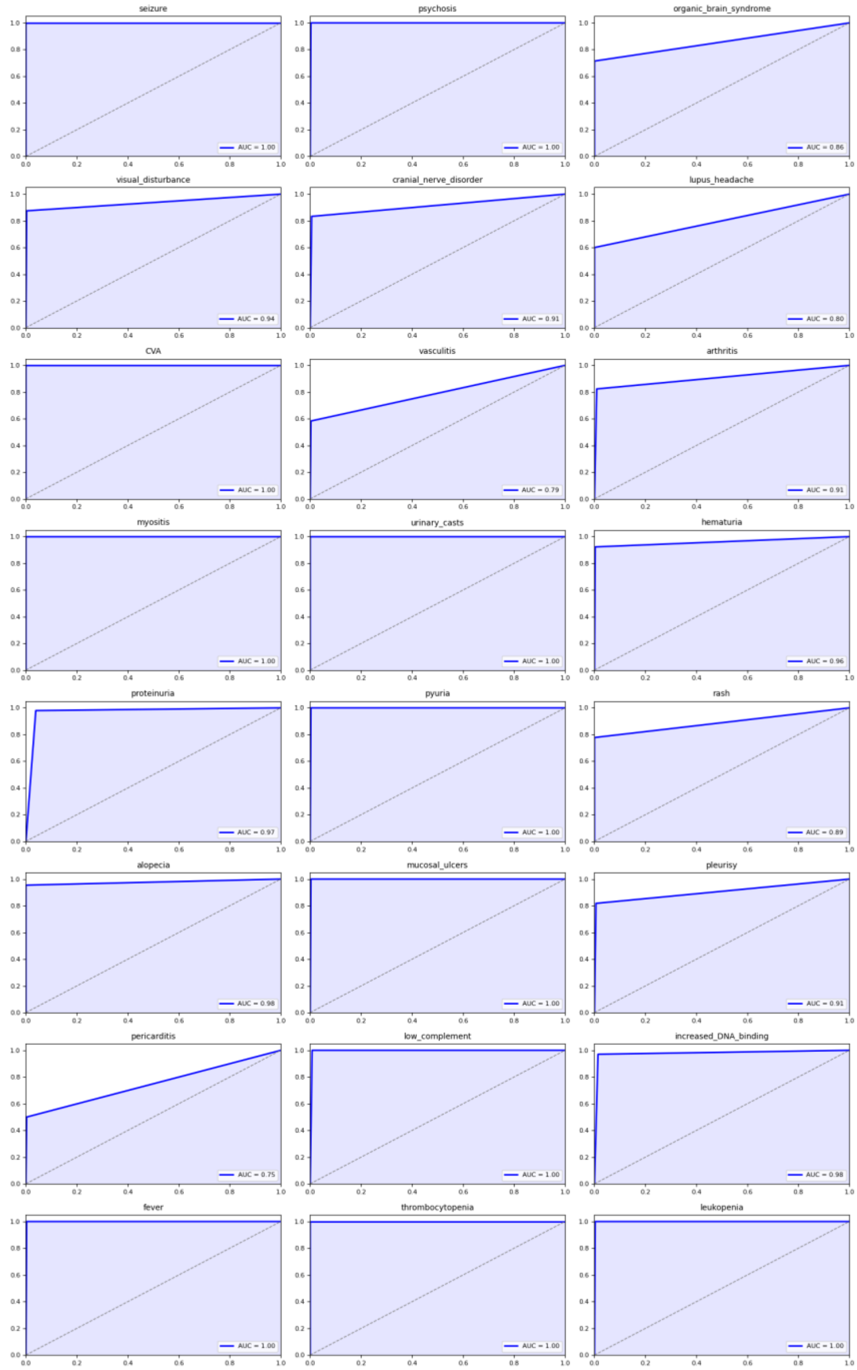

**Figure S5. ROC curves by descriptor on testing dataset (GPT-OSS-120B + verifier model).** Receiver operating characteristic (ROC) curves and AUC for each descriptor, evaluated on the testing set of 358 cases. Across most descriptors, the ROC curves indicate strong discriminative ability. Several descriptors (seizure, psychosis, organic brain syndrome, lupus headache, arthritis, and rash) show moderate discriminative patterns, while pleurisy and pericarditis display weaker separation between positives and controls.

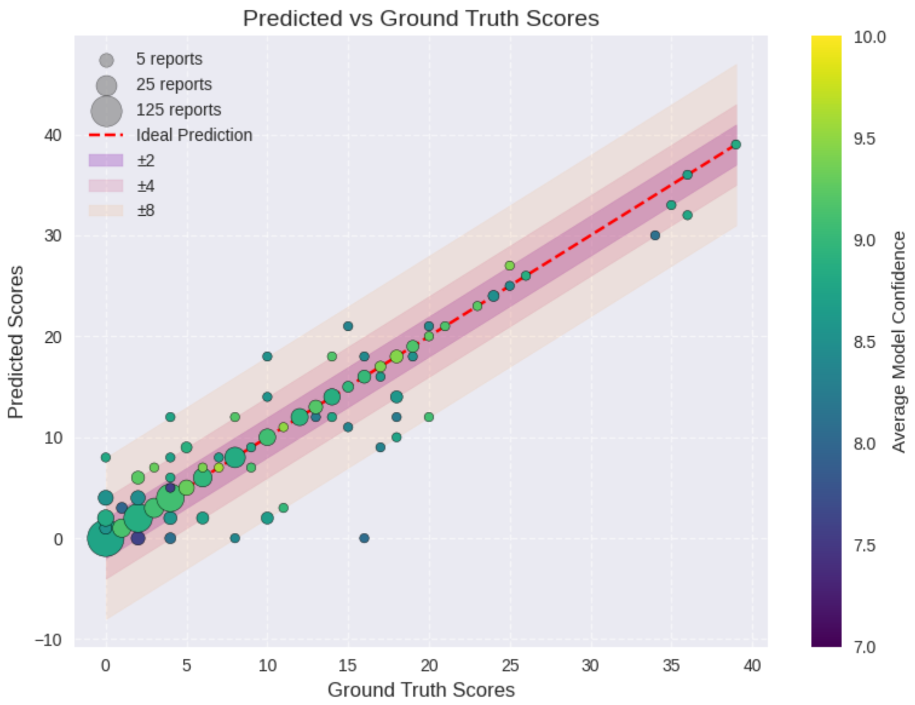

**Figure S6. Scatter plots illustrate the agreement between model-predicted scores and the actual ground truth scores (GPT-OSS-120B + verifier model).** Evaluated on the full dataset of 658 cases.

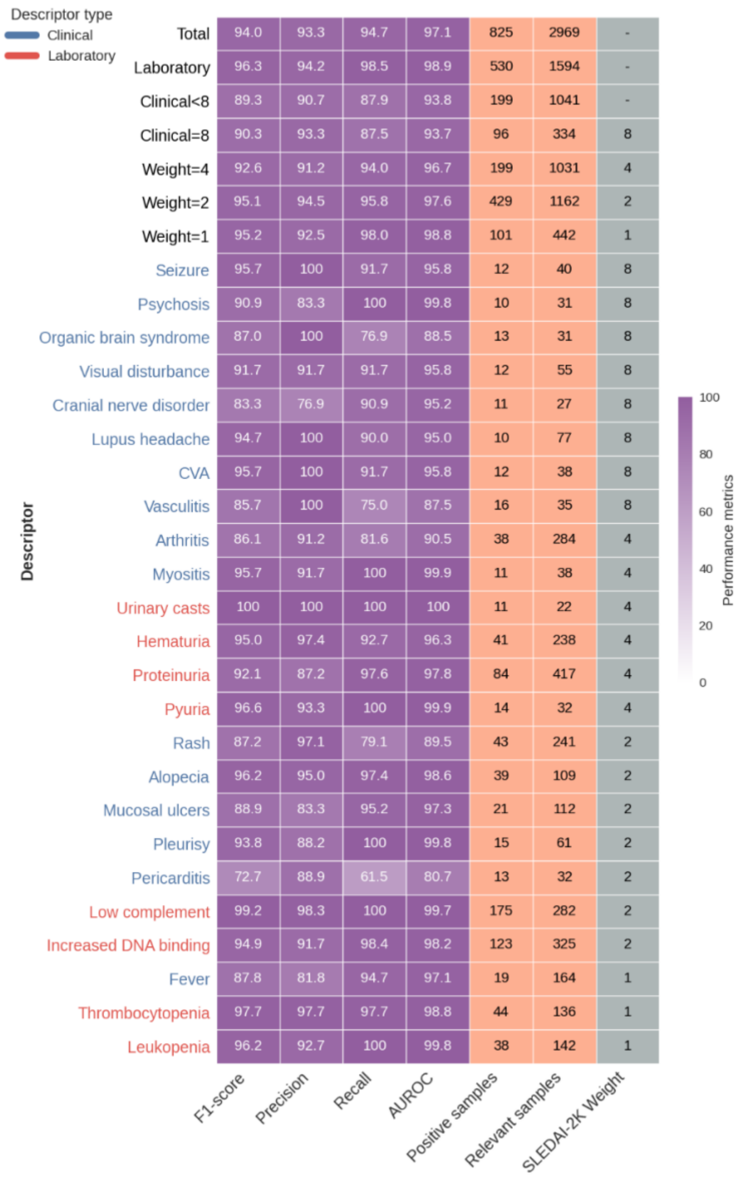

**Figure S7. Heatmap of descriptor-level performance on full dataset (GPT-OSS-120B + verifier model).** A heatmap table presenting precision, recall, and F1-score for each descriptor entity, along with the overall micro-averaged F1 score, for the best-performing GPT-OSS-120B + verifier model evaluated on the full dataset of 658 cases.

(a)

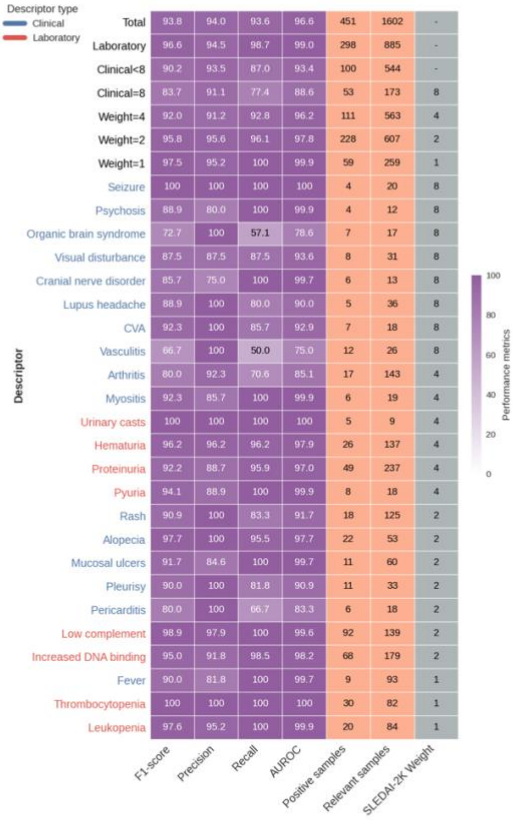
Descriptor type
Clinical
Laboratory
Descriptor
Total 93.8 94.0 93.6 96.6 451 1602 -
Laboratory 96.6 94.5 98.7 99.0 298 885 -
Clinical<8 90.2 93.5 87.0 93.4 100 544 -
Clinical=8 83.7 91.1 77.4 88.6 53 173 8
Weight=4 92.0 91.2 92.8 96.2 111 563 4
Weight=2 95.8 95.6 96.1 97.8 228 607 2
Weight=1 97.5 95.2 100 99.9 59 259 1
Seizure 100 100 100 100 4 20 8
Psychosis 88.9 80.0 100 99.9 4 12 8
Organic brain syndrome 72.7 100 57.1 78.6 7 17 8
Visual disturbance 87.5 87.5 87.5 93.6 8 31 8
Cranial nerve disorder 85.7 75.0 100 99.7 6 13 8
Lupus headache 88.9 100 80.0 90.0 5 36 8
CVA 92.3 100 85.7 92.9 7 18 8
Vasculitis 66.7 100 50.0 75.0 12 26 8
Arthritis 80.0 92.3 70.6 85.1 17 143 4
Myositis 92.3 85.7 100 99.9 6 19 4
Urinary casts 100 100 100 100 5 9 4
Hematuria 96.2 96.2 96.2 97.9 26 137 4
Proteinuria 92.2 88.7 95.9 97.0 49 237 4
Pyuria 94.1 88.9 100 99.9 8 18 4
Rash 90.9 100 83.3 91.7 18 125 2
Alopecia 97.7 100 95.5 97.7 22 53 2
Mucosal ulcers 91.7 84.6 100 99.7 11 60 2
Pleurisy 90.0 100 81.8 90.9 11 33 2
Pericarditis 80.0 100 66.7 83.3 6 18 2
Low complement 98.9 97.9 100 99.6 92 139 2
Increased DNA binding 95.0 91.8 98.5 98.2 68 179 2
Fever 90.0 81.8 100 99.7 9 93 1
Thrombocytopenia 100 100 100 100 30 82 1
Leukopenia 97.6 95.2 100 99.9 20 84 1
F1-score
Precision
Recall
AUROC
Positive samples
Relevant samples
SLEDAI-2K Weight
Performance metrics
100
80
60
40
20
0

(b)

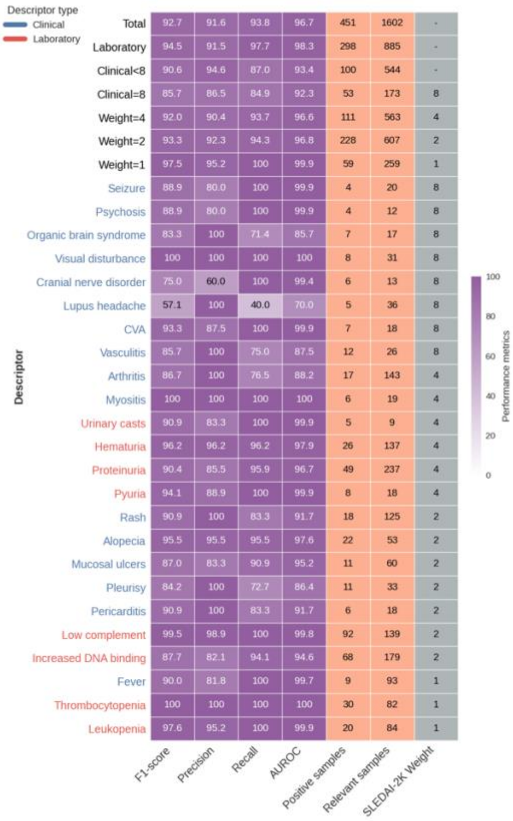
Descriptor type
Clinical
Laboratory
Descriptor
Total 92.7 91.6 93.8 96.7 451 1602 -
Laboratory 94.5 91.5 97.7 98.3 298 885 -
Clinical<8 90.6 94.6 87.0 93.4 100 544 -
Clinical=8 85.7 86.5 84.9 92.3 53 173 8
Weight=4 92.0 90.4 93.7 96.6 111 563 4
Weight=2 93.3 92.3 94.3 96.8 228 607 2
Weight=1 97.5 95.2 100 99.9 59 259 1
Seizure 88.9 80.0 100 99.9 4 20 8
Psychosis 88.9 80.0 100 99.9 4 12 8
Organic brain syndrome 83.3 100 71.4 85.7 7 17 8
Visual disturbance 100 100 100 100 8 31 8
Cranial nerve disorder 75.0 60.0 100 99.4 6 13 8
Lupus headache 57.1 100 40.0 70.0 5 36 8
CVA 93.3 87.5 100 99.9 7 18 8
Vasculitis 85.7 100 75.0 87.5 12 26 8
Arthritis 86.7 100 76.5 88.2 17 143 4
Myositis 100 100 100 100 6 19 4
Urinary casts 90.9 83.3 100 99.9 5 9 4
Hematuria 96.2 96.2 96.2 97.9 26 137 4
Proteinuria 90.4 85.5 95.9 96.7 49 237 4
Pyuria 94.1 88.9 100 99.9 8 18 4
Rash 90.9 100 83.3 91.7 18 125 2
Alopecia 95.5 95.5 95.5 97.6 22 53 2
Mucosal ulcers 87.0 83.3 90.9 95.2 11 60 2
Pleurisy 84.2 100 72.7 86.4 11 33 2
Pericarditis 90.9 100 83.3 91.7 6 18 2
Low complement 99.5 98.9 100 99.8 92 139 2
Increased DNA binding 87.7 82.1 94.1 94.6 68 179 2
Fever 90.0 81.8 100 99.7 9 93 1
Thrombocytopenia 100 100 100 100 30 82 1
Leukopenia 97.6 95.2 100 99.9 20 84 1
F1-score
Precision
Recall
AUROC
Positive samples
Relevant samples
SLEDAI-2K Weight
Performance metrics
100
80
60
40
20
0

(c)

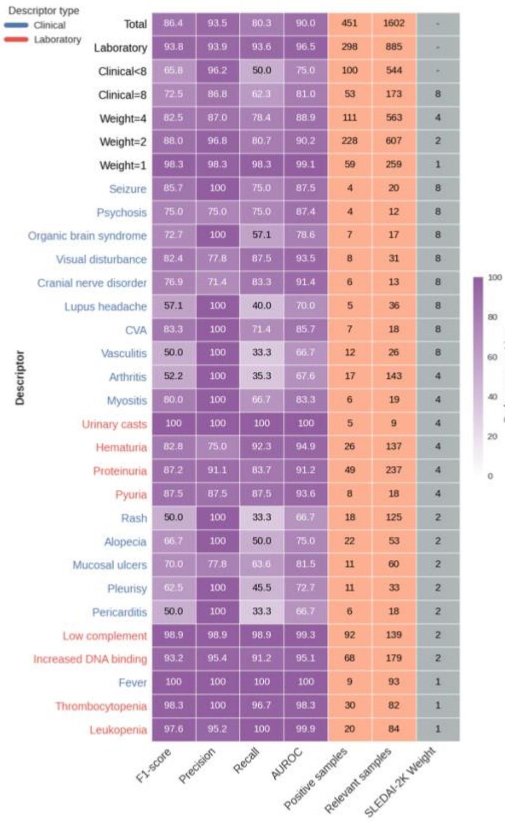

(d)

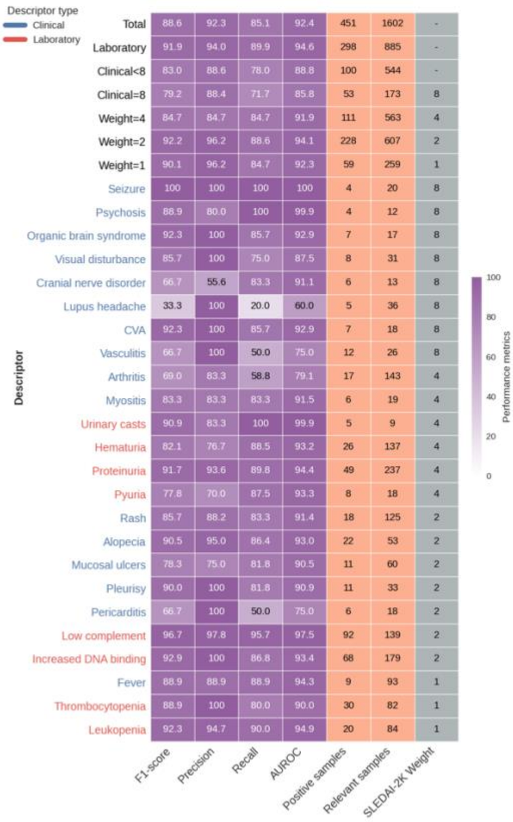

(e)

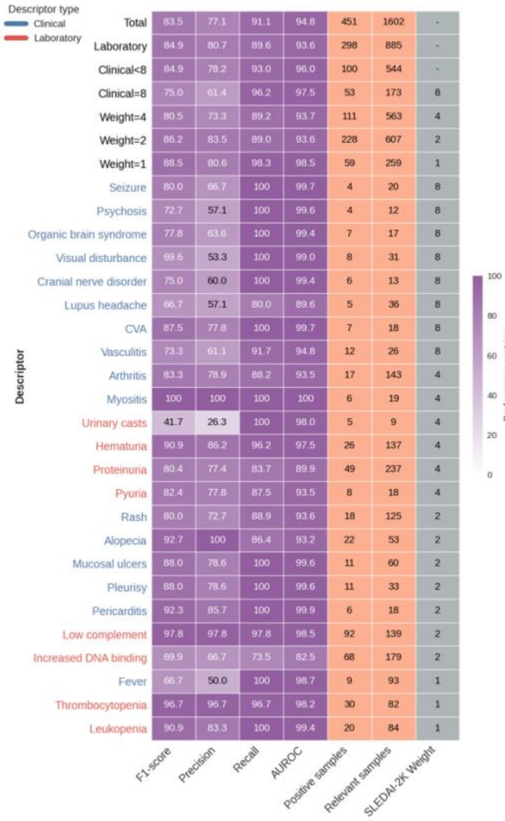

(f)

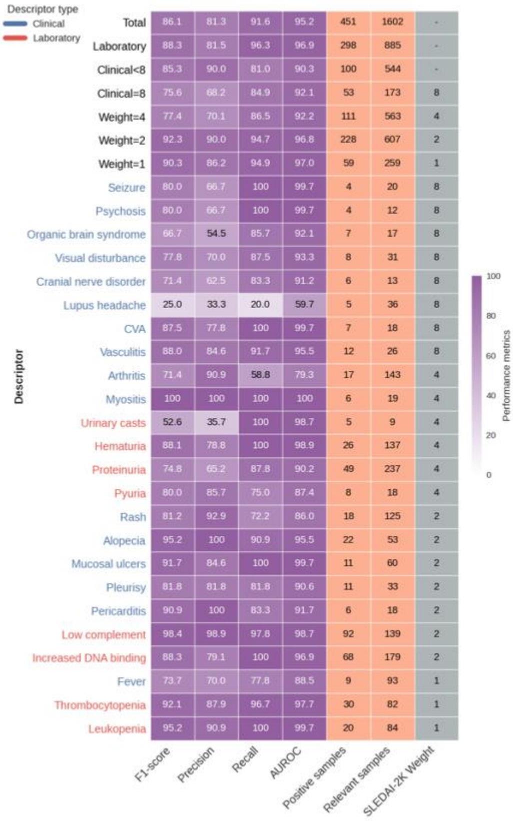

**Figure S8. Heatmaps of by-descriptor performance of LLMs on the testing dataset.** By-descriptor performance including F1, precision, recall, and AUROC on the testing set for (a) GPT-OSS-120B, (b) Qwen3-32B, (c) Qwen3-14B, (d) R1-Qwen3-8B, (e) Llama3.3-70B, and (f) Qwen3-32B-non-thinking.

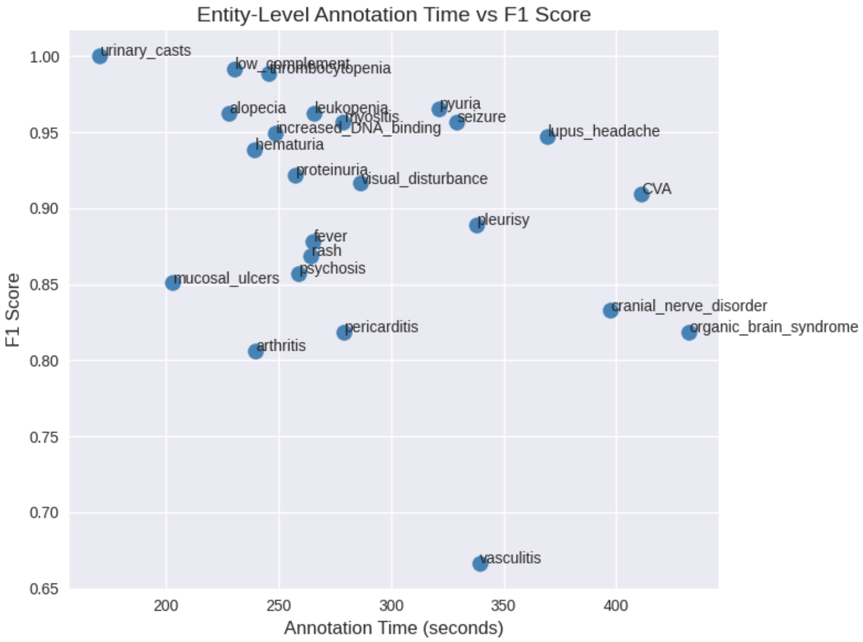

**Figure S9. Correlation plot between entity-level human annotation time vs model performance (GPT-OSS-120B model).** A scatter plot comparing average annotation time and model F1 performance for each entity type, evaluated on the full dataset of 658 cases. We observed a trend of moderate negative correlation between the average annotation time and F1 score across all descriptor entities (Pearson = -0.35, p = 0.098； Spearman = -0.37, p = 0.072). This suggests entities requiring a longer time for human annotation tend to be more challenging for the model prediction.

The average annotation time per entity type was computed by weighing the time proportionally to the count of each annotated entity type across all samples. For the entity type $e$, the average time was calculated as:

$$\mathrm{AvgTime}_e = \frac{\sum_{s \in S} \frac{C_{s,e}}{C_s} \cdot \mathrm{T_s}}{\sum_{s \in S} C_{s,e}}$$

Where $S$ is the set of all samples, $T_s$ is the annotation time for sample $s$, $C_{s,e}$ is the count of entities of type $e$ in sample $s$, and $C_s$ is the total number of annotated entities in sample $s$.

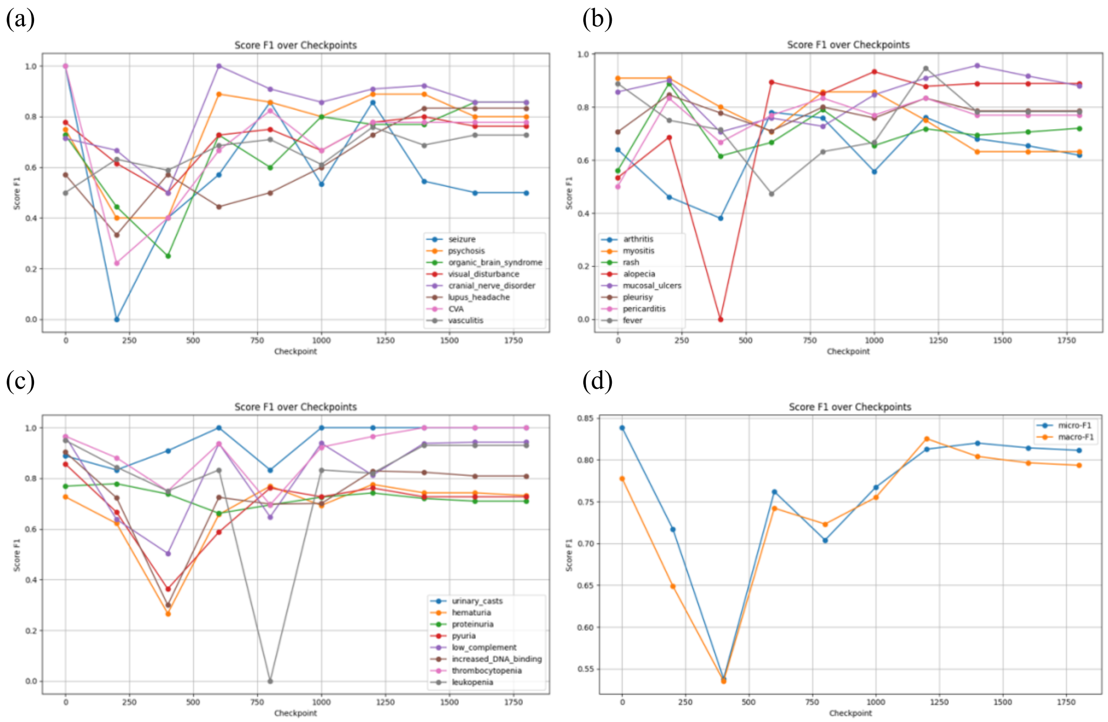

**Figure S10. GSPO performance across training steps.** Line plots illustrating model performance progression over the course of GSPO finetuning including (a) F1 scores for clinical descriptors with SLEDAI-2K weight of 8; (b) F1 scores for clinical descriptors with SLEDAI-2K weight < 8; (c) F1 scores for laboratory descriptors; (d) micro-F1 and macro-F1 scores across all descriptors. Overall performance reaches optimal at checkpoint 1200. F1 performance improved for 14 descriptors, where pericarditis, alopecia, and vasculitis improved over 50%; arthritis, psychosis, rash, cranial nerve disorder, and lupus headache improved over 25%; pleurisy and urinary casts over 10%; and hematuria, fever, mucosal ulcers, and organic brain syndrome over 5%. However, F1 performance decreased over 20% for CVA; over 10% for myositis, low complement, seizure, leukopenia, and pyuria; and over 5% for increased DNA binding.

**Table S6. Examples of model deduction processes of GPT-OSS-120B that yielded correct predictions for each descriptor.** Dates are masked to protect privacy.

| **descriptor** | **filename** | **LLM reasoning trace** |
|---|---|---|
| seizure | Abuhammad2024-F38-20231215.txt | **analysis**We need to evaluate.<br><br>First, check for diagnostic keywords: "seizure" appears: "c/o focal seizures". Also "EEG: focal temporal activity". Diagnostic criteria: need independent description of seizure by reliable witness OR EEG abnormalities. Here we have patient's complaint of focal seizures (subjective) and EEG shows focal temporal activity (abnormal). So criteria fulfilled likely. Need to check exclusions: any meds that cause seizures? quinolones, imipenem not present. Alcohol/drug withdrawal not mentioned. Electrolyte imbalance? not mentioned. organ failure? Not mentioned. infection? Not mentioned. Past irreversible CNS damage? Not mentioned. So none.<br><br>Relevant = true.<br><br>Now nature_of_intention_to_treat: Plan includes Pred 40mg OD (steroid) and HCQ and LEV (levetiracetam) seizure prophylaxis. This is escalation of lupus-specific treatment (Pred). So treat_escalated.<br><br>Now time: Note dated 15/12/2023. Mention "c/o focal seizures". No explicit timing, but seems current complaint. No "onset 3 months ago". So assume within_10days. Use default.<br><br>Thus criteria = fulfilled.<br><br>Score = 1 (since criteria fulfilled and time within_10days).<br><br>Confidence: High, because clear EEG abnormality and mention of seizures, and treatment escalated. I'd give 9 or 10. Choose 9 maybe because some could argue patient self-reported seizure may need witness but EEG provides. So high confidence. 9.<br><br>Now format JSON.<br><br>**assistantfinal**{<br>"descriptor": "seizure",<br>"relevant": true,<br>"criteria": "fulfilled",<br>"nature_of_intention_to_treat": "treat_escalated",<br>"time": "within_10days",<br>"score": 1, |

| | | |
|---|---|---|
| | | "confidence_score": 9,<br>"rationale": "The note reports \"c/o focal seizures\" and an EEG showing \"focal temporal activity,\" meeting the seizure diagnostic criteria (witness description and EEG abnormality). No exclusion factors are mentioned, and the treatment plan escalates lupus therapy with prednisone, confirming active management."<br>} |
| psychosis | Nayak2012-F20-20110115.txt | **analysis**We need to process the note. Keywords: "acute psychosis (auditory hallucinations, persecutory/referential delusions)" => psychosis keyword present, also symptoms hallucinations, delusions. So relevant true.<br><br>Now evaluate diagnostic criteria.<br><br>We have psychosis with auditory hallucinations and delusions (criterion A1 and A2). Need also B, C, D. No mention of distress/impairment, but likely causing impairment. Not exclusive to delirium, no delirium mentioned. Exclusions: steroid psychosis excluded (low dose prednisolone 30mg and onset pre-steroids). Substance/drug exclusion? No. Uremia etc not mentioned. So likely criteria fulfilled.<br><br>Check intention to treat: plan includes immunosuppression: prednisolone 30mg daily (increase? Already low-dose but it's being started). It is a lupus-specific treatment escalated (prednisolone used for NPSLE). So nature_of_intention_to_treat = treat_escalated.<br><br>Time: Note date is 15/01/2011. The complaint mentions acute psychosis (no exact date) but the duration of fatigue 3 months, psychosis seems acute now. No timeline given, but it's current as of note. According to guidelines, default to within_10days if no date but symptom appears current. So time = within_10days.<br><br>Score: criteria fulfilled, time within_10days => score 1.<br><br>Confidence: high confidence because explicit psychosis features, exclusion addressed, treatment escalation. I'd give confidence_score 9.<br><br>Now produce JSON.**assistantfinal**{<br>"descriptor": "psychosis",<br>"relevant": true,<br>"criteria": "fulfilled",<br>"nature_of_intention_to_treat": "treat_escalated",<br>"time": "within_10days",<br>"score": 1,<br>"confidence_score": 9,<br>"rationale": "The note documents acute psychosis with auditory hallucinations and persecutory delusions, meeting criterion A; no delirium or other exclusions are present, and the clinician plans to start prednisolone 30 mg, indicating treatment escalation for NPSLE. The symptom is described as |

| | | |
|---|---|---|
| | | current, so it is considered within 10 days."<br>} |
| organi c_brai n_synd rome | Samadder 2025-F25-20241215 .txt | **analysis**We need to parse note.<br><br>Descriptor organic_brain_syndrome relevant? Keywords: "Acute confusion" - does that count? The list includes "acute confusional state", "delirium", "encephalopathy". "Acute confusion" is similar but not exact keyword. However "NPSLE acute confusional status" includes "acute confusional". That is a diagnostic keyword (acute confusional state). So relevant = true.<br><br>Now evaluate criteria.<br><br>We have symptoms: "Acute confusion, mood swings, pressured speech, agitation." Also HPI: "2-week history of fluctuating behavior (irritability, confusion), with disorganized thoughts." PE: "Agitated, disorganized thought process."<br><br>We need A: disturbance in attention or awareness. Not explicitly mentioned but "confusion" and "disorganized thought process" imply reduced attention/awareness. The note says "Acute confusion" and "fluctuating behavior" and "disorganized thoughts". That suggests attention disturbance.<br><br>B: rapid onset and fluctuating over short period (hours-days). History 2 weeks of fluctuating behavior. That's longer than typical but still could be considered acute onset? 2 weeks is a bit longer than typical delirium (< days). But they call it acute confusional status. Might satisfy.<br><br>C: need at least two of: perceptual disturbance, incoherent speech, insomnia/daytime drowsiness, increased/decreased psychomotor activity. We have "pressured speech" and "agitation" (hyperactive). Pressured speech might count as incoherent speech? But incoherent speech explicitly: "disorganized thought process" possibly incoherent. So we have at least two: incoherent speech, hyperactive psychomotor.<br><br>Thus criteria likely fulfilled.<br><br>Exclusions: check metabolic disturbances. No glucose info beyond CSF glucose 2.7, serum glucose not given. Not high. Sodium not mentioned. No primary mental disorder. So no exclusions noted.<br><br>Thus diagnostic criteria: 'fulfilled'.<br><br>Nature of intention to treat: The plan includes IV methylprednisolone then oral pred 40 mg, cyclophosphamide initiation, HCQ addition. This is escalation of lupus-specific treatment (steroids, cyclophosphamide). So treat_escalated.<br><br>Time: The symptom onset? "2-week history of fluctuating behavior" active currently (confusion present). The note date 15/12/2024. Symptom ongoing for 14 days. That's >10 days, but <=30 days? Actually 14 days is within 11-30 days (since >10). So time = '11_to_30days'. |

| | | |
|---|---|---|
| | | Score: Both criteria fulfilled and time within 11-30days => score = 1.<br><br>Confidence: high, because clear mentions of acute confusional status, symptoms, treatment escalation. I'd give confidence 9 or 10. There's slight ambiguity about rapid onset criterion (2 weeks may be longer than typical delirium). But still considered acute confusional status; the physician labeled as such. So maybe 9.<br><br>We need JSON with single quotes? The example shows single quotes but typical JSON uses double quotes. We'll follow example: single quotes.<br><br>Now produce final answer with required fields.<br><br>**assistantfinal**{<br>'descriptor': 'organic_brain_syndrome',<br>'relevant': true,<br>'criteria': 'fulfilled',<br>'nature_of_intention_to_treat': 'treat_escalated',<br>'time': '11_to_30days',<br>'score': 1,<br>'confidence_score': 9,<br>'rationale': "The note explicitly states 'NPSLE acute confusional status' and describes acute confusion, disorganized thoughts, pressured speech, and agitation, meeting A-C of the organic brain syndrome criteria. Treatment is escalated with IV methylprednisolone, oral prednisone, and cyclophosphamide, confirming active management. Symptoms have been present for a 2-week (≈14 days) period, placing the time frame in the 11-to-30-day window."<br>} |
| visual_disturbance | Aldhefeery2023-M34-20230101.txt | **analysis**We need to parse note for visual disturbance. Keywords present: "painless bilateral vision loss ×3wk", "Bilateral CME, diffuse cotton wool spots, dot-blot hemorrhages". These are signs fitting retinal disease: cotton wool spots, hemorrhages. Diagnostic criteria: Retinopathy or choroidopathy with criteria A-D. Here we have cotton wool spots (cytoid bodies) and dot-blot hemorrhages (retinal hemorrhages). So criteria (1) is met: retinopathy with cotton-wool spots and hemorrhages. So criteria fulfilled.<br><br>Need relevance: true.<br><br>Nature of intention to treat: plan includes IV methylprednisolone then oral prednisone taper, plus HCQ, MMF. This is initiation/escalation of steroids. So treat_escalated.<br><br>Time: The vision loss is "painless bilateral vision loss ×3wk". Note dated 01/01/2023. 3 weeks ago from note date (~21 days). Within 10 days? It's |

| | | |
|---|---|---|
| | | within 30 days but >10 days. So time classification: 11_to_30days (active within 3 weeks). So time = '11_to_30days'.<br><br>Score: criteria fulfilled and time within 11_to_30days => score=1.<br><br>Confidence: high. Evidence is clear (cotton wool spots, hemorrhages, vision loss). So confidence maybe 9 or 10. Probably 9 because a bit of nuance but fairly certain. We'll give 9.<br><br>Now fill JSON.<br><br>Check exclusions: hypertension? BP normal. Infectious or drug? Not mentioned. MS family hx but not patient; but autoimmune ON excluded only if MS is patient cause. Not here. So no exclusions.<br><br>Thus criteria fulfilled.<br><br>Output JSON with required fields.**assistantfinal**{<br>"descriptor": "visual_disturbance",<br>"relevant": true,<br>"criteria": "fulfilled",<br>"nature_of_intention_to_treat": "treat_escalated",<br>"time": "11_to_30days",<br>"score": 1,<br>"confidence_score": 9,<br>"rationale": "The note describes painless bilateral vision loss for 3 weeks with objective findings of diffuse cotton-wool spots and dot-blot retinal hemorrhages, fulfilling the retinopathy criteria (cytoid bodies and hemorrhages). Treatment is escalated with IV methylprednisolone followed by oral prednisone, indicating active management. The symptom duration (~3 weeks) places it in the 11-to-30-day window, thus the descriptor scores 1."<br>} |
| cranial_nerve_disorder | Maniam2020-F47-20200410.txt | **analysis**We need to evaluate descriptor cranial_nerve_disorder.<br><br>Clinical note: Acute left eye diplopia + ptosis upon awakening 2 days. Neuro: Left CN III palsy (↓↓↓ & lateral eye deviation, nonreactive pupil, ptosis). Other CNs intact. Imaging negative. So this is a new onset focal motor neuropathy involving cranial nerve III. It meets criteria: oculomotor nerve palsy - ptosis, nonreactive pupil, etc. No exclusion (stroke, aneurysm etc excluded by CTA/MRA). So criteria fulfilled.<br><br>Relevant = true because diagnostic keywords: "CN III palsy", "palsy" etc. Also "isolated CN III palsy". So relevant true.<br><br>Nature of intention to treat: Management plan includes IV methylprednisolone 1g ×3d then oral prednisone 60 mg, restart HCQ. That's escalation of |

| | | |
|---|---|---|
| | | steroids (high dose) thus treat_escalated.<br><br>Time: The note is dated 10/04/2020. Onset: acute left eye diplopia + ptosis upon awakening 2 days. So symptom onset 2 days before note, within 10 days. So time = within_10days.<br><br>Score: criteria fulfilled + time within_10days => score =1.<br><br>Confidence: high, explicit mention of CN III palsy, no exclusions, clear treatment escalation. So confidence maybe 9 or 10. Use 10.<br><br>Now produce JSON with specified fields with single quotes? The example uses single quotes but JSON normally double quotes, but they want exactly as format? The example uses single quotes. We'll follow that.<br><br>Return with reasoning tags then answer.**assistantfinal**<reasoning><br>The note describes a new onset left CN III palsy (ptosis, nonreactive pupil, diplopia) documented 2 days ago, which directly matches the cranial nerve disorder criteria for the oculomotor nerve. Imaging excludes stroke/aneurysm, so no exclusion applies. The treatment plan escalates high-dose steroids and restarts HCQ, indicating “treat_escalated”. The symptom started within 2 days of the note date, thus time = within_10days. All conditions for a score of 1 are met, and the evidence is explicit, giving maximal confidence.<br></reasoning><br><answer><br>{<br>'descriptor': 'cranial_nerve_disorder',<br>'relevant': true,<br>'criteria': 'fulfilled',<br>'nature_of_intention_to_treat': 'treat_escalated',<br>'time': 'within_10days',<br>'score': 1,<br>'confidence_score': 10,<br>'rationale': "The note reports an acute left CN III palsy (ptosis, nonreactive pupil, diplopia) onset 2 days ago, with CTA/MRA negative for vascular causes, and the plan includes high-dose IV methylprednisolone then oral prednisone, representing treatment escalation."<br>}<br></answer> |
| lupus_headache | Rajasekharan2012-F14- | **analysis**We need to parse the note. Determine relevance: keywords list includes "lupus headache", "headache", "cephalagias", "migraine". The note has "bilateral throbbing hemicranial headache×4 weeks". So "headache" present, thus relevant = true.<br><br>Now assess diagnostic criteria for lupus_headache descriptor: It's a specific type of headache in lupus? The diagnostic criteria provided: severe |

| | | |
|---|---|---|
| | 20120415 .txt | disabling, persistent lasting ≥3 days, nonresponsive to narcotics, and types A-E (migraine, tension, cluster, intracranial hypertension, intractable). Note says "bilateral throbbing hemicranial headache×4 weeks—worse on standing + transient vision blackouts". Also papilledema, high CSF pressure, MRI signs, suggests intracranial hypertension (pseudotumor cerebri). That matches type D: Headache from intracranial hypertension. However need to see if meets all criteria: increased intracranial pressure (opening pressure >300 mmH2O) yes; normal neuro exam except papilledema and possible nerve VI palsy - we have bilateral papilledema and abducens weakness, which is VI palsy (ok). No mass lesion on imaging (MRI shows posterior scleral flattening and CSF herniation into sella; no mass). Normal CSF protein (protein 14.4, presumably normal), normal white cell (0 cells). No venous thrombosis (MRV no thrombosis). So criteria for intracranial hypertension are fulfilled.<br><br>Exclusions: Need to check for any exclusion: CNS infection, tumors, low pressure, trauma, metabolic, drug-induced pseudotumor cerebri, etc. Not present. So diagnostic criteria for lupus_headache are fulfilled (intracranial hypertension subtype).<br><br>But also need to consider intention to treat: The plan includes methylprednisolone, PSL, cyclophosphamide, HCQ – these are lupus-specific immunosuppressants. That's escalation (treat_escalated). So nature_of_intention_to_treat = treat_escalated.<br><br>However note: The impression is "Idiopathic intracranial hypertension". That's not necessarily lupus headache but the patient has SLE labs (ANA+, dsDNA high). So the headache is considered idiopathic intracranial hypertension; but within lupus context, perhaps still qualifies as lupus_headache descriptor given criteria. The instruction: "If diagnostic keywords are mentioned or criteria met, but nature_of_intention_to_treat is not treat_escalated, then criteria should be considered unfulfilled_diagnostic." Here we have treat_escalated, so okay.<br><br>Now time: The note says headache ×4 weeks. The note date is 15/04/2012. So headache has been present for 4 weeks, which is 28 days, within 11_to_30days (since 4 weeks = 28 days). According to time classification: within_10days if within last 10 days; 11 to 30 days if more than 10 days but less than 30 days. So time = '11_to_30days'.<br><br>Thus criteria = fulfilled, time = 11_to_30days, score = 1.<br><br>Confidence: high, clear evidence. I'd give confidence 9 or 10. However note the descriptor is lupus_headache which may require severe disabling, persistent etc. The headache is described as bilateral throbbing hemicranial lasting 4 weeks, seems persistent. Not mention narcotic responsiveness but irrelevant for intracranial hypertension type. Since criteria are satisfied, confidence high. I'd assign 9.<br><br>Now need to output JSON with fields.<br><br>Relevant true, criteria 'fulfilled', nature_of_intention_to_treat 'treat_escalated', time '11_to_30days', score 1, confidence_score 9, rationale with quoted text.<br><br>Let's craft. |

| | | |
|---|---|---|
| | | **assistantfinal**{<br>"descriptor": "lupus_headache",<br>"relevant": true,<br>"criteria": "fulfilled",<br>"nature_of_intention_to_treat": "treat_escalated",<br>"time": "11_to_30days",<br>"score": 1,<br>"confidence_score": 9,<br>"rationale": "The note documents a \"bilateral throbbing hemicranial headache×4 weeks\" with opening pressure >300 $mmH_2O$, papilledema, abducens weakness, normal CSF cells and protein, and no mass lesion or venous thrombosis, meeting the intracranial hypertension subtype of lupus headache. Treatment is escalated with high-dose steroids, cyclophosphamide and HCQ. The duration (~4 weeks) places the symptom in the 11-to-30-day window, supporting a score of 1."<br>} |
| CVA | Li2023-F25-20120220.txt | **analysis**We need to assess descriptor CVA (stroke). The note: "progressive right limb weakness ×2mo, acutely worsened ×3d with new R leg weakness + left facial deviation and headache." Also prior "lacunar infarct". Imaging: Brain MRI: Recent L basal ganglia infarct (DWI+). So there is evidence of stroke. The diagnostic criteria for CVA: stroke syndrome acute focal deficit persisting >24h, supported by imaging abnormality. Here we have acute worsening over 3 days with new deficits, imaging shows recent infarct. So fulfilled.<br><br>We must check exclusions: list includes drug-induced pseudotumor, infection, tumor, trauma, etc. No mention. So not excluded. The note says Dx: NPSLE, lupus nephritis, anemia. So they attribute it to neuropsychiatric SLE. That is fine.<br><br>Now relevant = true because keywords like "infarct", "hemiparesis", "facial deviation", "stroke"? The note uses "lacunar infarct". Not the word stroke but still relevant. The guideline says check diagnostic keywords list: stroke, subarachnoid, hemorrhage, cerebral venous, thrombosis, TIA, etc. "infarct" is not in list but it's a stroke. However we have signs like hemiparesis, unilateral weakness. Those are symptom/sign keywords: hemiplegia, unilateral weakness. So relevant true.<br><br>Nature_of_intention_to_treat: Management Plan includes IV methylprednisolone, prednisone, HCQ, MMF, warfarin. This is escalation of lupus treatment (steroids, immunosuppression). This qualifies as treat_escalated (new high dose steroid, adding MMF). So treat_escalated.<br><br>Time: The symptom onset progressive over 2 months, acutely worsened 3 days ago. The note: progressive right limb weakness 2 months, acutely worsened 3 days with new deficits. The present deficits are active now (the exam shows deficits). The timing: since acutely worsened 3 days ago, within 10 days. So time = within_10days.<br><br>Thus criteria: fulfilled. |

| | | |
|---|---|---|
| | | Score = 1 (criteria fulfilled and time within_10days).<br><br>Confidence: high, clear imaging and deficits. I would give confidence 9 or 10. Might be 10 because explicit. Use 10.<br><br>Now produce JSON.**assistantfinal**{<br>"descriptor": "CVA",<br>"relevant": true,<br>"criteria": "fulfilled",<br>"nature_of_intention_to_treat": "treat_escalated",<br>"time": "within_10days",<br>"score": 1,<br>"confidence_score": 10,<br>"rationale": "The note documents acute focal neurologic deficits (right hemiparesis, left facial deviation) persisting >24 h with MRI showing a recent basal ganglia infarct, satisfying stroke criteria. No exclusionary causes are mentioned, and treatment is escalated with high-dose steroids, MMF and warfarin, indicating active management of NPSLE. The acute worsening occurred 3 days ago, placing it within the 10-day window."<br>} |
| vasculitis | KUK2014-F45-20150101.txt | **analysis**We need to process the note.<br><br>First, check for diagnostic keywords: "cutaneous vasculitis", "vasculitic rash", etc. The note says "SLE Flare with Vasculitic Digital Ulcers". Not exactly "cutaneous vasculitis", but "vasculitic" and "digital ulcers" which are skin lesions. The descriptor is vasculitis. The note mentions "Painful necrotic ulcers R index/4th fingers x2w". That's necrotic ulcers on fingers, which are skin lesions. Also "Necrotic ulcers(R index/4th fingers, dry eschar)". This could be considered cutaneous vasculitis with ulceration.<br><br>Diagnostic criteria: Either clinician observed cutaneous vasculitis with at least one described lesion: ulceration, gangrene, etc. Here we have necrotic ulcers, which qualifies as skin ulceration. So A is fulfilled.<br><br>Exclusions: The note includes a negative for APS, negative ANCA, no infection, etc. So no exclusions.<br><br>Thus criteria = fulfilled.<br><br>Relevance: keywords present => true.<br><br>Nature of intention to treat: The plan includes Cyclophosphamide 150mg daily x6mo (an immunosuppressant for SLE vasculitis), plus Nifedipine, etc. This is escalation of therapy (adding cyclophosphamide). So nature_of_intention_to_treat = treat_escalated. |

| | | |
|---|---|---|
| | | Time frame: The note says "Painful necrotic ulcers R index/4th fingers x2w." Means for 2 weeks. Also "SLE Flare with Vasculitic Digital Ulcers". So currently present, within 2 weeks => within_10days? 2 weeks = 14 days, which is >10 days. The guidelines: classify as '11_to_30days' if active more than 10 days ago but less than 30 days ago. Since 2 weeks (14 days) is within that range. So time = '11_to_30days'.<br><br>Score: criteria fulfilled and time is in allowed range (within_10days or 11_to_30days). So score = 1.<br><br>Confidence: High, as clear evidence. I'd give confidence 9 or 10. There is explicit mention of necrotic ulcers, qualifies as skin ulceration. The time frame is clear: 2 weeks. So 9 or 10. I'll give 9 (some minor uncertainty about exact days). But note "x2w" meaning 2 weeks, which is 14 days. So definitely within 11-30 days. So confident.<br><br>Rationale: mention necrotic ulcers (skin ulceration) fulfilling criteria, no exclusions, treatment escalation, time 2 weeks => score 1.<br><br>Now produce JSON.<br><br>**assistantfinal**{<br>"descriptor": "vasculitis",<br>"relevant": true,<br>"criteria": "fulfilled",<br>"nature_of_intention_to_treat": "treat_escalated",<br>"time": "11_to_30days",<br>"score": 1,<br>"confidence_score": 9,<br>"rationale": "The note documents painful necrotic ulcers on the right index and fourth fingers ('skin ulceration'), meeting the cutaneous vasculitis criterion. No exclusions (infection, APS, drugs) are noted. Treatment is escalated with cyclophosphamide, indicating active management. The ulcers have been present for ~2 weeks ('x2w'), placing the event in the 11-to-30-day window, thus the descriptor scores 1."<br>} |
| arthritis | Narang2022-M20-20220101.txt | **analysis**We need to parse the note. Look for arthritis keywords. Words: "Bilateral wrist synovitis at presentation." Synovitis is a paraclinical keyword (synovitis). Also "Bilateral wrist synovitis (tender, swelling)." That's arthritis descriptor. So relevant = true.<br><br>Now assess criteria: Need either A: two or more joints with synovitis, pain, signs (tenderness, swelling, effusion). Document mentions bilateral wrist synovitis — that's two joints (both wrists) showing tenderness and swelling. So meets criteria A: two or more joints with synovitis signs. So criteria 'fulfilled'.<br><br>Check exclusions: any mention of infection causing joints? No. No gout, osteoarthritis. So no exclusions. |

| | | |
|---|---|---|
| | | Now time: note dated 01/01/2022. HPI says "Bilateral wrist synovitis at presentation." The present is the current visit. In the same note, "Prior Tx: Indomethacin...→ symptom resolution. Later started prednisone 20mg daily." But at presentation there is synovitis. So it's current. No explicit onset time. So assumed active now. So time classification: within_10days (active currently). So time = within_10days.<br><br>Thus score = 1 (criteria fulfilled and time within 10 days). Confidence: high. The note clearly mentions bilateral wrist synovitis with tenderness and swelling, two joints, meets criteria. So confidence maybe 9 or 10. Use 10.<br><br>Now produce JSON.<br><br>**assistantfinal**<reasoning><br>The note contains the phrase **"Bilateral wrist synovitis (tender, swelling)"**, which includes the diagnostic keyword "synovitis" and mentions two joints (both wrists) with tenderness and swelling. This satisfies criterion A (≥2 joints with synovitis and signs of inflammation). No exclusions (infection, gout, OA, etc.) are noted. The finding is described at the current presentation, so the timing is current – classified as **within_10days**. Therefore the descriptor is relevant, criteria are **fulfilled**, and the score should be **1**. The evidence is explicit, giving full confidence.<br></reasoning><br><answer><br>{<br>"descriptor": "arthritis",<br>"relevant": true,<br>"criteria": "fulfilled",<br>"time": "within_10days",<br>"score": 1,<br>"confidence_score": 10,<br>"rationale": "The note states 'Bilateral wrist synovitis (tender, swelling)', indicating two joints with synovitis and inflammatory signs, meeting the arthritis criteria, and it is described as present at the current visit."<br>}<br></answer> |
| myositis | Roy2022-F51-20180315.txt | **analysis**We need to evaluate myositis descriptor.<br><br>First, check relevance: Are keywords present? Yes, "myositis" appears in impression, also "muscle weakness", "EMG", biopsy. So relevant = true.<br><br>Now diagnostic criteria:<br><br>A. proximal muscle aching/myalgias/weakness: The note says "mild proximal muscle weakness (shoulders/hips) – peaked over past 2 years" and PE |

| | | |
|---|---|---|
| | | shows moderate symmetric weakness in neck, shoulders, hips. So criterion A met.<br><br>B. associated with elevated CK/aldolase or EMG changes or biopsy showing myositis: CK (CPK) is normal, aldolase normal. EMG shows fibrillations and myopathic units => EMG changes consistent. Biopsy shows endomysial CD20+ B-cell aggregates, MHC upregulation, described as "B-cell predominant myositis". So EMG changes or biopsy fulfilling criteria. So criterion B met.<br><br>Thus both A and B met. So criteria fulfilled.<br><br>Exclusions: Check for CK elevation explanations, statin, hydroxychloroquine myotoxicity, cancer, metabolic muscle disease. CK is normal, so no CK elevation. No mention of statin, no cancer. So no exclusions.<br><br>Therefore criteria = fulfilled.<br><br>Now time frame: Need to decide if symptom active within 10 days or 11-30 days. Note says progressive exertional dyspnea and mild proximal muscle weakness peaked over past 2 years, worst at presentation. The note is current presentation; the weakness is present now. There's no specific timeline such as "last 2 weeks". The mention of progressive over 2 years suggests ongoing. Since it's present at presentation (the note date 15/03/2018), we treat as current active. According to guidelines: if symptom ongoing and no resolution, consider within_10days default unless indication otherwise. So time = within_10days.<br><br>Thus score = 1 since criteria fulfilled and time within_10days.<br><br>Confidence: high (clear evidence). I'd give confidence 9 or 10. Since all criteria directly documented, no ambiguity. I'd set 10.<br><br>Rationale: mention that proximal weakness present and EMG findings and biopsy confirm myositis, active now.<br><br>Now construct JSON.<br><br>**assistantfinal**<reasoning><br>The note mentions “myositis” explicitly and describes proximal muscle weakness plus objective EMG findings and a muscle biopsy consistent with myositis. Both diagnostic components (A = proximal weakness, B = EMG changes/biopsy) are satisfied, and no exclusion (e.g., CK elevation due to other causes, statin use, cancer) is noted. The weakness is described as present at the current visit (“worst at presentation”), so the manifestation is active now, which maps to the “within_10days” time category. Therefore the descriptor meets the criteria and is temporally active, yielding a score of 1. The evidence is explicit and unambiguous, justifying a high confidence level.<br></reasoning><br><answer> |

| | | |
|---|---|---|
| | | {<br>"descriptor": "myositis",<br>"relevant": true,<br>"criteria": "fulfilled",<br>"time": "within_10days",<br>"score": 1,<br>"confidence_score": 10,<br>"rationale": "The note documents proximal muscle weakness (shoulders/hips) and EMG fibrillations with myopathic units plus a biceps biopsy showing B-cell predominant myositis, fulfilling both A and B criteria; the weakness is present at the current visit, so it is active within 10 days."<br>}<br></answer> |
| urinary_casts | pseudo026-F51-[masked].txt | **analysis**We need to evaluate urinary_casts descriptor. Diagnostic info: Heme-granular or RBC casts. Keywords: heme-granular casts, RBC casts.<br><br>Clinical note has: "Urine for RBC +++, Protein +++, red blood cell casts," So RBC casts present. That is diagnostic keyword present, so relevant = true.<br><br>Now criteria: Need to see if criteria fulfilled. Presence of RBC casts indicates positive. No exclusion. So criteria fulfilled.<br><br>Now time: The note date is [masked]. There's no explicit time for the casts, but it's part of current urine findings. Usually it's current labs. So assume active now => within_10days. No indication of timing being older. So time = within_10days.<br><br>Thus score = 1 (criteria fulfilled and time within_10days). Confidence: High, clear statement, so maybe 9 or 10. Let's give 9? It's explicit. No ambiguity. So 10.<br><br>Return JSON with fields.<br><br>**assistantfinal**<reasoning><br>The note explicitly mentions "red blood cell casts," which matches the keyword list (RBC casts). This makes the descriptor relevant. The presence of RBC casts directly fulfills the diagnostic criterion for urinary_casts; no exclusions are noted. The lab result is presented as part of the current assessment, implying it is recent (active at the time of the note), so the timing is classified as within_10days. Therefore, the score should be 1. The evidence is clear and unambiguous, giving high confidence.<br></reasoning><br><answer><br>{<br>"descriptor": "urinary_casts", |

| | | |
|---|---|---|
| | | "relevant": true,<br>"criteria": "fulfilled",<br>"time": "within_10days",<br>"score": 1,<br>"confidence_score": 10,<br>"rationale": "The note states 'red blood cell casts,' directly matching the keyword for urinary_casts and indicating current presence; no exclusions are noted, and the finding is part of the current lab results, so it is considered active within the last 10 days."<br>}<br></answer> |
| hematuria | Hao2021-F28-20140620.txt | analysisWe need to evaluate hematuria descriptor. Identify keywords: "U/A: blood 3+" indicates urine blood presence. Also "non-blanching dermohemia hands/feet" is skin bleeding, not urine. The note: "U/A: blood 3+, protein 3+".<br><br>Check criteria: need >5 RBC/hpf or urine RBC >=2+ etc. They have "blood 3+" on UA. That qualifies as Urine RBC >=2+ (blood 3+). So criterion A met.<br><br>Now need also abnormal urine protein: they have protein 3+ on dipstick; that fulfills criterion B. So both A and B are met => fulfilled.<br><br>Exclusions: no mention of stones, infection, menstruation causing urine blood. So no exclusion.<br><br>Now time frame: The note is dated 20/06/2014. The labs are from Jun 8 (labs). That's 12 days before note date. The descriptor "blood 3+" is from labs on Jun 8. Since note date is Jun 20, the finding is 12 days ago. Time classification: within_10days is up to 10 days. 11_to_30days is >10 days up to 30 days. This is 12 days => falls into 11_to_30days.<br><br>Thus criteria fulfilled, time = 11_to_30days, score = 1.<br><br>Confidence: high because clear evidence. So confidence 9 or 10. Maybe 10 because explicit.<br><br>Return JSON with fields.<br><br>Check relevance: keywords present => relevant true.<br><br>Criteria: 'fulfilled'.<br><br>Time: '11_to_30days'. |

| | | |
|---|---|---|
| | | Score: 1.<br><br>Confidence_score: 10.<br><br>Rationale: "U/A shows blood 3+ and protein 3+ on 08/06/2014, meeting hematuria and protein criteria; no exclusions noted; lab done 12 days prior to note, within 11-30 days, so active."<br><br>Provide JSON with correct quoting using single quotes? In example they used single quotes. Use same.<br><br>**assistantfinal**<reasoning><br>The clinical note contains the keyword "blood 3+" in the urinalysis (U/A) on 08/06/2014, indicating urine RBC ≥2+. It also shows "protein 3+" on dipstick, satisfying the abnormal urine protein requirement. No exclusions (stones, infection, menstruation) are mentioned. Thus both A and B criteria are met → 'fulfilled'. The lab result is dated 08/06/2014, 12 days before the note date (20/06/2014), placing the finding in the "11_to_30days" time window. Therefore the descriptor should be scored 1. The evidence is explicit and unambiguous, giving high confidence.<br></reasoning><br><answer><br>{<br>'descriptor': 'hematuria',<br>'relevant': true,<br>'criteria': 'fulfilled',<br>'time': '11_to_30days',<br>'score': 1,<br>'confidence_score': 10,<br>'rationale': "Urinalysis on 08/06/2014 shows blood 3+ (≥2+ RBC) and protein 3+, meeting both hematuria and protein criteria; no exclusion noted; result is 12 days before note date, classified as 11-30 days, so active."<br>}<br></answer> |
| protein uria | 128-F54-[masked].txt | **analysis**We need to parse note. Look for proteinuria keywords: "urine protein ++", "urine protein/Cr 5.24", "24 hours urine TP 3.12". So relevant = true.<br><br>Now evaluate diagnostic criteria. Need either 24-hr urine >0.50 gram/24h, or urine protein/creatinine >0.50 mg/mg (same as 0.5). The note gives:<br><br>- "urine protein/Cr 5.24 (daily urine protein 2-4g for years)". This is presumably ratio; units unspecified, but value 5.24 >0.5. So fulfilled.<br><br>- "24 hours urine TP 3.12". Units not given, likely grams per 24h? Probably >0.5 grams. So fulfilled. |

| | | |
|---|---|---|
| | | Thus criteria fulfilled.<br><br>Now time. Note date [masked]. No date specific for these lab results. They are described in current note without a date. Likely recent labs (most recent values). So assume within 10 days? The guidelines: If no date, default to within_10days unless indications suggest otherwise. So time = within_10days.<br><br>Thus score = 1 (criteria fulfilled, time within_10days). Confidence high.<br><br>Now need to ensure no exclusion statements: none. There is "urine protein ++" dipstick but also ratio.<br><br>Thus final JSON.<br><br>Confidence maybe 9 or 10. I'd give 10 because clear numeric values > threshold, time default to within_10days.<br><br>Provide rationale.<br><br>Now output.**assistantfinal**<reasoning><br>The note contains multiple proteinuria keywords: “urine protein ++”, “urine protein/Cr 5.24”, and “24 hours urine TP 3.12”. Numeric values are provided for both the urine protein/creatinine ratio (5.24) and the 24-hour urine total protein (3.12). Both exceed the diagnostic thresholds (>0.50). No exclusions are mentioned. The lab values are presented as part of the current assessment with no specific older date, so per the guidelines the default time frame is **within_10days**. Therefore the descriptor meets the SLE proteinuria criterion and is recent, giving a score of 1 with high confidence.<br></reasoning><br><answer><br>{<br>"descriptor": "proteinuria",<br>"relevant": true,<br>"criteria": "fulfilled",<br>"time": "within_10days",<br>"score": 1,<br>"confidence_score": 10,<br>"rationale": "The note reports urine protein/Cr = 5.24 and 24-hour urine protein = 3.12, both well above the >0.50 threshold, with no exclusion noted; labs are presented as current, so time is within 10 days."<br>}<br></answer> |

| | | |
|---|---|---|
| pyuria | 128-F54-[masked].txt | **analysis**We need to evaluate descriptor pyuria.<br><br>First, look for keywords: pyuria, urine WBC, UWBC, urine WC. The note includes "urine WBC 10-100 no growth". That's a keyword.<br><br>Thus relevant = true.<br><br>Now assess diagnostic criteria: Need >5 white blood cells/high-power field AND abnormal urine protein (dipstick positive (UP+ or above) or UPC >0.15 etc.) Also exclude infection. Also infection if antibiotics etc.<br><br>We have urine protein ++ (dipstick positive) (the note says "urine protein ++ sugar -"). That's abnormal protein.<br><br>Urine WBC 10-100 no growth. That qualifies as >5 WBC/hpf (they consider WBC >=10-50/uL as >5 WBC/hpf). So both criteria present.<br><br>Now need to consider infection exclusion. The note mentions "Repeated UTI". Also "urine WBC 10-100 no growth". That suggests there is a history of UTIs, but current culture no growth. Does that count as infection? The criteria says Exclude infection. If doctor suspects infection, treat with antibiotics, then pyuria is likely due to infection and should not count. Here they note "Repeated UTI" (history). Not clear if current infection present. They have no growth. No mention of antibiotics being started for this pyuria. So we may consider infection not confirmed, so we can still score as pyuria.<br><br>Thus criteria satisfied -> 'fulfilled'.<br><br>Now time frame: Need to determine when this pyuria is noted. The note is dated [masked]. The urinalysis is part of current labs (they list). No explicit timing, but it's presumably current (as labs). So time is within_10days.<br><br>Thus score = 1.<br><br>Confidence: high because explicit >5 WBC, abnormal protein, no infection. So confidence maybe 9 or 10. There is mention of "Repeated UTI" which could be a confounder, but still no growth now. So moderate high. I'd give confidence 8.<br><br>Now construct JSON. Note field names: descriptor, relevant, criteria, time, score, confidence_score, rationale.<br><br>Rationale: mention urine WBC 10-100, urine protein ++, no growth, no current infection.<br><br>Let's produce answer.**assistantfinal**<reasoning><br>The note contains the keyword “urine WBC 10-100” and “urine protein ++”, satisfying the pyuria definition (>5 WBC/hpf + abnormal protein). No |

| | | |
|---|---|---|
| | | current infection is documented (culture shows “no growth”) and no antibiotics are mentioned for this finding, so the infection exclusion does not apply. The labs are presented as part of the current assessment, implying they are recent (within the note date), thus the time frame is within_10days. Therefore the diagnostic criteria are **fulfilled** and the descriptor is active, giving a score of 1. Confidence is high because the required lab values are explicit and infection is excluded.<br></reasoning><br><answer><br>{<br>"descriptor": "pyuria",<br>"relevant": true,<br>"criteria": "fulfilled",<br>"time": "within_10days",<br>"score": 1,<br>"confidence_score": 8,<br>"rationale": "The note reports 'urine WBC 10-100' (≥5 WBC/hpf) and 'urine protein ++' (dipstick positive) with 'no growth' on culture, indicating pyuria without evidence of infection; labs are presented in the current encounter, so the finding is recent."<br>}<br></answer> |
| rash | Kato2005-F46-20030201.txt | **analysis**We need to parse note. Identify rash keywords.<br><br>Note includes: "Cutaneous flare (↑ erythema) → prednisolone ↑ to 40mg qd" and "PE lupus erythema, no ulceration". Also "rash"? Not directly but "Cutaneous flare (↑ erythema)" is a diagnostic keyword? It's in list: "cutaneous flare". That is diagnostic keyword. So relevant = true.<br><br>Now need to assess criteria: Is there evidence of lupus rash? The note says "Cutaneous flare (↑ erythema)" which suggests increased erythema, likely active cutaneous lupus. Also "lupus erythema" in PE. However, we need a specific lupus rash classification (ACLE, SCLE, CCLE). The note says "lupus erythema" - ambiguous maybe just erythema. But "cutaneous flare" is considered diagnostic keyword that confirms active cutaneous lupus manifestation fulfilling criteria? In guidelines: "The term 'flare' in clinical impressions acts as a diagnostic trigger: it confirms active manifestations fulfilling specific SLEDAI descriptors (e.g., cutaneous flare fulfills lupus rash)." So "Cutaneous flare" qualifies as meeting the descriptor: fulfilled.<br><br>Thus criteria = fulfilled.<br><br>Time: The note date is 01/02/2003. The cutaneous flare occurred Dec 30 2002 (as per note). That's about a month earlier (31 days? Dec 30 to Feb 1 is approx 33 days). The note says "Cutaneous flare (↑ erythema) → prednisolone ↑ to 40mg qd → After 1mo: Hypomania..." So flare was noted Dec 30 2002. The note is Feb 1 2003. That's slightly more than 30 days ago (around 33 days). Time categories: '30days_ago' is for entity present 30 days ago. Might be considered beyond 30 days? They have categories: within_10days, 11_to_30days, 30days_ago (entity was present 30 days ago), time_uncertain. The flare was about 33 days ago, >30 days, maybe it's outside the categories, maybe time_uncertain. But note: "Cutaneous flare (↑ |

| | | |
|---|---|---|
| | | erythema) → prednisolone ↑ to 40mg qd" Not specify resolution; seems still active? They increased steroids after flare; now on Feb 1 they are tapering from 30mg → goal ≤20 mg, monitor skin/SLE activity. That suggests the rash may still be present? The phrase "monitor skin/SLE activity" implies ongoing activity. The initial flare was Dec 30. The steroid increase likely controlled rash? Not explicit. But "lupus erythema, no ulceration" in PE suggests currently there is lupus erythema present at exam during this visit (Feb 1). So rash is present currently, i.e., within 10 days. Since PE notes "lupus erythema". So time = within_10days.<br><br>Thus criteria fulfilled, time within_10days, score =1.<br><br>Confidence: high, because explicit diagnostic keyword "cutaneous flare" and exam shows "lupus erythema". So confidence maybe 9.<br><br>Now produce JSON with fields.<br><br>relevant true, criteria fulfilled, time within_10days, score 1, confidence 9, rationale.<br><br>**assistantfinal**{<br>"descriptor": "rash",<br>"relevant": true,<br>"criteria": "fulfilled",<br>"time": "within_10days",<br>"score": 1,<br>"confidence_score": 9,<br>"rationale": "The note documents a “Cutaneous flare (↑ erythema)” and a physical exam finding of “lupus erythema,” both diagnostic keywords indicating an active lupus rash. The exam was performed on the note date (01/02/2003), confirming current presence, thus the time frame is within 10 days."<br>} |
| alopecia | 116-F38-[masked].txt | **analysis**We need to follow instructions. Clinical note: "Noted patchy alopecia and occ joint pain". Contains alopecia keyword. So relevant = true.<br><br>Now assess criteria: diagnostic criteria for alopecia: hair loss or alopecia. The note says "Noted patchy alopecia". That's evidence. Need to consider exclusions: alopecia areata, iron deficiency, androgenic alopecia, scarring alopecia, drug-induced alopecia. No mention of these exclusions. So criteria likely fulfilled.<br><br>Time: note date [masked]. No timing info about when alopecia occurred. The phrase "Noted patchy alopecia" suggests it's observed now (current). No mention of resolved. So assume within_10days (default). According to guidelines, default to within_10days unless indications otherwise.<br><br>Thus criteria fulfilled, time within_10days -> score=1. |

| | | |
|---|---|---|
| | | Confidence: high, as explicit mention of alopecia, no exclusions, current observation. I'd give 9 or 10. Considering maybe uncertainty about chronicity? But we have clear evidence. Confidence 9.<br><br>Provide JSON with fields.<br><br>Also rational: mention noted patchy alopecia, no exclusions, presumed current.<br><br>Now output.<br><br>**assistantfinal**<reasoning><br>The note contains the keyword “alopecia” (“Noted patchy alopecia”), so the descriptor is relevant.<br>The statement directly confirms the presence of hair loss, which satisfies the diagnostic criterion (hair loss or alopecia). No exclusions (e.g., alopecia areata, iron deficiency, androgenic, scarring, drug-induced) are mentioned.<br>No timing details are provided, and the finding is described as currently “noted,” which under the guidelines defaults to **within_10days**.<br>Therefore the criteria are **fulfilled**, the time window is acceptable, and the descriptor should be scored 1. Confidence is high because the evidence is explicit and unambiguous.<br></reasoning><br><answer><br>{<br>"descriptor": "alopecia",<br>"relevant": true,<br>"criteria": "fulfilled",<br>"time": "within_10days",<br>"score": 1,<br>"confidence_score": 9,<br>"rationale": "The note states 'Noted patchy alopecia', directly confirming hair loss without any listed exclusions, and the observation is current (defaulted to within_10days)."<br>}<br></answer> |
| mucosal_ulcers | 022-F49-[masked].txt | **analysis**We need to parse note for mucosal_ulcers. Keywords: "A few oral ulcer". That's relevant. So relevant = true.<br><br>Now assess criteria: descriptor is oral ulcer observed by clinician. The note says "A few oral ulcer". That's a symptom reported by patient? It appears in the note: "? aloplecia" then "A few oral ulcer". It's likely observed? It's mentioned in assessment, not sure if clinician observed. It is in note: after "? aloplecia" line: "A few oral ulcer". Likely clinician noted. No mention of exclusions (vasculitis, Behcet's, infection). No mention of herpes infection. |

| | | |
|---|---|---|
| | | So criteria likely fulfilled? However need to consider whether it's sufficient: "Oral or nasal ulceration observed by a clinician". Yes. So 'fulfilled'.<br><br>Time: note date [masked]. The ulcer is described in present tense "A few oral ulcer". No time frame given. Default to within_10days unless indicated otherwise. There's no timeframe, so assume current. So time = within_10days.<br><br>Score: since criteria fulfilled and time within 10days, score = 1.<br><br>Confidence: high. The note clearly states "A few oral ulcer". It's present. So confidence maybe 9.<br><br>Provide JSON.<br><br>**assistantfinal**<reasoning><br>The note contains the phrase **“A few oral ulcer”**, which directly matches the diagnostic keywords for mucosal ulcers (oral ulceration). This indicates the clinician observed oral ulcers, satisfying the diagnostic criterion. No exclusions (vasculitis, Behcet’s, infection, etc.) are mentioned. The timing is not specified, but the descriptor is presented in the current assessment, so per guidelines we default to **within_10days**. Therefore the criteria are **fulfilled** and the time window is appropriate, leading to a score of **1**. Confidence is high because the evidence is explicit and unambiguous.<br></reasoning><br><answer><br>{<br>"descriptor": "mucosal_ulcers",<br>"relevant": true,<br>"criteria": "fulfilled",<br>"time": "within_10days",<br>"score": 1,<br>"confidence_score": 9,<br>"rationale": "The note states \"A few oral ulcer,\" indicating clinician-observed oral ulceration with no listed exclusions; timing is current (default within_10days)."<br>}<br></answer> |
| pleurisy | Narang2022-M20-20220101.txt | **analysis**We need to parse note.<br><br>First, relevance: keywords list includes serositis, pleurisy, pleuritis. Also symptoms: pleuritic chest pain, pleural rub, etc. Note includes "Acute pleuritic chest pain, SOB." That's a symptom keyword (pleuritic chest pain). Also CT: "12mm PE + bilateral pleural effusions." That's pleural effusion imaging. So relevant = true. |

Now criteria: need both A and B.

A: pleuritic chest pain – present (Acute pleuritic chest pain). B: need pleural rub OR pleural thickening OR imaging confirmed pleuritic effusion. The CT shows pleural effusion (imaging confirmed pleuritic effusion). So B satisfied. Also presence of pericardial rub but not needed.

Exclusions: infection – note says DDx: Viral pericarditis excluded (neg infectious workup). No mention of infection for pleurisy. Not uremia; labs not BUN. So no exclusion.

Thus criteria fulfilled.

Time: note dated 01/01/2022. The complaint is current, "Acute pleuritic chest pain" suggests present now. No timeframe given. By default assume within_10days. So time = within_10days.

Score: both criteria fulfilled and time within_10days => score 1.

Confidence: High confidence because explicit mention of pleuritic chest pain and imaging pleural effusion. So perhaps 9 or 10. Use 10.

Rationale: Provide short explanation.

Now create JSON with proper quoting (single quotes in example but JSON requires double quotes, but they used single quotes in sample. We'll follow same style, using single quotes). The fields:

descriptor: 'pleurisy'
relevant: true
criteria: 'fulfilled'
time: 'within_10days'
score: 1
confidence_score: 10
rationale: include quoted text.

Rationale sentence: "The note reports acute pleuritic chest pain and a CT showing bilateral pleural effusions, satisfying both A (pleuritic chest pain) and B (imaging-confirmed effusion) with no exclusions, and the findings are described as current, thus within 10 days."

Now produce answer with reasoning and answer tags.

| | | |
|---|---|---|
| | | **assistantfinal**<reasoning><br>The clinical note mentions “Acute pleuritic chest pain” (symptom keyword for criterion A) and a CT scan showing “bilateral pleural effusions” (imaging-confirmed effusion fulfilling criterion B). No exclusions (infection, uremia) are noted. The findings are presented as the current acute complaint, implying they are active at the time of documentation (01/01/2022), which fits the ‘within_10days’ time category. Therefore the descriptor meets the diagnostic criteria and timing requirements, giving a score of 1 with high confidence.<br></reasoning><br><answer><br>{<br>'descriptor': 'pleurisy',<br>'relevant': true,<br>'criteria': 'fulfilled',<br>'time': 'within_10days',<br>'score': 1,<br>'confidence_score': 10,<br>'rationale': "The note reports acute pleuritic chest pain and a CT showing bilateral pleural effusions, satisfying both A (pleuritic chest pain) and B (imaging-confirmed effusion) with no exclusions, and the findings are described as current, thus within 10 days."<br>}<br></answer> |
| pericarditis | Gilbert2021-F26-20211015.txt | **analysis**We need to process.<br><br>First, check relevance: Keywords pericarditis appear: "recurrent pericarditis", "pericardial inflammation", "pericardial pain"? There's "Worsening left-sided chest tightness x1 year, exacerbated by breathing, relieved at rest." That's chest tightness, not pericardial pain exactly. But "pericarditis" keyword present in PMH and impression: "SLE with recurrent pericarditis", "Active SLE flare with recurrent pericarditis". So relevant = true.<br><br>Now evaluate criteria: Need both A (pericardial pain) and B (pericardial rub OR ECG changes OR imaging pericardial effusion). Actually diagnostic criteria say both A and B need to be present. Let's see.<br><br>A: pericardial pain typical sharp, worse with inspiration, improved by leaning forward. The note says "Worsening left-sided chest tightness ×1 year, exacerbated by breathing, relieved at rest." That's chest tightness not mention of sharp, improved leaning forward. But it's chest tightness worsened by breathing; could be pericardial pain. However does it match the symptom keyword list? Symptoms/signs keywords: pericardial pain, pericardial chest pain, pericardial rub, cardiac tamponade. The note does not contain the exact phrase "pericardial pain" or "pericardial chest pain". It says "chest tightness" and "exacerbated by breathing". That might be considered a symptom but not explicit pericardial pain keyword. According to guidelines, we check for presence of specific symptoms/signs keywords. If not present, then not satisfied. However the diagnostic keyword list includes "pericarditis, hydropericardium" – present. That indicates descriptor maybe diagnosed. The guideline says: "Diagnostic keywords: Check for the presence of |

| | | |
|---|---|---|
| | | diagnostic keywords which are strong indicators confirming criteria fulfilment. (In such cases, it is not necessary to verify each point within the criteria)". So presence of diagnostic keyword "pericarditis" may be strong indicator, meaning criteria fulfilled without needing to verify A and B. Indeed they said if diagnostic keywords are present (e.g., "pericarditis") we can consider that as confirming criteria. So we can consider criteria fulfilled.<br><br>But need to consider exclusions: infection, uremia (BUN >40), Dressler's. No mention. So none.<br><br>Thus criteria = fulfilled.<br><br>Now time: The note date is 15/10/2021. Need to assess when the pericarditis active. The note says "Active SLE flare with recurrent pericarditis (cMRI-confirmed inflammation)". That suggests currently active. Also chest tightness is present now as CC. The symptom has been for 1 year but currently present. The time classification: active within 10 days? The note expresses current active flare. No explicit date. So default to within_10days as per guideline: if no date specified, default to within_10days unless indications suggest otherwise. So time = within_10days.<br><br>Score = 1 (criteria fulfilled and time within 10 days/11-30). So 1.<br><br>Confidence: fairly high because diagnostic keyword present plus imaging confirmation, active flare. I'd assign maybe 9.<br><br>Now produce JSON.<br><br>Check fields: descriptor 'pericarditis', relevant true, criteria 'fulfilled', time 'within_10days', score 1, confidence_score 9, rationale.<br><br>Rationale: mention presence of diagnostic keyword "pericarditis", cMRI confirmed pericardial inflammation, active flare noted, no exclusions, timeframe current.<br><br>Let's produce.**assistantfinal**<reasoning><br>The note contains the diagnostic keyword “pericarditis” (PMH: SLE with recurrent pericarditis; Impression: Active SLE flare with recurrent pericarditis) indicating the descriptor is relevant. According to the guideline, a diagnostic keyword is a strong indicator that fulfills the criteria without needing to verify each sub-criterion, so the criteria are considered **fulfilled**. No exclusion (infection, uremia, Dressler’s) is mentioned.<br><br>The impression states the pericarditis is “active” and the chief complaint of chest tightness exacerbated by breathing is ongoing, which means the manifestation is current. With no specific date given, the default is **within_10days**.<br><br>Since the criteria are fulfilled and the timing is within the allowed window, the descriptor receives a score of **1**. Confidence is high because the note explicitly confirms active pericarditis and provides imaging evidence, with no contradictory information. |

| | | |
|---|---|---|
| | | </reasoning><br><answer><br>{<br>"descriptor": "pericarditis",<br>"relevant": true,<br>"criteria": "fulfilled",<br>"time": "within_10days",<br>"score": 1,<br>"confidence_score": 9,<br>"rationale": "The note explicitly mentions \"recurrent pericarditis\" and an \"Active SLE flare with recurrent pericarditis (cMRI-confirmed inflammation)\", fulfilling diagnostic criteria; no exclusions are noted, and the condition is described as active, thus considered within the last 10 days."<br>}<br></answer> |
| low_complement | 026-F51-[masked].txt | **analysis**We need to parse the note. Descriptor low_complement. Look for keywords: C3, C4, complements, etc. In note: "C3/C4 63/15". So relevant true.<br><br>Now evaluate criteria: Need to determine if low complement is fulfilled. Values: C3 = 63, C4 = 15. Need reference thresholds: For date after 24/12/2018, thresholds: C3 <90 mg/dL is low; C4 <10 mg/dL is low. Note date: [masked], which is before 24/12/2018. So we use older thresholds: C3 <76, C4 <9. Values: C3 63 (<76) => low. C4 15 (>9) => not low. At least one low? The descriptor is "Decrease in CH50, C3, or C4". So if any of them low, criteria fulfilled? Probably yes: presence of low complement (any of them). So C3 low fulfills.<br><br>Check flags: no explicit flag "low" but numeric value is low according to threshold.<br><br>Thus criteria: fulfilled.<br><br>Now time: Note date [masked]. No explicit timing phrase for complement. The labs are presumably current to the note. No mention of when labs were drawn. Usually labs are recent as part of note. So we consider time as within_10days (default if no date specified). According to instruction: "if no date is specified => default to within_10days unless indications suggest otherwise". So time = within_10days.<br><br>Thus score = 1 (criteria fulfilled and time within_10days).<br><br>Confidence: fairly high. We have clear data: C3 value 63, threshold <76, value low. Note date earlier than threshold change date, we used correct threshold. So confidence maybe 9.<br><br>Rationale: mention that C3 63 is below reference (<76) indicating low complement; timing considered current; thus descriptor met. |

| | | |
|---|---|---|
| | | Now construct JSON.<br><br>Make sure correct field names etc. Use single quotes? The example uses single quotes but JSON requires double quotes, but spec shows single quotes. We'll follow spec example: use single quotes.<br><br>Return final.**assistantfinal**<reasoning><br>The note contains the keyword “C3/C4 63/15”, indicating complement levels are reported.<br>The note date ([masked]) is before 24/12/2018, so the applicable reference ranges are C3 < 76 mg/dL and C4 < 9 mg/dL.<br>C3 = 63 mg/dL is below the threshold, confirming low complement.<br>No timing qualifiers are provided; per guidelines the default is “within_10days”.<br>Thus the diagnostic criteria are **fulfilled**, the time classification is **within_10days**, leading to a score of 1.<br>Confidence is high because the numeric value directly meets the low-complement threshold and no contradictory information is present.<br></reasoning><br><answer><br>{<br>'descriptor': 'low_complement',<br>'relevant': true,<br>'criteria': 'fulfilled',<br>'time': 'within_10days',<br>'score': 1,<br>'confidence_score': 9,<br>'rationale': 'C3 is reported as 63, which is below the pre-2018 reference limit of <76 mg/dL, indicating low complement; the labs are presented in the current note with no time qualifiers, so they are treated as recent (within_10days).'<br>}<br></answer> |
| increased_DNA_binding | 045-F59-[masked].txt | **analysis**We need to parse the note.<br><br>We need to evaluate descriptor increased_DNA_binding.<br><br>First, check for relevant keywords: anti-ds DNA, anti DNA, dsDNA.<br><br>In note we have "Anti-dsDNA 63 → 72 → 49". So keywords present. Therefore relevant = true.<br><br>Now evaluate diagnostic criteria. |

| | | |
|---|---|---|
| | | We need to determine if increased DNA binding is present (i.e., elevated anti-dsDNA). The note lists three values: 63 → 72 → 49. Likely chronological values. Need the latest value: The most recent is 49 (assuming the order is older to newer? Typically arrow indicates progression over time, maybe 63 then 72 then 49. So the latest is 49. We need to see if that's above reference for ELISA. Method not mentioned, assume ELISA. Use reference for ELISA based on date. Note date is [masked], which is after 1/2/2021 (since [masked] > 2021), so reference is >10 IU/ml.<br><br>Values are likely IU/ml (units not given). 49 > 10, so it's elevated. However, need to check the rule: "Qualitative values (such as "high", "+", "rising", or similar terms) should be considered uncertain for ELISA". Our values are numeric, so they are quantitative.<br><br>Thus increased DNA binding is above reference.<br><br>But need to consider "twice above laboratory reference range". The descriptor definition: increased DNA binding above laboratory reference range, except ELISA: twice above laboratory reference range. Hmm note: "Increased DNA binding above laboratory reference range, except ELISA: twice above laboratory reference range." So for ELISA method, the increase must be at least twice the reference range? Let's parse: The descriptor condition says increased DNA binding above reference range, except for ELISA where it must be twice above reference range. So for ELISA, we need value >= 2 * reference.<br><br>Reference is >10 IU/ml (i.e., cutoff >10). Twice reference would be >20 IU/ml. 49 > 20, thus meets twice reference. So fulfills.<br><br>Now check if any exclusions. None.<br><br>Treatment response not needed because we have direct test.<br><br>Now time: When was the test performed? Not explicitly dated. We have three values: 63 → 72 → 49. No dates given. The note is dated [masked]. We assume the latest value is most recent, likely recent within the note. No explicit time given; default to within_10days unless indicated otherwise? The guidelines: "If no date is specified => default to within_10days unless indications suggest otherwise". So time = within_10days.<br><br>Thus criteria: fulfilled. time: within_10days. Score = 1.<br><br>Confidence: The note gives numeric values and reference cutoffs consistent; we are assuming ELISA method not specified but default. That's fairly confident. Maybe 8 or 9? There's a small uncertainty about method assumption and whether "twice above" required. But we have quantitative value well above. I'd give confidence 9.<br><br>Now produce JSON. |

| | | |
|---|---|---|
| | | **assistantfinal**<reasoning><br>The note contains the keyword “Anti-dsDNA”, indicating relevance. Three quantitative anti-dsDNA levels are listed (63 → 72 → 49). The most recent value is 49 IU/ml. No assay methodology is mentioned, so per instructions we assume ELISA. For ELISA the reference cutoff after 01/02/2021 is >10 IU/ml, and the descriptor requires at least twice that level (>20 IU/ml). The value 49 IU/ml exceeds both the basic cutoff and the “twice reference” requirement, satisfying the increased_DNA_binding criterion. No exclusions are noted. The test result is presented without a specific date; guidelines state that when no date is given we default to “within_10days”. Therefore the descriptor is fulfilled, the timing is within_10days, and the score should be 1. Confidence is high because the numeric result clearly meets the defined thresholds and there is no contradictory information, though the method assumption adds a slight uncertainty, so a confidence of 9/10 is appropriate.<br></reasoning><br><answer><br>{<br>"descriptor": "increased_DNA_binding",<br>"relevant": true,<br>"criteria": "fulfilled",<br>"time": "within_10days",<br>"score": 1,<br>"confidence_score": 9,<br>"rationale": "The note lists 'Anti-dsDNA 63 → 72 → 49' with the latest value 49 IU/ml. Assuming ELISA (default when method not given), the reference after 01/02/2021 is >10 IU/ml and the descriptor requires at least twice that (>20 IU/ml); 49 exceeds this, fulfilling the criterion. No exclusion is noted and no date is provided, so default timing is within_10days."<br>}<br></answer> |
| fever | Kato2005-M48-20041006.txt | **analysis**We need to assess fever descriptor. The note:<br><br>T 38.1°C, BP... c/o: 1-month history of fever, malaise... So fever present. Temperature recorded 38.1°C (>38). That meets diagnostic criterion (>38°C). Need to exclude infection. There is mention: "DDx: CNS infection ruled out ; metabolic/toxic causes excluded (labs nl)." Also labs show no infection signs? No WBC high, CRP not given. So infection appears excluded. No malignancy mentioned. So criteria fulfilled.<br><br>Time frame: note date 06/10/2004. Fever described as "1-month history of fever". So has been ongoing for a month. The symptom is still present (c/o fever). So active. Since it's within 1 month, it's more than 10 days ago but less than 30 days? Actually 1 month = approx 30 days. Need categories: within_10days or 11_to_30days. Since 1-month history suggests continuous up to now, likely within 30 days. Might be exactly 30 days, but still qualifies for 11_to_30days? The rule: active more than 10 days ago but less than 30 days ago => 11_to_30days. Since it's 1 month, could be 30 days, borderline. Could also treat as 11_to_30days. Probably time = 11_to_30days.<br><br>Thus criteria fulfilled, time within allowed, score =1. |

| | | |
|---|---|---|
| | | Confidence: high, as clear temperature >38, infection ruled out, time reasonable. Confidence maybe 9.<br><br>Now format JSON with fields.<br><br>Also need 'relevant': true (keywords present). criteria: 'fulfilled'. time: '11_to_30days'. Score 1. confidence_score 9. rationale: mention temperature 38.1°C, 1-month history, infection ruled out, etc.<br><br>Ok.**assistantfinal**<reasoning><br>The note contains a temperature reading of 38.1 °C and a statement of a “1-month history of fever,” fulfilling the SLE fever criterion of >38 °C. The differential diagnosis explicitly states “CNS infection ruled out” and no infectious or malignant causes are described, satisfying the exclusion requirement. Fever is therefore considered a lupus manifestation. The duration is described as a month, which places the active fever within the 11-to-30-day window (greater than 10 days but less than 30 days from the note date). Both criteria (fulfilled + appropriate time) are met, so the descriptor should receive a score of 1. Confidence is high because the evidence is direct and unambiguous.<br></reasoning><br><answer><br>{<br>"descriptor": "fever",<br>"relevant": true,<br>"criteria": "fulfilled",<br>"time": "11_to_30days",<br>"score": 1,<br>"confidence_score": 9,<br>"rationale": "The note records a temperature of 38.1 °C and a 1-month history of fever, satisfying the >38 °C criterion, and explicitly rules out CNS infection, meeting the exclusion rule; the fever is ongoing within the 11-to-30-day window, supporting a score of 1."<br>}<br></answer> |
| thrombocytopenia | Natarajan 2021-F23-20200930.txt | **analysis**We need to parse note for thrombocytopenia.<br><br>Keywords list: PLT, platelet. Note includes "Plt 70↓". That's a numeric value 70 (presumably 70,000/mm3) which is <100,000. So thrombocytopenia present. Need to consider exclusions: no mention of drug causing it, other causes? They have pancytopenia from SLE likely. So criteria: fulfilled if platelet count <100k, and no other known cause. No exclusion noted. So criteria = fulfilled.<br><br>Now time: note dated 30/09/2020. The labs are presumably recent, no date mentioned but part of current assessment. The symptom is present now. So time classification: within_10days (active). Could also be 11_to_30days but default is within_10days if not specified, assume current labs. So time = |

| | | |
|---|---|---|
| | | within_10days.<br><br>Thus score = 1 (criteria fulfilled and time within_10days).<br><br>Confidence: high, because explicit numeric value, clear criteria, no exclusions. Confidence maybe 9 or 10. Since note includes numeric platelet value 70 (presumably 70k), and recent labs; I think 10.<br><br>Now generate JSON.<br><br>Also need fields: relevant true because keyword "Plt" present.<br><br>criteria = 'fulfilled'.<br><br>time = 'within_10days'.<br><br>score 1.<br><br>confidence_score 10.<br><br>rationale: "Plt 70↓" indicates platelet count 70,000 (<100,000) confirming thrombocytopenia; labs are current in note dated 30/09/2020, no exclusion noted, so criteria fulfilled and active now."<br><br>Return format.**assistantfinal**<reasoning><br>The note contains the keyword **Plt** with a numerical value: "Plt 70↓". This indicates a platelet count of $70 \times 10^3$/µL, which is below the threshold of $100 \times 10^3$/µL required for thrombocytopenia. No alternative cause (e.g., drug effect, portal hypertension, TTP) is mentioned, so the SLE-related criterion is satisfied.<br><br>The laboratory values are presented as part of the current work-up on the note dated **30/09/2020**, implying they are recent. No resolution is stated, therefore the finding is considered active now, i.e., **within_10days**.<br><br>Thus the descriptor is relevant, the diagnostic criteria are **fulfilled**, the timing is appropriate, and the score should be **1**. The evidence is explicit and unambiguous, giving a high confidence.<br></reasoning><br><answer><br>{ |

| | | |
|---|---|---|
| | | "descriptor": "thrombocytopenia",<br>"relevant": true,<br>"criteria": "fulfilled",<br>"time": "within_10days",<br>"score": 1,<br>"confidence_score": 10,<br>"rationale": "The note reports 'Plt 70↓', a platelet count of 70,000/mm³ (<100,000), meeting the thrombocytopenia threshold. No alternative cause is documented, and the labs are presented as current (note dated 30/09/2020), indicating the finding is active now."<br>}<br></answer> |
| leukopenia | Nishimura2003-F55-20020122.txt | **analysis**We need to evaluate leukopenia descriptor.<br><br>Keywords: WCC, WBC. In note: "Labs (Jan 18): WBC 2.5↓". So relevant = true.<br><br>Now determine criteria. Diagnostic threshold: leukocyte count <3,000/mm3 or <3 x10^9/L. The note says WBC 2.5 ↓. Units default 1000/mm3 or 10^9/L. So 2.5 indicates 2.5 x10^9/L = 2500/mm3, which is below 3. So meets threshold. It's a numeric value; counts as fulfillment.<br><br>Check for exclusions: Felty's, drug causes, portal hypertension. No mention. Could be steroids causing leukopenia? But steroids usually cause neutrophilia, not leukopenia. Drug causes: maybe cyclophosphamide? But that's later. The leukopenia is noted Jan 18 before cyclophosphamide started Jan 29. So no drug cause noted. So criteria fulfilled.<br><br>Now time: The note date is 22/01/2002. Labs date Jan 18. That's 4 days before note. So within_10days. Also still present as part of impression (Leukopenia). So time = within_10days.<br><br>Thus criteria fulfilled, time within_10days => score 1.<br><br>Confidence: We have clear numeric value; clear timeframe; no exclusions. So high confidence, maybe 10.<br><br>Return JSON.**assistantfinal**<reasoning><br>The clinical note contains the keyword **WBC** with a specific numeric value: “WBC 2.5↓” on 18 Jan. This value (2.5 × $10^9$/L) is below the leukopenia threshold of <3 × $10^9$/L, fulfilling the diagnostic criterion. No exclusions (Felty’s, drug-induced, portal hypertension) are mentioned. The laboratory result is dated 18 Jan, which is 4 days before the note date (22 Jan/2002), placing the finding in the **within_10days** time window. Therefore the descriptor is relevant, criteria are fulfilled, time is recent, and the score should be 1 with high confidence.<br></reasoning><br><answer> |

| | | |
|---|---|---|
| | | {<br>"descriptor": "leukopenia",<br>"relevant": true,<br>"criteria": "fulfilled",<br>"time": "within_10days",<br>"score": 1,<br>"confidence_score": 10,<br>"rationale": "Lab result shows WBC 2.5 (↓), which is $<3 \times 10^9$/L, meeting the leukopenia threshold; no exclusions are noted; the lab was performed 4 days before the note date, so the finding is within 10 days."<br>}<br></answer> |

(a) (b)

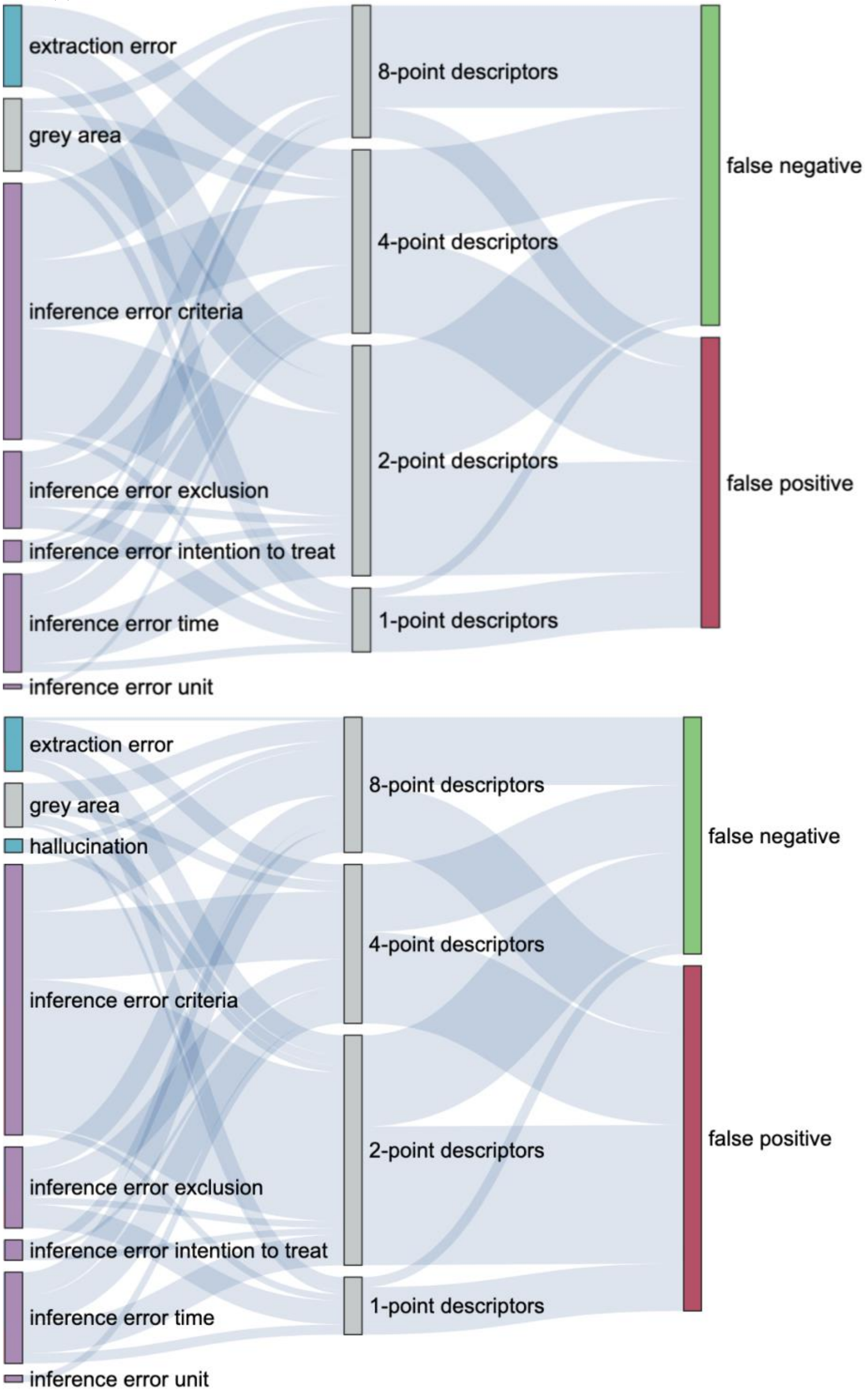

**Figure S11. Error analysis of all error records at the descriptor level (714 cases pooled together).** (a) A Sankey plot showing error records generated by first-layer GPT-OSS-120B. (b) A Sankey plot showing error records generated by first-layer Qwen3-32B.

**Table S7. Examples of error types.** Dates are masked to protect privacy.

| filename | data | descriptor | misclassification | LLM answer (majority) | vote agreement | LLM output rationale | correct answer | error type | Comments from doctors |
|---|---|---|---|---|---|---|---|---|---|
| 054-F47-[masked].txt | training | arthritis | false positive | 1 | 1 | The note reports 'multi-joint pain' with 'swelling +/-', indicating ≥2 inflamed joints (criterion A). No exclusionary causes are mentioned. The episode started ~3 weeks ago and resolved, fitting the 11-to-30-day window, supporting a score of 1. | 0 | Extraction error | LLM overlooked "self increase dose of prednisolone to 10mg daily for recent 3/53 no more pain after 3-4 days then currently nil compliaint" , the pain was resolved by self-treatment, currently nil pain, no active arthritis |
| 032-F51-[masked].txt | testing | hematuria | false positive | 1 | 0.8 | The note states 'Urine RBC- P++', which meets the ≥2+ threshold for hematuria; no exclusions are listed and the finding is presented as current, so it is considered active within the past 10 days. | 0 | Extraction error | the value for urine RBC here is "-" |
| 053-F69-[masked].txt | training | increased_DNA_binding | false negative | 0 | 1 | The note reports anti-dsDNA 21 with a reference range of 21-25 IU/ml, which is within normal limits for ELISA (>10 IU/ml positive); thus increased DNA binding is not demonstrated. The test is recent, but the diagnostic threshold is not met. | 1 | Extraction error | "21 <---21 <---25" was indicating the dynamic changes of dsDNA, but not the reference |
| Prasad 2022-F36-20220502.txt | testing | psychosis | false positive | 1 | 0.6 | The note records persecutory delusions, auditory hallucinations and incoherent speech ('persecutory delusions, auditory hallucinations' and 'incoherent speech') onset 3 days ago, meeting the psychosis criteria. No exclusion (e.g., uremia, drug-induced) is noted. The treatment plan escalates lupus therapy with high-dose dexamethasone and cyclophosphamide, indicating treat_escalated intent. | 0 | Inference error criteria | The LLM failed to differentiate between psychosis and delirium. diagnosis clearly stated "Delirium". Key features for this case are "acute onset“ and "fluctuating orientation". |
| pseudo 068-F52- | training | arthritis | false negative | 0 | 0.6 | The note lists 'arthritis (MCPs, PIPs)' but provides no mention of tenderness, swelling, effusion, or prolonged morning stiffness required for SLE arthritis criteria; thus the criteria are not met despite the current timing. | 1 | Inference error criteria | diagnostic keyword "arthritis", no need to meet all criteria |

| | | | | | | | | | |
|---|---|---|---|---|---|---|---|---|---|
| [masked].txt | | | | | | | | | |
| Nayak2012-F17-20110715.txt | training | arthritis | false negative | 0 | 0.6 | Physical exam notes 'both hand PIPs tenderness' (two tender joints) but lacks swelling, effusion, synovitis, or documented ≥30 min morning stiffness required for arthritis criteria; thus criteria are not met despite current (within 10 days) presence. | 1 | Inference error criteria | "both hand PIPs tenderness" with active lupus psychosis (the patient had a altered mental status, so cannot report pain properly) |
| Kutky2018-M74-20180101.txt | training | proteinuria | false positive | 1 | 1 | The note records a UACR of 305 mg/mmol, well above the >0.5 threshold, and the impression cites active lupus nephritis, confirming SLE-related proteinuria; the finding is current, thus within 10 days. | 0 | Inference error criteria | UACR does not count |
| Narang2022-M20-20220101.txt | testing | pericarditis | false negative | 0 | 1 | Physical exam notes a pericardial rub, satisfying criterion B, but no pericardial pain (criterion A) is documented. Without both criteria the descriptor is unfulfilled; the rub is present now, so time is within 10 days, leading to a score of 0. | 1 | Inference error criteria | diagnostic keyword "Serositis", no need to meet all criteria |
| 091-F54-[masked].txt | testing | increased_DNA_binding | false positive | 1 | 0.6 | The note states 'serology improving dsDNA 85'; assuming ELISA, the reference for [masked] is >67.5 IU/ml, so 85 is elevated, meeting increased DNA binding. The result is presented in the current note, implying recent testing (within 10 days). No exclusions are noted. | 0 | Inference error criteria | The LLM didn't double the ELISA upper limit of the reference |
| Saini2013-F0-20131015.txt | training | seizure | false negative | 0 | 0.8 | The note describes acute left focal seizures ('acute left focal seizures + lethargy ×14d', 'left clonic activity'), satisfying the seizure description criterion, but a documented calcium deficiency ($Ca^{2+}$ 6.6↓) is an explicit exclusion for seizures, making the diagnostic criteria unfulfilled. The seizures began ~14 days ago, placing the timing in the 11-to-30-day window, and the management plan does not involve escalation of lupus-specific therapy. | 1 | Inference error exclusion | diagnosis specified "Neonatal Lupus with CNS Vasculitis", despite low calcium may also contribute to seizure, the cause of lupus is certain |

| | | | | | | | | | |
|---|---|---|---|---|---|---|---|---|---|
| 053-F68-[masked].txt | training | arthritis | false negative | 0 | 0.8 | The note describes joint pain in the hands, right hip, and knee but attributes it to right AVN and provides no signs of inflammation (swelling, tenderness, effusion) or prolonged morning stiffness; AVN is an explicit exclusion for lupus arthritis. | 1 | Inference error exclusion | R hip / knee was due to AVN, but there was also a "joint flare" where "Increasing small hand joint pains with EMS" |
| 058-F49-[masked].txt | testing | pyuria | false positive | 1 | 1 | The note reports 'Urine showed WBC 10-50' ( >5 WBC/HPF) and 'UP+' (positive urine protein dipstick). No significant infection is noted (insignificant growth, AFB smear negative), satisfying pyuria criteria. Findings are presented as current lab values, so the time is considered within 10 days. | 0 | Inference error exclusion | "insignificant growth AFB smear -ve await culture ", infection cannot be fully excluded |
| 025-F62-[masked].txt | training | fever | false positive | 1 | 0.6 | Temperatures recorded as 38.1 °C today ("Temp 38.1 on measurement") meet the >38 °C requirement; no explicit infection exclusion is documented, and the symptom is noted as occurring today, placing it within the 10-day window. | 0 | Inference error exclusion | fever with cough, treatment with Levofloxacin, likely due to URI, which is an exclusion |
| 010-F46-[masked].txt | training | fever | false positive | 1 | 0.8 | The note states 'recently admitted x PUO', indicating fever ≥38 °C; infection work-up is negative, satisfying exclusion criteria. The term 'recently' places the fever within the last 10 days. | 0 | Inference error exclusion | The treatment intention was empirical anti-TB Rx, doctors not consider fever to be lupus related |
| 026-F55-[masked].txt | testing | arthritis | false negative | 0 | 1 | The note reports increased morning stiffness and polyarthralgia (multiple joint pain) but does not specify ≥2 tender joints with ≥30 min stiffness or objective synovitis, so the arthritis criteria are not fulfilled despite the symptom being active today. | 1 | Inference error intention to treat | increase prednisolone 7.5mg qd for joint, indicating doctor's impression is lupus flare |
| 037-F39-[masked].txt | training | rash | false negative | 0 | 1 | The clinical note mentions a deteriorated skin condition but does not use the keyword 'rash' or any other diagnostic terms for lupus rash; thus the descriptor is not relevant. | 1 | Inference error intention to treat | "recently deteriorated in skin condition given plaguenil ( hydroxycholoquine 600 mg qd by American doctor )" should be score according to treatment intention |
| 099-F40-[masked].txt | testing | rash | false negative | 0 | 1 | The note reports a 'slight increased in facial rash' but does not label it as a lupus-specific rash (e.g., malar, discoid, SCLE) nor use diagnostic keywords, so the lupus rash criterion is not met; the rash is described as currently present, thus time = within_10days. | 1 | Inference error intention to treat | "Slight increased in facial rash, advised on avoidance of sunlight, patient not keen for increased HCQ", so treatment intention is HCQ, indicating a lupus rash |

| | | | | | | | | | |
|---|---|---|---|---|---|---|---|---|---|
| Kobayashi1992-F32-19890930.txt | training | cranial_nerve_disorder | false negative | 0 | 0.8 | The note describes bilateral sensorineural hearing loss (vestibulo-cochlear nerve involvement) but specifies a gradual decline since 1985, indicating a chronic rather than new-onset disorder; thus the diagnostic criterion is not met, leading to a score of 0 despite treatment escalation. | 1 | Temporal reasoning | The patient had a refractory bilateral sensorineural hearing loss, current still exist, therefore time_within10days |
| 027-F53-[masked].txt | testing | proteinuria | false positive | 1 | 1 | The note states "Latest 24 hour UP 2.4g/day", which exceeds the >0.50 g/24 h threshold for proteinuria and is recent (latest result), so the criteria are fulfilled and the symptom is active within 10 days. | 0 | Temporal reasoning | latest should be considered to be time_uncertain |
| 110-M61-[masked].txt | training | increased_DNA_binding | false positive | 1 | 0.8 | The note lists 'Anti-dsDNA 83.4 ← 117.9 ← 155.6' with the most recent value 83.4 IU/ml, which exceeds the ELISA reference (>25 IU/ml) and is more than twice that threshold. The test is from 'Bloods from last visit,' indicating a recent result, thus within_10days. | 0 | Temporal reasoning | last time is time uncertain |

**Table S8. Baseline Characteristics of Patients included in the clinical/research implementation study.**

| | Overall | LLM-determined SSLDA* | LLM-determined Non-SSLDA* | p | Missing (%) |
|---|---|---|---|---|---|
| N (%) | 108 | 45 (41.7) | 63 (58.3) | | |
| Gender (%) | | | | | |
| Female | 94 (87.0) | 41 ( 91.1) | 53 ( 84.1) | 0.438 | 0 |
| Male | 14 (13.0) | 4 ( 8.9) | 10 ( 15.9) | | |
| Age, year (median [IQR]) | 45.00 [39.00, 52.00] | 43.00 [38.00, 50.25] | 47.00 [39.00, 52.50] | 0.251 | 0 |
| ISN/RPS Classification of Lupus Nephritis (%) | | | | | |
| II | 6 ( 5.6) | 2 ( 4.4) | 4 ( 6.5) | 0.425 | 0.9 |
| III | 9 ( 8.4) | 4 ( 8.9) | 5 ( 8.1) | | |
| III+V | 4 ( 3.7) | 2 ( 4.4) | 2 ( 3.2) | | |
| IV | 29 (27.1) | 10 ( 22.2) | 19 ( 30.6) | | |
| IV+V | 5 ( 4.7) | 1 ( 2.2) | 4 ( 6.5) | | |
| V | 23 (21.5) | 9 ( 20.0) | 14 ( 22.6) | | |
| unknown biopsy | 17 (15.9) | 7 ( 15.6) | 10 ( 16.1) | | |
| non-LN | 14 (13.1) | 10 ( 22.2) | 4 ( 6.5) | | |
| LN duration, year (median [IQR]) | 12.08 [6.45, 17.90] | 13.01 [8.41, 15.53] | 9.85 [6.45, 18.07] | 0.451 | 29.6 |
| SLE duration, year (median [IQR]) | 14.55 [9.44, 21.07] | 14.76 [10.91, 21.48] | 13.61 [8.63, 20.71] | 0.579 | 8.3 |
| Time from first visit in QMH to baseline, year (median [IQR]) | 11.57 [6.56, 18.13] | 12.52 [6.72, 18.06] | 11.31 [6.56, 17.86] | 0.751 | 5.6 |
| Developed to stage 3 CKD within 5 years from baseline | 29 (26.9) | 5 ( 11.1) | 24 ( 38.1) | 0.004 | 0 |

*Sustained SLEDAI-2K low disease activity (SSLDA): defined as a SLEDAI-2K ≤4 for five years without major organ involvement (renal, central nervous system, cardiopulmonary, or vasculitis). The LLM used here is the best-performing two-layered GPT-OSS + verifier.

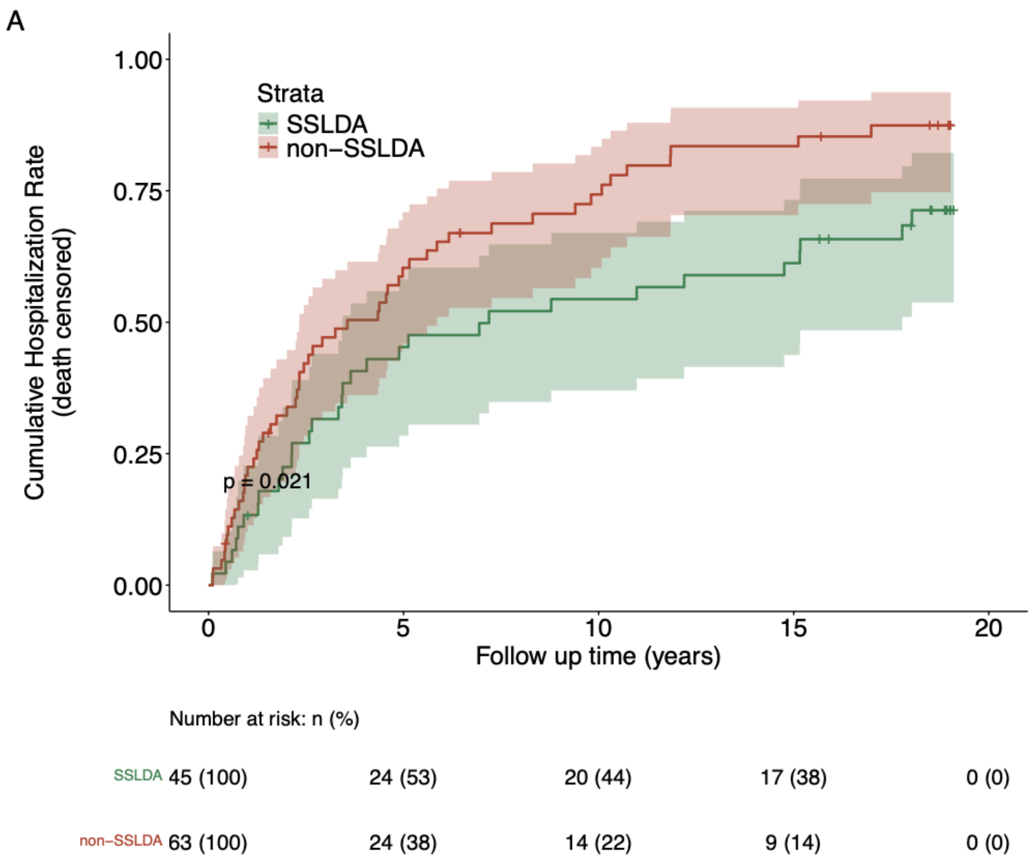

**Figure S12. Kaplan-Meier curve for time to hospitalization.** The clinical/research implementation study using large language models determined sustained SLEDAI-2K low disease activity (SSLDA) as a risk stratification tool for long-term outcomes in an LN-dominant SLE cohort.

**Table S9. Cox regression results for LLM-determined non-SSLDA in predicting long-term clinical outcomes.**

| Outcome | Predictors* | Models for adjustment# | Adjusted hazard ratio (aHR) | p value | Number of event/Number of patients |
|---|---|---|---|---|---|
| All-cause mortality | LLM-determined non-SSLDA | Model 1 | 1.18 (0.55-2.52) | 0.673 | 29/108 |
| | LLM-determined non-SSLDA | Model 2 | 1.62 (0.62-4.23) | 0.32 | 21/76 |
| | LLM-determined non-SSLDA | Model 3 | 1.33 (0.62-2.84) | 0.458 | 29/102 |
| Kidney replacement therapy | LLM-determined non-SSLDA | Model 1 | 6.15 (0.77-49.44) | 0.09 | 9/107 |
| | LLM-determined non-SSLDA | Model 2 | 5.74 (0.69-47.51) | 0.10 | 8/75 |
| | LLM-determined non-SSLDA | Model 3 | 5.98 (0.72-49.48) | 0.10 | 8/101 |
| Stage 3 chronic kidney disease | LLM-determined non-SSLDA | Model 1 | 3.11 (1.36-7.13) | **0.007** | 9/107 |
| | LLM-determined non-SSLDA | Model 2 | 5.66 (1.72-18.6) | **0.004** | 19/52 |

| | | | | | |
|---|---|---|---|---|---|
| | LLM-determined non-SSLDA | Model 3 | 3.18 (1.37-7.39) | **0.007** | 26/74 |
| Hospitalizatio n | LLM-determined non-SSLDA | Model 1 | 1.65 (1.05-2.6) | **0.03** | 88/108 |
| | LLM-determined non-SSLDA | Model 2 | 1.88 (1.06-3.33) | **0.03** | 58/76 |
| | LLM-determined non-SSLDA | Model 3 | 1.84 (1.16-2.9) | **0.009** | 82/102 |

*reference group is LLM-determined SSLDA. Sustained SLEDAI-2K low disease activity (SSLDA): defined as a SLEDAI-2K ≤4 for five years without major organ involvement (renal, central nervous system, cardiopulmonary, or vasculitis). The LLM used here is the best-performing two-layered GPT-OSS + verifier.

#Model 1: adjusted for baseline age and sex. Model 2: adjusted for age, sex, and LN duration. Model 3: adjusted for age, sex, and SLE duration.

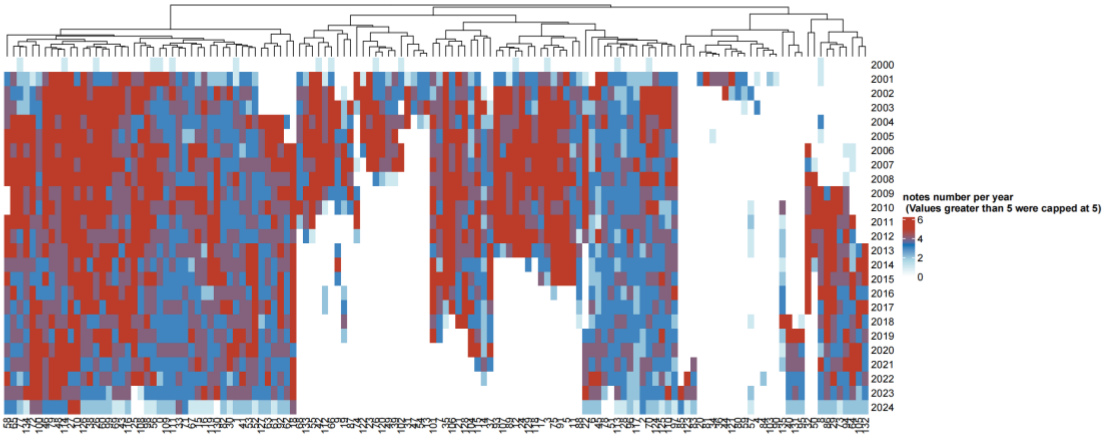

**Figure S13. Heatmap showing the number of notes for each SLE patient per year.** Each column indicates a patient (patient ID shown at the bottom), and each row indicates the year. The complete linkage method was used to perform hierarchical clustering based on Euclidean distance to better visualize the distribution of the notes. This study utilized electronic health records (EHRs) from the Hong Kong Hospital Authority's (HA) Clinical Management System (CMS). The HA launched the CMS II system, which introduced the foundational Electronic Patient Record (ePR) from 2000. This marked the beginning of a centralized, computerized system for patient data, replacing paper-based records in pilot clusters.

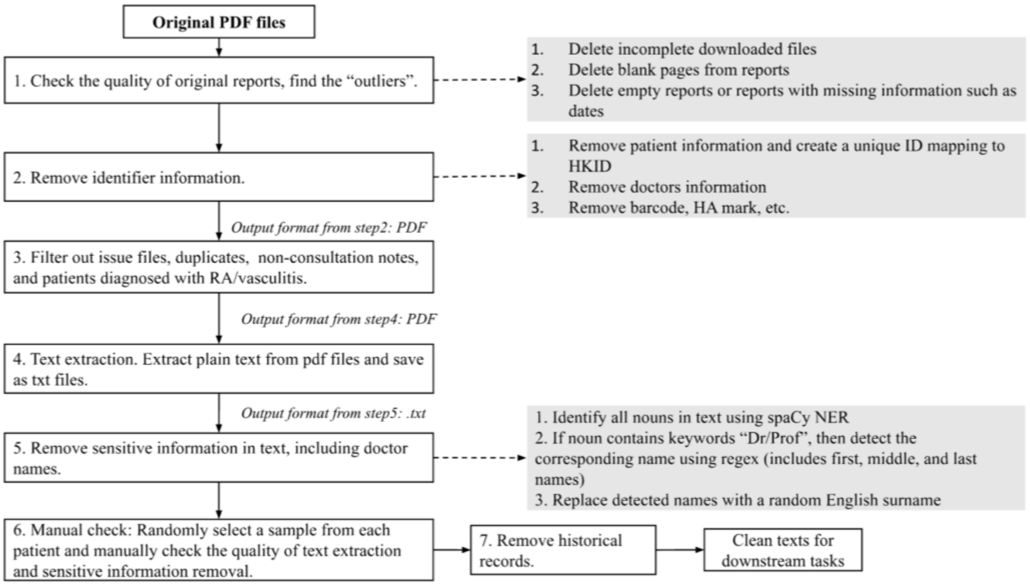

**Figure S14. Flowchart for data cleaning and de-identification.** A schematic diagram illustrating the step-by-step process for preparing the raw clinical data. This includes procedures for removing protected health information (PHI) and standardizing formats to generate a de-identified, analysis-ready dataset. Clinical notes were downloaded from the Hong Kong Hospital Authority's (HA) Clinical Management System (CMS) in PDF format. As the primary provider of public healthcare in Hong Kong, the HA delivers over 90% of inpatient care and the vast majority of specialist outpatient services.[1] Incomplete downloads and notes missing essential metadata (e.g., dates) were removed. All patient and clinician identifiers, including names, HKIDs, barcodes, and institutional marks, were masked, and a unique ID was assigned to each patient. We excluded issue files, duplicates, and non-consultation notes. The remaining documents were converted to plain text, and additional sensitive information, such as clinician names, was removed using a combination of named entity recognition and regex matching. The processed notes were saved as text files, with each filename encoding the patient's sex, visit age, and visit date.

1 https://www.ha.org.hk/visitor/ha_visitor_text_index.asp?Content_ID=10006&Lang=ENG&Dimension=1#:~:text=Welcome%20Messages,determined%20to%20scale%20new%20heights.

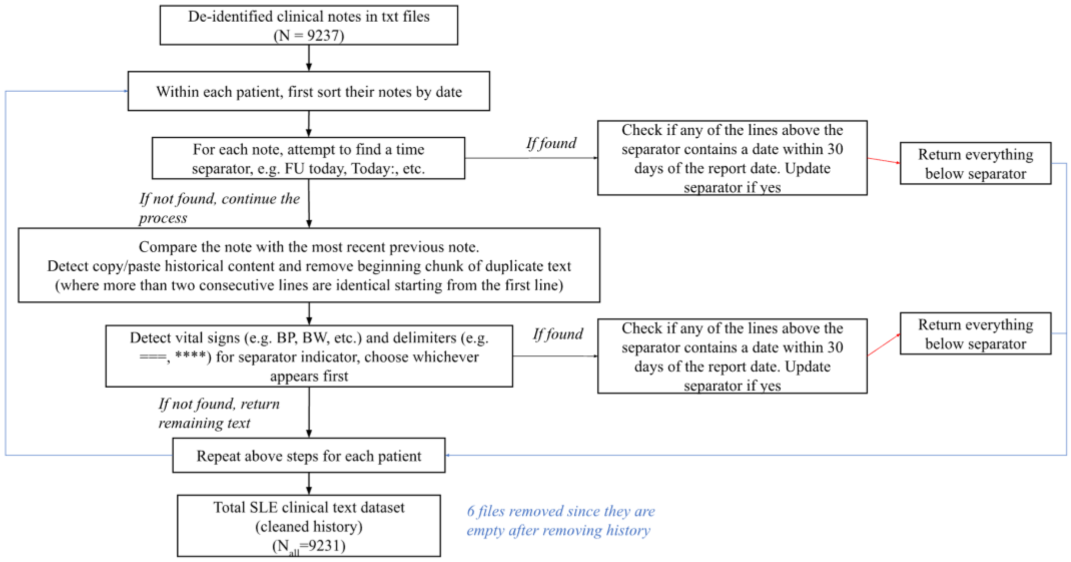

**Figure S15. Flowchart for historical content removal.** A flowchart detailing the algorithm and criteria used to identify and filter out historical information from clinical reports, ensuring that the analysis focuses solely on the current 30-day clinical episode. Clinical notes for each patient were first sorted chronologically. Each note was then processed in ascending order to identify a temporal delimiter (e.g., “FU today”, “today”) which typically marks the beginning of the current visit. If found, we examined the preceding lines for any date references within 30 days of the note’s documented date; if present, the delimiter was updated accordingly, and only the content below it was retained. If no delimiter was found, we compared the note to the most recent prior note to detect and remove copy-pasted historical content, defined as blocks of two or more consecutive identical lines from the beginning of the note. We also searched for indicators such as vital signs (e.g., blood pressure, body weight) and formatting delimiters (e.g., “====”, “****”) to infer the start of the current visit. If a suitable separator was identified, we again checked for recent date references above it and retained only the relevant portion. If no separator could be determined, the entire remaining note was preserved. This procedure was applied across all patients, resulting in a total of 9,231 cleaned clinical notes. Six notes were excluded due to being empty after history removal.

#### Supplementary Note 1. Step-by-step process of oversampling rare descriptors to get target notes

An initial seed set of 34 samples was selected from the population of 9,231 clinical notes for initial development and analysis. These were selected based on balancing the distribution of keyword presence, including samples in which relevant keywords were mentioned versus not mentioned, as well as those where keywords were mentioned in a negated versus non-negated context. To oversample rare descriptors, an additional 455 samples were selected according to the workflow shown in Figure S16 below.

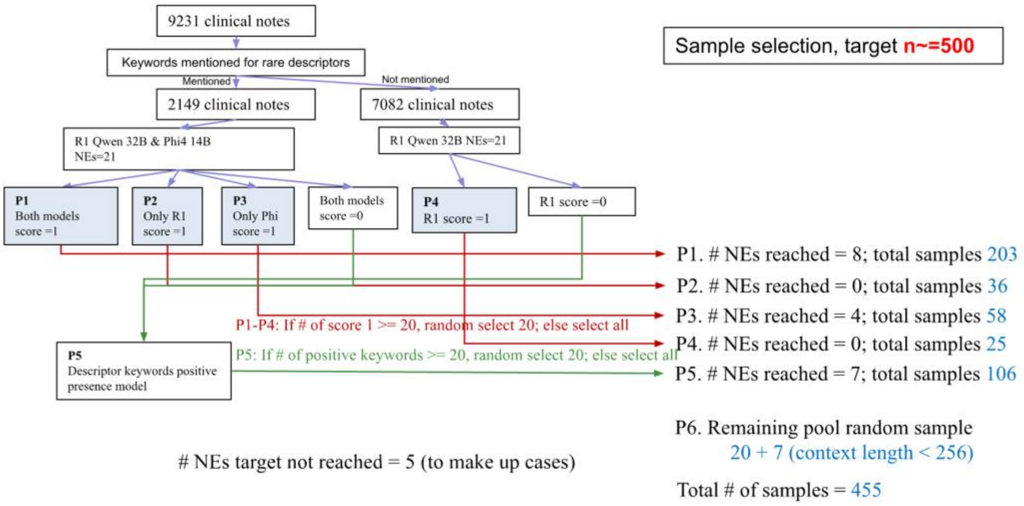

**Figure S16. Flowchart for selecting cases for annotation.** A diagram outlining the multi-step process for selecting 455 cases after an initial seed set of 34 (with a final cohort of 489) for manual annotation by clinical experts, including inclusion and exclusion criteria.

The sampling process began with an initial keyword-based filtering step using rare NE-related keywords indicative of potential relevance to the target descriptors (seen in annotation guidelines Tables 5 and 6). This filtering divided the notes into two pools: Pool A, consisting of 2,149 notes with at least one relevant keyword, and Pool B, comprising 7,082 notes without any keyword matches. Subsequently, each note was evaluated by large language models: Pool A, being smaller, was assessed using both DeepSeek-R1-Distill-Qwen-32B and Phi-4-14B; Pool B, due to its larger size, was evaluated solely with DeepSeek-R1-Distill-Qwen-32B to reduce computational cost. These models were selected based on superior performance in preliminary experiments, particularly in recall and computational efficiency, supporting the identification of potential positive samples at scale (see preliminary performance in Table S10 below).

**Table S10. NER performance of various models in a preliminary experiment.** A table showing the performance of various models evaluated on 50 cases in a preliminary experiment.

| Model | Precision | Recall | F1 | Runtime (sec) | # GPUs (RTX 4090) |
|---|---|---|---|---|---|
| DeepSeek-R1-Distill-Qwen-32B | 0.75 | **1.00** | 0.77 | 2849 | 4 |
| QwQ-32B | 0.58 | 0.75 | 0.79 | 5005 | 4 |
| Phi-4-14B | 0.30 | **0.94** | 0.36 | **1287** | **2** |
| Gemma-3-27B | 0.46 | **0.94** | 0.51 | 1460 | 4 |

Based on model outputs, notes were categorized into five subsets per descriptor entity: P1 (Pool A notes identified as positive by both models), P2/P3 (Pool A notes identified as positive by only one model), P4 (Pool B notes identified as positive by DeepSeek-R1-Distill-Qwen-32B), and P5 (notes from either pool classified as negative by all models, despite containing non-negated keywords (e.g. "oral ulcers+" vs negated form "no oral ulcers")). A stratified selection was then performed to prioritize subsets enriched with NEs. For P1 to P5, if an entity had 20 or more associated notes, 20 were randomly selected; otherwise, all notes for that entity were included. Since the target number of five named entities remained short after this selection, a final random top-up of 27 notes was drawn from the remaining notes. Together with the initial seed set, a total of 489 cases were selected from the collection of real clinical notes.

#### Supplementary Note 2. Pseudo-case creation and sources

To supplement rare descriptor cases that are either absent or insufficiently captured in the original data, we curated hypothetical cases into the dataset. Specifically, we required each descriptor to have at least 10 positive and 10 relevant but negative cases. This was done using two approaches: (1) augmenting real clinical notes by inserting additional text, such as treatments, symptoms, or details that fulfill criteria for underrepresented descriptors, and (2) collecting published case reports that describe rare descriptors meeting SLE diagnostic criteria and converting them into clinical note format using LLMs. We utilized an LLM prompt to help extract and structure clinical information into semi-structured clinical notes, covering age, sex, visit date, vital signs, chief complaint, history, physical examination findings, paraclinical test results, impression, and management plan. Ultimately, we generated 59 pseudo-notes derived from QMH cases and 110 pseudo-notes from published case reports (Table S11 below). All pseudo cases were reviewed, modified, and confirmed by three clinical experts (E.W., S.C., and D.Y.) to ensure diversity, uncertainty, representativeness, and consistency with the standards of a well-curated clinical note suitable for LLM analysis.

The following prompt was used in conjunction with DeepSeek-V3.1:

```
Transform the provided Systemic Lupus Erythematosus (SLE) case report
{{ case_report }}
into an outpatient clinical note documenting disease flare. Using this structure, remove the subtitle/subtitle number/bullet point and try to make it look like it was written by a human (with some abbreviations and personal writing style is okay):

Title: `[filename]-[Sex][age]-[Visit Date in YYYYMMDD format]`
*(Example: Appenzeller2009-F35-20230915)*
*Use the date corresponding to the most severe disease activity*

Note Body:
1.Vitals: [BP, HR, Temp, RR, O2 sat - state "-" if absent]

2. Complaint & History of present illness & past medical history
Patient Complaint: Chief symptom(s) at peak activity
Prior Tx Response: Key treatments tried & outcomes from other institutions & symptom progression/changes

3.PE
Focus on active SLE findings (e.g., "Malar rash", "Synovitis in MCPs 2-3", "Pericardial rub")

4.Paraclinical tests including laboratory testa, imaging, Electrophysiological:
(Examples: dsDNA 312 IU/mL↑(30/09/23), C3 0.48 g/L↓; UP: 2+ )
(Examples: CXR on 23/12/10: no lung lesion, just pleural effusion)

4. impression
  • impression: Confirm `Active SLE` with key features (cite organ involvement)
  • DDx: [Relevant differentials w/ruling rationale]
   *(Example: "RA excluded by negative CCP/absence of erosions; TTP ruled out by normal ADAMTS13")*
5. Management Plan:
Specific drug/dose changes (e.g., "MMF increase to 1.5g BID")
Prophylaxis/Vax if applicable
Critical labs/imaging/follow-up timing

Extraction Rules:
• Prioritize findings demonstrating active SLE (e.g., lupus nephritis, serositis...)
• Convert narrative lab data to abbreviated clinical format (e.g., "WBC 3.2↓" instead of "leukopenia")
• If case report lacks dates, consider the publication date and estimate timing relative to symptom flare.
```

**Table S11: DOIs of the 110 pseudo-cases generated from published case reports.**

| no | filename | DOI | SLEDAI-2K Descriptor scoring |
|---|---|---|---|
| 1 | Abuhammad2024-F20-20240131.txt | 10.30574/wjarr.2024.21.2.0385 | seizure, rash, pericarditis, increased_DNA_binding |
| 2 | Abuhammad2024-F38-20231215.txt | 10.30574/wjarr.2024.21.2.0385 | seizure, rash, alopecia, low_complement |
| 3 | Adelowo2012-F27-20120401.txt | 10.1136/bcr-2012-006259 | vasculitis, arthritis |
| 4 | Aldhefeery2023-M34-20230101.txt | 10.12659/AJCR.942085 | visual_disturbance, arthritis, low_complement, increased_DNA_binding |
| 5 | Alharmoodi2025-F60-20250215.txt | 10.7759/cureus.87266 | negative control |
| 6 | Amalia2023-F27-20221215.txt | 10.2147/JBM.S387075 | CVA, low_complement |
| 7 | Appenzeller2009-F69-20081215.txt | 10.1177/0961203308101546 | negative control |
| 8 | Aryal2023-F70-20221223.txt | 10.1002/ccr3.8192 | CVA |
| 9 | Atawneh2025-F22-20250222.txt | 10.52338/jocc.2025.4536 | seizure, arthritis, hematuria, pyuria, rash, mucosal_ulcers, low_complement |
| 10 | Bogaczewicz2013-M31-20130915.txt | 10.1177/0961203313513818 | arthritis |
| 11 | Calabuig2025-F52-20240415.txt | 10.3899/jrheum.2025-0390.PV305 | lupus_headache, arthritis, rash |
| 12 | Castillo2014-F55-20130615.txt | 10.1177/0961203314534303 | negative control |
| 13 | Chen2025-F17-20230315.txt | 10.1097/MD.0000000000043428 | seizure, proteinuria, low_complement, thrombocytopenia |
| 14 | Cordero2021-M30-20210410.txt | 10.38207/jmcrcs2021/0212110 | psychosis |
| 15 | Cravero2021-F17-20210715.txt | 10.26502/acmcr.96550431 | negative control |
| 16 | Cure2023-F35-20221215.txt | 10.1111/1756-185X.14652 | vasculitis, low_complement, increased_DNA_binding |
| 17 | Dey2020-F28-20200130.txt | 10.4103/jfmpc.jfmpc_186_20 | arthritis, myositis, proteinuria, low_complement, leukopenia |
| 18 | Dharamdasani2021-F24-20130902.txt | 10.14260/jemds/2021/382 | visual_disturbance |
| 19 | Ebrahimi2024-F53-20241020.txt | 10.34172/mj.025.33626. Persian | organic_brain_syndrome, hematuria, proteinuria, rash, low_complement, increased_DNA_binding, leukopenia |
| 20 | Feng2022-M58-20220515.txt | 10.3389/fneur.2022.934335 | organic_brain_syndrome, low_complement |
| 21 | Freter1994-F46-19930701.txt | 10.1001/archinte.1994.00420040133016 | urinary_casts, hematuria, proteinuria, pyuria, low_complement |
| 22 | Geng2025-M15-20241207.txt | 10.5498/wjp.v15.i2.102259 | seizure, organic_brain_syndrome, low_complement, increased_DNA_binding |

| | | | |
|---|---|---|---|
| **23** | Gilbert2021-F26-20211015.txt | 10.1016/j.chest.2021.07.2058 | pericarditis |
| **24** | Hajihashemi2020-F20-20200215.txt | 10.1177/0961203320961473 | negative control |
| **25** | Hao2021-F28-20140620.txt | 10.1177/03000605211029766 | seizure, hematuria, proteinuria, rash, low_complement, increased_DNA_binding, thrombocytopenia, leukopenia |
| **26** | Hasan2025-F38-20250101.txt | 10.26502/acmcr.96550712 | visual_disturbance, arthritis, proteinuria, low_complement |
| **27** | Heckman2001-F82-20000415.txt | 10.1046/j.1532-5415.2001.49093.x | organic_brain_syndrome, arthritis, alopecia, low_complement, increased_DNA_binding |
| **28** | Heckman2019-F44-20180101.txt | 10.12659/AJCR.912875 | visual_disturbance, increased_DNA_binding |
| **29** | Herath2020-F15-20191015.txt | 10.1186/s41983-020-0149-9 | seizure, vasculitis, urinary_casts, hematuria, proteinuria, pyuria, low_complement, increased_DNA_binding |
| **30** | Hernando2009-F20-19680110.txt | 10.1177/0961203309104429 | lupus_headache, low_complement, increased_DNA_binding, leukopenia |
| **31** | Hernando2009-F25-20080322.txt | 10.1177/0961203309104429 | lupus_headache, low_complement |
| **32** | Hsu2011-F49-20110715.txt | 10.1111/j.1601-5215.2011.00575.x | negative control |
| **33** | Humala2024-F32-20231115.txt | 10.1016/j.crwh.2024.e00662 | pericarditis, low_complement, fever |
| **34** | Ibrahim2024-F18-20240615.txt | 10.7759/cureus.70090 | vasculitis, myositis, rash |
| **35** | Ikechukwu2023-F19-20230915.txt | 10.31579/2690-8808/173 | seizure, psychosis, hematuria, proteinuria, pyuria, rash, alopecia, pleurisy, low_complement |
| **36** | Im2002-F21-20020101.txt | 10.3341/kjo.2002.16.1.52 | visual_disturbance, arthritis, rash, low_complement, thrombocytopenia |
| **37** | Ioannidisa2018-M37-20180115.txt | doi.org/10.1016/j.ensci.2018.11.001 | CVA, low_complement, increased_DNA_binding |
| **38** | Jeon2022-F13-20210601.txt | doi.org/10.4078/jrd.2022.29.1.52 | visual_disturbance, rash, alopecia, low_complement, thrombocytopenia, leukopenia |
| **39** | Jusakos2021-F61-20210801.txt | 10.38207/JMCRCS/2021/0212184 | psychosis |
| **40** | Kato2005-F46-20030201.txt | 10.4088/pcc.v07n0610b | rash |
| **41** | Kato2005-M48-20041006.txt | 10.4088/pcc.v07n0610b | organic_brain_syndrome, myositis, urinary_casts, hematuria, proteinuria, pyuria, rash, alopecia, pericarditis, low_complement, |

| | | | |
|---|---|---|---|
| | | | increased_DNA_binding, fever |
| **42** | Khalidi2008-F33-20080101.txt | 10.1017/S0022215108001783 | cranial_nerve_disorder |
| **43** | Khan2015-F21-20150501.txt | 10.1155/2015/373201 | visual_disturbance, CVA, alopecia, thrombocytopenia |
| **44** | KishoreKumar2017-F65-20150101.txt | 10.1159/000456538 | negative control |
| **45** | KishoreKumar2017-M75-20150215.txt | 10.1159/000456538 | negative control |
| **46** | Kobayashi1992-F32-19890930.txt | 10.2169/internalmedicine.31.778 | cranial_nerve_disorder, low_complement, fever |
| **47** | Kodera2020-F40-20190501.txt | 10.1093/mrcr/rxab003 | lupus_headache, alopecia, low_complement |
| **48** | Kodera2020-F44-20190201.txt | 10.1093/mrcr/rxab003 | lupus_headache, increased_DNA_binding |
| **49** | KRAUS1990-M33-19890120.txt | 10.1136/ard.49.6.421-a | cranial_nerve_disorder, vasculitis, arthritis, hematuria, proteinuria, pyuria, mucosal_ulcers |
| **50** | KUK2014-F45-20150101.txt | 10.5606/tjr.2014.3171 | vasculitis |
| **51** | Kumar2021-F22-20201020.txt | 10.4103/jfmpc.jfmpc_1475_20 | psychosis |
| **52** | Kumaran2017-M73-20140715.txt | 10.5430/crim.v4n1p50 | organic_brain_syndrome, proteinuria, rash, low_complement, increased_DNA_binding, fever, thrombocytopenia |
| **53** | Kuroda2021_2-F78-20210430.txt | 10.12659/AJCR.932959 | cranial_nerve_disorder, arthritis, low_complement, increased_DNA_binding, fever, leukopenia |
| **54** | Kuroda2021-F78-20210430.txt | 10.12659/AJCR.932959 | cranial_nerve_disorder, arthritis, low_complement, increased_DNA_binding, leukopenia |
| **55** | Kutky2018-M74-20180101.txt | 10.1177/2054358118793397 | urinary_casts, hematuria, pyuria, low_complement |
| **56** | Kwok2008-M32-20060904.txt | 10.3346/jkms.2008.23.1.153 | low_complement |
| **57** | Lateef2007-F26-20060601.txt | 10.47102/annals-acadmedsg.V36N2p146 | visual_disturbance, rash, low_complement |
| **58** | Lee2022-F18-20220124.txt | 10.31728/jnn.2022.00112 | CVA, proteinuria |
| **59** | Li2023-F16-20240115.txt | 10.1002/iid3.1183 | CVA, low_complement, thrombocytopenia, leukopenia |
| **60** | Li2023-F25-20120220.txt | 10.1002/iid3.1183 | CVA, proteinuria, low_complement |
| **61** | Li2023-F39-20120220.txt | 10.1002/iid3.1183 | CVA, low_complement |
| **62** | Madurapperuma2025-F32-20250315.txt | 10.1186/s13256-025-05158-6 | visual_disturbance, proteinuria, rash, |

| | | | |
|---|---|---|---|
| | | | mucosal_ulcers, low_complement, fever |
| **63** | Majdak2015-F40-20150115.txt | 10.1016/j.jns.2015.12.014 | CVA |
| **64** | Makol2011-F60-20110101.txt | 10.1002/acr.20536 | negative control |
| **65** | Maloney2013-F14-20120415.txt | 10.1136/bcr-2013-010223 | lupus_headache, arthritis, hematuria, alopecia, low_complement, leukopenia |
| **66** | Maniam2020-F47-20200410.txt | 10.25270/con.2020.04.00009 | cranial_nerve_disorder, low_complement |
| **67** | Memon2020-F23-20201104.txt | 10.7759/cureus.11643 | organic_brain_syndrome, pleurisy, increased_DNA_binding |
| **68** | Mesquita2025-F55-20250125.txt | 10.24911/ejmcr.9-1606 | proteinuria |
| **69** | Nakamagoe2018-F61-20150601.txt | 10.2169/internalmedicine.1106-18 | cranial_nerve_disorder, proteinuria, low_complement |
| **70** | Narang2022-F40-20220115.txt | 10.1177/23247096221077832 | pericarditis, fever |
| **71** | Narang2022-F70-20220210.txt | 10.1177/23247096221077832 | arthritis, pericarditis, low_complement |
| **72** | Narang2022-M20-20220101.txt | 10.1177/23247096221077832 | arthritis, pleurisy, pericarditis, fever |
| **73** | Nast2006-F23-20060115.txt | 10.2215/CJN.03910907 | urinary_casts, hematuria, proteinuria, pericarditis, low_complement, fever |
| **74** | Natarajan2021-F23-20200930.txt | 10.1080/01658107.2021.1912113 | visual_disturbance, cranial_nerve_disorder, rash, low_complement, increased_DNA_binding, fever, thrombocytopenia, leukopenia |
| **75** | Nayak2012-F17-20110715.txt | 10.4103/0253-7176.96170 | psychosis, arthritis, alopecia, mucosal_ulcers, increased_DNA_binding |
| **76** | Nayak2012-F20-20110115.txt | 10.4103/0253-7176.96170 | psychosis, alopecia |
| **77** | Nayak2012-F35-20110220.txt | 10.4103/0253-7176.96170 | psychosis, alopecia, mucosal_ulcers, increased_DNA_binding |
| **78** | Nishimura2003-F55-20020122.txt | 10.2190/P2T8-Y12E-ANKM-8MWF | organic_brain_syndrome, rash, low_complement, fever, leukopenia |
| **79** | Noor2023-F65-20220101.txt | 10.1681/ASN.20233411S1274c | negative control |
| **80** | Opadola2024-F37-20200715.txt | 10.56557/jocrims/2024/v10i18916 | psychosis, hematuria, proteinuria, alopecia, pleurisy, low_complement, increased_DNA_binding |
| **81** | Patel2024-F18-20231216.txt | 10.18231/j.ijn.2024.022 | seizure, proteinuria, rash, alopecia, mucosal_ulcers, thrombocytopenia |
| **82** | Pathak2022-F12-20221020.txt | 10.1016/j.amsu.2022.104843 | negative control |
| **83** | Penarrocha2009-F55-20000301.txt | 10.1016/j.joms.2009.04.079 | negative control |

| | | | |
|---|---|---|---|
| **84** | Penarrocha2009-M27-20000115.txt | 10.1016/j.joms.2009.04.079 | negative control |
| **85** | Perez2017-F59-20171023.txt | 10.7759/cureus.1793 | organic_brain_syndrome, hematuria, proteinuria, increased_DNA_binding |
| **86** | Prasad2022-F36-20220502.txt | 10.7860/JCDR/2022/52527.16334 | organic_brain_syndrome, proteinuria, rash, thrombocytopenia |
| **87** | Pugliese2023-F19-20220915.txt | 10.1016/j.heliyon.2024.e38880 | lupus_headache, proteinuria, alopecia, low_complement, increased_DNA_binding |
| **88** | Rajasekharan2012-F14-20120415.txt | 10.1136/bcr-2012-007886 | lupus_headache, vasculitis, arthritis, low_complement, increased_DNA_binding, fever |
| **89** | Raju2021-M23-20200930.txt | 10.4103/ajim.ajim_115_22 | cranial_nerve_disorder, arthritis, pericarditis, thrombocytopenia |
| **90** | Rao2023-F24-20201001.txt | 10.1177/03000605231163716 | visual_disturbance, proteinuria, pleurisy, low_complement, increased_DNA_binding |
| **91** | Rivera2024-F25-20240925.txt | 10.1016/j.chest.2024.06.1639 | psychosis, low_complement |
| **92** | Roy2022-F51-20180315.txt | 10.1186/s41927-021-00240-0 | myositis, low_complement |
| **93** | Saini2013-F0-20131015.txt | 10.1016/j.ejpn.2014.01.007 | seizure, CVA |
| **94** | Saleh2010-F48-20070315.txt | 10.1177/0961203309345722 | cranial_nerve_disorder, increased_DNA_binding |
| **95** | Samadder2025-F25-20241215.txt | 10.1136/bcr-2024-262637 | organic_brain_syndrome, rash, low_complement, increased_DNA_binding |
| **96** | Shah2021-F42-20210715.txt | 10.47746/FMCR.2021.2504 | seizure, CVA, low_complement, increased_DNA_binding |
| **97** | Shih2019-F47-20190610.txt | 10.4081/ni.2019.8182 | CVA, low_complement, increased_DNA_binding |
| **98** | Shreemit2017-F24-20170120.txt | 10.25107/2474-1655.1337 | negative control |
| **99** | Shreemit2017-F24-20170415.txt | 10.25107/2474-1655.1337 | negative control |
| **100** | Sommerlad2015-F58-20150209.txt | 10.1136/bcr-2014-208215 | organic_brain_syndrome, rash |
| **101** | Sonkar2019-F40-20191101.txt | 10.1136/bcr-2019-230869 | vasculitis, increased_DNA_binding |
| **102** | Srimanan2022-F11-20220415.txt | 10.1177/11795476221122649 | visual_disturbance, lupus_headache, proteinuria, pyuria, rash, mucosal_ulcers, low_complement, increased_DNA_binding, fever, thrombocytopenia, leukopenia |
| **103** | Tsukamoto2019-F29-20180201.txt | 10.5606/ArchRheumatol.2019.7026 | rash, fever, thrombocytopenia |
| **104** | Varghese2015-F24-20150718.txt | Varghese, G. H., Alexande, H., & Swethalekshmi, T. N. | rash, pericarditis, fever |

| | | | |
|---|---|---|---|
| | | (2015). Case report on steroid induce psychosis. World J Pharm Pharm Sci, 4(8), 769-774. | |
| **105** | Warnatz2003-F33-20030101.txt | 10.1136/ard.62.1.50 | negative control |
| **106** | Warner2023-F30-20230115.txt | 10.1016/j.jen.2022.12.012 | seizure |
| **107** | Xie2025-F20-20250210.txt | 10.3389/fimmu.2025.1626721 | psychosis, alopecia, low_complement, leukopenia |
| **108** | Yoo2001-F30-20001015.txt | 10.3346/jkms.2001.16.6.805 | lupus_headache, rash, low_complement, increased_DNA_binding |
| **109** | Yoon2015-F20-20150120.txt | 10.12779/dnd.2015.14.1.48 | organic_brain_syndrome |
| **110** | Yu2007-F30-20020815.txt | 10.1177/0961203307080632 | cranial_nerve_disorder, low_complement, increased_DNA_binding, fever |

##### Supplementary Note 3. Step-by-step process of splitting notes into training and testing sets

The **Comprehensive SLE Clinical Dataset** was split into training and testing sets using a stratified random sampling approach with a ratio of approximately 1:1. To better preserve the distribution of all descriptors across sets, we employed the following procedure: First, all descriptors were ranked in ascending order according to their frequency of occurrence. The algorithm then iterated through this sorted list, allocating cases associated with each descriptor to either the training or testing set. This stratified process guaranteed the inclusion of every descriptor in both subsets. Moreover, all notes originating from a single patient were exclusively assigned to either the training or testing set, thereby eliminating the potential for pre-learning patterns from the same patient. All subsequent analyses of LLM performance are based on the testing set.

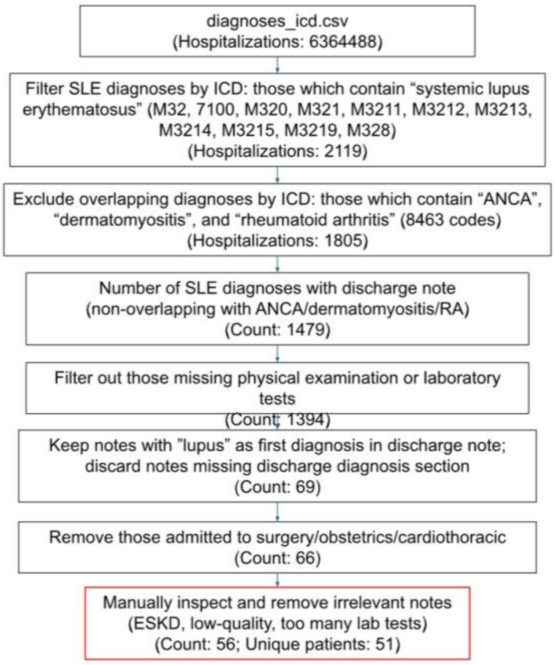

**Figure S17. Flowchart for selecting MIMIC-IV cases for annotation.** A flowchart demonstrating the step-by-step process for selecting MIMIC-IV SLE data for annotation and external validation. We first filtered hospitalization records by ICD codes containing the term “systemic lupus erythematosus”. To reduce diagnostic ambiguity, we excluded cases with overlapping autoimmune syndromes based on ICD, including ANCA, dermatomyositis, and rheumatoid arthritis, etc. From the remaining records, we selected those with associated discharge summaries and further excluded entries missing laboratory test results or physical examination documentation. Within the discharge summaries, we retained only cases where the first diagnosis item explicitly contained the term “lupus” in the discharge diagnosis section. To ensure SLE relevance, we excluded admissions to surgery, obstetrics, and cardiothoracic issues, resulting in 66 candidate cases. A final manual review was conducted to remove low-quality or irrelevant notes, such as cases unrelated to lupus, admissions not directly related to SLE, yielding a curated set of 56 discharge summaries.

**Table S12: Annotation of MIMIC-IV Discharge Note Sections for Admission SLEDAI Calculation.**

| MIMIC-IV Discharge Note Section | Selected for annotation notes (✓) |
|---|---|
| **Chief Complaints** | ✓ |
| Major Surgical or Invasive Procedure | |
| **History of Present Illness** | ✓ |
| **In ER** | ✓ |
| **Review of Systems** | ✓ |
| Past Medical History | |
| Social History | |
| Family History | |
| **Physical Exam** | ✓ |
| **Pertinent Results (admission labs, relevant labs, discharge labs)** | **✓(only admission labs and relevant labs)** |
| On discharge | |
| **Micro/Microbiology** | ✓ |
| **Imaging** | ✓ |
| Brief Hospital Course | |
| **Medications on Admission** | ✓ |
| Discharge Medications | |
| Discharge Disposition | |
| **Discharge Diagnosis** | ✓ |
| Discharge Condition | |
| Discharge Instructions | |
| Followup Instructions | |

The SLEDAI was assessed at the time of hospital admission only. This decision was made because discharge summaries are often extensive, and disease activity can change rapidly following intensive inpatient treatment. At admission, each patient underwent a comprehensive physical examination, and further laboratory data were obtained upon transfer from the emergency department. Therefore, to calculate the admission SLEDAI, we extracted the relevant clinical and laboratory data from the corresponding sections of the discharge summary. As the MIMIC notes were de-identified data without date information for all the tests, all laboratory results mentioned in the notes were considered present. **In cases of duplicate results, only the first recorded test was used for the assessment.**

#### Supplementary Note 4. Development of annotation guidelines

The **annotation guidelines** were co-developed by clinicians and an AI expert (a physician with training in rheumatology (D.Z.), three clinical experts in Rheumatology, Nephrology, and Cardiology (S.C., D.Y., and E.W., respectively), and an expert in developing and implementing LLM models in clinical settings (Z.W.)). It consists of two parts: one is the detailed definition for each descriptor in SLEDAI-2K, and the other is the annotation structure. A 30-day window for SLEDAI-2K was used instead of the original 10 days [23]. We implemented necessary refinements for the original SLEDAI-2K criteria [2] to enhance its clarity. For example, we incorporated detailed criteria from lupus classification criteria (e.g., SLICC, ACR) for rash, and the ACR nomenclature and case definitions for neuropsychiatric SLE (NPSLE). Moreover, reference ranges for lab tests applicable to QMH were provided. We also provided the exclusions and added rules on how to interpret treatment intentions. (see ***annotation guidelines Table 8 & 9***) As for the annotation structure, there were 25 named entities (24 SLEDAI-2K descriptors and Time), 8 events, 3 relations, and 4 attributes designed for the annotation. Among all the 24 descriptors from SLEDAI-2K, 16 descriptors are categorized as ***clinical*** and 8 descriptors as ***laboratory***; each type was given a distinct annotation structure reflective of their diagnostic basis. The assessment on clinical descriptors requires a comprehensive clinical appraisal: NPSLE, visual disturbance, vasculitis, arthritis, myositis, rash, alopecia, mucosal ulcers, pleurisy, pericarditis, and fever. In contrast, the remaining laboratory descriptors rely heavily on laboratory findings. (***annotation guidelines Table 2***) Using the text annotation tool Brat (http://brat.nlplab.org/), our annotators (D.Z. and Y.L.) performed annotations independently (Y.L. is an experienced researcher in the field of SLE with a medical background) following the annotation guideline. Annotated data for each case was stored in .ann files. The step-by-step annotation process and examples are provided in the ***annotation guidelines***. An inter-rater reliability (Cohen's κ) was assessed upon the completion of human annotation. Annotations with discrepancies were escalated to three senior experts (E.W., S.C., and D.Y.) for a final consensus ground truth.

**Table S13: Full names and release dates of the large language models (LLMs).**

| Model name | Release date |
|---|---|
| openai/gpt-oss-120b | August 5, 2025 |
| qwen/qwen3-32b | April 28, 2025 |
| qwen/qwen3-14b | April 28, 2025 |
| deepseek-ai/deepSeek-R1-0528-qwen3-8b | May 28, 2025 |
| meta-llama/llama-3.3-70b-instruct | December 6, 2024 |

Note: The open-weight models covering different sizes were downloaded from Huggingface[2] and evaluated in the study. For model inference, we utilized the vLLM[3] library, which facilitates the rapid inference of large language models. Inference was performed on L40S GPUs, with the number of GPUs allocated depending on the model size: Mid-sized models (e.g., Qwen3-32GB) were run on two GPUs, while larger models (e.g., Llama3.3-70B) required greater VRAM and were run on four GPUs. We used the official recommended temperature settings and a batch size of 16. The full testing set (358 cases) was used for inference. With this setting, model behaviour is equivalent to *Instruct* type of models.

[2] https://huggingface.co/

[3] https://github.com/vllm-project/vllm

#### Supplementary Note 5. Prompting strategies

We experimented with the following prompting strategies.

**Individual-descriptor prompt**: Prompts are processed by descriptor so that the LLM can focus on descriptor-specific guidelines. ***(Supplementary Prompts section 1)***

**Grouped-descriptors prompt**: Prompts are processed by groups of descriptors, determined by descriptor type. This is designed such that the model can differentiate symptoms from correlated descriptors, e.g. psychosis and organic brain syndrome, with more information about other descriptors. ***(Supplementary Prompts section 2)***

**Second-layer verifier**: This model is a LLM-as-a-judge which reads the output of the first-layer model, identifies and corrects any errors. This is only triggered when the descriptor confidence is below a threshold. ***(Confidence formula defined in Supplementary Note 7)*** In this study, we used a threshold of 5. However, it is important to note that confidence calculation can be tailored for clinical application. For example, if a more advanced LLM is utilized, the confidence threshold could be lowered. Alternatively, thresholds could be adjusted based on clinician preferences for specific descriptors. ***(Supplementary Prompts section 3)***

#### Supplementary Note 6. Model finetuning specifications

We first pretrained the BioClinicalBERT[4] model using a span-level masking strategy[5] on the annotation guidelines, allowing the model to learn domain-specific knowledge. We then finetuned the model on the training set annotated under our SLE NER framework in a supervised learning setup. A portion of the training set was held out as a validation set to guide early stopping. Training was run for a maximum of 30 epochs, and the final model was selected based on the best validation performance. A learning rate of $1e^{-4}$ and a batch size of 16 were used.
During inference, the finetuned model identifies descriptor entities and their associated attributes directly from the clinical note text. Based on extracted criteria and time attributes, we then infer the descriptor score classification for each note.

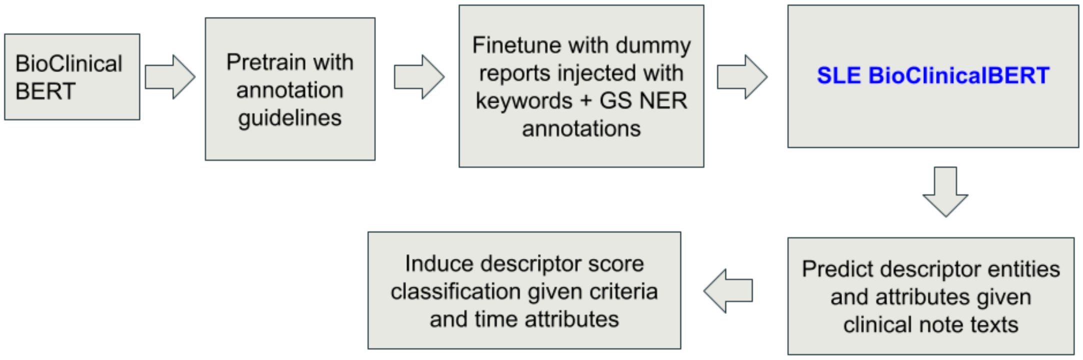

**Figure S18. BERT model architecture and training.** A schematic of the natural language processing model pipeline, based on a pre-trained BioClinicalBERT model that was subsequently fine-tuned using a combination of keyword-based sampling and Gold Standard (GS) expert annotations.

For GSPO, we employed the Unsloth[6] framework on a single L40S GPU to finetune the Qwen3-14B dense model on the training set, guided by reward signals derived from comparisons with ground truth annotations, thereby enhancing the model's reasoning quality and encouraging it to generate outputs that align more closely with the expected reasoning patterns and task objectives.

---

[4] https://huggingface.co/emilyalsentzer/Bio_ClinicalBERT
[5] Joshi, Mandar, et al. "Spanbert: Improving pre-training by representing and predicting spans." Transactions of the association for computational linguistics 8 (2020): 64-77.
[6] https://github.com/unslothai/unsloth

Format rewards encourage structural consistency to support parsing, correctness rewards reflect the overall accuracy of the descriptor classification, and reasoning rewards evaluate the model's ability to judge more fine-grained attributes such as diagnostic criteria fulfilment, temporal classification, and treatment intent. The total reward is computed as:

$$\text{Total Reward} = R_{\text{format}} + R_{\text{correctness}} + R_{\text{reasoning}} \quad (S1)$$

Where:

- Format rewards:
  - XML-formatted responses receive +0.125 for each correctly used tag (e.g., <reasoning>or <answer>)
  - Properly formatted JSON or integer responses earn +0.5
- Correctness reward: +2 if descriptor classification answer is correct; −2 if incorrect
- Reasoning rewards:
  - NE identification: +1 if identified; -1 if missing
  - Criteria fulfilment judgment: +1 if correct; -1 if incorrect
  - Time classification (e.g., within 30 days or not): +1 if correct; -1 if incorrect
  - Intention-to-treat judgment: +1 if correct; −1 if incorrect

A critical aspect in GSPO training is the need for balanced exposure to both positive and negative samples to guide policy learning. To address the class imbalance in the data and ensure consistent exposure to informative training signals, we constructed a specialized subset from the training set. For each query descriptor, we curated a balanced mix of samples, maintaining a 1:1:1 ratio between positive, relevant, but negative, and irrelevant samples. To prevent overrepresentation of more common descriptors, we imposed a cap of 20 samples per category. The resulting distribution of samples across categories and descriptors is presented in Figure S7. Training was run for 1800 steps using a learning rate of 5e-5.

(a)

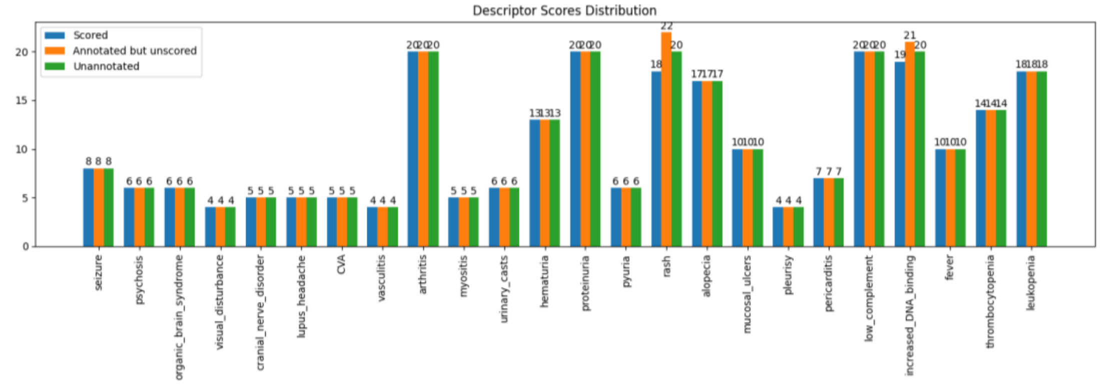

(b)

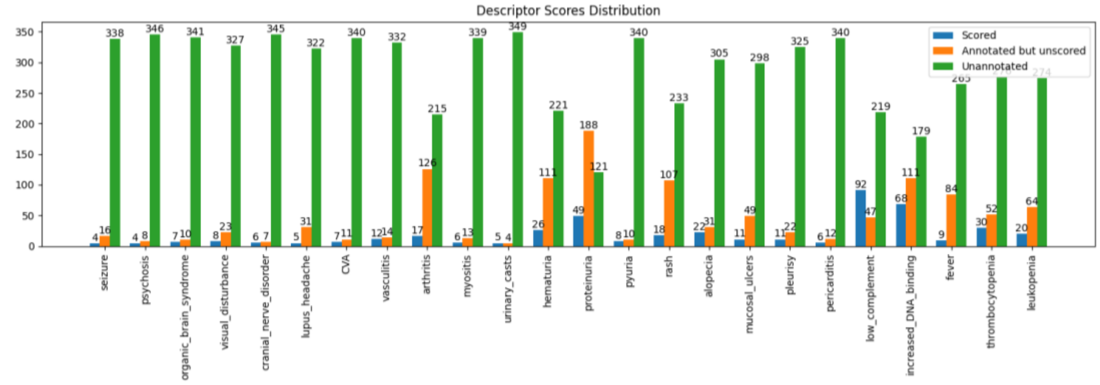

**Figure S19. Train-test split for GSPO reinforcement learning.** (a) For GSPO finetuning, we constructed a subset from the training set using a balanced 1:1:1 ratio of three sample categories: positive samples, relevant but negative samples, and irrelevant samples, with a maximum of 20 samples in each category. This stratified sampling strategy ensures that the reinforcement learning model encounters sufficient examples from each category. (b) For evaluation, the full testing set was used, reflecting the performance under the natural distribution of the dataset.

#### Supplementary Note 7. Computation of case confidence

Below are the formulae for the descriptor confidence and case confidence to measure reliability and certainty of the LLM. We adopted a confidence normalization approach for computing descriptor confidence. This addresses the need to account for minority signals and ensure they are not overlooked, aligning with concerns raised in prior work[7] [1]. By considering both majority and minority confidences simultaneously, the formula provides a straightforward way to measure the dominance of majority confidence relative to competing evidence. This approach also resembles other verbalized confidence estimation methods[8,9] [2,3], where confidence is derived from the normalized probability of positive vs negative confidence tokens. In the below example of dsDNA, we observe high confidence scores in both majority and minority outputs, creating a contradiction that weakens the reliability of the majority outcome. Thus, the normalization adjusts this contradiction in the resulting confidence score. To avoid inflating the overall case confidence, we limited the calculation to descriptors predicted as relevant. This is because the model often assigns high confidence to irrelevant descriptors, mainly due to their absence in the note.

**If no minority votes (i.e. agreement = 1):**

$$\text{Descriptor Confidence} = \text{Average confidence}_{\text{majority}} \qquad (S2)$$

**If a minority vote exists:**

$$\text{Descriptor Confidence} = 10 \cdot \frac{\text{Average confidence}_{\text{majority}}}{\text{Average confidence}_{\text{majority}} + \text{Average confidence}_{\text{minority}}} \qquad (S3)$$

**If all descriptors are predicted as negative:**

$$\text{Case Confidence} = \frac{1}{24}\sum_{i=1}^{24} C(d_i) \qquad (S4)$$

**If at least one descriptor is predicted *relevant*:**

$$\text{Case Confidence} = \frac{1}{N_+}\sum_{d_i \in D_+} C(d_i) \qquad (S5)$$

Where:

- $C(d_i)$ is the confidence score for the descriptor $d_i$
- $D_+ \subseteq D$ is the set of descriptors predicted ***relevant*** within a case
- $N_+ = |D_+|$ is the number of descriptors predicted ***relevant*** within a case

**Demo case**

**Proteinuria**

| | | | | | |
|---|---|---|---|---|---|
| Relevancy | 1 | 1 | 1 | 1 | 1 |
| Score | 1 | 1 | 1 | 1 | 0 |
| Confidence | 10 | 8 | 9 | 9 | 2 |

Final relevancy: 1
Final score: 1
Average majority confidence: 9
Average minority confidence: 2
Descriptor confidence: 10 * 9/(9+2)=8.2

---

[7] Huang, S., et al. Mirror-Consistency: Harnessing Inconsistency in Majority Voting. 2024. Miami, Florida, USA: Association for Computational Linguistics.
[8] Kapoor, S., et al., Large language models must be taught to know what they don't know. Advances in Neural Information Processing Systems, 2024. 37: p. 85932–85972.
[9] Yeom, J., et al., EpiCaR: Knowing What You Don't Know Matters for Better Reasoning in LLMs. arXiv preprint arXiv:2601.06786, 2026.

**C3**
Final score: 1
Final relevancy: 1
Descriptor confidence: 6

**Leukopenia**
Final score: 0
Final relevancy: 1
Descriptor confidence: 5.5

**dsDNA**

| | | | | | |
|---|---|---|---|---|---|
| Relevancy | 1 | 1 | 1 | 0 | 1 |
| Score | 1 | 1 | 0 | 0 | 0 |
| Confidence | 10 | 8 | 9 | 9 | 9 |

Final relevancy: 1
Final score: 0
Average majority confidence: 9
Average minority confidence: 9
Descriptor confidence: 10 * 9/(9+9)=5.0

**Other 20 descriptors….**

| | | | | | |
|---|---|---|---|---|---|
| Relevancy | 0 | 0 | 0 | 0 | 0 |
| Score | 0 | 0 | 0 | 0 | 0 |
| Confidence | 10 | 10 | 10 | 10 | 10 |

Final relevancy: 0
Final score: 0
Descriptor confidence: 10

---

Case confidence = (8.2 + 6.0 + 5.5 + 5.0)/4=6.18

#### Supplementary Note 8. From the output to final SLEDAI score

In NER, entity detection measures whether the model correctly identifies which entities are relevant or irrelevant within the input clinical note, while scoring classification measures whether the model assigns the correct binary score to each entity. For SLEDAI-2K score prediction, we computed the total score for each clinical note as follows:

$$\text{SLEDAI-2K} = \sum_{d \in D} \widehat{y_d} \cdot w_d \qquad (S6)$$

Where $\widehat{y_d} \in \{0, 1\}$ is the model-predicted binary classification of descriptor $d$, and $w_d$ is the predefined weight assigned to descriptor $d$ in the SLEDAI-2K scoring system.

Clinical SLEDAI-2K excludes increased DNA binding and low complement. Renal SLEDAI-2K includes four renal descriptors: hematuria, pyuria, proteinuria, and urinary casts.

#### Supplementary Note 9. Model evaluation metrics

LLMs' performance on descriptor concept relevancy, descriptor classification and total SLEDAI-2K score was assessed on the testing set and the MIMIC-IV validation set. Individual descriptor performance including relevancy and classification was evaluated using F1, precision, and recall scores. These metrics were chosen due to negative cases (including cases not mentioned, mentioned but explicitly negated, and mentioned but criteria unfulfilled) significantly outnumbering positive cases for each descriptor. **For evaluating cross-descriptor and overall performance, micro-F1 serves as the primary metric.** For total SLEDAI-2K score, we compute the proportion of samples exactly matching the ground truth, as well as within 1 and 2 tolerance of the ground truth. Additionally, the root mean squared error (RMSE) was used to quantify the prediction error between the predictions and the ground truths. We classified model errors into three primary categories: inference errors, extraction errors, and hallucinations.

Precision is the proportion of true positive predictions among all positive classifications, quantifying prediction reliability. Lower precision indicates more false positives.

$$Precision = \frac{TP}{TP + FP} \qquad (S7)$$

Recall is the proportion of actual positives correctly identified, also called sensitivity. Lower recall indicates more false negatives.

$$Recall = \frac{TP}{TP + FN} \qquad (S8)$$

The F1 score provides a balanced performance metric of precision and recall.

$$F1Score = 2 \times \frac{Precision \times Recall}{Precision + Recall} \qquad (S9)$$

Micro F1 pools all predictions, giving more weight to larger classes in the dataset, reflecting overall performance in proportion to the natural frequency of each descriptor:

$$MicroF1 = 2 \times \frac{(Micro\ Precision \times Micro\ Recall)}{Micro\ Precision + Micro\ Recall} \qquad (S10)$$

Where: $MicroPrecision = \frac{\sum TP_i}{\sum TP_i + \sum FP_i}, MicroRecall = \frac{\sum TP_i}{\sum TP_i + \sum FN_i}$

TPi is the true positive for each descriptor i
FPi is the false positive for descriptor i
FNi is the false negative for descriptor i

AUROC is the area under the curve in the plot of True Positive Rate (TPR) against the False Positive Rate (FPR) across all classification thresholds.

$$AUROC = \int_0^1 TPR(FPR)\ d(FPR) \qquad (S11)$$

Where: $TPR = \frac{TP}{TP+FN}, FPR = \frac{FP}{FP+TN}$

The total weight of misclassified descriptors is computed as follows:

$$\text{Total weight of misclassified descriptors} = \sum_i^M \sum_{d \in D} w_d \cdot 1\{\widehat{y_{d,i}} \neq y_{d,i}\} \qquad (S12)$$

Where: $\widehat{y_{d,i}}$ is the predicted descriptor classification for descriptor d in case i
$y_{d,i}$ is the ground-truth descriptor classification for descriptor d in case i
$w_d$ is the predefined SLEDAI-2K weight assigned to descriptor $d$
M is the total number of cases

The ***exact match of a SLEDAI-2K score*** refers to the total weight of misclassified descriptors equalling 0, whereas a ***tolerance within k*** refers to the total weight of misclassified descriptors ≤ k.

The root mean squared error (RMSE) was used to quantify the prediction error between the total SLEDAI-2K scores predicted by the LLMs and the ground truth values.

$$\text{RMSE} = \sqrt{\frac{1}{M}\sum(\hat{y_i} - y_i)^2} \qquad (S13)$$

Where $\hat{y_i}$ is the predicted SLEDAI-2K score for case i, $y_i$ is the ground-truth SLEDAI-2K score for case i, M is the total number of cases.

#### Supplementary Note 10. Error analysis

**Inference errors** were characterized by flawed logical reasoning and were further sub-categorized into errors regarding criteria, temporal reasoning, unit conversions, exclusions, and intention to treat, etc. **Extraction errors** occurred when the model omitted key information or pulled flawed information from the source text. **Hallucinations** were identified as instances where the model generated content that did not exist in the provided context. Finally, a **grey area** arises when the LLM's reasoning is plausible but diverges from the ground truth, typically due to ambiguity in the description or the existence of an alternative explanation.

#### Supplementary Note 11. Simulated clinical implementation

The time to first stage 3 chronic kidney disease (CKD) was defined as the date when a patient's estimated glomerular filtration rate (eGFR) was first documented to be persistently below 60 ml/min/1.73m² for at least 3 months, without intervening recovery to eGFR ≥ 60 ml/min/1.73m² during this period. We used eGFR values (calculated via either the MDRD or CKD-EPI equations) provided by the central laboratory. For visits with serum creatinine but missing the central calculated eGFR, eGFR was calculated using the 2021 CKD-EPI creatinine equation[10]. Kidney replacement therapy (KRT) is the initiation of hemodialysis, peritoneal dialysis, or kidney transplantation.
Baseline data are presented as mean ± standard deviation (SD), or non-normally distributed data as median with interquartile range [IQR], based on their distribution as assessed by the Kolmogorov-Smirnov test. Comparisons of these continuous variables were performed using the t-test or the Mann-Whitney U test, as appropriate. Categorical variables were compared using the Chi-square test or Fisher’s exact test, as appropriate.

---

[10] Inker, L. A., Eneanya, N. D., Coresh, J., Tighiouart, H., Wang, D., Sang, Y., ... & Levey, A. S. (2021). New creatinine-and cystatin C–based equations to estimate GFR without race. New England Journal of Medicine, 385(19), 1737-1749.

### Supplementary Prompts

First, we employed a prompt template that can be applied to each descriptor, which allows the model to focus narrowly on descriptor-specific information, including associated keywords, diagnostic criteria, and exclusion rules. Second, we experimented with grouping descriptors by type (e.g., clinical and laboratory) and evaluating them collectively within a single prompt. This approach helps the model leverage contextual relationships among related descriptors, such as distinguishing between "rash" and "vasculitic rash," or "psychosis" and "organic brain syndrome", thereby reducing ambiguity. Finally, we incorporated a second verification layer to identify and correct potential errors from the initial outputs. This step aims to mitigate the model's tendency to overlook details or produce misclassifications when processing information-dense and complex clinical descriptions. The model was instructed to return its answer in JSON format, from which we parsed and extracted NE and scoring classification results. Moreover, to facilitate error analysis by clinicians, the model was mandated to output its chain-of-thought reasoning alongside its confidence score (ranging from 0 to 10) for each descriptor and total SLEDAI-2K score.

As mentioned in the main study, we employed error-feedback workflows to guide the design and refinement of prompts. This involved systematically analyzing the model's outputs to identify common recurring errors, which often arose either from insufficient domain knowledge or from misinterpretation of task-specific rules. These errors were then grouped into categories to enable targeted corrective strategies. Importantly, the refinement process was deliberately constrained to these common, categorized errors to prevent excessive over-engineering of prompts. For example, in the preliminary experiments, specialized clinicians analyzed some erroneous examples and found that the model did not have enough knowledge to distinguish between rash caused by SLE versus other reasons. Therefore, we added a formal vocabulary list in the diagnostic criteria for rash to provide sufficient information, such as erythematous versus scaling versus hyperpigmentation versus scarring.

Below are the finalized prompts used in the study. The Haystack[11] library was used to systematically construct prompts from the templates. Each prompt was populated with the descriptor name, relevant keywords, and diagnostic information of each descriptor, as well as the date of the clinical note and the main text body.

##### 1. Prompt template for individual descriptor

The universal, high-level instructions are provided to the language models to ensure consistent output style and format across all tasks. Each prompt targets a single descriptor within a clinical note.

```
You specialize in evaluating clinical notes for the Systemic lupus erythematosus (SLE) disease.
Given the descriptor {{ descriptor }}, score it as 0 or 1 based on the clinical note and information.
Please follow this guideline carefully:
1. Diagnostic criteria:
Given the information for {{ descriptor }}, assess the clinical note for evidence and evaluate
whether the diagnostic criteria are met. Possible areas include:
- Diagnostic keywords: Check for the presence of diagnostic keywords which are strong indicators
confirming criteria fulfilment. (In such cases, it is not necessary to verify each point within the
criteria)
- Symptoms/signs keywords: Check for the presence of specific symptoms or signs
- Tests: Check for the value and unit of test results (check latest values; use default units if
unspecified). And assess whether they meet criteria thresholds.
Relevancy check
- If no keywords are mentioned => `relevant = false`, `score = 0`, and skip remaining steps.
- If keywords are mentioned => `relevant = true`, and continue this guideline.
```

[11] https://github.com/deepset-ai/haystack

[8-point items] - Intention to treat: The doctor's treatment plans must include the initiation or escalation of steroids (e.g. prednisone, prednisolone, glucocorticoids), immunosuppressants (e.g. mycophenolate mofetil, cyclophosphamide, azathioprine, cyclosporine, tacrolimus), or biologics (e.g. belimumab, rituximab).
Classify the nature_of_intention_to_treat:
- 'treat_escalated' (lupus-specific treatment is initiated or escalated)
- 'treat_de_escalate' (treatment is reduced, tapered, or discontinued)
- 'wait_and_see' (no treatment change is made; intent is to monitor and reassess)
Important: If diagnostic keywords are mentioned or the diagnostic criteria are met, but `nature_of_intention_to_treat` is not treat_escalated, then the criteria should be considered unfulfilled_diagnostic (see below).

[non 8-point items] - Treatment response: For cases of {{ descriptor }} where documented positive symptoms/signs/tests do not not fully meet diagnostic thresholds, criteria could be fulfilled if either:
(a) a documented positive response to lupus-directed therapeutic trials (e.g. steroids, antimalarials, immunosuppressants, or biologics), or
(b) doctor's stated intention to initiate or escalate lupus-directed treatment.
Exception: Rash that improves with steroids alone is insufficient to establish lupus etiology.

Exclusions:
- Look for exclusions in the clinical note that may explain the symptom, but not SLE itself. These exclusions may justify scoring the descriptor but make the criteria `unfulfilled_diagnostic`.
- Only count exclusions if they are explicitly written in the note. If not mentioned, assume no exclusions.
- Drug exclusions only apply if the symptom is a common side effect, or if the doctor suspects the drug caused it.
If `relevant = true`, classify the diagnostic criteria as:
- 'fulfilled' (clear evidence the descriptor is met)
- 'unfulfilled_negated' (the absence of symptoms/diagnosis is explicitly stated in the clinical note)
- 'unfulfilled_diagnostic' (not enough evidence)
- 'uncertain' (doctor is uncertain or diagnostic ambiguity)

Notes and nuances:
- Terms such as "decrease/subsided/less/reduced/improved/similar/no increase in" still imply current symptom presence
- '?' or 'vs' indicates physician uncertainty, so criteria should be 'uncertain'
- If multiple values are present for the same test, use the most recent. Watch for collection dates or historical references (terms like 'last'/'prev'). For results 'pending', it is uncertain.
- If multiple reference ranges exist for the descriptor, check the report date and apply the reference range that corresponds to the time of testing
- Scan the entire clinical note for all mentions of descriptor keywords. Do not stop at the first mention.
- If keywords appear in multiple parts of the note, check whether they refer to the same time frame or episode. If yes, evaluate them together. If no, otherwise, evaluate each mention independently.
- If any independent mention fulfills the criteria, it can support final scoring of the descriptor.
Additional tips:
- Core lupus drug abbreviations include HCQ (hydroxychloroquine), P/PRED (prednisone), MMF/MPA (mycophenolate), AZA (azathioprine), BEL (belimumab), LEF(Leflunomide), RTX (rituximab), CTX/CYC (cyclophosphamide), TAC/FK506/FK (tacrolimus), and CsA/CYA (cyclosporine).
- 'P' followed by a number (e.g. P5, P7.5, P20) refers to prescribing and administering oral prednisone at the specified dose in milligrams (mg).
- The term 'flare' in clinical impressions acts as a diagnostic trigger: it confirms active manifestations fulfilling specific SLEDAI descriptors (e.g. cutaneous flare fulfills lupus rash).
- In medical documentation, X/52 denotes X weeks and X/12 denotes X months, reflecting durations based on yearly cycles (52 weeks or 12 months).(e.g., 2/52 = 2 weeks, 1/12 = 1 month).

- The value "trace" for test results means +/- (neither positive nor negative)

2. Time frame:
After identifying relevant keywords, symptoms, or test results in the diagnostic assessment, determine how recently the identified entity or finding occurred.
Time classification should reflect the current status of the symptom as of the documentation date, not when it first began. For example, if the note states 'onset 3 months ago' or '3 months already', it implies the symptom has been present continuously and is still active currently, so the time is still considered within_10days.
If the note explicitly states that the symptom has resolved, then the time should reflect when it was last active.
Assign one of the following categories from the date of the clinical note:
- 'within_10days' (descriptor is active within the last 10 days)
- '11_to_30days' (active more than 10 days ago but less than 30 days ago)
- '30days_ago' (entity was present 30 days ago)
- 'time_uncertain' (timing of entity cannot be determined)
Tips:
- Use context from the same part of the note where the descriptor was identified
- "recent/frequent/recurrent" => within_10days
- "occasional/sometimes/last visit/last time/on and off" => time_uncertain
- if only month and year provided: if match note date => 11_to_30days; otherwise => time_uncertain
- if no date is specified => default to within_10days unless indications suggest otherwise
Note that all dates mentioned should follow the format dd/mm/yyyy, unless it doesn't make sense.
For time_uncertain or 30days_ago, the descriptor score should be 0.

3. Score calculation:
Descriptor `score = 1` only if both:
(1) criteria must be 'fulfilled'
(2) time is either 'within_10days' or '11_to_30days'
Output the score in JSON format.
Include:
- "confidence_score": a number from 0 to 10 indicating how confident you are that your predicted score (0 or 1) accurately reflects whether the diagnostic criteria are met, based solely on the information available in the clinical note.
  - 10 = You are fully confident your prediction is correct, based on clear and direct evidence in the note. This includes explicit confirmation of the criteria (for score 1), explicit negations or absence of relevant keywords (for score 0).
  - 0 = You have no confidence in your prediction due to ambiguity, conflicting statements, vague language, or missing context in the note.
  - Scores between 1-9 reflect varying degrees of uncertainty, partial evidence, indirect phrasing, or incomplete documentation.
  - Note: Your confidence should reflect the clarity and completeness of the clinical note — not the doctor's intent, external medical knowledge, or pending test results. For example:
    - If the note says "? alopecia", you may be highly confident that the physician is uncertain, and therefore confident in assigning score 0.
    - If a test is mentioned but no result is provided, you may be confident that the criteria are not met, even if the test result is pending.
Your confidence should reflect how well the note supports your prediction — not whether the criteria are truly met in reality.
- "rationale": a 1-3 sentence explanation of the score. Mention relevant symptoms/signs, tests, any exclusions, time, etc. that helped you conclude this score. Use exact quoted text if citing clinical notes.

Diagnostic information for descriptor {{ descriptor }}:
{{ information }}

```
{{ keywords }}

=============================================================
The clinical note is dated {{ date }} (dd/mm/yyyy).

Clinical Note:
{{ clinical_note }}

======================================================
Output JSON format:
```
{
    'descriptor': '{{ descriptor }}', // required
    'relevant': true or false, // required; write true if keywords are mentioned in the clinical note
    'criteria': 'unfulfilled_negated' | 'unfulfilled_diagnostic' | 'fulfilled' | 'uncertain' | null, // write null if 'relevant' field is false
    {{ nature_of_intention_to_treat }}'time': 'within_10days' | '11_to_30days' | '30days_ago' | 'time_uncertain' | null, // write null if 'relevant' field is false
    'score': 0 or 1, // required
    'confidence_score': 0 to 10, // required
    'rationale': '' // required
}
```
```

Note: In the MIMIC-IV data, SLEDAI-2K is assessed at the time of admission instead of the date of the clinical note. Thus, the prompt template for the MIMIC-IV data gives additional details on interpreting time for discharge notes, such as ED vs hospital admission.

```
Discharge note rules:
- In the history of present illness section: if timing is not explicitly stated (e.g. "one week ago"), classify as time_uncertain.
- In the emergency department (ED) documentation: unless otherwise specified, classify findings as within_10days
- At hospital admission (or on the floor): findings are prioritized and should be classified as within_10days.
```

#### 2. Prompt template for grouped descriptors

Similar to the individual descriptor prompt format, the group-based prompt approach organizes descriptors by type: clinical descriptors are grouped in one prompt, and laboratory descriptors in another. This structure allows the model to jointly reason over related descriptors.

```
You specialize in evaluating clinical notes for the Systemic lupus erythematosus (SLE) disease.

Given the provided descriptors, score each as 0 or 1 based on the clinical note and information.

Descriptors to evaluate: {{ descriptor_list }}

For each descriptor, please follow this guideline carefully:

1. Diagnostic criteria:

Given the information for the descriptor, assess the clinical note for evidence and evaluate whether
the diagnostic criteria are met. Possible areas include:

- Diagnostic keywords: Check for the presence of diagnostic keywords which are strong indicators
confirming criteria fulfilment. (In such cases, it is not necessary to verify each point within the
criteria)

- Symptoms/signs keywords: Check for the presence of specific symptoms or signs

- Tests: Check for the value and unit of test results (check latest values; use default units if
unspecified). And assess whether they meet criteria thresholds.

Relevancy check

- If no keywords are mentioned => `relevant = false`, `score = 0`, and skip remaining steps.

- If keywords are mentioned => `relevant = true`, and continue this guideline.

(For 8-point descriptors only) - Intention to treat: The doctor's treatment plans must include the
initiation or escalation of steroids (e.g. prednisone, prednisolone, glucocorticoids),
immunosuppressants (e.g. mycophenolate mofetil, cyclophosphamide, azathioprine, cyclosporine,
tacrolimus), or biologics (e.g. belimumab, rituximab).

Classify the nature_of_intention_to_treat:

- 'treat_escalated' (lupus-specific treament is initiated or escalated)

- 'treat_de_escalate' (treament is reduced, tapered, or discontinued)

- 'wait_and_see' (no treatment change is made; intent is to monitor and reassess)

Important: If diagnostic keywords are mentioned or the diagnostic criteria are met, but
`nature_of_intention_to_treat` is not treat_escalated, then the criteria should be considered
unfulfilled_diagnostic (see below).
```

(For non 8-point descriptors only) - Treatment response: For cases of descriptors where documented positive symptoms/signs/tests do not not fully meet diagnostic thresholds, criteria could be fulfilled if either:

(a) a documented positive response to lupus-directed therapeutic trials (e.g. steroids, antimalarials, immunosuppressants, or biologics), or

(b) doctor's stated intention to initiate or escalate lupus-directed treatment.

Exception: Rash that improves with steroids alone is insufficient to establish lupus etiology.

Exclusions:

- Look for exclusions in the clinical note that may explain the symptom, but not SLE itself. These exclusions may justify scoring the descriptor but make the criteria `unfulfilled_diagnostic`.

- Only count exclusions if they are explicitly written in the note. If not mentioned, assume no exclusions.

- Drug exclusions only apply if the symptom is a common side effect, or if the doctor suspects the drug caused it.

If `relevant = true`, classify the diagnostic criteria as:

- 'fulfilled' (clear evidence the descriptor is met)

- 'unfulfilled_negated' (the absence of symptoms/diagnosis is explicitly stated in the clinical note)

- 'unfulfilled_diagnostic' (not enough evidence)

- 'uncertain' (doctor is uncertain or diagnostic ambiguity)

Notes and nuances:

- Terms such as "decrease/subsided/less/reduced/improved/similar/no increase in" still imply current symptom presence

- '?' or 'vs' indicates physician uncertainty, so criteria should be 'uncertain'

- If multiple values are present for the same test, use the most recent. Watch for collection dates or historical references (terms like 'last'/'prev'). For results 'pending', it is uncertain.

- If multiple reference ranges exist for the descriptor, check the report date and apply the reference range that corresponds to the time of testing

- Scan the entire clinical note for all mentions of descriptor keywords. Do not stop at the first mention.

- If keywords appear in multiple parts of the note, check whether they refer to the same time frame or episode. If yes, evaluate them together. If no, otherwise, evaluate each mention independently.

- If any independent mention fulfills the criteria, it can support final scoring of the descriptor.

Additional tips:

- Core lupus drug abbreviations include HCQ (hydroxychloroquine), P/PRED (prednisone), MMF/MPA (mycophenolate), AZA (azathioprine), BEL (belimumab), LEF(Leflunomide), RTX (rituximab), CTX/CYC (cyclophosphamide), TAC/FK506/FK (tacrolimus), and CsA/CYA (cyclosporine).

- 'P' followed by a number (e.g. P5, P7.5, P20) refers to prescribing and administering oral prednisone at the specified dose in milligrams (mg).

- The term 'flare' in clinical impressions acts as a diagnostic trigger: it confirms active manifestations fulfilling specific SLEDAI descriptors (e.g. cutaneous flare fulfills lupus rash).

- In medical documentation, X/52 denotes X weeks and X/12 denotes X months, reflecting durations based on yearly cycles (52 weeks or 12 months).(e.g., 2/52 = 2 weeks, 1/12 = 1 month).

- The value "trace" for test results means +/- (neither positive nor negative)

2. Time frame:

After identifying relevant keywords, symptoms, or test results in the diagnostic assessment, determine how recently the identified entity or finding occurred.

Time classification should reflect the current status of the symptom as of the documentation date, not when it first began. For example, if the note states 'onset 3 months ago' or '3 months already', it implies the symptom has been present continuously and is still active currently, so the time is still considered within_10days.

If the note explicitly states that the symptom has resolved, then the time should reflect when it was last active.

Assign one of the following categories from the date of the clinical note:

- 'within_10days' (descriptor is active within the last 10 days)

- '11_to_30days' (active more than 10 days ago but less than 30 days ago)

- '30days_ago' (entity was present 30 days ago)

- 'time_uncertain' (timing of entity cannot be determined)

Tips:

- Use context from the same part of the note where the descriptor was identified

- "recent/frequent/recurrent" => within_10days

- "occasional/sometimes/last visit/last time/on and off" => time_uncertain

- if only month and year provided: if match note date => 11_to_30days; otherwise => time_uncertain

- if no date is specified => default to within_10days unless indications suggest otherwise

Note that all dates mentioned should follow the format dd/mm/yyyy, unless it doesn't make sense.

For time_uncertain or 30days_ago, the descriptor score should be 0.

3. Score calculation:

Descriptor `score = 1` only if both:

(1) criteria must be 'fulfilled'

(2) time is either 'within_10days' or '11_to_30days'

Output the score in JSON format.

Include:

- "confidence_score": a number from 0 to 10 indicating how confident you are that your predicted score (0 or 1) accurately reflects whether the diagnostic criteria are met, based solely on the information available in the clinical note.

  - 10 = You are fully confident your prediction is correct, based on clear and direct evidence in the note. This includes explicit confirmation of the criteria (for score 1), explicit negations or absence of relevant keywords (for score 0).

  - 0 = You have no confidence in your prediction due to ambiguity, conflicting statements, vague language, or missing context in the note.

  - Scores between 1-9 reflect varying degrees of uncertainty, partial evidence, indirect phrasing, or incomplete documentation.

  - Note: Your confidence should reflect the clarity and completeness of the clinical note — not the doctor's intent, external medical knowledge, or pending test results. For example:

    - If the note says "? alopecia", you may be highly confident that the physician is uncertain, and therefore confident in assigning score 0.

    - If a test is mentioned but no result is provided, you may be confident that the criteria are not met, even if the test result is pending.

Your confidence should reflect how well the note supports your prediction — not whether the criteria are truly met in reality.

- "rationale": a 1-3 sentence explanation of the score. Mention relevant symptoms/signs, tests, any exclusions, time, etc. that helped you conclude this score. Use exact quoted text if citing clinical notes.

```
Diagnostic information for each descriptor:

{{ information }}

'''

note_template = '''

==========================================================

The clinical note is dated {{ date }} (dd/mm/yyyy).

Clinical Note:

{{ clinical_note }}

===================================================

Output JSON format:

```

[

    {

        'descriptor': '{ descriptor }', // required

        'relevant': true or false, // required; write true if keywords for the descriptor are mentioned in
the clinical note

        'nature_of_intention_to_treat': 'treat_escalated' | 'wait_and_see' | 'treat_de_escalate' | null, //
write null only if non 8-point descriptor or if 'relevant' field is false

        'criteria': 'unfulfilled_negated' | 'unfulfilled_diagnostic' | 'fulfilled' | 'uncertain' | null, // write null
only if 'relevant' field is false

        'time': 'within_10days' | '11_to_30days' | '30days_ago' | 'time_uncertain' | null, write null only if
'relevant' field is false

        'score': 0 or 1, // required

        'confidence_score': 0 to 10, // required

        'rationale': '' // required

    },

    // add more descriptors
```
```

]

```
```

#### 3. Verifier prompt template

The second-layer verifier prompt serves as a quality control mechanism within the LLM pipeline. It aims to evaluate outputs generated by the first-layer model, identify and correct potential errors, and output the final response. The prompt takes as input (i) the rules – including the general guidelines, descriptor diagnostic criteria, and keywords, (ii) the original clinical note, and (iii) the output of the first-layer model.

```
You are a verifier tasked with ensuring that a model-generated output complies with a set of predefined rules.

Below are the rules:

{{ guidelines (same as rules defined in the general prompt template) }}

Common errors to watch for (not limited to):

- Time classification: terms like "onset" indicate symptoms are still active.

- Diagnostic keyword interpretation: the presence of diagnostic keywords may be sufficient to mark criteria_fulfilled, even if not all formal criteria elements are explicitly listed in the note.

- Time and criteria validation: To support `score = 1`, time must fall within either within_10days or 11_to_30days, and the corresponding criteria must be fulfilled.

- Incomplete keyword scanning: ensure all relevant keyword mentions are identified across the entire clinical note, not just in one area.

========================================

Here is the clinical note dated {{ date }} (dd/mm/yyyy):

{{ clinical_note }}

========================================

Here is the model output to evaluate:

{{ model_output }}

========================================

Your task:

1. Review the output against each rule.
```

| |
|---|
| 2. Identify any errors or omissions.<br><br>3. Correct the output to fully comply with the rules.<br><br>4. Return the final corrected output only.<br><br>Important:<br><br>- The final output must follow the same format as the original.<br><br>- If the original output is already correct, return it unchanged. |

#### 4. Descriptor information prompt

The diagnostic information is used to guide prompt construction for each descriptor. For each descriptor, we include (i) the core diagnostic criteria based on clinical definitions and the annotation guidelines, and (ii) exclusion rules specifying conditions or contexts in which the descriptor criteria should not be fulfilled.

| | |
|---|---|
| seizure | Diagnostic criteria:<br>A with or without the presence of B:<br>A. Independent description of seizure by a reliable witness<br>B. EEG abnormalities<br><br>Exclusions:<br>- Medications: quinolones, imipenem<br>- Alcohol and drug withdrawal (phenothiazines, antipsychotics)<br>- electrolyte imbalance (acidosis, serum sodium, calcium, and hypoglycemia)<br>- organ failure (hepatic and renal failure)<br>- infection<br>- due to past irreversible CNS damage<br>- Vasovagal syncope<br>- Cardiac syncope<br>- Hysteria<br>- Hyperventilation<br>- Tics<br>- Narcolepsy and cataplexy<br>- Labyrinthitis<br>- Subarachnoid hemorrhage<br>- Trauma<br>- Panic attacks, conversion disorders, and malingering<br><br>- hypersensitivity encephalopathy |
| psychosis | Diagnostic criteria:<br>All of the following:<br>A. At least one of the following:<br>1. Delusions<br>2. Hallucinations without insight (visual, olfactory, gustatory, tactile, or auditory)<br>3. Disorganized thinking (speech) (frequent derailment; loose associations; incoherence; “word salad”)<br>4. Grossly disorganized or abnormal motor behavior (childlike “silliness”; unpredictable agitation; catatonic behavior; negativism; |

| | |
|---|---|
| | mutism and stupor; catatonic excitement; stereotyped movements, staring; grimacing, mutism; echoing of speech)<br>5. Negative symptoms (diminished emotional expression; avolition; alogia; anhedonia; asociality)<br><br>B. The disturbance causes clinical distress or impairment in social, occupational, or other relevant areas of functioning.<br>C. The disturbance does not occur exclusively during the course of a delirium.<br>D. The disturbance is not better accounted for by another mental disorder (e.g., mania).<br>Exclusions:<br>- Uremia (BUN > 40 mg/dL)<br>- Substance- or drug-induced psychotic disorder (including NSAIDs, antimalarials)<br>- acidosis, or electrolyte imbalance<br>- delirium<br>- Primary psychotic disorder unrelated to SLE (e.g., schizophrenia)<br>- Psychologically mediated reaction to SLE (brief reactive psychosis with major stressor) |
| organic_brain_syndrome | Diagnostic Synonymous:<br>organic brain syndrome<br>organic brain disorder<br>acute confusional state<br>delirium<br>encephalopathy<br>neurocognitive disorders<br><br>Diagnostic criteria:<br>All of the following:<br>A. A disturbance in attention (i.e., reduced ability to direct, focus, sustain, and shift attention) or awareness (reduced orientation to the environment)<br>B. Rapid onset and fluctuating clinical features: develops over a short period of time (usually hours to a few days), represents a change from baseline attention and awareness, and tends to fluctuate in severity during the course of a day<br>C. Plus at least two of the following:<br>1. perceptual disturbance (misinterpretations, illusions, or hallucinations)<br>2. incoherent speech<br>3. insomnia or daytime drowsiness<br>4. increased or decreased psychomotor activity (Hyperactive/ Hypoactive)<br><br>Exclusions:<br>- Primary mental/neurologic disorder not related to SLE.<br>- Metabolic disturbances (glucose > 25; serum sodium <125 or >155; serum calcium >3)<br><br>NB: Preexisting cognitive deficits are not an exclusion. If acute confusional state is superimposed on preexisting cognitive deficits, diagnose both. |
| visual_disturbance | Diagnostic criteria: |

| | |
|---|---|
| | Either (1) or (2)<br>(1) Retinopathy or choroidopathy with one criteria below, usually bilateral:<br>A. cytoid bodies (cotton-wool spots or retinal soft exudates)<br>B. retinal hemorrhages<br>C. serous exudate in the choroid<br>D. serous hemorrhages in the choroid<br><br>(2) Optic neuritis<br>Clinical criteria (A), (B), or (C) if seen acutely; or at least one Paraclinical criteria with a medical history suggestive of optic neuritis<br>1. Clinical criteria:<br>A. Monocular, subacute loss of vision associated with orbital pain worsening oneye movements, reduced contrast and colour vision, and relative afferent pupillary deficit (RAPD)<br>B. Painless with all other features of (A).<br>C. Binocular loss of vision with all features of (A) or (B).<br>2. Paraclinical criteria:<br>A. OCT: Corresponding optic disc swelling acutely or an inter-eye difference in the mGCIPL of >4% or >4 µm or in the pRNFL of >5% or >5 µm within 3 months after onset.<br>B. MRI: Contrast enhancement of the symptomatic optic nerve and sheaths acutely or an intrinsic signal (looking brighter) increase within 3 months.<br><br>Exclusions:<br>- hypertension (BP > 180/120)<br>- Infectious, post-infectious or post-vaccination<br>- drug caused<br>- drusen (for cytoid bodies)<br>- Autoimmune ON: multiple sclerosis (MS), Aquaporin4 IgG antibodies-associated neuromyelitis optica spectrum disorder (NMOSD) or Anti-myelin oligodendrocytes glycoprotein antibody-associated disease (MOGAD) |
| cranial_nerve_disorder | Diagnostic criteria:<br>New onset of focal sensory or motor neuropathy attributable to cranial nerve dysfunction:<br>I. Olfactory nerve: Loss of sense of smell, distortion of smell, and loss of olfactory discrimination<br>II. Optic nerve: [Intentionally omitted. Not evaluated under cranial nerve disorder criteria.]<br>III. Oculomotor nerve: Ptosis of the upper eyelid and inability to rotate eye upward, downward, or inward (complete lesion), and/or dilated nonreactive pupil and paralysis of accommodation (interruption of parasympathetic fibers only)<br>IV. Trochlear nerve: Extortion and weakness of downward movement of affected eye<br>V. Abducens nerve: Weakness of eye abduction<br>VI. Trigeminal nerve: Paroxysm of pain in lips, gums, cheek, or chin initiated by stimuli in trigger zone (trigeminal neuralgia) and sensory loss of the face or weakness of jaw muscles<br>VII. Facial nerve: Unilateral or bilateral paralysis or facial expression muscles, impairment of taste, and hyperacusis (painful sensitivity to sounds)<br>VIII. Vestibulo-cochlear nerve: Deafness, tinnitus (cochlear), dizziness and/or vertigo (vestibular)<br>IX. Glossopharyngeal nerve: Swallowing difficulty, deviation of soft palate to normal side, anesthesia of posterior pharynx and/or |

| | |
|---|---|
| | glossopharyngeal neuralgia (unilateral stabbing pain in root of tongue and throat, triggered by coughing, sneezing, swallowing, and pressure on ear tragus)<br>X. Vagus nerve: Soft palate droop, loss of the gag reflex, hoarseness, nasal voice, and/or loss of sensation at external auditory meatus.<br>XI. Accessory nerve: Weakness and atrophy of sternocleidomastoid muscle and upper part of trapezius muscle.<br>XII. Hypoglossal nerve: Paralysis of one side of tongue with deviation to the affected side<br><br>Exclusions:<br>- Skull fracture<br>- Tumor: meningioma, carcinomatous meningitis, aneurysm<br>- Infection: herpes zoster, neuroborreliosis, syphilis, mucormycosis<br>- Miller Fisher syndrome |
| lupus_headache | Diagnostic criteria:<br>Either type of headache below, and should be severe (disabling headache), persistent (lasts ≥3 days) also nonresponsive to narcotic analgesia (e.g. heroin, fentanyl, byprenorpine, oxycodone methadone, and morphine).<br>A. Migraine<br><br>Migraine without aura: Idiopathic, recurrent headache manifested by attacks lasting 4-72 hours. Typical characteristics are unilateral location, pulsating quality, moderate to severe intensity, aggravation by routine physical activity, and associated with nausea, vomiting, photo- and phonophobia. At least 5 attacks fulfilling the above criteria.<br>Migraine with aura: Idiopathic, recurrent disorder manifested by attacks of neurologic symptoms localizable to cerebral cortex or brain stem, usually gradually developing over 5-20 minutes and lasting less than 60 minutes. Headache, nausea, and/or photophobia usually follow neurologic aura symptoms directly or after an interval of less than 1 hour. Headache usually lasts 4-72 hours, but may be completely absent.<br>B. Tension headache (episodic tension type headache)<br><br>Recurrent episodes of headaches lasting minutes to days. Pain typically pressing/tightening in quality, of mild to moderate intensity, bilateral in location, and does not worsen with routine physical activity. Nausea is rare, but photophobia and phonophobia may be present. At least 10 previous headaches fulfilling these criteria.<br>C. Cluster headache<br><br>Attacks of severe, strictly unilateral pain, orbital, supraorbital, and/or temporal, usually lasting 15-180 minutes and occurring from at least once every other day up to 8 times per day. Associated with one or more of the following: conjunctival injection, lacrimation, nasal congestion, rhinorrhea, forehead and facial sweating, myosis, ptosis, eyelid edema. Attacks occur in series for weeks or months ("cluster" periods) separated by remissions of usually months or years.<br>D. Headache from intracranial hypertension (Pseudotumor cerebri, benign intracranial hypertension)<br><br>All of the following: |

| | |
|---|---|
| | Increased intracranial pressure (200 mm HiO) measured by lumbar puncture<br>Normal neurologic findings except for papilledema and possible nerve VI palsy<br>No mass lesion and no ventricular enlargement on neuroimaging<br>Normal or low protein and normal white cell count in CSF<br>No evidence of venous sinus thrombosis<br>E. Intractable headache, nonspecific<br><br><br>Exclusions:<br>- Aseptic meningitis (including drug-induced)<br>- Drug-induced pseudotumor cerebri (oral contraceptives, sulfonamides, trimethoprim, etc.)<br>- CNS infection (meningitis/encephalitis)<br>- Tumors and other structural lesions<br>- Low intracranial pressure<br>- Trauma<br>- Metabolic headache that remits with elimination of cause (carbon monoxide exposure)<br>- Withdrawal (caffeine, etc.)<br>- Seizure/postictal state<br>- Sepsis<br>- Intracranial hemorrhage or vascular occlusion |
| CVA | Diagnostic criteria:<br>One of the following and supporting radio imaging study, exclused arteriosclerosis:<br>A. Stroke syndrome: acute focal neurologic deficit persisting more than 24 hours (or lasting less than 24 hours with CT or MRI abnormality consistent with physical findings/symptoms<br>B. Transient ischemic attack: acute, focal neurologic deficit with clinical resolution within 24 hours (without corresponding lesion on CT or MRI)<br>C. Subarachnoid and intracranial hemorrhage: bleeding documented by CSF findings or MRI/CT<br>D. Sinus thrombosis: Acute, focal neurologic deficit in the presence of increased intracranial pressure<br>NB: The finding of unidentified bright objects on MRI without clinical manifestations is not classified at the present time.<br><br><br>Exclusions:<br>- Aseptic meningitis (including drug-induced)<br>- Drug-induced pseudotumor cerebri (oral contraceptives, sulfonamides, trimethoprim, etc.)<br>- CNS infection<br>- Tumors and other structural lesions<br>- Low intracranial pressure<br>- Trauma<br>- Metabolic headache that remits with elimination of cause (carbon monoxide exposure)<br>- Withdrawal (caffeine, etc.)<br>- Seizure/postictal state<br>- Sepsis<br>- Intracranial hemorrhage or vascular occlusion |
| vasculitis | Diagnostic criteria:<br>Either one:<br>A. clinician observed cutaneous vasculitis with at least one of the following description of lesions: Ulceration, gangrene, lender finger nodules, periungual infarction (or necrosis), splinter hemorrhages |

| | |
|---|---|
| | B. biopsy or angiogram proof of cutaneous vasculitis<br><br>Exclusions:<br>-mimics of vasculitis (Atheroembolic disease, Atheromatous vascular disease, Anti-phospholipid syndrome, Multiple myeloma, Infective endocarditis, Para-neoplastic syndromes, Genetic vascular disorders (e.g. Marfan's syndrome), Autoinflammatory syndromes, Hypersensitivity reactions, Cocaine and amphetamine abuse)<br>-Infections (Tuberculosis, Hepatitis B, Hepatitis C, HIV)<br>-Malignancy (Lymphoma, Solid organ malignancy)<br>-Drugs (Penicillamine, Propylthiouracil, Hydralazine, Minocycline, Cocaine)<br>-Environmental exposure (including Dusts and Silica) |
| arthritis | Diagnostic criteria:<br>Either one:<br>A. two or more joints (synovitis) with pain and signs of inflammation (e.g., tenderness, swelling, or effusion)<br>B. two or more joints tenderness and 30 minutes or more of morning stiffness<br><br>Exclusions:<br>- infection (a septic joint or as osteomyelitis): tuberculosis, Bacterial, viral<br>- stress fractures<br>- Crystal arthritis: Gout, calcium pyrophosphate deposition disease<br>- Noninflammatory arthritis, such as osteoarthritis and avascular necrosis (AVN) (especially for hip, knee or shoulder)<br>- Rheumatoid arthritis |
| myositis | Diagnostic criteria:<br>All of the following:<br>A. proximal muscle aching(myalgias)/weakness,<br>B. associated with elevated creatine phosphokinase/aldolase or electromyogram (EMG) changes or a biopsy showing myositis<br><br>Exclusions:<br>- CK elevation was unexplained, isolated, felt to be related to rhabdomyolysis, exertion, infection, or the toxic effect of medication (i.e., statin,hydroxychloroquine-induced myotoxicity)<br>- cancer-associated myositis<br>- metabolic muscle disease |
| urinary_casts | Heme-granular or red blood cell casts. |
| hematuria | Diagnostic criteria:<br>All of the following:<br>A. >5 red blood cells/high-power field. Acceptable equivalents include:<br>- Urine RBC >= 2+ (e.g., 2+, ++, 2+ve, ++ve, +++ve, etc.)<br>- Urine RBC >30-58/uL<br>Important: "urine RBC +" should not be fulfilled.<br>Important: Values such as "high/moderate" or similar terms should be considered as uncertain.<br>Note: Microscopic haematuria is most commonly defined as >3 red blood cells per high-power field on urinary microscopy, but this study uses a threshold of >5. Therefore, it should be considered uncertain.<br>Exclusions: stones, infection or other cause (menstruation). |

| | |
|---|---|
| | B. abnormal urine protein:<br>- UP dipstick positive, or<br>- Urine protein/creatinine > 0.15mg/mg (equivalent to 15mg/mmol or 0.15g/g), or<br>- 24-hour urine protein > 0.15gram/24 hours).<br>In cases where multiple urine protein assessments are available and the results conflict, please prioritize according to the following hierarchy:<br>24-hour urine protein > urine protein/creatinine (UPC) > dipstick |
| proteinuria | Either one:<br>A. 24-hour urine : >0.50 gram/24 hours.<br>B. Urine protein/creatinine > 0.50 mg/mg. (This is equivalent to 0.50 g/g or 50 mg/mmol. Do not convert further - these units are already standardized and interchangeable.)<br><br>NB: UP dipstick results (e.g., "+", "+++", "+ve") are generally uncertain. Numeric values must meet criteria above.<br>Exception: A dipstick reading of 3+ or greater may be scored as fulfilled when correlated with a physician's clinical impression of a lupus flare (i.e., one necessitating escalation of therapy for lupus nephritis).<br><br>If the two results are conflicting at the same time, the 24-hour protein result shall prevail.<br>If UPC unit is unspecified, presume mg/mmol scale for values >20, mg/mg for values <5, and flag uncertain for values in between. |
| pyuria | >5 white blood cells/high-power field and abnormal urine protein (dipstick positive (UP+ or above) or Urine protein/creatinine > 0.15mg/mg (15mg/mmol or 0.15g/g) or 24-hour urine protein >0.15gram/24 hours).<br>In cases where multiple urine protein assessments are available and the results conflict, please prioritize according to the following hierarchy:<br>24-hour urine protein > urine protein/creatinine (UPC) > dipstick<br>Exclude infection. Possible urine infection types include E.coli, Enterococcus, Klebsiella, Pseudomonas, AFB/tuberculosis.<br>NB: in our study, urine WBC 2+, urine WBC ++, etc.,, or urine WBC >= 10-50/uL can be considered as >5 white blood cells/high-power field.<br>Important: "urine WBC +" should not be counted.<br>Note: if the doctor decides to treat patient with antibiotics or conduct further tests to identify the specific pathogen causing the symptoms, the presence of pyuria is likely due to an infection. |
| rash | Diagnostic criteria:<br>Either one form of lupus rash from these three class of lupus erythematosus:<br>A. Acute cutaneous lupus erythematosus (ACLE), either one:<br>1. lupus malar rash;<br>2. bullous lupus;<br>3. toxic epidermal necrolysis variant of SLE;<br>4. maculopapular lupus rash;<br>5. photosensitive lupus rash in the absence of dermatomyositis;<br>B. subacute cutaneous lupus erythematosus (SCLE), either one:<br>1. Annular form<br>2. Papulosquamous form<br>C. Chronic cutaneous lupus erythematosus (CCLE), either one:<br>1. classical discoid lupus erythematosus (DLE)<br>2. localized DLE (above the neck)<br>3. generalized DLE (above and below the neck)<br>4. hypertrophic (verrucous) lupus<br>5. lupus panniculitis (profundus) |

| | |
|---|---|
| | 6. mucosal lupus<br>7. lupus erythematosus tumidus<br>8. chillblains lupus (CHLE, pernio)<br>9. lichen planus overlap<br><br>Note: A diagnosis of lupus rash cannot be made solely based on describing the visual characteristics and location of a rash.<br>Here is the vocabulary to help distinguish between lupus activity versus damage:<br>Activity (erythema):<br>pink, faint erythema, pinkish, faint erythema,<br>faintly erythematous, mild erythema, mildly erythematous, mild redness, mildly red,<br>erythematous, erythema, moderate erythema, moderately erythematous. moderate erythematous,<br>red, redness, reddish, dark red, deep red, deeply red, deep reddish, deeply reddish, purple, purplish, violaceous<br><br>Activity (scale/hypertrophy):<br>Scale, scaling, scaly, follicular plugging, follicular prominence, carpet tacking, carpet tack sign, perifollicular scale<br>Hypertrophy, hypertrophic, hyperkeratosis, hyperkeratotic<br><br>Damage (Dyspigmentation):<br>Hypopigmentation, hypopigmented, hyperpigmentation, hyperpigmented,<br>dyspigmentation, dyspigmented, pigmentary changes, brown, depigment, white<br><br>Damage (scarring/atrophy/panniculitis):<br>Scarring, scar, Atrophic scarring, atrophic, panniculitis, lipoatrophy, atrophic scar<br><br>- Exclusions for ACLE:<br>• Localized form: rosacea, seborrheic eczema, perioral dermatitis, tinea faciei, erysipelas<br>• Generalized form: dermatomyositis, viral and drug-induced rash, erythema multiforme, TEN<br>• Bullous lupus erythematosus (BLE): Epidermolysis bullosa acquisita, dermatitis herpetiformis (Duhring's disease), bullous pemphigoid, linear IgA-dermatosis, drug-induced bullous disorder, porphyria cutanea tarda<br><br>- Exclusions for SCLE:<br>• Psoriasis vulgaris, tinea corporis, mycosis fungoides, erythema annulare centrifugum, dermatomyositis, pityriasis rubra pilaris, nummular eczema, drug-induced rash, seborrheic eczema, erythema multiforme /TEN, erythema gyratum repens<br><br>- Exclusions for Discoid lupus erythematosus (DLE).:<br>• Actinic keratosis, tinea faciei, sarcoidosis, lupus vulgaris |

| | |
|---|---|
| | - Exclusions for Lupus erythematodes profundus (LEP):<br>• Various forms of panniculitis, malignant lymphoma (especially subcutaneous panniculitic T-cell lymphoma), subcutaneous sarcoidosis, panarteritis nodosa, morphea profunda, subcutaneous granuloma annulare<br><br>- Exclusions for Chilblain lupus erythematosus (CHLE):<br>• Pernio (chilblains), lupus pernio (chronic form of skin sarcoidosis of the acral regions), acral vasculitis/vasculopathy<br><br>- Exclusions for Lupus erythematosus tumidus (LET):<br>• Lymphocytic infiltration Jessner-Kanof or erythema arciforme et palpabile (see text), polymorphic light eruption, pseudolymphoma, B-cell lymphoma, plaque-like cutaneous mucinosis, solar urticaria |
| alopecia | Diagnostic criteria:<br>Hair loss or alopecia<br><br>Exclusions:<br>- alopecia areata<br>- iron deficiency<br>- androgenic alopecia<br>- scarring alopecia<br>- drug-induced alopecia |
| mucosal_ulcers | Diagnostic criteria:<br><br>Oral or nasal (nasopharyngeal) ulceration observed by a clinician, including palate, buccal, tongue or nasal ulcers<br><br>Exclusions:<br>- Vasculitis<br>- Behcet's<br>- infection (herpes)<br>- inflammatory bowel disease<br>- reactive |
| pleurisy | Diagnostic criteria:<br>Both A and B:<br>A. Pleuritic chest pain (typically sharp, worse with inspiration, improved by shallow breathing)<br>B. pleural rub OR pleural thickening OR imaging confirmed pleuritic effusion (such as ultrasound, x-ray, CT scan, MRI)<br>Exclusions:<br>- infection<br>- uremia (BUN > 40 mg/dL) |
| pericarditis | Diagnostic criteria:<br>Both A and B:<br>A. pericardial pain (typically sharp, worse with inspiration, improved by leaning forward)<br>B. pericardial rub OR EKG with new widespread ST-elevation or PR depression OR new/worsen imaging confirmed pericardial effusion (such as ultrasound, x-ray, CT scan, MRI)<br><br>Exclusions:<br>- infection (coxsackievirus, mycoplasma, tuberculosis)<br>- uremia (BUN > 40 mg/dL) |

<table>
<tr><td></td><td>- Dressler's pericarditis (post-myocardial infarction syndrome)</td></tr>
<tr><td>low_complement</td><td>Decrease in CH50, C3, or C4:<br>- Use lab flags first:<br>- "low”, “reduced”, or similar terms => fulfilled<br>- “nl” (normal), "high" => unfulfilled<br>- trend-based terms like “lower” => uncertain<br>- If a flag is present, it overrides the numeric value even if the value doesn't meet the threshold.<br>- If no flags or reference ranges are provided, use the hospital thresholds below:<br>Reference for complement in this study (no test for CH50 in our study):<br>Time period | item | Reference (mg/dL)<br>24/12/2018 till now | C3 | <90<br>before 24/12/2018 | C3 | <76<br>24/12/2018 till now | C4 | <10<br>before 24/12/2018 | C4 | <9<br><br>Presume mg/dL scale for values >10, g/L for values <2, and flag uncertain for values in between.</td></tr>
<tr><td>increased_DNA_binding</td><td>Increased DNA binding above laboratory reference range, except ELISA: twice above laboratory reference range.<br><br>NB: If the methodology for increased DNA binding is not mentioned, we assume it is ELISA.<br>Qualitative values (such as "high", "+", "rising", or similar terms) should be considered uncertain for ELISA, but should be considered fulfilled for other methods.<br><br>Laboratory reference range for anti-dsDNA:<br>Time period | Reference<br>1/2/2021 till now | >10 IU/ml<br>21/1/2013 to 1/2/2021 | >25 IU/ml<br>before 21/1/2013 | >67.5 IU/ml<br>Important:<br>- If the clinical note states a reference range (e.g. immunofluorescence), often in brackets, use its provided reference.<br>- “nl” means normal range</td></tr>
<tr><td>fever</td><td>Diagnostic criteria:<br>>38°C (Exclude infectious cause)<br><br>Exclusions:<br>- Infection (usually have high WBC, high CRP or definite localizing infective foci)<br>- malignancy (especially lymphoma)</td></tr>
<tr><td>thrombocytopenia</td><td>Thrombocytopenia (platelets <100,000/mm3 or <100 x 10^9/L), in the absence of other known causes such as drugs, portal hypertension, and thrombotic thrombocytopenic purpura.<br>Important: Qualitative values such as "low", "mild", or similar terms should be considered as uncertain. The exact numerical value must be stated and meet the criteria above.<br>In clinical practice, the default unit is 1000/mm3 or 10^9/L</td></tr>
<tr><td>leukopenia</td><td>Leucocyte count, < 3,000/mm3 or < 3 x 10^9/L white blood cells.<br>Exclude Felty's, drug causes and portal hypertension.</td></tr>
</table>

| | |
|---|---|
| | Important: Qualitative values such as "low" or similar terms should be considered as uncertain. The exact numerical value must be stated and meet the criteria above.<br>Also, only consider leucocyte counts labeled as WCC or WBC. Do not consider ANC.<br>In clinical practice, the default unit is 1000/mm3 or 10^9/L |

Note: In the MIMIC-IV data, different reference ranges are used for some lab tests, and SLEDAI-2K is assessed at the time of admission instead of the date of the clinical note. Thus, the following descriptors have modified diagnostic criteria (or additional rules) for MIMIC.
