## Supplementary material for "Automated Disease Activity Assessment in Systemic Lupus Erythematosus Using Privacy-Preserving Large Language Models": Annotation Guidelines

### Annotation Guidelines for SLEDAI-2K

21. Increased DNA binding ... 80
21.1. Clinical definition of concepts ... 80
21.2. Common ways to describe this concept ... 81
21.3. SLEDAI-2K definition in this study ... 81
21.4. Examples present of SLEDAI-2K score ... 81
21.5. Examples absent of SLEDAI-2K score ... 81
22. Fever ... 82
22.1. Clinical definition of concepts ... 82
22.2. Common ways to describe this concept ... 82
22.3. SLEDAI-2K definition in this study ... 82
22.4. Examples present of SLEDAI-2K score ... 82
22.5. Examples absent of SLEDAI-2K score ... 83
23. Thrombocytopenia ... 84
23.1. Clinical definition of concepts ... 84
23.2. Common ways to describe this concept ... 84
23.3. SLEDAI-2K definition in this study ... 85
23.4. Examples present of SLEDAI-2K score ... 85
23.5. Examples absent of SLEDAI-2K score ... 85
24. Leukopenia ... 85
24.1. Clinical definition of concepts ... 85
24.2. SLEDAI-2K definition in this study ... 86
24.3. Examples present of SLEDAI-2K score ... 86
24.4. Examples absent of SLEDAI-2K score ... 86
References: ... 93

### 1. Chapter 1-Background

Systemic lupus erythematosus (SLE) is an autoimmune disorder characterized by multisystem inflammation and diverse clinical manifestations. It predominantly affects women, accounting for more than 90% of cases. The disease remains incurable, with approximately 70% of patients experiencing a relapsing-remitting course throughout their lives(1), making continuous disease monitoring crucial.

The Systemic Lupus Erythematosus Disease Activity Index (SLEDAI) (2) is one of the most popular tools for measuring activity and predicting long-term damage and mortality. The instrument itself is comprehensive, assessing activity across nearly all organ systems, including mucocutaneous, neuropsychiatric, musculoskeletal, cardiorespiratory, ophthalmic, renal, and hematologic, using 24 different disease descriptors (Table 1). Therefore, accurate scoring requires sophisticated clinical reasoning that synthesizes symptoms, signs, and paraclinical test results. However, its clinical utility is limited by the labor-intensive scoring process and a shortage of rheumatologists, often resulting in incomplete documentation in practice.

The advent of generative AI and large language models (LLMs) offers a potential solution by automating such complex clinical evaluations from electronic clinical notes. More and more open source/weighted models like Llama-3, Mistral-7B, Deepseek-2.5, and Qwen3 show strong inference capabilities.(3) This study aimed to test the potential of LLMs to automate the assessment of the SLEDAI-2K.

To accomplish our study objective, we need to develop a high-quality, clinician-annotated ground-truth dataset for SLEDAI-2K. In this dataset, the SLEDAI-2K score is annotated for each clinical note within the BRAT annotation system. Tailored for SLEDAI-2K, this guideline serves to provide annotators with comprehensive, essential details for the annotation task. Furthermore, we provide practical refinements to the SLEDAI-2K definitions, supplemented with abundant annotation examples. This ensures our annotation process is professional, reproducible, and transparent. The guideline comprises two main parts: 1) the BRAT annotation structure tailored for SLEDAI-2K, and 2) the detailed criteria for each SLEDAI-2K descriptor, accompanied by annotation examples.

*Table 1: SLEDAI-2K data collection form with 24 descriptors.(4) (Enter weight in SLEDAI Score column if the descriptor is present at the time of the visit or in the preceding 30 days.(5))*

| Weight | Score | Descriptor | Definition |
|---|---|---|---|
| **8** | | Seizure | Recent onset, excluding metabolic, infectious, or drug causes |
| **8** | | Psychosis | Altered ability to function in normal activity due to severe disturbance in the perception of reality. Include hallucinations, incoherence, marked loose associations, impoverished thought content, marked illogical thinking, bizarre, disorganized, or catatonic behavior. Exclude uremia and drug causes |
| **8** | | Organic brain syndrome | Altered mental function with impaired orientation, memory, or other intellectual function, with rapid onset, and fluctuating clinical features. Inability to sustain attention to the environment, plus at least two of the following: perceptual disturbance, incoherent speech, insomnia or daytime drowsiness, or increased or decreased psychomotor activity. Exclude metabolic, infectious, or drug causes |
| **8** | | Visual disturbance | Retinal changes of SLE. Include cytoid bodies, retinal hemorrhages, serous exudate or hemorrhages in the choroid, or optic neuritis. Exclude hypertension, infection, or drug causes |
| **8** | | Cranial nerve disorder | New onset of sensory or motor neuropathy involving cranial nerves |

| | | | |
|---|---|---|---|
| **8** | | Lupus headache | Severe, persistent headache; may be migrainous, but must be nonresponsive to narcotic analgesia |
| **8** | | CVA | New onset of cerebrovascular accident(s). Exclude arteriosclerosis. |
| **8** | | Vasculitis | Ulceration, gangrene, lender finger nodules, periungual infarction, splinter hemorrhages, or biopsy or angiogram proof of vasculitis |
| **4** | | Arthritis | ≥2 joints with pain and signs of inflammation (e.g., tenderness, swelling, or effusion) |
| **4** | | Myositis | Proximal muscle aching/weakness, associated with elevated creatine phosphokinase/aldolase or electromyogram changes or a biopsy showing myositis |
| **4** | | Urinary casts | Heme-granular or red blood cell casts. |
| **4** | | Hematuria | >5 red blood cells/high-power field. Exclude stone, infection or other cause. |
| **4** | | Proteinuria | >0.5 gram/24 hours. |
| **4** | | Pyuria | >5 white blood cells/high-power field. Exclude infection. |
| **2** | | Rash | Inflammatory-type rash |
| **2** | | Alopecia | Abnormal, patchy, or diffuse hair loss |
| **2** | | Mucosal ulcers | Oral or nasal ulceration |
| **2** | | Pleurisy | Pleuritic chest pain with pleural rub or effusion, or pleural thickening. |
| **2** | | Pericarditis | Pericardial pain with at least one of the following: rub, effusion, or electrocardiogram or echocardiogram confirmation. |
| **2** | | Low complement | Decrease in CH50, C3, or C4 below the lower limit of normal for testing laboratory |
| **2** | | Increased DNA binding | Increased DNA binding by Farr assay above normal range for testing laboratory. |
| **1** | | Fever | >38°C. Exclude infectious cause |
| **1** | | Thrombocytopenia | <100,000 platelets/$\times 10^9$/L. Exclude drug causes |
| **1** | | Leukopenia | <3,000 white blood cells/$\times 10^9$/L. Exclude drug causes |

**Total score:_____**

### 2. Chapter 2-Annotation with BRAT

#### 1. Introduction to the BRAT system

BRAT is a web-based tool for annotating text-based structured documents (https://brat.nlplab.org/standoff.html) (6). BRAT is specifically designed for ***structured annotation*** rather than free-form notetaking. The fixed form can be automatically processed and interpreted by a computer. BRAT supports three annotation types: ***text span (entities and events)***, ***relations***, and ***attributes***. This framework can be specifically tailored to model the complex information found in clinical notes.

Our objective was to create an annotation schema tailored for SLE clinical notes. The schema is designed to identify 24 descriptors or ***entities*** (e.g., diagnoses for certain descriptors) and link them, via specific ***relations***, to associated clinical ***events*** such as symptoms, signs, paraclinical findings, and treatments.

As shown on the right, the word "arthritis" is annotated as an ***entity*** representing a relevant SLEDAI-2K descriptor. The terms "gouty" and "bilat big toe" were annotated as symptoms/signs events, providing contextual detail for this diagnosis. The ***relation*** annotations, depicted by arrows, show that "gouty" and "bilat big toe" are details pertaining to the diagnosis of "arthritis," and not linked to other diagnostic entities. In this example, we see ***text span annotations*** (entity and event) and ***relation annotations*** (the links).

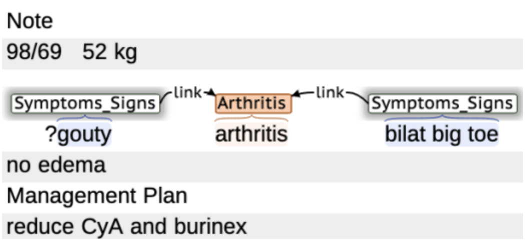

In addition to a clinical diagnosis, SLEDAI-2 K incorporates descriptors based on laboratory tests. Within our annotation schema, the ***entity*** represents the name of the laboratory examination, while the related events capture the test value and its unit. For example, the term "24 hour UP" is annotated as an ***entity*** [proteinuria]. The value "0.67" and the unit "g/d" are annotated as ***events*** linked to this entity. The ***relation*** annotations explicitly associate the value with the specific laboratory test with the entity [links_to], and the unit with a specific value [is_unit_of].

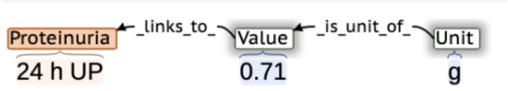

Within the BRAT annotation framework, we also assigned ***attributes*** to each ***entity***. As seen with arthritis ***entity*** shown on the right side, its associated symptoms/signs events indicated a gout attack. This presentation did not fulfill the diagnostic criteria for lupus arthritis according to SLEDAI-2K. Consequently, we assigned a [criteria_unfulfilled] ***attribute*** to the ***entity*** to denote that it did not meet the SLEDAI-2K definition.

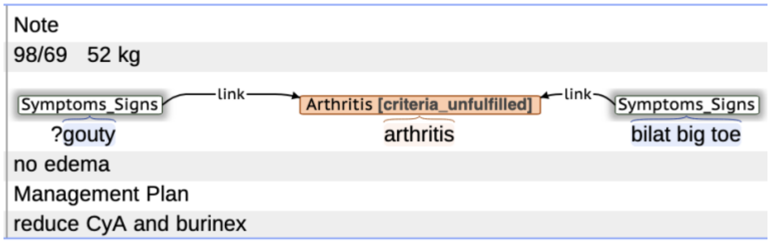

The result of 24-hour urine protein on the right fulfilled the criteria of SLEDAI-2K. Therefore, we gave an ***attribute*** of [criteria_fulfilled] to indicate that it is an active proteinuria and should be scored.

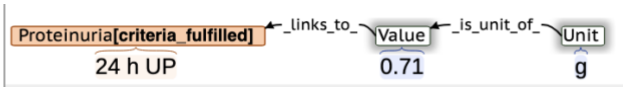

#### 2. Annotation structure for scoring SLEDAI-2K in the BRAT system

We differentiated the annotation structures for clinical and laboratory descriptors within the SLEDAI-2K framework to accurately reflect their distinct natures. 16 ***Clinical descriptors*** demand complex clinical judgment, integrating symptoms, signs, exclusionary diagnoses, paraclinical data, and treatment outcomes; thus, their accurate annotation relies on comprehensive clinical notes. 8 ***Laboratory descriptors*** are determined directly by objective laboratory results, such as proteinuria and low complement. This difference in interpretation rationale led us to develop specialized annotation schemas for each category to better capture their textual and relational elements (**Table 2** below).

Another important factor in SLEDAI-2K assessment is time. Whereas the original SLEDAI-2K evaluates the disease activity in the preceding 10 days, we selected a 30-day window to enhance clinical relevance. This longer timeframe was chosen for its increased informativeness in clinical practice, where a lag often exists between laboratory tests and physician consultation. We defined time as a specialized ***entity*** to capture this timeline accurately.

*Figure 1. The BRAT annotation workflow for evaluating SLEDAI-2K*

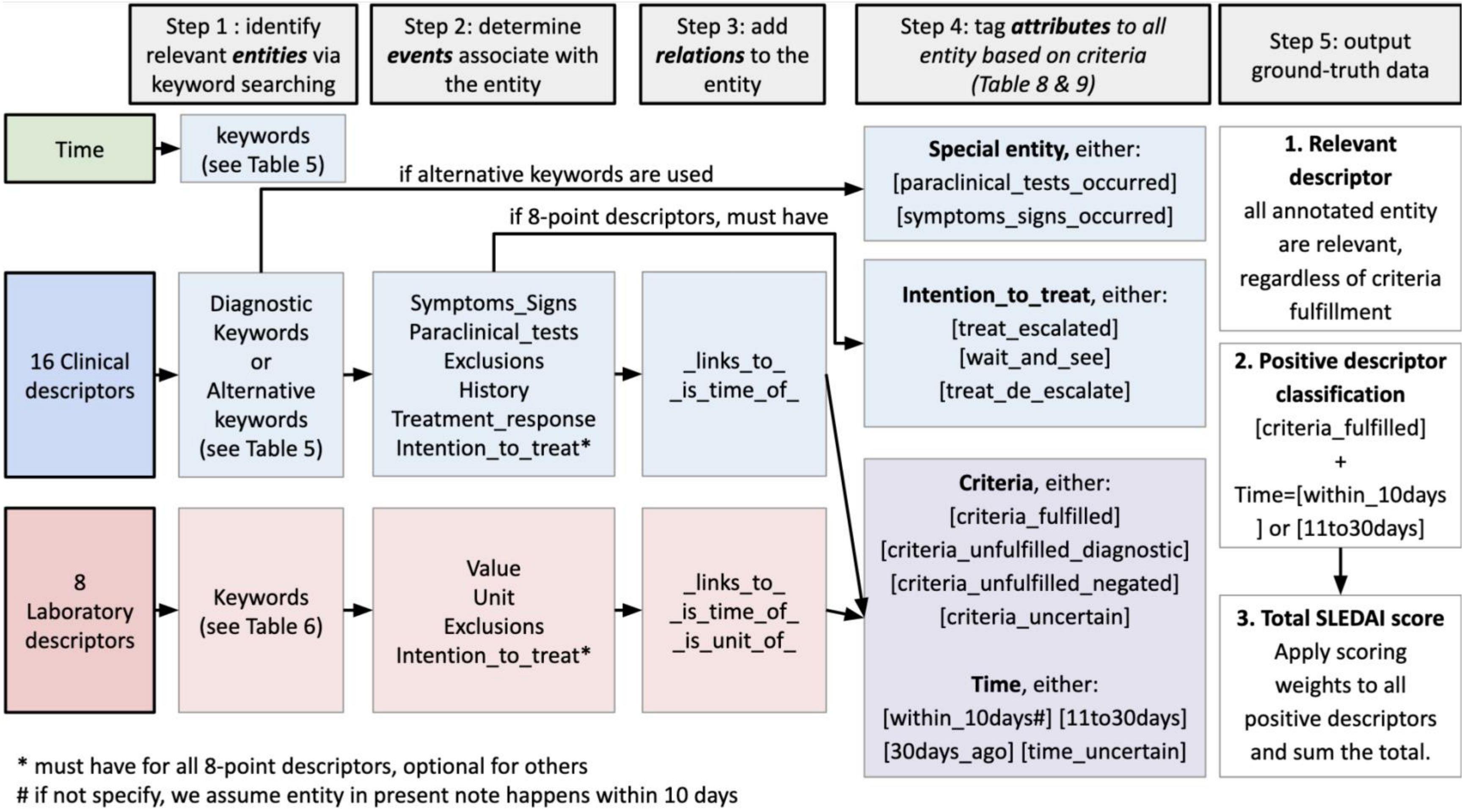

*Table 2. The BRAT annotation structure for evaluating SLEDAI-2K: text span and attributes*

| | **Entities** (time+24 descriptors) | **Events** | **Attributes** of entities (SLEDAI criteria) | **Attributes** of entities (time) | **Attributes** of entities (special entity) | **Attributes** of entities **(Intention_to_treat)** |
|---|---|---|---|---|---|---|
| Time | Time | / | / | **[within_10days]**<br>**[11to30days]**<br>**[30days_ago]**<br>**[time_uncertain]** | / | |
| **Clinical descriptors N=16** | Seizure<br>Psychosis<br>Organic brain syndrome<br>Visual disturbance<br>Cranial nerve disorder<br>Lupus headache<br>CVA<br>Vasculitis<br>Arthritis<br>Myositis<br>Rash<br>Alopecia<br>Mucosal ulcers<br>Pleurisy<br>Pericarditis<br>Fever | symptoms_signs<br>paraclinical_tests<br>exclusions<br>history<br>treatment_response<br>Intention_to_treat* | **[criteria_fulfilled]** fulfill the criteria of SLEDAI-2K (without considering the time issue)<br>**[criteria_unfulfilled_diagnostic]** unfulfill the diagnostic criteria of SLEDAI-2K (without considering the time issue)<br>**[criteria_unfulfilled_negated]** denial of the concept<br>**[criteria_uncertain]** | **[within_10days]** happened within 10 days<br>**[11to30days]** happened within a 11-30 days window<br>**[30days_ago]** happened 30 days ago<br>**[time_uncertain]** time is not clear | **[]** is blank when diagnosic keywords were used<br>**[special_entity_symptoms_signs_occurred]** alternative keywords were used<br>**[special_entity_paraclinical_tests_occurred]** alternative keywords were used | **[treat_escalated]** initiation or escalation of steroids, immunosuppressants, biologics.<br>**[wait_and_see]**<br>**[treat_de_escalate]** |
| **Laboratory descriptors N=8** | Urinary casts<br>Hematuria<br>Proteinuria<br>Pyuria<br>Low complement<br>Increased DNA binding<br>Thrombocytopenia | value<br>unit<br>exclusions<br>Intention_to_treat* | **[criteria_fulfilled]**<br>**[criteria_unfulfilled_diagnostic]**<br>**[criteria_unfulfilled_negated]**<br>**[criteria_uncertain]** | **[within_10days]**<br>**[11to30days]**<br>**[30days_ago]**<br>**[time_uncertain]** | / | |

| | |
|---|---|
| | Leukopenia |

*Intention_to_treat (doctors' management plan) is the additional criterion for all 8-point descriptors: Seizure, Psychosis, Organic Brain Syndrome, Visual Disturbance, Cranial Nerve Disorder, Lupus Headache, and Vasculitis. It is optional for other descriptors

Negation rules: Keywords for descriptors with clear negation words such as "no" or "absent of", are considered negation.

*Table 3. The BRAT annotation structure for evaluating SLEDAI-2K-relations*

| Three types of relations | Explanation |
|---|---|
| **_links_to_** | All events will be given at least one link to entity. |
| **_is_time_of_** | Time, if specifically mentioned, should be annotated as time entity and linked to the relevant descriptor entity. |
| **_is_unit_of_** | Specific for laboratory descriptors. All unit will be given a link to the corresponding value. |

### 3. Separating notes for present illness from historical notes

Because the SLEDAI-2K assesses current disease activity, our annotation was restricted to notes documenting the patient's present condition to reduce the annotation burden. A critical preliminary step involved distinguishing current free-text notes from historical documentation. This step is essential for determining the temporal context of each entity. There are some indicators to start the current notes (***Table 4***). Cases in which the beginning of the current illness record could not be confirmed were deemed of poor quality and were excluded.

*Table 4. Indicators for the beginning of the current notes.*

| Indicators | Examples |
|---|---|
| FU<br>FU today<br>Currently<br>now | Note<br>Class V LN 1992 (Bx some chronic changes); on CyA, stopped in 94<br>Pulmonary TB in 1993; treated for 1 year<br>Persistent proteinuria since 1994<br>Small lacunar infarcts in 1997<br>Thyrotoxicosis with RAI 11/99 and post RAI hypothyroidism 9/00<br>Echo: 22/6/2018→EF ~65%; mildly thickened AMVL with mild dooming<br>FU today<br>BP 154/62 P 77 BW 44.8kg<br>TFT not checked this time<br>Anti-dsDNA 9.8->36.5->70.2<br>C3/C4/CRP normal<br>Cr 155 eGFR 26 static<br>K 5.1<br>Clinically no rash/oral ulcer/alopecia<br>Static finger joints pain with OA changes<br>Static proteinuria |
| Starts with vital signs | Clerical work<br>Non-smoker, non-dinker<br>no known drug allergy<br>BP 95/61; P 83 urine RBC negative; albumin negative<br>WCC normal; Hb 13.6; PLT 205;<br>CRP 0.7; Anti-dsDNA 142; C3 49<br>RFT normal; AST↑ 87/ ALT ↑ 72<br>Complained of<br>(1) fever and diarrhoea; on-off low grade<br>(2) insomnia<br>(3) frequent aphthous ulcer<br>No definite contact/ travel history<br>P/E:<br>→ GC well; not spetic looking; T =<br>→ chest clear'; abdomen soft and non-tender<br>Management Plan<br>CXR (wet films), AXR (Erect)<br>Check CBP, L/RFT, LDH, AST<br>Stool x C/ST, C. difficile<br>Recheck CBP, L/RFT, LDH, CPK for trend<br><br>Note: While, doctors typically begin progress notes after recording the vital signs, exceptions exist based on clinical context. |

| | |
|---|---|
| =====<br>****** | Note<br>SLE since 1980, with skin, joint and renal involvement<br>Class IV nephritis 10/05 given steroid+MMF<br>MMF stopped on 11/10/07 → Aza<br>Off Aza since 29/10/2015<br>==<br>seen alone<br>well<br>HBPM 100-110/50-60<br>serology quiescent<br>Cr 85 stable<br>count stable |

#### 4. Identify relevant entities via keyword searching -Step 1

The first step in obtaining a SLEDAI-2K score is to indentify relevant descriptor that occurred within the clinical note, irrespective of whether the diagnostic criteria are fulfilled. To ensure comprehensive capture, we implemented a **keyword searching** strategy for identifying each entity. These keyword lists were designed to encompass medical synonyms and subcategories of disease entities (e.g., 'grand mal' for seizures, 'TIA' for CVA). Synonym variation arises from differences in physician writing styles, the use of abbreviations, and the historical evolution of medical terminology (i.e., the replacement of 'organic brain syndrome' with 'delirium').

For Clinical descriptors, the ideal keywords to annotate as entities are formal diagnostic terms, named as ***Diagnostic keywords***. When a clear diagnosis is absent, clinical features that satisfy the diagnostic criteria, including characteristic symptoms, signs, or paraclinical findings, serve as the ***Alternative keywords***. For Laboratory descriptors, the keywords for entities are the names of the laboratory tests. As summarized in Tables 4 and 5, we compiled a set of keywords to identify each entity. This lexicon was developed through a comprehensive literature review (***see Chapter 3***) and inputs from experts.

*Table 5. Keywords list for identifying entities of Time and Clinical descriptors.*

| Entities | **Diagnostic Keywords** | **Alternative keywords** (when no diagnostic keywords) | Examples for Entities |
|---|---|---|---|
| Time | mm/yyyy<br>FU<br>currently<br>now<br>today<br>? days ago<br>day(s)<br>?/52 (? indicating weeks)<br>?/12 (? indicating month)<br>wk(s)<br>on and off<br>occasionally/occasional<br>sometimes<br>last visit/last | / | treatment started in Oct 06 for relapse of lupus nephritis<br>Time<br>but 24 hr urine x p 3.9 in Mar 07 |

| | | | |
|---|---|---|---|
| Seizure | NPSLE<br>seizure<br>epilepsy<br>grand mal<br>petit mal<br>drop attack | **[Special_entity_ symptoms_signs_occurred]**<br>loss of consciousness (LOC)<br>loss of awareness<br>tonic<br>clonic<br>myoclonic<br>atonic<br>generalized stiffness<br>twitching<br>uprolling eyeballs<br>tongue bite<br>syncope | Well. No ankle oedema.<br>c/o LOC for few minutes<br>Seizure<br>? epileptic attack |
| Psychosis | NPSLE<br>psychosis<br>psychotic disorder | **[Special_entity_ symptoms_signs_occurred]**<br>psychotic<br><br>delusions<br>hallucinations<br>disorganized thinking/speech<br>grossly disorganized or abnormal motor behavior<br>catatonic behavior<br><br>paranoia<br>psychotic beliefs<br>false beliefs<br>bizarre beliefs<br>weird beliefs<br><br>sensory misperceptions<br>perceptual disturbances<br>illusions<br>phantom sensations | Psychosis **[special_entity_symptoms_signs_occurred]**<br>Also c/o some disorganized speech and reduced GC<br><br>this monring, BP fell to 120/80 after medicine<br>has some stress recently, with poor sleep<br>Psychosis **[special_entity_symptoms_signs_occurred]**<br>occ abnormal motor behavior |
| Organic brain syndrome | NPSLE<br>organic brain syndrome<br>organic brain disorder<br>acute confusional state<br>delirium<br>encephalopathy<br>neurocognitive disorders | **[Special_entity_ symptoms_signs_occurred]**<br>disturbance of consciousness<br>impaired/reduced/ shift attention<br>attention deficit<br>impaired/reduced sustain<br>reduced ability to direct/ focus/sustain<br><br>impaired/decreased/reduced awareness<br>impaired orientation<br>disorientation<br><br>impaired cognition<br>cognitive impairment<br>decreased/impaired cognitive<br><br>memory deficit/impairment/loss<br>impaired recall<br>hypomnesia<br>amnesia<br><br>incoherent speech<br>disorder/impaired language ability<br><br>impaired perception<br>misinterpretations<br>illusions<br>hallucinations<br><br>impaired mental function | Organic_brain_syndrome**[special_entity_symptoms_signs_occurred]**<br>cognitive decline<br>(forgetfulness, mood fluctuations).<br><br>Feels dizzy with irregular periods and menorrhagia, also tranisent mild headahce noted, and<br>Organic_brain_syndrome**[special_entity_symptoms_signs_occurred]**<br>reduced ability to focus |

| | | | |
|---|---|---|---|
| | | hyper-aroused/hyperactive<br>hypo-aroused/hypoactive<br><br>coma<br>sleepiness<br>insomnia<br>difficulty falling asleep<br>nighttime agitation<br>lack of responsiveness<br>drowsiness | |
| Visual disturbance | retinal disease(s)<br>retinopathy<br>choroidopathy<br>chorioretinopathy<br>optic neuritis<br>retrobulbar neuritis<br>retinal vasculitis<br>optic neuropathy | [Special_entity_ symptoms_signs_occurred]<br>loss of vision<br>blurring of vision<br>visual impairment<br>orbital pain<br>eye pain<br>relative afferent pupillary deficit (RAPD)<br>choroid hemorrhage<br>retinal hemorrhages<br>cotton-wool spots<br>exudates<br>cytoid bodies | Visual_disturbance<br>History of HCQ retinopathy<br><br>Recently seen private eye doctor, diagnosed to have Migraine.<br>Unilateral associated with<br>Visual_disturbance[special_entity_symptoms_signs_occurred]<br>blurring of vision, like water |
| Cranial nerve disorder | NPSLE<br>Bell's palsy<br>cranial mononeuropathy<br>cranial polyneuropathy<br>cranial neuritis<br><br>**PALSY, paralysis, neuropathy, neuritis, or disorder** with the following nerve:<br>cranial nerve<br>olfactory nerve<br>optic nerve<br>oculomotor nerve<br>trochlear nerve<br>abducens nerve<br>trigeminal nerve<br>facial nerve<br>vestibulo-cochlear nerve<br>glossopharyngeal nerve<br>vagus nerve<br>accessory nerve<br>hypoglossal nerve<br>CN I<br>CN II<br>…<br>CN XII<br>1st NERVE<br>2nd NERVE<br>…<br>12th NERVE) | [Special_entity_ symptoms_signs_occurred]<br>nerve paralysis<br>nerve PALSY<br>nerve neuropathy | PE<br><br>Cranial_nerve_disorder<br>CN VI Palsy (R): Abducens nerve weakness.<br>RAPD (L): Afferent pupillary defect.<br>R Hemiparesis: 3/5 strength UE/LE.<br>R Facial/Hypoglossal Palsy. |
| Lupus headache | lupus headache | [Special_entity_ symptoms_signs_occurred]<br>headache<br>cephalalgias<br>migraine | Occasional mild frontal<br>Lupus_headache[special_entity_symptoms_signs_occurred]<br>headache, relieved by<br>simple analgesics |
| CVA | stroke<br>subarachnoid hemorrhage<br>cerebral venous | [Special_entity_ symptoms_signs_occurred]<br>hemiplegia<br>unilateral weakness | CVA [special_entity_paraclinical_tests_occurred]<br>- CT brain: ld right MCA infarct and right internal capsule lacunar infarct |

| | | | |
|---|---|---|---|
| | thrombosis<br>TIA<br>Transient ischaemic attack<br>Sinus thrombosis | | |
| Vasculitis | cutaneous vasculitis<br>vasculitic rash<br>vasculitic ulcer<br>vasculitis (only when indicating cutaneous) | [Special_entity_ symptoms_signs_occurred]<br>skin ulceration<br>skin ulcer(s)<br>gangrene<br>necorsis<br>lender finger nodules<br>periungual infarction<br>splinter hemorrhages | No complain<br>No joint pain/skin rash/oral ulcer<br>Vasculitis [special_entity_symptoms_signs_occurred]<br>Right leg ulcer healing well,<br>Vasculitis [special_entity_symptoms_signs_occurred]<br>but new ulceration over inner<br>aspect of right ankle joint. |
| Arthritis | arthritis<br>pauciarticular<br>polyarthritis<br>polyarticular | [Special_entity_ symptoms_signs_occurred]<br>arthralgia<br>tenderness<br>swelling<br>effusion<br>morning stiffness<br>joint pain<br>joint inflammation<br>jt pain<br>pain <u>in these location</u>:<br>MCP,DIP,PIP,knee,hip,writst<br>[Special_entity_ paraclinical_tests_occurred ] synovial fluid<br>synovitis<br>synovial inflammation<br>synovial proliferation | Arthritis [special_entity_symptoms_signs_occurred]<br>no alopecia/joint pain/oral ulcer<br>Arthritis<br>no arthritis/facial skin rash |
| Myositis | myositis | [Special_entity_ symptoms_signs_occurred]<br>myalgia<br>muscle aching<br>muscle weakness<br><br>[Special_entity_ paraclinical_tests_occurred ] elevated CK<br>CK+<br>↑CK<br>elevated CPK<br>CPK+<br>↑CPK | Myositis[special_entity_paraclinical_tests_occurred]<br>CK elevated<br>complained of myalgia |

| | | | |
|---|---|---|---|
| Rash | cutaneous flare<br>cutaneous lupus<br>CLE<br>lupus erythematosus (LE)<br>malar rash<br>bullous lupus<br>toxic epidermal necrolysis variant (TEN)<br>generalized maculopapular rash<br>maculopapular lupus rash<br>photosensitive<br>photosensitivity<br>sunlight reaction<br>discoid<br>DLE<br>hypertrophic (verrucous) lupus<br>lupus panniculitis (profundus)<br>mucosal lupus<br>lupus erythematosus tumidus<br>chillblains<br>lichen planus<br>overlap | **[Special_entity_ symptoms_signs_occurred]**<br>rash<br>erythema<br><br>plaques<br>nodules<br>vesicles<br>exanthema<br>enanthema<br><br>maculopapular<br>morbilliform<br>telangiectases<br>red lunula<br>annular form<br>papulosquamous<br>psoriasis-like<br>scale<br>scaling<br>scaly<br>perifollicular scale<br>erythematous-violaceous lesions<br>follicular hyperkeratosis<br>follicular plugging<br>follicular prominence<br>carpet tacking<br>carpet tack sign<br>reddened inflammation<br>hypertrophy<br>hypertrophic<br>hyperkeratosis<br>hyperkeratotic | PE:<br>Anxious, disoriented, poor eye contact. Restless, uncooperative.<br>Rash<br>Pallor +, malar rash.<br><br>PE<br>hyperpigmented maculopapular<br>Rash[special_entity_symptoms_signs_occurred]<br>rash<br>(face/neck/back/extremities; spares nasolabial folds). |
| Alopecia | hair loss<br>hair<br>alopecia<br>aloperic | | oral ulcer + now<br>Alopecia<br>↑ hair loss |
| Mucosal ulcers | ulceration<br>ulcers<br>ulcer<br>stomatitis<br>(note: the position of the ulcers should be follows: oral, mouth, mucosal, nasal, nasopharyngeal, palate, buccal, tongue)<br><br>aphthous ulcer<br>salt blister<br>canker sores | | small area of rash over L. cheek<br>Mucosal_ulcers<br>oral ulcer + now<br>↑ hair loss<br><br>PE:<br>Mucosal_ulcers<br>Discoid rash, ulcer on the hard palate |
| Pleurisy | serositis<br>pleurisy<br>pleuritis | **[Special_entity_ symptoms_signs_occurred]**<br>pleuritic chest pain<br>pleuritic pain<br>pleural rub<br>pleural thickening<br><br>**[Special_entity_ paraclinical_tests_occurred ]**<br>pleural effusion<br>pleural fluid | CXR increase CTR apical<br>Pleurisy[special_entity_paraclinical_tests_occurred]<br>pleural thickening<br>Pending next CT on 14/12/2022<br>Known right pleural effusion and chest pain worsened by deep breathing<br>Pleurisy<br>previously refused tapping; clinically lupus pleurisy; to perform pleural tap to rule out alternative causes |
| Pericarditis | serositis | **[Special_entity_ symptoms_signs_occurred]** | Case 1: |

| | | | |
|---|---|---|---|
| | pericarditis<br>hydropericardium | pericardial pain<br>pericardial chest pain<br>pericardial rub<br>cardiac tamponade<br><br>**[Special_entity_ paraclinical_tests_occurred ]** pericardial effusion<br>pericardial fluid | FU today<br>came alone, walk unaided<br>Pericarditis [special_entity_symptoms_signs_occurred]<br>c/o pericardial pain<br>Case 2:<br>Impression<br>Active SLE flare w/ Steroid-induced delirium (Naranjo score 7)<br>Pericarditis<br>Pericarditis (rub + effusion) |
| Fever | | **[Special_entity_ symptoms_signs_occurred]**<br>PUO<br>PVO<br>fever<br>(any records of tempature more than 38 in below format, ? below should be a number more than 38)<br>Temp ?<br>? ℃<br>? C<br>febrile | Fever [special_entity_symptoms_signs_occurred]<br>98/60; P 116; 51 kg,Temp 38.5 C; |

*Table 6. Keywords list for identifying entities of Laboratory descriptors.*

| Entities | Keywords for ***Entities*** | Examples of Entities |
|---|---|---|
| Urinary casts | heme-granular casts<br>RBC casts | Urinary_casts Urinary_casts<br>24h UP 2.5, WBC 3+, RBC 2+, heme-granular casts, RBC casts, |
| Hematuria | hematuria<br>urine RBC<br>URBC<br>Urine RC<br>Urine blood | Hematuria<br>Urine for RBC +++ Protein +++ |
| Proteinuria | UP<br>urine protein<br>urine P<br>upr<br>P (only with KSP or K/S/P)<br>UPC<br>UPCR<br>UP/C<br>P/Cr<br>PC ratio<br>P:C ratio<br>P/C<br>24 hr urine protein<br>24 hr urine TP<br>24 hr urine pr<br>24 hr urine x p<br>24hUP | Proteinuria<br>urine P +<br>serology inactive<br>no joint / skin rash / occasional oral ulcer<br>Proteinuria<br>24 hr urine 1.58 g |
| Pyuria | pyuria<br>urine WBC<br>UWBC<br>Urine WC | Pyuria<br>MSU: WCC 10-50 |
| Low complement | C3<br>C4<br>C3/4<br>complements<br>complement<br>hypocomplementemia | Low_complement<br>antidsDNA 47 C3 63 |

| | | |
|---|---|---|
| Increased DNA binding | anti-ds DNA<br>anti-dsDNA<br>antiDNA<br>anti DNA<br>dsDNA | 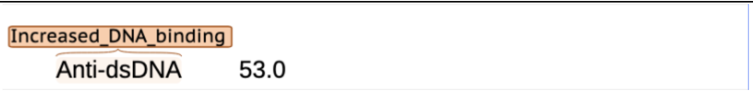 |
| Thrombocytopenia | PLT<br>platelet | 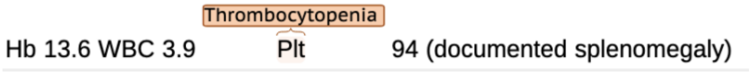 |
| Leukopenia | WCC<br>WBC | 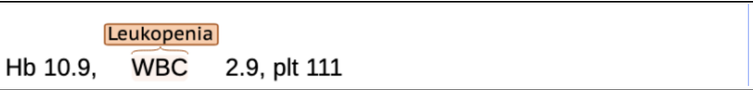 |

#### 5. Determine events associated with each entity-Step 2 & 3

Once entities are identified, we proceed to capture all relevant information, categorized as ***events***. The event structure for ***Clinical descriptors*** (see Table 2) is based on the diagnostic rationale for a lupus descriptor, including symptoms, signs, paraclinical findings, differential diagnoses, history, treatment response, and therapeutic intent. Accurately identifying these events necessitates a nuanced and coherent interpretation of the clinical context. For ***Laboratory descriptors***, the relevant events are the numerical value, the unit of test, and differential diagnoses. All events should be linked to the entity.

#### 6. Annotate the entity with attributes based on criteria - Step 4

After finishing the previous text-span and relation annotation, we assign attributes to each entity by comprehensively analyzing relevant ***events or time***. Given that the original SLEDAI-2K criteria may require further specification in different clinical settings, ***Tables 8 & 9*** in ***Chapter 3*** elaborates detailed criteria applicable to our study.

#### 7. Get human ground-truth for further analysis - Step 5

After completing the previous human annotation part, we obtain the structured annotation (ann.) files from BRAT. As mentioned in the manuscript, the step generates ***three key outputs*** for use as ground-truth: 1) ***Descriptor Relevancy***: all annotated entities, 2) ***Descriptor Classification***: positive descriptors with criteria_fulfilled and within 30-day window, and 3) ***Total SLEDAI Score***: computed by summing the weighted points of all positive descriptors. These three outcomes serve to measure the LLMs performance. Moreover, the ann. files provide the important information for fine-tuning.

#### 8. Practice with demo cases

Figures 2 and 3 below depict step-by-step annotation examples for Clinical and Laboratory descriptors, respectively. Table 7 presents the complete BRAT annotation results for a full clinical note.

*Figure 2. A demo annotation of a Clinical descriptor*

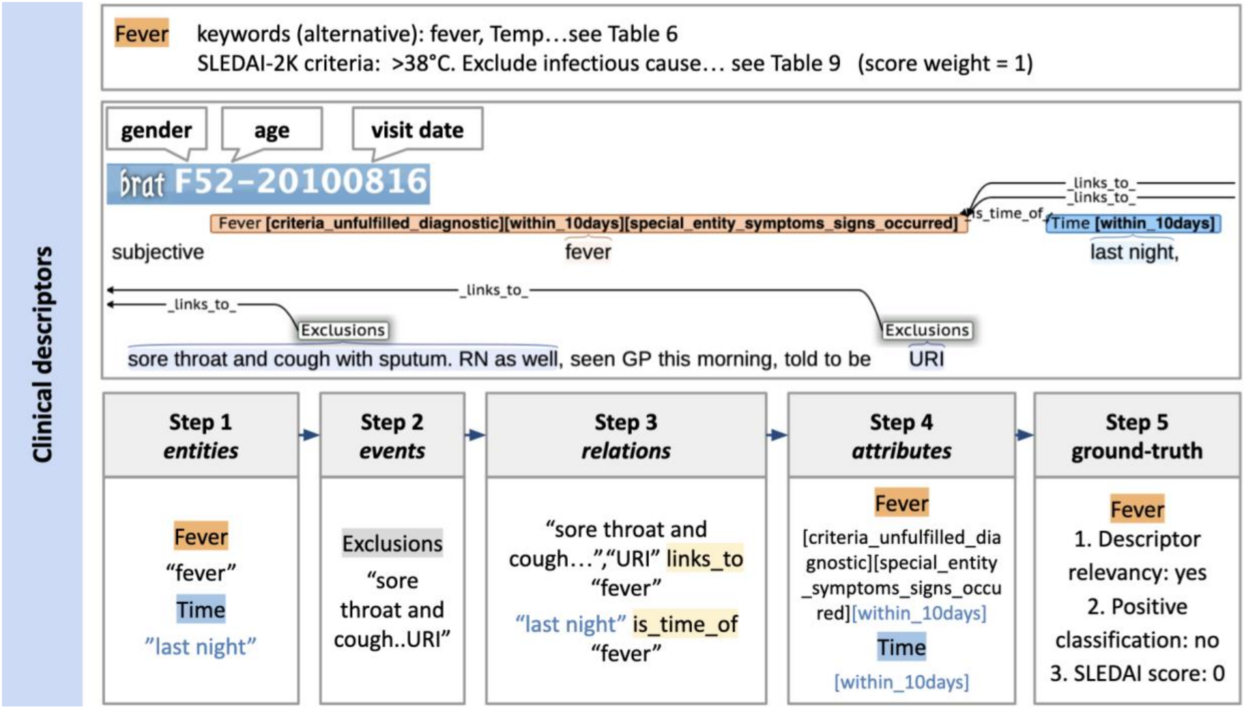

*Figure 3. A demo annotation of a Laboratory descriptor*

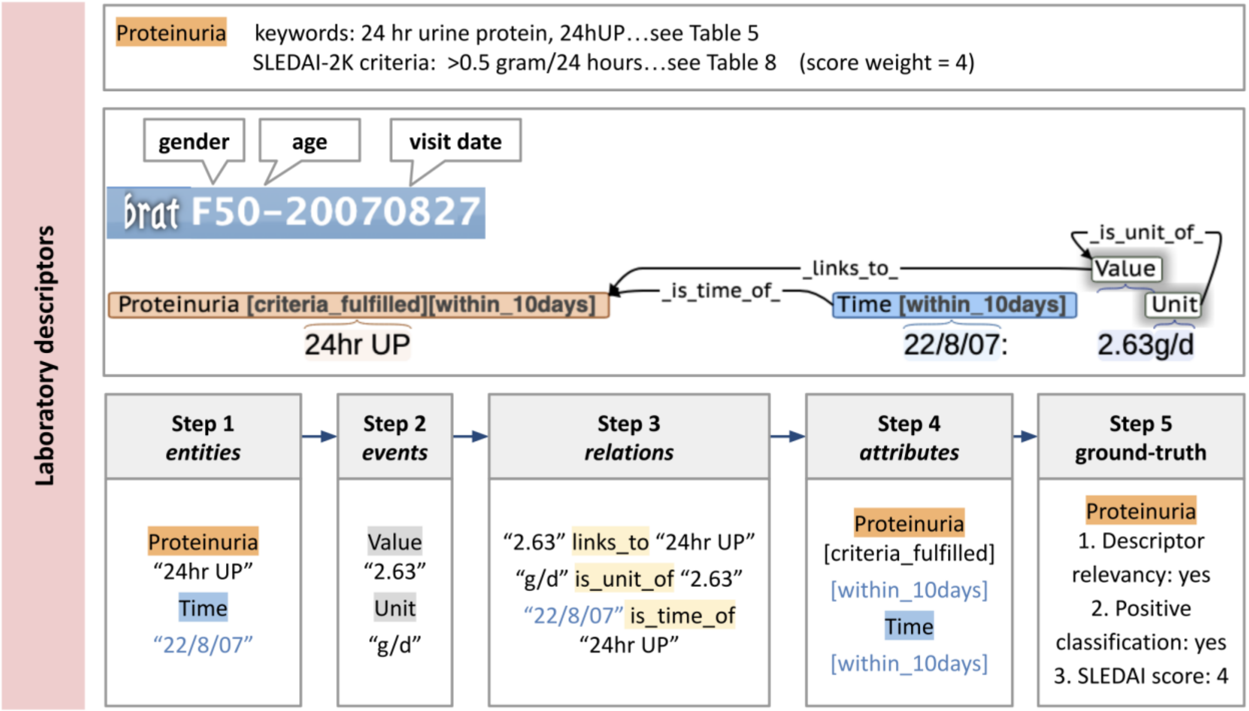

*Table 7. Examples for SLEDAI-2K annotation with a complete note*

<table>
<tr><td colspan="2">Annotations in BRAT<br>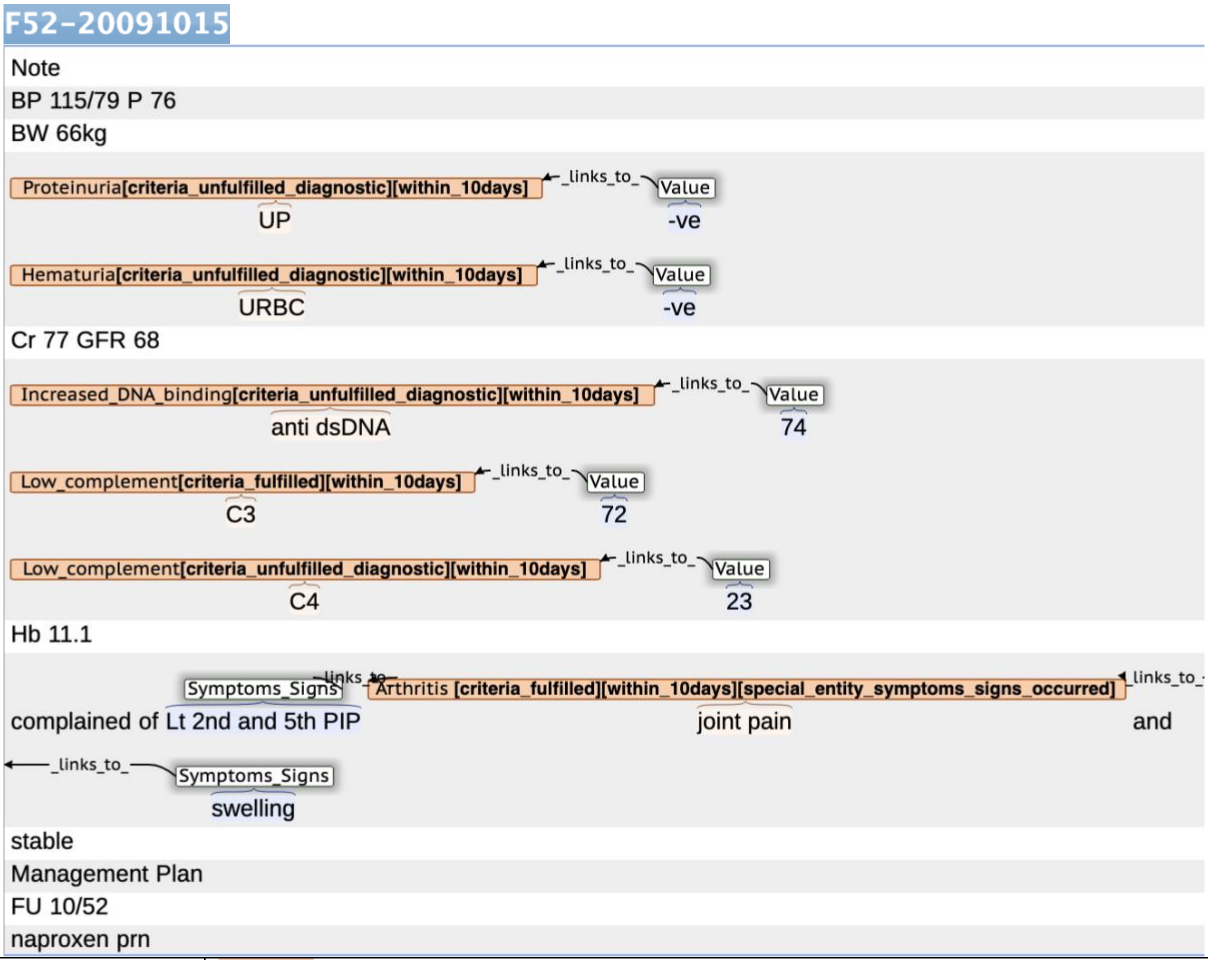
</td></tr>
<tr><td>Descriptor relevancy</td><td>Arthritis: yes. No diagnostic keywords, use alternative keyword “joint pain”.<br>Proteinuria: yes<br>Hematuria: yes<br>Increased DNA Binding: yes<br>Low Complement: yes</td></tr>
<tr><td>Descriptor classification</td><td>Positive:<br>Arthritis: as shown in Table 8, the description of “joint pain of PIPs + swelling” fulfilled criteria A of “two or more joints with pain and signs of inflammation (e.g., swelling)”. Given time within 10 days, Arthritis is positive.<br>Low Complement: As shown in Table 9, the reference for C3 is <76 mg/dL. 72<76, therefore, the criteria are fulfilled. Given time within 10 days, Low Complement 3 is positive.<br>Negative:<br>Increased DNA Binding: As shown in Table 9, the reference for dsDNA is >67.5 IU/ml, and the assumed method is ELISA. 74 < 135, so the criteria are unfulfilled.<br>Hematuria (URBC -ve) not fulfilled.<br>Proteinuria (UP -ve) - not fulfilled.</td></tr>
<tr><td>Total score</td><td>Arthritis (Score: 4) + Low Complement (Score: 2) = 6</td></tr>
</table>

### 3. Chapter 3-detailed criteria for SLEDAI-2K and annotation example for each descriptor

This study utilizes the SLEDAI for 30-day window (5). Consequently, scoring a descriptor requires meeting two criteria: 1) the disease manifestation must fulfill the SLEDAI definition, with necessary differential diagnoses excluded; and 2) it must have been present or occurred within the preceding 30 days.

Recognizing that the original SLEDAI-2K definitions can lack clarity or practicality for center-specific application, we refined them to enhance consistency between physicians and LLMs (see Tables 8 and 9 below). Where the SLEDAI-2K definition was ambiguous or impractical, we mainly incorporated supplementary criteria developed by established organizations, including the American College of Rheumatology (ACR) (7-9), the European League Against Rheumatism (EULAR) (10), and the Systemic Lupus International Collaborating Clinics (SLICC) (11) (Table 10). For each descriptor, we provide detailed rationales for the definitions used, cite source references, and include annotation examples to illustrate the application of our evaluation criteria, as seen in below sections.

*Table 8. Detailed SLEDAI-2K criteria for the **Clinical descriptor**. The sources with references to supplementary definitions used are discussed in Chapter 3.*

| **Clinical descriptors** | **definition** |
|---|---|
| Seizure | Diagnostic criteria:<br>A with or without the presence of B:<br><br>A. Independent description of seizure by a reliable witness<br>B. EEG abnormalities<br><br>Exclusions:<br>- Medications: quinolones, imipenem<br>- Alcohol and drug withdrawal (phenothiazines, antipsychotics)<br>- electrolyte imbalance (acidosis, serum sodium, calcium, and hypoglycemia)<br>- organ failure (hepatic and renal failure)<br>- infection<br>- due to past irreversible CNS damage<br>- Vasovagal syncope<br>- Cardiac syncope<br>- Hysteria<br>- Hyperventilation<br>- Tics<br>- Narcolepsy and cataplexy<br>- Labyrinthitis<br>- Subarachnoid hemorrhage<br>- Trauma<br>- Panic attacks, conversion disorders, and malingering<br>- hypersensitivity encephalopathy |
| Psychosis | Diagnostic criteria:<br>All of the following:<br><br>A. At least one of the following:<br><br>1. Delusions<br>2. Hallucinations without insight (visual, olfactory, gustatory, tactile, or auditory) |

| | |
|---|---|
| | 3. Disorganized thinking (speech) (frequent derailment; loose associations; incoherence; "word salad")<br>4. Grossly disorganized or abnormal motor behavior (childlike "silliness"; unpredictable agitation; catatonic behavior; negativism; mutism and stupor; catatonic excitement; stereotyped movements, staring; grimacing, mutism; echoing of speech)<br>5. Negative symptoms (diminished emotional expression; avolition; alogia; anhedonia; asociality)<br><br>B. The disturbance causes clinical distress or impairment in social, occupational, or other relevant areas of functioning.<br>C. The disturbance does not occur exclusively during the course of a delirium.<br>D. The disturbance is not better accounted for by another mental disorder (e.g., mania).<br>Exclusions:<br>- Uremia (BUN > 40 mg/dL)<br>- Substance- or drug-induced psychotic disorder (including NSAIDs, antimalarials)<br>- acidosis, or electrolyte imbalance<br>- delirium<br>- Primary psychotic disorder unrelated to SLE (e.g., schizophrenia)<br>- Psychologically mediated reaction to SLE (brief reactive psychosis with major stressor) |
| Organic brain syndrome | Dianostic Synonymous:<br>organic brain syndrome<br>organic brain disorder<br>acute confusional state<br>delirium<br>encephalopathy<br>neurocognitive disorders<br><br>Diagnostic criteria:<br>All of the following:<br><br>A. A disturbance in attention (i.e., reduced ability to direct, focus, sustain, and shift attention) or awareness (reduced orientation to the environment)<br>B. Rapid onset and fluctuating clinical features: develops over a short period of time (usually hours to a few days), represents a change from baseline attention and awareness, and tends to fluctuate in severity during the course of a day<br>C. Plus at least two of the following:<br><br>1. perceptual disturbance (misinterpretations, illusions, or hallucinations)<br>2. incoherent speech<br>3. insomnia or daytime drowsiness<br>4. increased or decreased psychomotor activity (Hyperactive/ Hypoactive)<br><br>Exclusions:<br>- Primary mental/neurologic disorder not related to SLE.<br>- Metabolic disturbances (glucose > 25; serum sodium < 125 or > 155; serum calcium > 3)<br><br>NB: Preexisting cognitive deficits are not an exclusion. If acute confusional state is superimposed on preexisting cognitive deficits, diagnose both. |
| Visual disturbance | Diagnostic criteria:<br>Either ① or ② |

| | |
|---|---|
| | ① Retinopathy or choroidopathy with one criteria below, usually bilateral:<br><br>A. cytoid bodies (cotton-wool spots or retinal soft exudates)<br>B. retinal hemorrhages<br>C. serous exudate in the choroid<br>D. serous hemorrhages in the choroid<br><br>② Optic neuritis<br>Clinical criteria (A), (B), or (C) if seen acutely; or at least one Paraclinical criteria with a medical history suggestive of optic neuritis<br><br>1. Clinical criteria:<br><br>A. Monocular, subacute loss of vision associated with orbital pain worsening oneye movements, reduced contrast and colour vision, and relative afferent pupillary deficit (RAPD)<br>B. Painless with all other features of (A).<br>C. Binocular loss of vision with all features of (A) or (B).<br><br>2. Paraclinical criteria:<br><br>A. OCT: Corresponding optic disc swelling acutely or an inter-eye difference in the mGCIPL of >4% or >4 µm or in the pRNFL of >5% or >5 µm within 3 months after onset.<br>B. MRI: Contrast enhancement of the symptomatic optic nerve and sheaths acutely or an intrinsic signal (looking brighter) increase within 3 months.<br><br>Exclusions:<br>- hypertension (BP > 180/120)<br>- Infectious, post-infectious or post-vaccination<br>- drug caused<br>- drusen (for cytoid bodies)<br>- Autoimmune ON: multiple sclerosis (MS), Aquaporin4 IgG antibodies-associated neuromyelitis optica spectrum disorder (NMOSD) or Anti-myelin oligodendrocytes glycoprotein antibody-associated disease (MOGAD) |
| Cranial nerve disorder | Diagnostic criteria:<br>New onset of sensory or motor neuropathy involving cranial nerves.<br>Syndrome corresponding to specific nerve function:<br>I. Olfactory nerve: Loss of sense of smell, distortion of smell, and loss of olfactory discrimination<br>II. ~~Optic nerve: Decrease or loss of visual acuity, diminished color perception, afferent pupillary defect and visual field deficits~~<br>II. Optic nerve: In the SLEDAI-2K scoring system, optic nerve disorder is categorized under visual disturbance. Please refer to the specific criteria for visual disturbance for details.<br>III. Oculomotor nerve: Ptosis of the upper eyelid and inability to rotate eye upward, downward, or inward (complete lesion), and/or dilated nonreactive pupil and paralysis of accommodation (interruption of parasympathetic fibers only)<br>IV. Trochlear nerve: Extorsion and weakness of downward movement of affected eye<br>V. Abducens nerve: Weakness of eye abduction<br>VI. Trigeminal nerve: Paroxysm of pain in lips, gums, cheek, or chin initiated by stimuli in |

| | |
|---|---|
| | trigger zone (trigeminal neuralgia) and sensory loss of the face or weakness of jaw muscles<br>VII. Facial nerve: Unilateral or bilateral paralysis or facial expression muscles, impairment of taste, and hyperacusis (painful sensitivity to sounds)<br>VIII. Vestibulo-cochlear nerve: Deafness, tinnitus (cochlear), dizziness and/or vertigo (vestibular)<br>IX. Glossopharyngeal nerve: Swallowing difficulty, deviation of soft palate to normal side, anesthesia of posterior pharynx and/or glossopharyngeal neuralgia (unilateral stabbing pain in root of tongue and throat, triggered by coughing, sneezing, swallowing, and pressure on ear tragus)<br>X. Vagus nerve: Soft palate droop, loss of the gag reflex, hoarseness, nasal voice, and/or loss of sensation at external auditory meatus.<br>XI. Accessory nerve: Weakness and atrophy of sternocleidomastoid muscle and upper part of trapezius muscle.<br>XII. Hypoglossal nerve: Paralysis of one side of tongue with deviation to the affected side<br><br>Exclusions:<br>- nerve palsy caused by stroke, seizure, or intracranial hypertension<br>- Skull fracture<br>- Tumor: meningioma, carcinomatous meningitis, aneurysm<br>- Infection: herpes zoster, neuroborreliosis, syphilis, mucormycosis<br>- Miller Fisher syndrome |
| Lupus Headache | Diagnostic criteria:<br>Either type of headache below, and should be severe (disabling headache), persistent (lasts ≥3 days) also nonresponsive to narcotic analgesia.<br><br>A. Migraine<br><br>Migraine withot aura: Idiopathic, recurrent headache manifested by attacks lasting 4-72 hours. Typical characteristics are unilateral location, pulsating quality, moderate to severe intensity, aggravation by routine physical activity, and associated with nausea, vomiting, photo- and phonophobia. At least 5 attacks fulfilling the above criteria.<br>Migraine with aura: Idiopathic, recurrent disorder manifested by attacks of neurologic symptoms localizable to cerebral cortex or brain stem, usually gradually developing over 5-20 minutes and lasting less than 60 minutes. Headache, nausea, and/or photophobia usually follow neurologic aura symptoms directly or after an interval of less than 1 hour. Headache usually lasts 4-72 hours, but may be completely absent.<br><br>B. Tension headache (episodic tension type headache)<br><br>Recurrent episodes of headaches lasting minutes to days. Pain typically pressing/tightening in quality, of mild to moderate intensity, bilateral in location, and does not worsen with routine physical activity. Nausea is rare, but photophobia and phonophobia may be present. At least 10 previous headaches fulfilling these criteria.<br><br>C. Cluster headache<br><br>Attacks of severe, strictly unilateral pain, orbital, supraorbital, and/or temporal, usually lasting 15-180 minutes and occurring from at least once every other day up to 8 times per day. Associated with one or more of the following: conjunctival injection, lacrimation, nasal congestion, rhinorrhea, forehead and facial sweating, myosis, ptosis, eyelid edema. Attacks occur in series for weeks or months ("cluster" periods) separated by remissions of usually months or years. |

| | |
|---|---|
| | D. Headache from intracranial hypertension (Pseudotumor cerebri, benign intracranial hypertension)<br><br>All of the following:<br>Increased intracranial pressure (200 mm HiO) measured by lumbar puncture<br>Normal neurologic findings except for papilledema and possible nerve VI palsy<br>No mass lesion and no ventricular enlargement on neuroimaging<br>Normal or low protein and normal white cell count in CSF<br>No evidence of venous sinus thrombosis<br><br>E. Intractable headache, nonspecific<br><br>Exclusions:<br>- Aseptic meningitis (including drug-induced)<br>- Drug-induced pseudotumor cerebri (oral contraceptives, sulfonamides, trimethoprim, etc.)<br>- CNS infection (meningitis/encephalitis)<br>- Tumors and other structural lesions<br>- Low intracranial pressure<br>- Trauma<br>- Metabolic headache that remits with elimination of cause (carbon monoxide exposure)<br>- Withdrawal (caffeine, etc.)<br>- Seizure/postictal state<br>- Sepsis<br>- Intracranial hemorrhage or vascular occlusion |
| CVA | Diagnostic criteria:<br>One of the following and supporting radio imaging study, exclused arteriosclerosis:<br><br>A. Stroke syndrome: acute focal neurologic deficit persisting more than 24 hours (or lasting less than 24 hours with CT or MRI abnormality consistent with physical findings/symptoms<br>B. Transient ischemic attack: acute, focal neurologic deficit with clinical resolution within 24 hours (without corresponding lesion on CT or MRI)<br>C. Subarachnoid and intracranial hemorrhage: bleeding documented by CSF findings or MRI/CT<br>D. Sinus thrombosis: Acute, focal neurologic deficit in the presence of increased intracranial pressure<br>NB: The finding of unidentified bright objects on MRI without clinical manifestations is not classified at the present time.<br><br>Exclusions:<br>- Arteriosclerosis<br>- Infection with space occupying lesions in the brain<br>- Intracranial tumor<br>- Trauma<br>- Vascular malformation<br>- Hypoglycemia |
| Vasculitis | Diagnostic criteria:<br>Either one：<br><br>A. clinician observed cutaneous vasculitis with at least one of the following discription of lesions: Ulceration, gangrene, lender finger nodules, periungual infarction (or necrosis), splinter hemorrhages |

| | |
|---|---|
| | B. biopsy or angiogram proof of cutaneous vasculitis<br><br>Exclusions:<br>- mimics of vasculitis (Atheroembolic disease, Atheromatous vascular disease, Anti-phospholipid syndrome, Multiple myeloma, Infective endocarditis, Para-neoplastic syndromes, Genetic vascular disorders (e.g. Marfan's syndrome), Autoinflammatory syndromes, Hypersensitivity reactions, Cocaine and amphetamine abuse)<br>- Infections (Tuberculosis, Hepatitis B, Hepatitis C, HIV)<br>- Malignancy (Lymphoma, Solid organ malignancy)<br>- Drugs (Penicillamine, Propylthiouracil, Hydralazine, Minocycline, Cocaine)<br>- Environmental exposure (including Dusts and Silica) |
| Arthritis | Diagnostic criteria:<br>Either condition:<br><br>A. two or more joints with pain and signs of inflammation (e.g., tenderness, swelling, or effusion)<br>B. synovitis involving 2 or more joints characterized by swelling or effusion<br>C. two or more joints tenderness and 30 minutes or more of morning stiffness<br><br>Exclusions:<br>- infection (a septic joint or as osteomyelitis): tuberculosis, Bacterial, viral<br>- stress fractures<br>- Crystal arthritis: Gout, calcium pyrophosphate deposition disease<br>- Noninflammatory arthritis, such as osteoarthritis and avascular necrosis (AVN) (especially for hip, knee or shoulder)<br>- Rheumatoid arthritis |
| Myositis | Diagnostic criteria:<br>All of the following:<br><br>A. proximal muscle aching (myalgias)/weakness,<br>B. associated with elevated creatine phosphokinase/aldolase or electromyogram (EMG) changes or a biopsy showing myositis<br><br>Exclusions:<br>- CK elevation was unexplained, isolated, felt to be related to rhabdomyolysis, exertion, infection, or the toxic effect of medication (i.e., statin, hydroxychloroquine-induced myotoxicity)<br>- cancer-associated myositis<br>- metabolic muscle disease |
| Rash | Diagnostic criteria:<br>Either one form of lupus rash from these three class of lupus erythematosus:<br><br>A. Acute cutaneous lupus erythematosus (ACLE), either one:<br><br>1. lupus malar rash;<br>2. bullous lupus;<br>3. toxic epidermal necrolysis variant of SLE;<br>4. maculopapular lupus rash;<br>5. photosensitive lupus rash in the absence of dermatomyositis;<br><br>B. subacute cutaneous lupus erythematosus (SCLE), either one: |

| | |
|---|---|
| | 1. Annular form<br>2. Papulosquamous form<br><br>C. Chronic cutaneous lupus erythematosus (CCLE), either one:<br><br>1. classical discoid lupus erythematosus (DLE)<br>2. localized DLE (above the neck)<br>3. generalized DLE (above and below the neck)<br>4. hypertrophic (verrucous) lupus<br>5. lupus panniculitis (profundus)<br>6. mucosal lupus<br>7. lupus erythematosus tumidus<br>8. chillblains lupus (CHLE, pernio)<br>9. lichen planus overlap<br><br>Note: A "lupus rash" was only scored if it was clinically diagnosed as such. Descriptions of skin changes based solely on morphology and location were considered insufficient for scoring a “lupus rash”, unless the rash was documented as a classic manifestation (e.g., typical malar distribution). Furthermore, rashes described solely as "skin damage" or residual changes were not scored, as the SLEDAI-2K is designed to capture active inflammatory rash.<br><br>Here is the vocabulary to help distinguish between lupus activity versus damage:<br>-Activity (erythema): pink, faint erythema, pinkish, faint erythema, faintly erythematous, mild erythema, mildly erythematous, mild redness, mildly red, erythematous, erythema, moderate erythema, moderately erythematous. moderate erythematous, red, redness, reddish, dark red, deep red, deeply red, deep reddish, deeply reddish, purple, purplish, violaceous<br><br>-Activity (scale/hypertrophy): Scale, scaling, scaly, follicular plugging, follicular prominence, carpet tacking, carpet tack sign, perifollicular scale<br>Hypertrophy, hypertrophic, hyperkeratosis, hyperkeratotic<br><br>-Damage (Dyspigmentation): Hypopigmentation, hypopigmented, hyperpigmentation, hyperpigmented, dyspigmentation, dyspigmented, pigmentary changes, brown, depigment, white<br><br>-Damage (scarring/atrophy/panniculitis): Scarring, scar, Atrophic scarring, atrophic, panniculitis, lipoatrophy, atrophic scar<br><br>- Exclusions for ACLE:<br>• Localized form: rosacea, seborrheic eczema, perioral dermatitis, tinea faciei, erysipelas<br>• Generalized form: dermatomyositis, viral and drug-induced rash, erythema multiforme, TEN<br>• Bullous lupus erythematosus (BLE): Epidermolysis bullosa acquisita, dermatitis herpetiformis (Duhring's disease), bullous pemphigoid, linear IgA-dermatosis, drug-induced bullous disorder, porphyria cutanea tarda<br><br>- Exclusions for SCLE:<br>• Psoriasis vulgaris, tinea corporis, mycosis fungoides, erythema annulare centrifugum, dermatomyositis, pityriasis rubra pilaris, nummular eczema, drug-induced rash, seborrheic eczema, erythema multiforme /TEN, erythema gyratum repens |

| | |
|---|---|
| | - Exclusions for Discoid lupus erythematosus (DLE).:<br>• Actinic keratosis, tinea faciei, sarcoidosis, lupus vulgaris<br><br>- Exclusions for Lupus erythematodes profundus (LEP):<br>• Various forms of panniculitis, malignant lymphoma (especially subcutaneous panniculitic T-cell lymphoma), subcutaneous sarcoidosis, panarteritis nodosa, morphea profunda, subcutaneous granuloma annulare<br><br>- Exclusions for Chilblain lupus erythematosus (CHLE):<br>• Pernio (chilblains), lupus pernio (chronic form of skin sarcoidosis of the acral regions), acral vasculitis/vasculopathy<br><br>- Exclusions for Lupus erythematosus tumidus (LET):<br>• Lymphocytic infiltration Jessner-Kanof or erythema arciforme et palpabile (see text), polymorphic light eruption, pseudolymphoma, B-cell lymphoma, plaque-like cutaneous mucinosis, solar urticaria |
| Alopecia | Diagnostic criteria:<br>Abnormal, patchy, or diffuse nonscarring hair loss or alopecia: Diffuse thinning or hair fragility with visible broken hairs in the absence of other causes such as alopecia areata, drugs, iron deficiency and androgenic alopecia<br><br>Exclusions:<br>- alopecia areata<br>- iron deficiency<br>- androgenic alopecia<br>- scarring alopecia |
| Mucosal ulcers | Diagnostic criteria:<br><br>Oral or nasal (nasopharyngeal) ulceration observed by a clinician, including palate, buccal, tongue or nasal ulcers<br><br>Exclusions:<br>- Vasculitis<br>- Behcet's<br>- infection (herpes)<br>- inflammatory bowel disease<br>- reactive |
| Pleurisy | Diagnostic criteria:<br>Both A and B:<br><br>A. Pleuritic chest pain (typically sharp, worse with inspiration, improved by shallow breathing)<br>B. pleural rub OR pleural thickening OR imaging confirmed pleuritic effusion (such as ultrasound, x-ray, CT scan, MRI)<br>Exclusions:<br>- infection<br>- uremia (BUN > 40 mg/dL) |
| Pericarditis | Diagnostic criteria:<br>Both A and B:<br><br>A. pericardial pain (typically sharp, worse with inspiration, improved by leaning forward) |

| | |
|---|---|
| | B. pericardial rub OR EKG with new widespread ST-elevation or PR depression OR new/worsen imaging confirmed pericardial effusion (such as ultrasound, x-ray, CT scan, MRI)<br><br>Exclusions:<br>- infection (coxsackievirus, mycoplasma, tuberculosis)<br>- uremia (BUN > 40 mg/dL)<br>- Dressler's pericarditis (post-myocardial infarction syndrome) |
| Fever | Diagnostic criteria:<br>>38°C (Exclude infectious cause)<br><br>Exclusions:<br>- Infection (usually have high WBC, high CRP or definite localizing infective foci)<br>- malignancy (especially lymphoma) |

An additional criterion should be fulfilled for 8-point descriptor, including Seizure, Psychosis, Organic Brain Syndrome, Visual Disturbance, Cranial Nerve Disorder, Lupus Headache, and Vasculitis: The doctors' treatment plans must include the initiation or escalation of steroids (e.g., prednisone, prednisolone, glucocorticoids), immunosuppressants (e.g., mycophenolate mofetil, cyclophosphamide, azathioprine, cyclosporine, tacrolimus), or biologics (e.g., belimumab, rituximab). Without these treatment plans, the diagnosis remains uncertain. Immunosuppressant optimization for steroid minimization or switching immunosuppressants for maintenance does not constitute therapy escalation.

For non-8-point descriptors with documented positive symptoms/signs/tests but not fully meeting diagnostic thresholds, either both of the following may support scoring: a) positive response to lupus-directed therapeutic trials (e.g., steroids, antimalarial, immunosuppressants, or biologics) or b) doctors' treatment intention of a lupus-directed therapy. Exception: Steroid-responsive rash alone does not establish lupus etiology.

*Table 9: Detailed SLEDAI-2K definitions for Laboratory descriptors. The sources of the supplementary definitions used are discussed in the sections below.*

| **Laboratory descriptors** | **Definition** |
|---|---|
| Urinary casts | Heme-granular or red blood cell casts. |
| Hematuria | >5 red blood cells/high-power field with abnormal urine protein (urine protein dipstick positive or urine protein/creatinine > 0.15 mg/mg (15mg/mmol or 0.15g/g) or 24-hour urine protein: >0.15 gram/24 hours). Exclude stone, infection or other cause (menstruation).<br>NB: in our study, urine RBC ++ or RBC > 30-58/µL can be considered as >5 red blood cells/high-power field. Microscopic haematuria is most commonly defined as >3 red blood cells per high-power field on urinary microscopy. |
| Proteinuria | Either one:<br><br>A. 24-hour urine protein: >0.5 gram/24 hours.<br>B. Urine protein/creatinine > 0.5 mg/mg (50mg/mmol or 0.5g/g)<br>NB: If the two results are conflict at the same time, the 24-hour protein result shall prevail. While dipstick urinalysis provides only a qualitative assessment and is generally indicative of uncertain proteinuria, a reading of 3+ or greater can be scored as significant proteinuria when correlated with a physician's clinical impression of a lupus flare (i.e., one necessitating escalation of therapy for lupus nephritis).<br><br>Exclude stone, infection or other cause. |

<table>
<tr><td></td><td>If the urine protein/creatinine value is more than 20, we suspect the unit should be mg/mmol. Interpret UPC values using heuristic unit detection: Suspect mg/mmol for values >20, mg/mg for values <5, and flag uncertain if in between.</td></tr>
<tr><td>Pyuria</td><td>>5 white blood cells/high-power field with abnormal urine protein (urine protein dipstick positive or urine protein/creatinine > 0.15 mg/mg (15mg/mmol or 0.15g/g) or 24-hour urine protein: >0.15 gram/24 hours). Exclude infection.<br>NB: in our study, urine WBC ++ or WBC ≥ 10-50/μL can be considered as ≥ 5 white blood cells/high-power field.</td></tr>
<tr><td>Low complement</td><td>Decrease in CH50,C3, or C4 below the lower limit of normal for testing laboratory.<br><br>Reference intervals for complement levels are laboratory-dependent. Values should first be interpreted based on the flags provided in the clinical reports (e.g., nl, low, high) or the reference range provided by the notes. When no flags or reference range were provided, the reference intervals below from our hospital should be applied. A lower complement than the reference range can fulfill the criteria.<br><br>
<table>
<tr><td>Time period</td><td>item</td><td>Reference</td></tr>
<tr><td>24/12/2018 till now</td><td>C3</td><td><90 mg/dL</td></tr>
<tr><td>before 24/12/2018</td><td>C3</td><td><76 mg/dL</td></tr>
<tr><td>24/12/2018 till now</td><td>C4</td><td><10 mg/dL</td></tr>
<tr><td>before 24/12/2018</td><td>C4</td><td><9</td></tr>
</table>
<br>Interpret C3 values using heuristic unit detection: Presume mg/dL scale for values more than 10, g/L for values below 2, and flag uncertain values if in between.</td></tr>
<tr><td>Increased DNA binding</td><td>Increased DNA binding above laboratory reference range, except ELISA: twice above laboratory reference range.<br><br>NB: If the methodology for increased DNA binding was not mentioned, we assumed it was ELISA. Qualitative values (such as "high", "+", "rising", or similar terms) should be considered criteria_uncertain for ELISA but criteria_fulfilled for other methods.<br><br>The cut-off for the ELISA method was based on reference range provided by clinical notes when available; otherwise, our QMH's reference range for ELISA was applied:
<table>
<tr><td>Time period</td><td>laboratory reference range</td></tr>
<tr><td>1/2/2021 till now</td><td>>10 IU/ml</td></tr>
<tr><td>21/1/2013 to 1/2/2021</td><td>>25 IU/ml</td></tr>
<tr><td>before 21/1/2013</td><td>>67.5 IU/ml</td></tr>
</table></td></tr>
<tr><td>Thrombocytopenia</td><td>Thrombocytopenia (platelets <100,000/$mm^3$ or <100 ×$10^9$/L), in the absence of other known causes such as drugs, portal hypertension, and thrombotic thrombocytopenic purpura<br><br>Note: In clinical practice, the default unit is 1000/mm3 or 109/L for platelets and leukocyte count.</td></tr>
<tr><td>Leukopenia</td><td>Leucocyte count, < 3,000/$mm^3$ or < 3 ×$10^9$/L white blood cells. Exclude Felty's, drug causes and portal hypertension.<br><br>Note: In clinical practice, the default unit is 1000/mm3 or 109/L for platelets and leukocyte count.</td></tr>
</table>

### 1. Seizure

#### 1.1. Clinical definition of concepts

According to the International League Against Epilepsy (ILAE), an ***epileptic seizure*** is a transient sign and/or symptom of the brain generated by aberrant neuronal discharge from the brain. **Epilepsy** is defined as recurrent (i.e. ≥ 2) epileptic seizures with enduring alteration in the brain (12). In the ILAE 2017 seizure classification, seizures are divided into three categories according to the onset: ***focal onset seizures (corresponding to the 1981 term "partial seizures")***, which occur in one cerebral hemisphere, ***generalized onset seizures***, which involve both cerebral hemispheres, and ***unknown onset*** (13, 14). Seizures can affect at least one of the following brain functions: sensory, motor, and autonomic function; consciousness; emotional state; memory; cognition; or behavior, with motor and awareness (knowledge of self and environment) being the most important signs and symptoms of seizures (semiology) (12). Semiology is used as a basis for the three categories of seizures. Based on these two important semiologies, ***focal onset seizures*** are subdivided into focal awareness (corresponding to the 1981 term 'simple partial seizure') versus focal impaired awareness (corresponding to the 1981 term 'complex partial seizure') and motor versus nonmotor. **Generalized onset seizures** are subdivided into motor and non-motor (absence) seizures (most are associated with impaired awareness). (13, 14)
***Electroencephalography*** (EEG) is a routine and primary diagnostic tool for seizures. To rule out non-epileptic behavioral symptoms, simultaneous observations of both behavioral and EEG changes (spikes and sharp waves) allow confirmation of seizures with a high degree of confidence (15). ***Subclinical seizures***, often referred to as ***electrographic seizures***, exhibit prolonged epileptiform EEG patterns but do not exhibit clinical symptoms. Moreover, epileptiform EEG episodes can be occasionally found in normal people. Taken together, the fact that documenting actual seizure episodes is uncommon during routine EEG, the diagnosis of seizures depends heavily on clinical judgment of historical episodes (16).

#### 1.2. Common ways to describe this concept

The ILAE 2017 provided a glossary and rules for classifying seizures. An ideal clinical term for seizure type contains two parts: categories of seizures with key signs and symptoms of seizures (semiology). Except for clinical seizure type, common descriptors of behaviors during and after seizures, and mapping of old to new seizure classifying terms were also taken into consideration for keywords (14).

Table. Seizure classifying terms and abbreviations according to ILAE 2017

| **Classification or categories of onset** | | **Seizure type (abbreviations)** | **Signs and symptoms (semiology)** | **Example** |
|---|---|---|---|---|
| Focal onset [Partial/Occipital lobe/Parietal lobe/Frontal lobe] | Either one: | Focal aware seizure (FAS) [Simple partial seizure] | / | focal aware nonmotor autonomic seizures |
| | | Focal impaired awareness seizure (FIAS) [Complex partial seizure or Dialeptic] | / | |
| | Either one: | Focal motor seizures (FMS) | atonic (stiffening) [Drop attack], automatisms, clonic (sustained rhythmic jerking), epileptic spasms, or hyperkinetic, myoclonic, or tonic [Drop attack] activity | |
| | | Focal nonmotor-onset (FNMS) | autonomic, behavior arrest, cognitive, emotional, or sensory dysfunction | |

| | | | | |
|---|---|---|---|---|
| Focal to bilateral [Secondarily generalized] | | Focal to bilateral tonic–clonic seizure (FBTCS) [Grand mal] | / | / |
| Generalized onset | Either one： | Generalized motor seizure (GMS) | atonic, clonic, epileptic spasms, myoclonic, myoclonic–atonic, myoclonic–tonic–clonic, tonic, or tonic–clonic | Generalized tonic–clonic<br><br>(Generalized) absence |
| | | Generalized non-motor (absence) seizures (GAS) [absence or Petit mal] | prominent myoclonic activity or eyelid myoclonia | |
| | | Generalized tonic–clonic seizure (GTCS) [Grand mal] | | |
| Unknown onset | | unknown onset | motor, nonmotor, tonic–clonic [Grand mal], epileptic spasms, or behavior arrest. | |

Note: inside [] are old terms mapping to new seizure classifying terms.(14)

#### 1.3. SLEDAI-2K definition in this study

As shown in Table 8, the diagnostic criteria for this study were primarily sourced from the supplementary materials of the ACR nomenclature and case definitions for NPSLE in 1999 (9). We incorporated exclusion criteria from the ACR 1997 classification criteria (7, 8) and another publication (17). Of note, while electroencephalography (EEG) is a sensitive diagnostic tool for epilepsy, it is not mandatory for diagnosis. Clinicians can diagnose seizures based on a reliable witness account alone.

The SLEDAI-2K does not restrict seizure types to those secondary to SLE but emphasizes recent onset and the exclusion of other causes, which is a principle consistent with the ACR 1997 (7, 8) and EULAR/ACR 2019 Classification Criteria (10). ACR also mentioned that the seizure criteria employed are based on the International League Against Epilepsy (ILAE) terminology (18). A scoping review indicates that the most common seizure presentation in SLE is tonic-clonic (60–88%), including secondarily generalized seizures, followed by seizures with impaired consciousness (12%–18%) and seizures with retained consciousness (3.5%–19%) (19).

#### 1.4. Examples present of SLEDAI-2K score

| How to determine. | Examples |
|---|---|
| A focal seizure was diagnosed clinically.<br>This diagnosis was supported by an EEG that revealed focal temporal activity, confirming ictal abnormalities.<br>Consequently, the treatment was steroids. | Complaint & HPI/PMH:<br>[Symptoms_Signs]<br>c/o focal seizures (dizziness → left facial paralysis), diplopia, epigastric pain, alopecia, photosensitivity.<br>No prior SLE dx. Partial response to IV fluids/iron.<br><br>PE:<br>[Symptoms_Signs]<br>Neuro: Conscious, oriented; dysdiadochokinesia (-).<br>[Symptoms_Signs]<br>EOM full with no definite binocular diplopia on exam<br>Derm: Alopecia, photosensitivity.<br>Abd: Non-tender; no rash/organomegaly.<br><br>ANA+, anti-dsDNA+, hypocomplementemia, Hb 6.6↓, ESR 180↑, MCV 65↓. AChR and autoimmune encephabilities ab-ve<br>LP showed TCC<1, protein elevated, no oligoclonal band.<br>[Paraclinical_tests]<br>Imaging: Brain MRI/CT (-); EEG: focal temporal activity. Chest CT: no Thymoma.<br>EMG with RNS showed no evidence of MG.<br><br>Impression:<br>Seizure **[criteria_fulfilled][within_10days][treat_escalated]**<br>New SLE, neuro (focal seizures), hematologic (anemia)<br><br>Plan:<br>[Intention_to_treat]<br>Meds: HCQ 200mg BID, Pred 40mg OD, $FeSO_4$, $CaCO_3$/Vit D.<br>LEV 500mg BID (seizure prophylaxis). |

| | |
|---|---|
| The presentation included recurrent generalized tonic-clonic seizures accompanied by aphasia and vision loss. Brain MRI demonstrated worsening T2/FLAIR hyperintensity, indicating a recurrence of posterior reversible encephalopathy syndrome (PRES).<br>This led to an escalation of immunosuppressive therapy with increased doses of steroids and cyclophosphamide (CTX). | BP 230/130<br><br>Symptoms_Signs<br>Patient Complaint: Recurrent generalized tonic-clonic seizures, aphasia, vision loss, frequent vomiting.<br>HPI/PMH<br>Initial PRES (Dec 2022): Methylprednisolone 500mg IV ×3d + cyclophosphamide → resolved with BP control<br>Maintenance: Prednisone 50mg OD, cyclophosphamide 0.4g monthly → borderline BP control at home<br><br>PE<br>Neuro: No focal neurology<br>Renal: Grade 3 pitting edema (lower extremities)<br>Hematologic: Pallor (no active rash)<br>CV: Sinus tachycardia (HR 105 baseline)<br><br>Labs: Hb 10.7↓, Plt 9↓; Blood film no red cell fragment. U/A: protein 3+; Cr 515↑ (hemodialysis-dependent); C3 0.38↓, C4 0.12↓; ANA 1:160<br>Paraclinical_tests<br>MRI: PRES recurrence - worsened T2/FLAIR hyperintensity in frontal/parieto-occipital lobes vs prior. Repeat LP showed no evidence of infection. Dilantin level: subtherapeutic.<br>Impression<br>Seizure **[criteria_fulfilled][within_10days][treat_escalated]**<br>Active SLE with PRES recurrence:Seizures<br>Hematologic: Anemia/thrombocytopenia<br>Lupus nephritis: ESRD on HD + proteinuria<br>Immunologic: Hypocomplementemia<br><br>Management Plan<br>Meds:<br>Intention_to_treat<br>IV methylprednisolone 40 mg OD (replace oral prednisone)<br>Intention_to_treat<br>Step up CTX to NIH protocol. Step up dilantin, and consult neurology for AED titration.<br>Aggressive BP control: IV nitroprusside → transition to valsartan/amlodipine 80mg/5mg OD + carvedilol 25mg OD |

#### 1.5. Examples absent SLEDAI-2K score

| Why is it an exception? | List of examples (notes) |
|---|---|
| - Serology was quiescent.<br>- Imaging revealed evidence of old structural brain damage.<br>- The physician did not escalate immunosuppressant therapy.<br>- The seizures were not likely attributed to lupus flare. | History<br>1/2013: Anaemia, Hb 5.3 and breakthrough seizure<br>History<br>both Dilantin and epilim level was on the low side<br>History<br>(during in hospital stay)<br>OGD: NAD, colonoscopy?<br>History<br>2/2013: Breakthrough seizure with left sided twitching<br>Paraclinical_tests<br>CT brain: Encephalomalacia noted at right parietal and occipital lobes.<br>Paraclinical_tests<br>Old lacunar infarcts are seen at basal ganglia.<br>No intracranial mass, haemorrhage or midline shift is seen. No hydrocephalus or extra-axial collection is noted.<br>BP 144/ 83 P 69<br>no urine<br>SErology in jan -- C3/4 76/ 13<br>ds DNA 5.2<br>Cr 59 alb 24<br>Hb 7.9<br>? miss her colonoscopy appointment<br>but Fe stauts ok<br>B12 folate ok<br>Seizure, with LP done.<br>Paraclinical_tests<br>CSF protein 0.65<br>drug levels low<br>currently well.<br>Seizure **[criteria_unfulfilled_diagnostic][within_10days][wait_and_see]**<br>? seizure more frequent.<br>previously once every 1-2 months<br>Intention_to_treat<br>to step up epilim<br>see drug level again. |

### 2. Psychosis

#### 2.1. Clinical definition of concepts

According to SLEDAI-2K psychosis is altered ability to function in normal activity due to severe disturbance in the perception of reality. According to the Diagnostic and Statistical Manual of Mental Disorders (DSM-V) (20) published by the American Psychiatric Association (APA), the ***schizophrenia spectrum and other psychotic disorders*** are defined by abnormalities in one or more of the following five domains: ***delusions***, ***hallucinations***,

***disorganized thinking (speech)***, ***grossly disorganized or abnormal motor behavior (including catatonia)***, and ***negative symptoms***. ***Delusions*** are fixed beliefs that are not amenable to change in light of conflicting evidence. Their content may include a variety of themes (e.g., persecutory, referential, somatic, religious, grandiose). ***Hallucinations*** are perception-like experiences that occur without an external stimulus. They are vivid and clear, with the full force and impact of normal perceptions, and not under voluntary control. ***Disorganized thinking/Speech*** (formal thought disorder) is typically inferred from the individual's speech. The individual may switch from one topic to another (derailment or loose associations). Answers to questions may be obliquely related or completely unrelated (tangentiality). Rarely, speech may be so severely disorganized that it is nearly incomprehensible and resembles receptive aphasia in its linguistic disorganization (incoherence or "word salad"). ***Grossly disorganized or abnormal motor behavior*** may manifest itself in a variety of ways, ranging from childlike "silliness" to unpredictable agitation. ***Catatonic behavior*** is a marked decrease in reactivity to the environment. This ranges from resistance to instructions (negativism); to maintaining a rigid, inappropriate, or bizarre posture to a complete lack of verbal and motor responses (mutism and stupor). It can also include purposeless and excessive motor activity without an obvious cause (catatonic excitement). Other features are repeated stereotyped movements, staring, grimacing, mutism, and the echoing of speech. ***Negative symptoms*** account for a substantial portion of the morbidity associated with schizophrenia but are less prominent in other psychotic disorders. (20)

As recommended by ACR nomenclature and case definitions for NPSLE in 1999, the term "schizophrenia" is defined as a primary disease and should not be used to describe psychosis due to SLE (9). The case definition for lupus psychosis is, therefore, based on the DSM-IV entity "***psychosis due to a general medical condition***" (DSM-IV 293.81/82) (21) or on the DSM-V entity "***psychotic Disorder Due to Another Medical Condition***" (DSM-V 293.81/82) (20).

#### 2.2. Common ways to describe this concept

Table. Five domains for psychotic disorder according to DSM-V. (20)

| | |
|---|---|
| ***Delusions*** | Specify whether:<br>***Persecutory type:*** belief that one is going to be harmed, harassed, and so forth by an individual, organization, or other group(most common)<br>***Erotomanic type:*** when an individual believes falsely that another person is in love with him or her<br>***Referential type:*** belief that certain gestures, comments, environmental cues, and so forth are directed at oneself<br>***Grandiose type:*** an individual believes that he or she has exceptional abilities, wealth, or fame<br>***Jealous type:*** belief his or her spouse or lover is unfaithful<br>**Somatic** type: focus on preoccupations regarding health and organ function<br>***Nihilistic type:*** involve the conviction that a major catastrophe will occur<br>***Mixed type***<br>***Unspecified type***<br><br>Specify if:<br>With ***bizarre*** content: clearly implausible and not understandable to same-culture peers and do not derive from ordinary life experiences |
| ***Hallucinations*** | visual, olfactory, gustatory, tactile, or auditory hallucinations |
| ***Disorganized speech*** | frequent derailment; loose associations; incoherence; "word salad" |
| ***Grossly Disorganized or Abnormal Motor Behavior (Including Catatonia)*** | childlike "silliness"; unpredictable agitation; catatonic behavior; negativism; mutism and stupor; catatonic excitement; stereotyped movements, staring; grimacing, mutism; echoing of speech |
| ***Negative symptoms*** | Diminished emotional expression; avolition; alogia; anhedonia; asociality |

#### 2.3. SLEDAI-2K definition in this study

As shown in Table 8, the SLEDAI-2K definition of psychosis encompasses the five domains listed under psychotic disorders in DSM-5, making it broader than the definitions from 1999 ACR nomenclature for NPSLE (9) and 2019 EULAR/ACR Classification Criteria (10), which are limited to delusions and hallucinations. To employ more precise terminology, we adopted the DSM-5 framework to define psychosis (20). Our exclusion criteria incorporated those specified in the SLEDAI-2K, as well as additional exclusions from the 1999 ACR (9) and 2019 EULAR/ACR Classification Criteria (10).

#### 2.4. Examples present of SLEDAI-2K score

| How to determine. | Examples |
|---|---|
| - The diagnosis specified SLE flare with lupus psychosis<br>- The treatment plan (immunosuppression and antipsychotics) | Symptoms_Signs<br>Complaint: Acute psychosis (persecutory/referential delusions, incoherent speech,<br>Symptoms_Signs<br>irritability, impulsive behavior) triggered by stress.Known SLE with predominant cutaneous involvement. On HCQ only.<br><br>PE<br>Neuro: Alert, disorganized thought, paranoid delusions.<br>Skin: Alopecia.<br>Ext: Mild bilateral LL pitting edema.<br>MSK: No active synovitis.<br>CV/Resp: No rub/effusion.<br>Labs (Feb 2025): WBC 1.86↓, Hb 96↓, ESR 114↑, C3 56↓, C4 10↓; ANA +1:1000, Anti-Smith ++, Anti-dsDNA ++, ACL 46.3↑ RU/mL.<br>MRI Brain (Feb 2025): R ant frontal horn WM lesion (T1/T2↑, FLAIR rim↑) -<br>Paraclinical_tests<br>stable chronic encephalomalacia/gliosis.<br>EEG: Normal. LP normal.<br>Paraclinical_tests<br>Psych Scales: SAPS 71 (↑↑ positive Sx).<br><br>Impression<br>Active SLE flare w/ severe Neuropsychiatric<br>Psychosis**[criteria_fulfilled][within_10days][treat_escalated]**<br>(Psychosis) & Hematologic (Leukopenia, Anemia) involvement.<br><br>Plan<br>Immunosuppression:<br>Intention_to_treat<br>Steroids: Methylprednisolone 500mg IV daily x3d → taper (250mg x3d → 120mg x3d → 80mg daily).<br>Intention_to_treat<br>IV Cyclophosphamide<br>HCQ: Continue 0.2g BID.<br>Psych: Olanzapine ↑ to 15mg QHS, add Clonazepam 2mg QHS & Valproate SR 0.25g BID.<br>Prophylaxis: ASA 100mg daily (↑ACL). |

#### 2.5. Examples absent of SLEDAI-2K score

| Why is it an exception? | Examples |
|---|---|
| - Steriod-induced psychosis is the exclusion of lupus psychosis.<br>- The management plan focuses on steroid tapering and antipsychotic medication. | Symptoms_Signs<br>Patient Complaint: Acute manic episode (psychomotor excitement, pressured speech) + insomnia.<br><br>Prior Tx Response:<br>SLE Dx 20y ago; prednisolone 12.5mg qd ×5y (stable).Chronic insomnia: Triazolam 7.5mg or flunitrazepam 2mg qd ×15y. no prior psych Hx<br><br>History<br>Dec 30 2002: Cutaneous flare (↑ erythema) → prednisolone ↑ to 40mg qd →<br>History<br>After 1mo: Hypomania (hyperactivity, ↑ talkativeness)<br><br>PE<br>lupus erythema, no ulceration<br>Psychomotor agitation, no focal deficits.<br><br>Paraclinical<br>Paraclinical_tests<br>CT head/CSF: WNL (no structural/neuro infx).<br>labs: dsDNA16 , C3/C4 -ve, urine -ve<br><br>Impression<br>Exclusions –links to– Psychosis [criteria_unfulfilled_diagnostic][within_10days][treat_de_escalate]<br>steroid-induced psychosis (temporal link to ↑ prednisolone).<br><br>DDx:<br>SLE psychosis: Less likely (no renal/serositis/CNS involvement).<br>Primary mania: Excluded by acute onset post-steroid ↑.<br><br>Management Plan<br>Intention_to_treat<br>Steroid taper: Prednisolone ↓ from 30mg → goal ≤20mg (monitor skin/SLE activity).<br>Valproic acid ↑ to 800mg qd<br>Risperidone 4mg qd |

### 3. Organic Brain Syndrome

#### 3.1. Clinical definition of concepts

***Organic brain syndrome***, also known as ***organic brain disorder***, is an obsolete general term from psychiatry. Originally, the term was created to distinguish physical (termed "organic") causes of mental impairment from psychiatric (termed "functional") disorders. According to ACR definitions for NPSLE in 1999 (7), "organic brain syndrome" is not recommended for usage since it is not a defined entity in the DSM-IV (21). "***acute confusional state***" is more used ("acute" means "of recent onset"). This state is equivalent to "***delirium***" defined in DSM-IV and ICD-9 as an observable state of impaired consciousness, cognition (including perception), mood, affect, and behavior. Neurologists often use "***encephalopathy***" where psychiatrists use "***delirium***" to describe the same clinical state. In DSM-V, delirium was categorized into the ***neurocognitive disorders*** (NCD). The essential feature of a ***delirium*** is a disturbance of ***consciousness*** that is accompanied by a change in ***cognition*** that cannot be better accounted for by a preexisting or evolving dementia(DSM-IV). (21) The diagnostic criteria for both DSM-IV(21) and DSM-V(20) emphasize both disturbance in ***awareness*** and ***cognition***. The syndrome develops over a short period of time, usually hours to days, tends to fluctuate during the course of the day, and encompasses a spectrum from mild disturbances of consciousness to coma, including ***hypo-aroused***/***hypoactive*** (e.g., somnolence, stupor) and ***hyper-aroused***/***hyperactive*** (e.g., delirium tremens) states (9).

#### 3.2. Common ways to describe this concept

| | |
|---|---|
| Similar terms for Organic Brain syndrome | ***acute confusional state***<br>***delirium***<br>***encephalopathy***<br>***neurocognitive disorders (NCD)*** |
| diagnostic criteria of delirium from DSM-V(20) | A. A disturbance in ***attention*** (i.e., reduced ability to direct, focus, sustain, and shift attention) and ***awareness*** (reduced orientation to the environment).<br>B. The disturbance develops over a short period of time (usually hours to a few days), represents a change from baseline attention and awareness, and tends to fluctuate in severity during the course of a day.<br>C. An additional disturbance in ***cognition*** (e.g., memory deficit,disorientation, language, visuospatial ability, or perception).<br>D. The disturbances in Criteria A and C are not better explained by another preexisting, established, or evolving neurocognitive disorder and do not occur in the context of a severely reduced level of arousal, such as coma.<br>E. There is evidence from the history, physical examination, or laboratory findings that the disturbance is a direct physiological consequence of another medical condition, substance intoxication or withdrawal (i.e., due to a drug of abuse or to a medication), or exposure to a toxin, or is due to multiple etiologies.<br><br>*Specify* if:<br>***Acute*****:** Lasting a few hours or days.<br>***Persistent*****:** Lasting weeks or months.<br><br>*Specify* if:<br>***Hyperactive*****:** The individual has a hyperactive level of psychomotor activity that may be accompanied by mood lability, agitation, and/or refusal to cooperate with medical care.<br>***Hypoactive*****:** The individual has a hypoactive level of psychomotor activity that may be accompanied by sluggishness and lethargy that approaches stupor.<br>***Mixed level of activity:*** The individual has a normal level of psychomotor activity even though attention and awareness are disturbed. Also includes individuals whose activity level rapidly fluctuates. |
| diagnostic criteria of delirium from DSM-IV(21) | The disturbance in consciousness is manifested by a reduced clarity of ***awareness*** of the environment. The ability to focus, sustain, or shift attention is impaired (Criterion A).<br>There is an accompanying change in ***cognition*** (which may include memory impairment, disorientation, or language disturbance) or development of a ***perceptual disturbance*** (Criterion B).<br>The disturbance develops over a short period of time and tends to fluctuate during the course of the day (Criterion C). |

#### 3.3. SLEDAI-2K definition in this study

As shown in Table 8, we applied the definition of SLEDAI-2K and added some details according to DSM-V(20). The SLEDAI-2K definition of ***organic brain syndrome*** covers all the diagnostic criteria mentioned in the DSM definition: 1) disturbance in ***attention*** (sustained); 2) disturbance in ***cognition*** (impaired orientation, memory, or other intellectual function, including perceptual disturbance and incoherent speech); 3) rapid onset with fluctuating clinical features. Importantly, SLEDAI-2K requires at least two of the following for scoring this descriptor on top of: ***perceptual disturbance***, ***incoherent speech***, ***insomnia or daytime drowsiness***, or ***increased or decreased psychomotor activity***. Moreover, the SLEDAI-2K definition of active organic brain syndrome is stricter than that from ACR nomenclature and case definitions for NPSLE in 1999(9).

#### 3.4. Examples present of SLEDAI-2K score

| How to determine. | Examples |
|---|---|
| A clinical diagnosis of acute confusional state was made.<br>The management plan involves initiating immunosuppressive therapy. | Symptoms_Signs<br>Acute confusion, mood swings, pressured speech, agitation.<br>Symptoms_Signs<br>HPI:2-week history of flunctuating behavior (irritability, confusion), with disorganized thoughts. No prior psychiatric history.<br>Labs:<br>Hb 96 ↓, WBC 6.1, Plt 371, CRP 3.3<br>C3 0.40 g/L ↓ (0.9–1.8), C4 0.05 g/L ↓ (0.1–0.4), Anti-dsDNA 349 IU/mL ↑ (<35), anti-RNP >643 ↑ (<19.9), anti-Sm >693.5 ↑ (<19.9)<br>APLA/anti-B2GP neg, tox screen neg, QuantiFERON neg<br>Paraclinical_tests<br>Imaging: MRI brain (GA): nl<br>Paraclinical_tests<br>CSF: WBC 20/μL (100% lymph), protein 0.50 g/L, glucose 2.7 mmol/L (serum 5.1); cultures neg<br>Paraclinical_tests<br>EEG: nl, no epileptiform activity<br>Paraclinical_tests<br>Serology: Antineuronal Ab neg<br>Organic_brain_syndrome **[criteria_fulfilled][within_10days][treat_escalated]**<br>NPSLE acute confusional status<br>Management Plan<br>Risperidone 3mg/day OD (taper after CYC pulses)<br>Intention_to_treat<br>IV methylprednisolone 500 mg ×3 days → transition to oral pred 40 mg OD |

#### 3.5. Examples absent of SLEDAI-2K score

| Why is it an exception? | Examples |
|---|---|
| Cases identified as steroid-induced delirium were excluded from the diagnosis of lupus acute confusional state.<br>Therefore, the management plan focused on steroid tapering. | 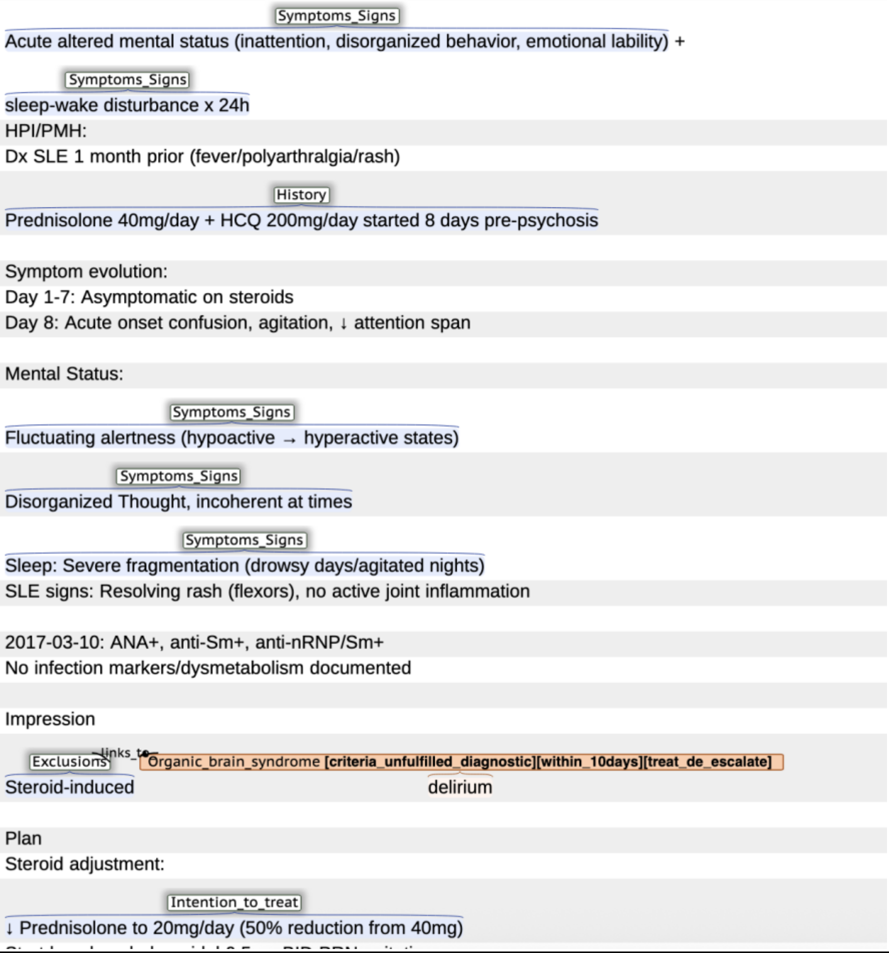<br>Symptoms_Signs<br>Acute altered mental status (inattention, disorganized behavior, emotional lability) +<br>Symptoms_Signs<br>sleep-wake disturbance x 24h<br>HPI/PMH:<br>Dx SLE 1 month prior (fever/polyarthralgia/rash)<br>History<br>Prednisolone 40mg/day + HCQ 200mg/day started 8 days pre-psychosis<br><br>Symptom evolution:<br>Day 1-7: Asymptomatic on steroids<br>Day 8: Acute onset confusion, agitation, ↓ attention span<br><br>Mental Status:<br>Symptoms_Signs<br>Fluctuating alertness (hypoactive → hyperactive states)<br>Symptoms_Signs<br>Disorganized Thought, incoherent at times<br>Symptoms_Signs<br>Sleep: Severe fragmentation (drowsy days/agitated nights)<br>SLE signs: Resolving rash (flexors), no active joint inflammation<br><br>2017-03-10: ANA+, anti-Sm+, anti-nRNP/Sm+<br>No infection markers/dysmetabolism documented<br><br>Impression<br>Exclusions —links_to→ Organic_brain_syndrome [criteria_unfulfilled_diagnostic][within_10days][treat_de_escalate]<br>Steroid-induced delirium<br><br>Plan<br>Steroid adjustment:<br>Intention_to_treat<br>↓ Prednisolone to 20mg/day (50% reduction from 40mg)<br>[illegible] |

### 4. Visual disturbance

#### 4.1. Clinical definition of concepts

The visual disturbance items of the SLEDAI-2k encompass three clinical entities: ***retinopathy***, ***choroidopathy***, and ***optic neuritis***.
The ***retina*** is the light-sensitive layer of tissue at the back of the eyeball. Images come through the eye's lens and are then focused on the retina. The retina converts these images to electric signals and sends them along the optic nerve to the brain. Lying behind the ***retina*** is the ***choroid***, which is also known as the choroidea or choroid coat, is the vascular layer of the eye. The ***retinal pigment epithelium (RPE)*** is the outermost layer of the retina and separates the neurosensory retina from the choroid. Retinal changes cause vision symptoms: changes in sharpness of vision, loss of color perception, flashes of light or floaters, distorted vision (straight lines look wavy).
***Optic neuritis (ON)*** is an optic nerve inflammation, including the ganglion cell axons, intradural space, or the optic nerve meningeal sheaths. Inflammation of the optic nerve posterior to the globe is often labelled as ***retrobulbar neuritis***. Etiologically, ON is divided into three basic groups, (1) Autoimmune ON, which is often relapsing and is associated with multiple sclerosis (MS), Aquaporin4 IgG antibodies-associated neuromyelitis

optica spectrum disorder (NMOSD) or Anti-myelin oligodendrocytes glycoprotein antibody-associated disease (MOGAD); (2) Infectious or systemic ON; and (3) idiopathic.(22, 23)

#### 4.2. Common ways to describe this concept

| | | |
|---|---|---|
| ***Retinopathy or choroidopathy*** | ***Retinal or Choroid hemorrhages*** | ***Retinal hemorrhages*** may occur on the surface of the retina (***preretinal***), under the retina (***subretinal***), or within the retinal tissue (***intraretinal***). Optical coherence tomography (OCT) can determine the location of the haemorrhages. (24) Unless the haemorrhages obscure the macula, patients may remain asymptomatic. |
| | ***Cotton-wool spots (CWSs)*** | ***Cotton-wool spots(CWSs),*** *previous* called the ***retinal soft exudates,*** are greyish-white, fudgy-bordered swellings of the retinal nerve fibre layer (axons of the retinal ganglion cells) resulting from an ischemic insult to the retinal nerve fibres which results in the blockade of the axoplasmic flow (22). The occurrence of CWSs is a sign of recent serious vascular damage (last for 4–6 weeks and disappear). The "principal constituent of the CWS" are ***cytoid bodies***. Histological examination showed ***cytoid bodies*** corresponded to areas in which the retinal nerve-fiber layer was swollen, revealing a central eosinophile globule.(25) |
| | ***Hard exudates*** | ***Hard exudates*** in the retina indicates a breakdown in the blood-retinal barrier with the leakage of plasma and macromolecules. ***Hard exudates*** have a waxy glistening appearance with sharp margins, and vary in size from a pinpoint to massive mounds measuring several disc diameters. Unlike cotton-wool spots, the retinal hard exudates often last for months or even years if the pathology that caused these continues to persist. (22) |
| ***Optic neuritis*** | ***Optic neuritis*** (ON) | ***Optic neuritis*** (ON) is an optic nerve inflammation, including the ganglion cell axons, intradural space, or the optic nerve meningeal sheaths. (22) |

#### 4.3. SLEDAI-2K definition in this study

As shown in Table 8, this study adopts the original SLEDAI-2K definition for retinopathy and choroidopathy, while utilizing supplementary diagnostic criteria for optic neuritis from two sources: Ophthalmic Signs in Practice of Medicine (22) and Diagnosis and Classification of Optic Neuritis (23). We listed autoimmune optic neuritis as an additional exclusion criterion.

SLE ***retinopathy*** is characterized by bilateral multiple cotton wool spots resulting from occlusion of the precapillary arterioles and manifests as SLE microangiopathy. SLE may also uncommonly present with choroidopathy, mostly bilateral, and presents with multifocal serous detachments of the retina.(26) There is no consensus regarding the precise diagnostic criteria until 2022 (23). The criteria of definite optic neuritis given by the consensus are very strict; therefore, we allow for possible optic neuritis to be considered as optic neuritis based on the clinical notes.

#### 4.4. Examples present of SLEDAI-2K score

| How to determine. | Examples |
|---|---|

| | |
|---|---|
| A clinical diagnosis of retinopathy was made, with cotton-wool spots identified as a typical manifestation in SLE.<br>The management plan included immunosuppressive therapy. | Ocular OD:<br>Paraclinical_tests<br>Temporal disc pallor + blurred nasal margin<br>Paraclinical_tests<br>Widespread cotton wool spots (5,8,10,11 o'clock)<br>Systemic:<br>Polyarticular tenderness (2+/4), mild edema<br>No rash/serositis/renal edema<br><br>Labs:<br>ESR↑, ANA+, dsDNA+, C3/C4↓<br>Renal: UP 3+, UTP 2.4g/24h<br>Ophthalmic:<br>Paraclinical_tests<br>FFA OD: Disc pallor, cotton wool spots<br>OCT OD: Normal macula<br>SLEDAI: 23 (very severe)<br><br>Impression<br>Active SLE flare: New-onset lupus<br>Visual_disturbance[criteria_fulfilled][within_10days][treat_escalated]<br>retinopathy (cotton wool spots + disc pallor) + lupus nephritis (proteinuria) + polyarthritis.<br><br>Management Plan<br>Induction:<br>Intention_to_treat<br>IV methylprednisolone 1g ×3d → oral prednisolone 1mg/kg/day |

| | |
|---|---|
| A diagnosis of optic neuritis was made.<br>The clinical presentation included bilateral vision loss and a relative afferent pupillary defect (RAPD).<br>The management plan involved immunosuppressive therapy. | Symptoms_Signs<br>Pt c/o acute bilateral vision loss (CF both eyes) ×2 days. Developed malar rash + polyarthritis (3 joints) during workup. Rapid progression to oliguric renal failure + thrombocytopenia within days.<br><br>PE<br>Neuro-ophthalmic:<br>Bilateral RAPD, non-reactive pupils<br>Symptoms_Signs<br>Optic disc hyperemia + blurred margins OU<br>Dermatologic: Malar rash<br>Musculoskeletal: Active synovitis (3 peripheral joints)<br><br>Labs:<br>Hb 8.8↓ g/dL, Plt 21.9↓×10$^3$/mm$^3$, ESR 56↑ mm/hr, CRP 9.5↑ mg/dL<br>Renal: BUN 72↑ mg/dL, Cr 2.8↑ mg/dL, UP 2.4 g/dl<br>Serology: ANA+, dsDNA+, LE cell+, lupus anticoagulant+ (initially), C3/C4↓<br>ant-NMO -ve. VEP equivocal<br><br>Imaging:<br>Paraclinical_tests<br>FFA: Bilateral disc leakage, retinal non-perfusion, venous tortuosity<br>Brain MRI: No demyelination (MS excluded)<br>Electrophysio: VEP ↑latency ↓amplitude OU<br><br>Impression<br>Active SLE flare: New-onset bilateral<br>Visual_disturbance[criteria_fulfilled][within_10days][treat_escalated]<br>optic neuritis + CRVO, lupus nephritis (class IV biopsy), hematologic crisis (thrombocytopenia/anemia), malar rash, polyarthritis.<br><br>Management Plan<br>Intention_to_treat<br>Pulse therapy: IV methylprednisolone 1g ×3d → prednisone 1mg/kg/day |

#### 4.5. Examples absent of SLEDAI-2K score

| Why is it an exception? | Examples |
|---|---|

| | |
|---|---|
| The blurring of vision was likely attributed to a hydroxychloroquine side effect. | 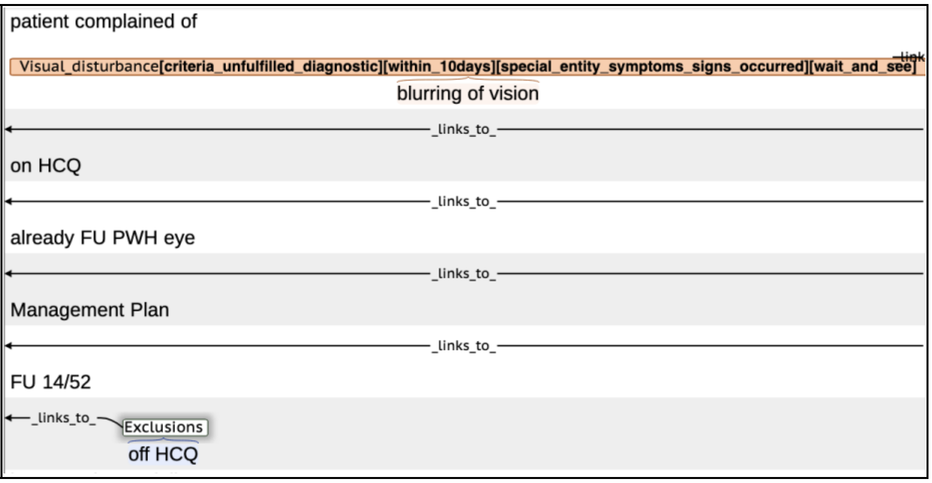 |

### 5. Cranial nerve disorder

#### 5.1. Clinical definition of concepts

***Cranial nerve disorders*** (***Cranial neuropathies***) are characterized by a pathological disruption of the structure or function of one or more cranial nerve pairs, producing sensory, motor, or autonomic deficits. The etiology is heterogeneous, encompassing inflammatory, infectious, vascular, traumatic, tumoral, and neurodegenerative processes. The specific clinical presentation is determined by the identity of the affected nerve(s) and the location of the lesion along its anatomical course.

#### 5.2. Common ways to describe this concept

Table. Summary of Cranial Nerves: Function, Dysfunction, and CNS Coverage. (27)

| Number | Name | Primary Function | Main Dysfunctions<br>(S: sensory; M: motor) |
|---|---|---|---|
| **CN I** | **Olfactory** | Sensory (Smell) | Anosmia (loss of smell), parosmia (distorted smell) |
| **CN III** | **Oculomotor** | Motor (Eye movement, pupil constriction, eyelid elevation) | Ptosis, diplopia, "down and out" eye deviation, mydriasis (pupil dilation) |
| **CN IV** | **Trochlear** | Motor (Eye movement - intorsion, downward gaze) | Diplopia (worse when looking down and in, e.g., going down stairs), head tilt |
| **CN V** | **Trigeminal** | **Both**<br>S: Facial sensation<br>M: Mastication | **S:** Loss of facial sensation, trigeminal neuralgia<br>**M:** Jaw deviation, difficulty chewing |
| **CN VI** | **Abducens** | Motor (Eye movement - abduction) | Failure of lateral gaze (esotropia), diplopia |
| **CN VII** | **Facial** | **Both**<br>M: Facial expression<br>S: Taste (ant. 2/3 tongue) | **M:** Facial palsy (Bell's palsy), loss of facial expression<br>**S:** Loss of taste, hyperacusis |
| **CN VIII** | **Vesti-bulocochlear** | Sensory (Hearing and balance) | Sensorineural hearing loss, tinnitus, vertigo, nystagmus |
| **CN IX** | **Glosso-pharyngeal** | **Both**<br>S: Taste (post. 1/3 tongue), pharynx<br>M: Swallowing | **S:** Loss of gag reflex, loss of taste<br>**M:** Dysphagia (swallowing difficulty) |

| | | | |
|---|---|---|---|
| **CN X** | **Vagus** | **Both**<br>M: Palate, pharynx, larynx, parasympathetic (viscera)<br>S: Viscera, pharynx, larynx | Hoarseness, dysphagia, dysarthria, loss of gag reflex, autonomic dysfunction |
| **CN XI** | **Spinal Accessory** | Motor (SCM & Trapezius muscles) | Weakness turning head, shoulder droop, difficulty shrugging |
| **CN XII** | **Hypoglossal** | Motor (Tongue movement) | Tongue weakness, atrophy, fasciculations; deviation **toward** the side of the lesion |

#### 5.3. SLEDAI-2K definition in this study

As shown in Table 8, we refined the SLEDAI-2K lupus definition by integrating criteria from the ACR nomenclature and case definitions for NPSLE in 1999 (9). Of note, according to SLEDAI-2K Optic neuritis (CN II) involvement was a disease entity under visual disturbance, so we will not include this in cranial nerve disorder.

#### 5.4. Examples present of SLEDAI-2K score

| How to determine. | Examples |
|---|---|
| In a case of SLE-related CN III palsy, the patient presented with ptosis and diplopia confirmed by MRI.<br>Laboratory findings included serologic activity, leukopenia, renal involvement, supporting active lupus.<br>Management involved initiating MP treatment for NPSLE. | Symptoms_Signs<br>Acute R eye ptosis + diplopia + ocular pain 1 day.<br>Chronic progressive LL weakness six months ago → immune-mediated neuropathy? → Failed 2 IVIG courses for presumed immune neuropathy (nerve biopsy: severe axonal neuropathy)<br>PE<br>Symptoms_Signs<br>Neuro: Complete R CN III palsy (↓ & lateral gaze, ptosis, mydriasis 5mm, nonreactive pupil)<br>Motor: Severe LL weakness → wheelchair-dependent<br>No rash/arthritis/serositis<br>ANA+, dsDNA+, C3↓ C4↓, Coombs+, UPCR 135.2↑ (N<30), NAG 25.9↑ (N<15), β2M 567↑ (N<230)<br>CBC: Leukopenia + lymphopenia<br>Imaging:<br>Brain MRI<br>Paraclinical_tests<br>a contrast effect at the right oculomotor nerve in the prepontine cistern → inflammatory neuropathy<br>Neurophysiology: Absent peroneal/tibial/sural responses (diffuse axonal neuropathy)<br>Active SLE flare:<br>Cranial_nerve_disorder [criteria_fulfilled][within_10days][treat_escalated]<br>Neuropsychiatric (CN III palsy + radiculopathy)+ Renal + Hematologic (Coombs+ hemolysis/cytopenias)<br>Management Plan<br>Intention_to_treat<br>IV methylprednisolone 1g x3d → repeat x2 cycles (q1wk)<br>Prednisone 30mg daily post-pulse<br>HCQ 400mg OD (baseline retinal screening) |

#### 5.5. Examples absent of SLEDAI-2K score

| Why is it an exception? | Examples |
|---|---|
| Trigeminal neuropathy (cranial nerve V) caused by viral infection is an exclusion. | Symptoms_Signs<br>Acute-onset numbness in R lower lip/chin (V3 distribution).<br><br>Exclusions<br>Preceded by recurrent intraoral herpes lesions 2wks prior. No trauma/dental procedures. PMH: Recurrent herpes labialis.<br><br>Prior no history for neuropathy.<br>Neuro: Pinprick/cotton-touch hypoesthesia isolated to R mental nerve distribution. No motor weakness.<br><br>Exclusions<br>notice Healed herpes lesions (no active ulcers).<br><br>Labs: HSV IgG ↑(1/4,000 U↑), IgM-. ANA-, dsDNA-.<br>Imaging: Panoramic XR, craniofacial CT nl.<br>Impression:<br>Cranial_nerve_disorder **[criteria_unfulfilled_diagnostic][within_10days][wait_and_see]**<br>Acute Idiopathic Trigeminal Neuropathy (V3):<br>Exclusions<br>Post-herpetic sensory neuropathy.<br><br>Management Plan:<br>Intention_to_treat<br>Antiviral: Acyclovir 1000mg PO daily ×10d.<br>Monitoring: Sensory exam weekly.<br>F/U: 2wks. Expected resolution: 1-3mo (per prior episodes). |

| | |
|---|---|
| CN VI palsy caused by fungal menigitis. | BP nl , HR nl , T 39°C↑ , RR nl , $O_2$ sat -<br><br>Admitted 3d ago w/ horizontal diplopia & mild HA → today spiked fever (39°C), severe HA, N/V.<br>PMH: Class IV LN (dx 1994, 13x IV CYC pulses), on DFZ 24mg & MMF 1g daily.<br><br>Symptoms_Signs<br>PE:Alert, no meningismus , R sixth nerve palsy (esotropia) , Neuro: CN II-XII intact except VI<br>No rash/vasculitis, no jt pain<br><br>Labs: WBC 10.35k↑ , Plt 123k↓ , Cr 2.66↑ , C3 77.7↓ , CRP↑ (no value)<br>Exclusions<br>CSF: OP 42↑↑ , WBC 280↑ , Crypto Ag+ , India ink+ , Culture: C. neoformans<br>Initial CT/MRI brain nl<br>Repeat MRI (Day4): accentuation of leptomeningeal<br>contrast enhancement along the sulci of both cerebral hemisphere in T1-weighted image, suggestive of<br>Exclusions<br>meningitis.<br><br>Exclusions Cranial_nerve_disorder [criteria_unfulfilled_diagnostic][within_10days][treat_de_escalate]<br>Cryptococcal meningitis → R CN VI palsy,<br>CKD<br><br>Plan:<br>Intention_to_treat<br>• Discontinue MMF , Maintain DFZ 24mg •Antifungals: Amphotericin B + flucytosine IV (started) → switch to |

#### 6. Lupus headache

##### 6.1. Clinical definition of concepts

According to the 3rd edition of the International Classification of Headache Disorders (ICHD-3) (28), ***headaches*** are largely divided into ***primary headaches*** and ***secondary headaches***. Primary headaches are divided into migraine, tension-type headaches, trigeminal autonomic cephalalgias, and other primary headache disorders. Lupus-related headache is put into the secondary headaches categories, including 7.3.3 Headache attributed to other non-infectious inflammatory intracranial disease. (It remits after successful treatment of the autoimmune disorder.) and 10.8.2 Headache attributed to other metabolic or systemic disorder. Headache as a non-specific symptom is commonly seen in SLE with a prevalence of 57.1%, which does not differ from the prevalence observed in the population without the disease. (29) Lupus headache was reported with a prevalence of only 1.5% among SLE. (30)

##### 6.2. Common ways to describe this concept

| | |
|---|---|
| **Headache** | The common, general term for any pain located in the head. It describes the symptom itself. |
| **Cephalalgia/Cephalalgias (pl.)** | The formal medical word for "headache." It is the umbrella term used in medical classification for all headache disorders. |
| **Migraine** | A specific, common neurological disorder that causes severe, often one-sided, throbbing headaches, usually accompanied by nausea, and sensitivity to light and sound. |

| | |
|---|---|
| narcotic analgesics<br>(Opioids or opiates, opioid analgesics, or narcotics) | Narcotic analgesics are a class of medicines that are used to provide relief from moderate-to-severe acute or chronic pain. Natural/Semi-synthetic: Morphine, Codeine, Oxycodone (OxyContin), Hydrocodone (Vicodin).<br>Synthetic: Fentanyl, Methadone, Demerol. |

#### 6.3. SLEDAI-2K definition in this study

As shown in Table 8, we refined the SLEDAI-2K ***lupus headache*** definition by integrating criteria from two sources: the British Isles Lupus Assessment Group 2004 (BILAG 2004) index (31) and the 1999 American College of Rheumatology (ACR) nomenclature for neuropsychiatric SLE (NPSLE) (9).

The concept of "***lupus headache***" was first introduced by the Systemic Lupus Erythematosus Disease Activity Index 2000 (SLEDAI-2K), which described it as "intense, persistent headache that may be migrainous and is non-responsive to narcotic analgesia." However, researchers have questioned this vague concept for years, noting that terms such as "persistent" and "non-responsive" are subjective. Furthermore, opioids are not standard therapy for headaches. (32, 33). To address these issues and provide clear, measurable parameters for "severe" and "persistent," we applied the BILAG 2004 definition, which characterizes lupus headache as a disabling headache unresponsive to narcotic analgesia and lasting three days or longer (≥3 days) (31). To further clarify the disease entities comprising lupus headache, we used the classification from the 1999 American College of Rheumatology (ACR) nomenclature and case definitions for NPSLE (9). It classifies lupus headache into the following four entities: 1) ***migraine***; 2) ***tension-type headache***; 3) ***cluster headache***; and 4) ***headache secondary to intracranial hypertension (including pseudotumor cerebri)***.

Differential diagnosis is paramount when assessing lupus headache, as severe, non-responsive headaches in SLE patients can signal serious underlying pathology (e.g., seizures, cerebral edema, ischemia, or severe hypertension) and should not be treated in isolation. (33) Key secondary headaches to exclude include cerebral vasculitis, cerebral venous thrombosis, stroke, CNS infection, aseptic meningitis, intracranial hemorrhage, posterior reversible encephalopathy syndrome (PRES), reversible cerebral vasoconstriction syndrome (RCVS), intracranial neoplasms, and intracranial hypotension. (34)

#### 6.4. Examples present of SLEDAI-2K score

| How to determine. | Examples |
|---|---|

| | |
|---|---|
| In a case of SLE-related intracranial hypertension, the patient presented with a severe, persistent headache and vomiting lasting >3 days.<br>Diagnostic lumbar puncture showed an opening pressure exceeding 50 cm $H_2O$.<br>Laboratory findings included serologic activity, leukopenia, and anemia, supporting active lupus. Management involved initiating immunosuppressant treatment for NPSLE. | BP 118/76, HR 82, T 37.1°C, RR 16<br><br>Symptoms_Signs<br>Acute severe headache → vomiting x4d.<br><br>PMH: SLE dx 1y ago (rash/arthritis). Prior tx: HCQ 200mg BID<br><br>PE<br>Severe bilateral papilledema<br>CN II-XII intact, No synovitis/rash<br><br>Paraclinical_tests<br>LP: OP >50 cmH2O↑↑<br><br>WBC 2.8↓, Hb 9.2↓, Plt 210<br>dsDNA 285↑, C3 0.45↓, C4 <0.10↓,aCL IgM/IgG-, LA-<br>Prot 2+, No casts<br><br>Impression:<br>Active SLE with<br>Lupus_headache[criteria_fulfilled][within_10days][treat_escalated][special_entity_symptoms_signs_occurred]<br>Intracranial hypertension<br>Leukopenia, anemia<br><br>Plan:<br>Intention_to_treat<br>Methylprednisolone 1g IV x3d → Pred 60mg daily<br>Repeat LP in 72h if OP unchanged |

#### 6.5. Examples absent of SLEDAI-2K score

| Why is it an exception? | Examples |
|---|---|
| Headache is attributed to high blood pressure. | Time [within_10days]<br>attended AED yesterday for<br>Lupus_headache[criteria_unfulfilled_diagnostic][within_10days][special_entity_symptoms_signs_occurred]<br>headache<br>Exclusions<br>and HT, 150/100<br>was used to be around 120/80<br>Exclusions<br>noted BP increased for last 10 days |

| | |
|---|---|
| SLE is stable. The migraine is not disabling and does not require escalation to opioids or immunosuppressants. | Note<br>BP 105/64 P 57 UP- UK- UG- URBC trace BW 57kg<br>CBP/RFT and serology all stable<br>MRI brain normal<br>No psychotic disorder<br>Still with<br>Lupus_headache[criteria_unfulfilled_diagnostic][within_10days][special_entity_symptoms_signs_occurred][wait_and_see]<br>migraine<br>attacks, PYNEH neurology appt in 8/2005!<br>Intention_to_treat<br>Claimed that B blocker did not help her Sx and Ponstan not useful<br>Management Plan<br>CBP, RFT, antidsDNA, C3/4 today<br>Intention_to_treat<br>Try pizotifen for migraine prophylaxis |

### 7. CVA

#### 7.1. Clinical definition of concepts

The term ***cerebrovascular accidents*** (CVA) is an old but now discouraged medical term for ***stroke*** (35). ***Cerebrovascular events*** are a more widely used term. The World Health Organization defines stroke as the sudden onset of focal neurological signs of presumed vascular origin lasting longer than 24 hours or causing death. In the past, any focal cerebral ischemic event with symptoms like those of stroke lasting <24 hours were defined as a ***transient ischemic attack*** (TIA). The definition of TIA is evolving, and the statement supported endorsement of the following, tissue-based definition of TIA: a transient episode of neurological dysfunction caused by focal brain, spinal cord, or retinal ischemia, without acute infarction (36). A generalized definition of cerebrovascular accidents (cardiovascular events) should include TIA and stroke (37, 38). Stroke contains many clinical entities, depending on the site of the lesion of a blood vessel in the brain and the causes of the disease. An ideal classification of stroke first distinguishes between ischemic and hemorrhagic stroke, subarachnoid hemorrhage, cerebral venous thrombosis, and spinal cord stroke. Then the classification should identify the most likely etiology(ies) (i.e. atherothrombotic, cardioembolic, small vessel disease, and other causes). However, 25–39% strokes have failed to identify a definite cause.

#### 7.2. Common ways to describe this concept

Currently, there are many stroke subtype classifications according to causes, including Stroke Data Bank, Lausanne Stroke Registry, TOAST, and GÉNIC, which use different terminology for similar etiology. A more commonly used classification specifically for infarct stroke is OCSP classification, which is simple to apply and is of prognostic use. (39). The most common terminology that occurs in a clinical setting follows the basic structure of the classification mentioned above. Terminology for describing stroke subtypes, different causes and types of infarct subtypes are listed below.

Terminology for describing stroke.

| **Stroke subtypes** | **Stroke related etiology (from Stroke Data Bank, Lausanne Stroke Registry, TOAST and GÉNIC)** | **Type of infarct subtypes (OCSP classification)** |
|---|---|---|
| ***Ischemic stroke (acute ischemic stroke, AIS)*** | Atherothrombosis | Cerebral infarction |

| | | |
|---|---|---|
| ***Hemorrhagic stroke*** | Cardiac embolism/Cardioembolism/ Emboligenic heart disease/ Cardioembolic stroke | Lacunar infarct (LACI) or lacunar stroke (LACS) |
| ***Subarachnoid hemorrhage (SAH)*** | Small vessel occlusion /lacune/ Hypertensive arteriolopathy/ Lacunar stroke | Total anterior circulation infarct (TACI) or total anterior circulation stroke (TACS) |
| ***Cerebral venous thrombosis (CVT)*** | Other causes:<br><br>- arteritis, dissection, fibromuscular hyperplasia, sickle cell anemia.<br><br>- hematologic conditions (polycythemia, thrombocythemia, etc.)<br><br>- stroke in the setting of migraine or mycotic aneurysm | Partial anterior circulation infarct (PACI) or partial anterior circulation stroke (PACS) |
| ***Spinal cord stroke*** | | Posterior circulation infarcts (POCI) or posterior circulation stroke (POCS) |

#### 7.3. SLEDAI-2K definition in this study

As shown in Table 8, this study applied the definition and exclusions provided by ACR nomenclature and case definitions for NPSLE in 1999 (9). The definition of CVA in SLEDAI-2K didn't describe detailed clinical entities, so it is not clear whether TIA is counted. According to ACR nomenclature and case definitions for NPSLE in 1999 (9), common disease entities of lupus CVA include: 1) ***Stroke syndrome***; 2) ***Transient ischemic attack (TIA***); 3) ***Subarachnoid and intracranial hemorrhage,*** and 4) ***Sinus thrombosis***.
Although lupus erythematosus is a rare cause of CVA, lupus has an increased risk (relative risk 2.5) for stroke compared with the general population, particularly in those aged below 50 years (relative risk > 4.3), which cannot be explained by traditional cardiovascular risk factors (i.e., arteriosclerosis) (40, 41). Anticardiolipin antibody (aCL), lupus anticoagulant (LA), and cutaneous vasculitis are all risk factors for ischemic cerebrovascular disease in lupus, supporting pathological roles of lupus-related hypercoagulable state (the presence of antiphospholipid antibodies (aPLs)) and proinflammatory cytokine-mediated vasculitis in the disease process of CVA (42). Among cerebrovascular diseases in SLE, ischemic stroke and/or TIA comprise >80% of cases, intracerebral hemorrhage in 7–12%, subarachnoid hemorrhage in 3–5%, and sinus thrombosis in 2% (43). Paraclinical tests can be considered for differential diagnosis include aPLs, computed tomography (CT) and magnetic resonance imaging (MRI), carotid and transcranial Doppler ultrasound (CD/TCD), magnetic resonance angiography (MRA), digital subtraction angiography (DSA), echocardiography, and Holter studies.

#### 7.4. Examples present of SLEDAI-2K score

| How to determine. | Examples |
|---|---|

| | |
|---|---|
| The patient presented with an acute stroke, characterized by right hemiparesis and right hemisensory loss, with imaging confirming infarcts and vasculitis.<br>The management plan involved increasing steroids and immunosuppressants. | Symptoms_Signs<br>Acute right-sided weakness and numbness (onset 2 days prior).<br><br>Dx SLE 2 months ago (c/b lupus nephritis: severe proteinuria, edema).<br><br>Stable on prednisolone 20mg + mycophenolate 2g daily with partial renal symptom improvement.<br>Symptoms_Signs<br>Neuro: R hemiparesis (MRC 4/5), R hemisensory loss. No meningism.<br>Skin/msk: No active rash/joint swelling.<br>Cardiopulm: No rubs/murmurs (TTE normal).<br><br>Hb 9.3↓ \| Alb 2.1↓ \| LDL 333↑ \| ESR 126↑ \| CRP 0.95↑<br>UPC 415↑ \| dsDNA+ \| ANA+ \| Anti-SSA+<br>Negative: aPL, LA, anti-β2GP1, RF<br><br>Paraclinical_tests<br>Brain MRI DWI: Acute multifocal infarcts (L watershed zones):<br><br>Paraclinical_tests<br>Vessel wall MRI/MRA: Concentric wall thickening + enhancement from petrous to cervical ICA.<br>Paraclinical_tests<br>Stenosis: distal ICA, proximal PCA/ACA.<br>US: CDUS/EEG/TTE Normal.<br><br>impression:<br>CVA [criteria_fulfilled][within_10days][treat_escalated]<br>acute storke ( Large-vessel CNS vasculitis)<br>Lupus nephritis<br><br>Plan:<br>Intention_to_treat<br>↑prednisolone 40mg daily + continue mycophenolate 2g BID |

#### 7.5. Examples absent of SLEDAI-2K score

| Why is it an exception? | Examples |
|---|---|

| | |
|---|---|
| Stroke event with hemiplegia and imaging suggestive of local vascular stenosis.<br>The physician's judgment is therapy for a thrombotic/embolic event and immunomodulation for the lupus nephritis.<br>Consequently, this stroke is most likely due to coexisting conditions like atherosclerosis, rather than a manifestation of current lupus disease activity. | BP 110/64 P 98<br>UP 3+ URBC -ve<br>come on wheelchair<br>Cr 56 GFR >90<br>Hb 11.3 WBC 3.25 plt 135<br>serology not active<br>CRP 1.44<br>Time [11to30days]<br>admitted x CVA - x Lt<br>CVA[criteria_unfulfilled_diagnostic][11to30days][special_entity_symptoms_signs_occurred][wait_and_see]<br>hemiplegia<br>Paraclinical_tests<br>-CT brain showed Rt MCA infarct<br>Paraclinical_tests<br>-MRA : Rt MCA M2 stenosis<br>Intention_to_treat<br>-started Plavix for secondary stroke preventaion<br>-also noted proteinuria<br>-step up steroid 20mg daily --> 15mg daily and added aza as steroid sparing agent<br>-urine Pr/Cr ratio 2.22 ( ) --> 0.4 ( )<br>-noted Vit B12 def, anti IF <2<br>no ankle edema<br>no other complaint<br>Management Plan<br>FU 4/52<br>decrease prednisolone 12.5mg daily<br>check anti cardiolipin/ Vit B12, folate<br>check 24 hour urine protein |

### 8. Vasculitis

#### 8.1. Clinical definition of concepts

***Vasculitis*** is inflammation of vessel walls, characterized by infiltration of inflammatory cells and subsequent necrosis of vessel walls, leading to hemorrhage, ischemia, and infarction. (44) The diagnosis of vasculitis is a challenge because its clinical presentations are heterogeneous in severity and organ distribution. Currently, there are two mainstream classification system for vasculitis:1) Chapel Hill Consensus Conference (CHCC) based on pathological criteria (45) and 2) American College of Rheumatology (ACR) based predominantly on clinical findings (46). Based on histology, vasculitis can be classified on the size of vessels affected and the main immune cell mediating the inflammation (e.g. neutrophilic, granulomatous, or lymphocytic). Lupus vasculitis are mixed, predominately small and medium vessel vasculitis and can affect numerous organs. Vasculitic lesions in SLE can be largely divided into cutaneous and visceral vasculitis. The most frequent type of lupus vasculitis is cutaneous vasculitis.(47) Clinically, cutaneous vasculitis can present with morphologies that include urticaria, purpura, haemorrhagic vesicles, ulcers, nodules, livedo, infarcts, and/or digital gangrene. (48)

#### 8.2. Common ways to describe this concept

| | |
|---|---|
| ***ulceration/ulcer*** | An ulcer is a sore on the skin or a mucous membrane, accompanied by the disintegration of tissue. |

| | |
|---|---|
| ***gangrene*** | Skin gangrene is a type of tissue death caused by occlusion of skin vessels. Gangrene is a specific forms of necrosis. The tem ***necrosis*** is also considered to describe ischemic manifestations of vasculitis.(49) |
| ***lender finger nodules*** | Nodules correspond to palpable and solid lesions measuring of more than 1 cm. In the context of vasculitis, they reflect the involvement of vessels of the dermohypodermic junction or the hypodermis. Nodules can be located alongside an artery or a superficial vein, and can evolve into an ulcer or necrosis. (49) |
| ***periungual infarction*** | Injury or death of tissue resulting from inadequate blood supply that appear around the finger and toenails. |
| ***splinter hemorrhages*** | Splinter hemorrhages, small areas of hemorrhages under the fingernails or toenails, are rare in vasculitis and are more likely to be due to thromboembolic or thrombotic pathology. (49) |

#### 8.3. SLEDAI-2K definition in this study

As shown in Table 8, this study applied the original definition of ***vasculitis*** from SLEDAI-2K. Exclusions considered were from Jayne's work (50). In the SLEDAI-2K, the vasculitis items only refer to cutaneous vasculitis. Ideally, histologic and/or arteriographic confirmation should be used to determine the diagnosis of vasculitis; SLEDAI-2k allows for judgment depending on cutaneous lesions.

#### 8.4. Examples present of SLEDAI-2K score

| How to determine. | Examples |
|---|---|
| Diagnostic keyword for vasculitis occurred. | Impression:<br>Vasculitis [criteria_fulfilled][treat_escalated][within_10days]<br>New Dx SLE with Vasculitic Gangrene |
| Diagnostic keyword occcured. management plan was increasing steriods. | Vasculitis[criteria_fulfilled][within_10days][treat_escalated] Symptoms_Signs<br>vasculitic rash over ant chest and fingers<br>also warty growth over finger tips<br>no joint pain<br>imp: mild skin flare<br>Intention_to_treat<br>--> to step up Pred |

#### 8.5. Examples absent of SLEDAI-2K score

| Why is it an exception? | Examples |
|---|---|
| The leg ulcers might indicated a vasculitic rash. Given it was healed up, the clinicians tapered steroids. Consequently, no score. | FU podiatry for the<br>Vasculitis[criteria_unfulfilled_diagnostic][within_10days][special_entity_symptoms_signs_occurred][treat_de_escalate] links<br>leg ulcers,<br>_links_to_ Symptoms_Signs<br>left already healed up, right side still on self daily dressing<br>Management Plan<br>CBP, L/RFT, antidsDNA, C3/4, LDH today<br>Resume quinine 200mg qd prn<br>Intention_to_treat<br>Decrease prednisolone 7.5mg qd |

| | |
|---|---|
| The vasculitic rash on the hands remained stable. Given this stability, the clinicians did not initiate or escalate steroids or immunosuppressants. Consequently, no score. | BP 110/66 P 62 |
| | BW 55kg |
| | UP -ve URBC -ve |
| | Cr 59 |
| | serology anti dsDNA 51.6 |
| | Lipids: TC 5.8 LDL 3.5 |
| | A1c 6.3% |
| | advised lifestyle modifications |
| | Vasculitis [criteria_unfulfilled_diagnostic][within_10days][wait_and_see] |
| | Bilateral hands vasculitic rash static |
| | Management Plan |
| | FU 12/52 |
| | repeat Fasting blds bnv |

### 9. Arthritis

#### 9.1. Clinical definition of concepts

***Arthritis*** can largely be divided into inflammatory arthritis and noninflammatory arthritis conditions. Moreover, based on the number affected, it can also be divided into ***pauciarticular*** arthritis and ***polyarthritis*** (***polyarticular arthritis***). (51) ***Inflammatory arthritis*** is one of the most common manifestations of SLE. Although any joint may be affected, typical lupus arthritis is a ***symmetric polyarthritis*** that preferentially involves the small joints over the large joints. Swelling of the joint as a consequence of joint fluid or synovial proliferation can be present. Other signs and symptoms of lupus arthritis include joint erythema, pain with range of motion of the joint, and morning stiffness. The most common joints affected are the hand joints as well as the knees. Shoulders, ankles, elbows, and even sacroiliitis can also be involved. (52)

#### 9.2. Common ways to describe this concept

Table. Terminology and definitions for arthritis (51).

| | |
|---|---|
| ***Synovitis (synovial proliferation)*** | Inflammation of the synovial membrane, indicating active inflammatory arthritis |
| ***Inflammatory Arthritis*** | The presence of joint swelling, erythema, prolonged morning stiffness (more than one hour), and symmetric pain even at rest is suggestive of inflammatory conditions. |
| ***Noninflammatory arthritis conditions*** | weight bearing and movement worsens the pain, such as osteoarthritis |
| Classified based on number of joints affected | ***Pauciarticular arthritis***: Two to four joints are affected |
| | ***Polyarthritis (polyarticular arthritis)***: five or more joints are affected |
| Abbreviations for hand and toe joints | metacarpal phalangeal (MCP)<br>proximal interphalangeal (PIP)<br>distal interphalangeal (DIP)<br>metatarso Phalangeal (MTP)<br>interphalangeal (IP, big toe only has one joint) |

#### 9.3. SLEDAI-2K definition in this study

To augment the SLEDAI-2K definition, as shown in Table 8, the this study used supplementary criteria derived from SLICC 2012 (11). Exclusions to consider were derived from Grossman's work (52). The SLEDAI-2K incorporates the ACR 1997 arthritis definition. The SLICC group later expanded this definition to include "tenderness in two or more joints and at least 30 minutes of morning stiffness," a refinement that the subsequent EULAR/ACR classification criteria project validated as superior.

Paraclinical tests for evaluating arthritis include assessments of inflammatory markers, serum uric acid, synovial fluid, and specific pathogen serology, as well as imaging (51). Inflammatory markers such as ESR and CRP are typically higher in inflammatory arthritis than in non-inflammatory etiologies. Serum uric acid measurement is relevant for gout, although levels may be elevated even during intercritical periods. Synovial fluid analysis is crucial for diagnosing crystal-induced synovitis (e.g., gout, pseudogout) and septic arthritis. Serologic testing for pathogens like Borrelia burgdorferi, hepatitis B, hepatitis C, and parvovirus can identify infection-related arthritis. While imaging is widely used, its findings in conditions like lupus arthritis are often non-specific.

#### 9.4. Examples present of LEDAI-2K score

| How to determine. | Examples |
| --- | --- |
| ≥2 PIP joints with pain and signs of inflammation (swelling) | Symptoms_Signs<br>complained of Lt 2nd and 5th PIP<br>Arthritis[criteria_fulfilled][within_10days][special_entity_symptoms_signs_occurred]<br>joint pain and<br>Symptoms_Signs<br>swelling |
| ≥ 2 joints with pain and signs of inflammation (swelling) | Symptoms_Signs<br>Still increased temp ; mild swelling @ R hand / R elbow<br>Arthritis [criteria_fulfilled][within_10days][special_entity_symptoms_signs_occurred]<br>Some pain over various joints;<br>Symptoms_Signs<br>morning stiffness + |
| Synovitis is an indicator for active inflammatory arthritis | Arthritis [criteria_fulfilled][within_10days][special_entity_paraclinical_tests_occurred]<br>Pallor, synovitis in<br>bilateral ankles and right wrist.<br>No malar rash, oral ulcers, or skin lesions.<br>Proximal muscle weakness (deltoids/hips) noted<br>Symptoms_Signs<br>no dyspnea or dysphagia. Joint swelling tender but no deformities. |

#### 9.5. Examples absent of SLEDAI-2K score

| Why is it an exception? | Examples |
| --- | --- |

| | |
|---|---|
| Mechanical knee pain(one joint) is pain directly related to movement, weight-bearing, or specific actions. This is more likely a local structural problems like osteoarthritis, or meniscal injury. | Symptoms_Signs<br>C/o right mechanical<br>Arthritis[criteria_unfulfilled_diagnostic][within_10days][special_entity_symptoms_signs_occurred]<br>knee pain<br>Symptoms_Signs Symptoms_Signs<br>~2/52, no trauma/ fever, no effusion, tenderness+ over medial joint line<br>C/o progressive memory loss over the past few months, cannot give a definite example apart from forgetting to take<br>bloods, no focal neurology, FHx unremarkable<br>Management Plan<br>16/52<br>Bld taking today<br>Knee xray |
| Just pain of more than two joints without signs of inflammation were not counted as arthritis. | Same<br>Arthritis[criteria_unfulfilled_diagnostic][within_10days][special_entity_symptoms_signs_occurred] links_te<br>joint pain<br>←links_to Symptoms_Signs<br>over fingers, elbow and knees.<br>Knee pain.<br>Cannot squat down and climbing stairs due to knee pain.<br>Unlimited level ground walking unaided.<br>Symptoms_Signs<br>No active joint inflammation. |

### 10. Myositis

#### 10.1. Clinical definition of concepts

***Myositis*** (Idiopathic Inflammatory Myopathies, IIMs) refers to a heterogeneous group of chronic autoimmune disorders characterized by chronic inflammation of skeletal muscle, leading to progressive muscle weakness (typically proximal and symmetrical) and multi-organ systemic involvement.

#### 10.2. Common ways to describe this concept

There are two widely used classification criteria for IIM, namely the Bohan and Peter criteria (53) and the Tanimoto criteria (54), as summarized in the table below. The subtype of myositis includes dermatomyositis (DM), clinically amyopathic DM (CADM), immune-mediated necrotizing myopathy (IMNM), antisynthetase syndrome (ASS), inclusion body myositis (IBM), and overlap myositis. (55) Overlap myositis was defined as patients fulfilling criteria for IIM plus criteria for other CTD, including systemic lupus erythematosus.

Table. Classification criteria for DM/PM.

| | | |
|---|---|---|
| Bohan and Peter criteria(53) | First, rule out all other forms of myopathies<br>1. Symmetrical weakness, usually progressive, of the limb-girdle muscles with or without dysphagia and respiratory muscle weakness<br>2. Muscle biopsy evidence of myositis<br>Necrosis of type I and type II muscle fibers; phagocytosis, degeneration, and regeneration of myofibers with variation in | Definite PM: all first four elements, probable PM: 3 of first 4, possible PM: 2 of first 4. |

| | | |
|---|---|---|
| | myofiber size; endomysial, perimysial, perivascular, or interstitial mononuclear cells.<br>3. Elevation of serum levels of muscle-associated enzymes (CK, LDH, transaminases, aldolase)<br>4. EMG triad of myopathy<br>a. Short, small, low-amplitude polyphasic motor unit potentials<br>b. Fibrillation potentials, even at rest<br>c. Bizarre, high-frequency repetitive discharges<br>5. Characteristic rashes of dermatomyositis | Definite DM: rash plus 3 others, probable DM: rash plus 2 others, possible DM: rash plus 1 other |
| Tanimoto criteria (54) | 1.Skin lesions<br>a) Heliotrope rash (red purple edematous erythema on the upper palpebra)<br>b) Gottron's sign (red purple keratotic, atrophic erythema, or macules on the extensor surface of finger joints)<br>c) Erythema on the extensor surface of extremity joints: slightly raised red purple erythema over elbows or knees<br>2.Proximal muscle weakness (upper or lower extremity and trunk)<br>3.Elevated serum CK (creatine kinase) or aldolase level<br>4.Muscle pain on grasping or spontaneous pain<br>5.Myogenic changes on EMG (short-duration, polyphasic motor unit potentials with spontaneous fibrillation potentials)<br>6.Positive anti-Jo-1 (histadyl tRNA synthetase) antibody<br>7.Nondestructive arthritis or arthralgias<br>8.Systemic inflammatory signs (fever: more than 37°C at axilla, elevated serum CRP level or accelerated ESR of more than 20 mm/h by the Westergren method)<br>9. Pathological findings compatible with inflammatory myositis (inflammatory infiltration of skeletal muscle with degeneration or necrosis of muscle fibers; active phagocytosis, central nuclei, or evidence of active regeneration may be seen.) | At least 1 item from 1 and at least 4 items from 2 to 9 = DM.<br>At least 4 items from 2 to 9 = PM. |

#### 10.3. SLEDAI-2K definition in this study

As shown in Table 8, our study utilized the original SLEDAI-2K definition of myositis. Exclusions to consider were derived from Bitencourt's work (56). Clinically, magnetic resonance imaging (MRI) has emerged as a valuable diagnostic tool in contemporary clinical practice, where muscle edema found by MRI is a recognized radiological sign of active inflammation (57). The histological features of SLE myopathy in muscle biopsies may overlap with those of subtypes of myositis, including dermatomyositis, polymyositis, and necrotizing myopathy. With findings of necrosis/regeneration, perifascicular atrophy, primary inflammation, endomysial inflammation, and perimysial inflammation (57).

#### 10.4. Examples present of SLEDAI-2K score

| How to determine. | Examples |
|---|---|

| | |
|---|---|
| The patient presented with proximal muscle weakness, accompanied by an elevated creatine kinase level and electromyographic changes. With no other myositis-specific autoantibodies and drug-induced causes excluded, the physician's impression was a lupus flare, necessitating an increase in steroids. | Note<br>--------------------------<br>114/69; P 83; 53 kg; urine S-P trace<br>Currently on MMF 500/250, P15<br>CRP < 0.35 C3 86 C4 16 Plt static 122<br>Myositis[criteria_fulfilled][within_10days][special_entity_paraclinical_tests_occurred][treat_escalated]<br>CK ↑<br>Anti-dsDNA 201.8->140.6<br>No more finger numbness now<br>Symptoms_Signs Paraclinical_tests Exclusions<br>Still proximal muscle weakness EMG myopathic changes. MSA -ve. No statin use.<br>C/o recent SOBOE<br>No orthopnea<br>No fever<br>Symptoms_Signs<br>Generalized malaise<br>No nightsweating<br>Management Plan<br>Intention_to_treat<br>FU 6/52 Start P20 |

#### 10.5. Examples absent of SLEDAI-2K score

| Why is it an exception? | Examples |
|---|---|
| Elevated CK was likely attributed to infection. | Exclusions<br>complained of flu like symptoms,<br>Myositis[criteria_unfulfilled_diagnostic][within_10days][special_entity_symptoms_signs_occurred]<br>myalgia |
| Elevated CK was likely attributed to atorvastatin (Lipitor). | Time[time_uncertain] Exclusions<br>Previously took Lipitor but developed generalized malaise and<br>Myositis[criteria_unfulfilled_diagnostic][time_uncertain][special_entity_symptoms_signs_occurred]<br>myalgia |

### 11. Urinary casts

#### 11.1. Clinical definition of concepts

***Urinary casts*** are tube-shaped particles found in urine under the microscope. Urinary casts can be made up of cells or substances.

#### 11.2. Common ways to describe this concept

Table. Different types of urinary cast in urinalysis (58).

| Type of cast | Composition | |
|---|---|---|
| Cellular | include those comprised of red blood cells (RBCs or Erythrocyte), white blood cells | Nephritis, nephritic syndrome, interstitial nephritis, nephritic |

| | | |
|---|---|---|
| | (WBCs or Leukocyte), and renal tubule (epithelial) cells | syndrome etc. |
| Hyaline (transudation) | composed of mucoproteins | Advanced renal disease |
| Granular | Various cell types | |
| Waxy | comprised of a variety of cell types but have a homogenous structure | Advanced renal disease |
| Fatty | comprised of fat- or lipid-laden cells | Nephrotic syndrome, renal disease, hypothyroidism |
| Broad | Various cell types | End-stage renal disease |

#### 11.3. SLEDAI-2K definition in this study

As shown in Table 9, our study utilized the original SLEDAI-2K definition of urinary casts.

#### 11.4. Examples present of SLEDAI-2K score

| How to determine. | Examples |
|---|---|
| | Urinary_casts [criteria_fulfilled][within_10days]<br>24h UP 2.5, WBC 3+, RBC 2+, heme-granular casts,<br>Urinary_casts[criteria_fulfilled][within_10days]<br>RBC casts, |

#### 11.5. Examples absent of SLEDAI-2K score

| Why is it an exception? | Examples |
|---|---|
| | Hb 132 , WBC 9.7 , Lymph 1.26↓ , ESR 49↑ , Cr 451↑ (baseline 159) , GFR 42↓ , UPCR 2.49g/24h↑ , U/A: innum RBC, 14 WBC/hpf, granular/hyalin<br>Urinary_casts[criteria_unfulfilled_diagnostic][within_10days]<br>casts , ANA+ , dsDNA+.no c-ANCA |

### 12. Hematuria

#### 12.1. Clinical definition of concepts

***Hematuria*** is defined as the presence of blood in the urine and is classified into two main categories. ***Macrohematuria*** refers to visibly apparent blood, typically defined as more than 1 mL of blood per liter of urine. The coloration may range from bright red, suggesting arterial bleeding, to dark red or black, which is often indicative of venous or older hemorrhage. ***Microhematura (Microscopic hematuria)***, by contrast, is detectable only by microscopic examination and is formally defined as the presence of three or more red blood cells per high-power field in at least two out of three properly collected urine specimens (59). Hematuria is divided into glomerular, renal (i.e., nonglomerular), and urologic etiologies. (58)

#### 12.2. Common ways to describe this concept

| Terminology | Definition | Indication |
|---|---|---|
| **asymptomatic microhematuria (aMH)** | defined as the presence of microscopic blood in urine (≥3 red blood cells per high-power field on microscopy) without symptoms (e.g., pain, fever, urinary urgency). | Benign (80% of cases): Idiopathic/non-progressive.<br>Pathological (20%): May indicate occult malignancy, stones, or renal disease. |
| **Glomerular Hematuria** | Dysmorphic RBCs, acanthocytes (>5%), or RBC casts on microscopy. Often with proteinuria (>500 mg/24h). | Indicates renal pathology (e.g., IgA nephropathy, thin basement membrane disease). Nephrology referral needed. |
| **Non-Glomerular Hematuria** | Isomorphic RBCs; no casts. | Associated with urological causes (e.g., UTI, stones, tumors). Urological workup required if risk factors present. |
| **Acanthocytes (G1 cells)** | Ring-shaped RBCs with bleb-like protrusions. | >5% specificity for glomerular disease. |
| **Idiopathic Microhematuria** | Persistent microhematuria without identifiable cause. | ~80% of non-glomerular aMH cases; benign but requires monitoring. |

#### 12.3. SLEDAI-2K definition in this study

In contrast to the widely accepted clinical definition of hematuria as >3 red blood cells per high-power field (RBCs/HPF), the SLEDAI-2k index employs a more stringent threshold of >5 RBCs/HPF. In LN, hematuria is typically of glomerular origin and is often accompanied by significant proteinuria, erythrocyte casts, and the presence of dysmorphic red blood cells (58). Therefore, in SLE patients, isolated hematuria (i.e., hematuria without concomitant proteinuria) is not scored as indicative of active lupus nephritis. Exclusions to consider were derived from the work of Bolenz et al (59).

#### 12.4. Examples present of SLEDAI-2K score

| How to determine. | Examples |
|---|---|
| Given a urinary dipstick result of ≥3+ with low albumin level, treatment with increasing steroids. A corresponding hematuria score was assigned to quantify the possible LN flare, in keeping with the clinician's impression. | Note<br>T 38.2 C<br>↑↑antidsDNA 207 <-- 74<br>↓C3/C4 53/8.2<br>WBC 5.20, PLT 272, Hb 13.5<br>stable renal function and liver function<br>Treated with stepped up steroid on 11th Aug<br>172/86, p 82, bw 50.9, urine protien 3+ve,<br>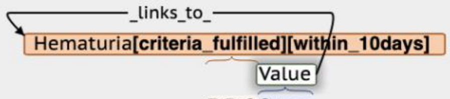<br>RBC3+ve<br>Granular casts: Occ.<br>no alopecia/joint pain/oral ulcer<br>no arthritis/facial skin rash<br>Noted to have low albumin level<br>for 24 hr urine Protein<br>Stepped up steroid to 20 mg QD<br>Management Plan<br>FU 2/52 |

| | |
|---|---|
| proteinuria >1.4g/d with dysmorphic RBC>5/HP, criteria fulfilled. | 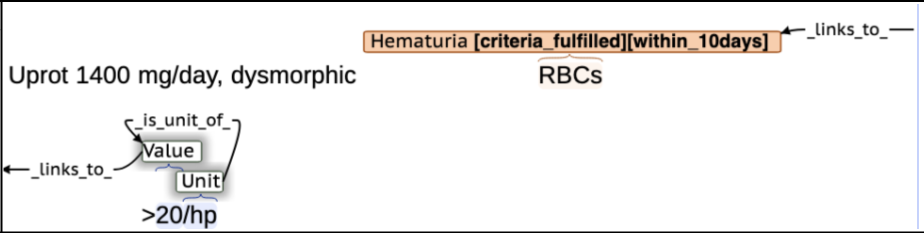 |

#### 12.5. Examples absent of SLEDAI-2K score

| Why is it an exception? | Examples |
|---|---|
| The urine sample may have been contaminated with menstrual blood, which could affect the interpretation of the results. | 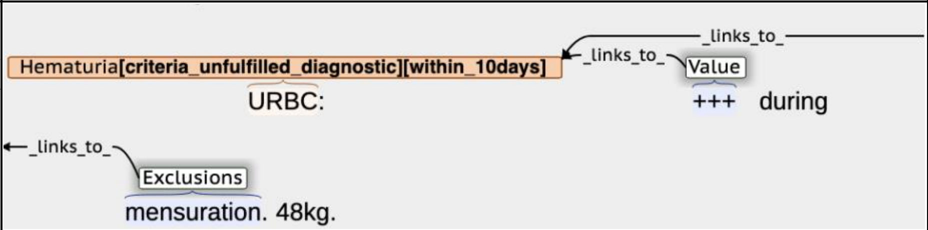 |
| hematuria without concomitant proteinuria is not likely lupus related. | 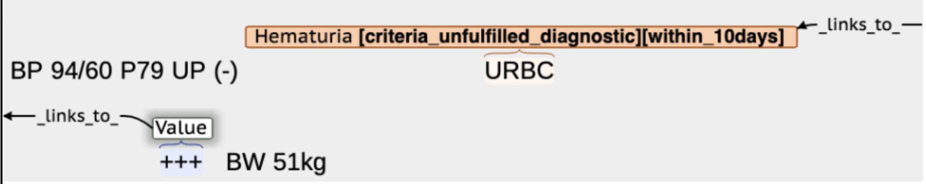 |

### 13. Proteinuria

#### 13.1. Clinical definition of concepts

***Proteinuria*** is defined as the presence of protein in urine, specifically indicating abnormally elevated levels of protein excreted by the kidneys. It serves as a key biomarker for kidney disease and carries significant prognostic value. (60)

#### 13.2. Common ways to describe this concept

Table. Laboratory methods for detecting proteinuria (60).

| **Specific Test/Technique** | **Reference Range (Normal)** | **Clinical Use** |
|---|---|---|
| **Semiquantitative** | | |
| Nonspecific Dipstick (e.g., Combur-9 Test®) | Trace or ≥1+ (≈30 mg/dL) = abnormal | Screening to rule out proteinuria if quantitative tests unavailable |
| Albumin-Specific Dipstick (e.g., Micral-Test®) | ≥2 mg/dL albumin = abnormal | Early albuminuria detection; not ideal for screening due to false positives |
| **Quantitative** | | |
| 24-Hour Urine Collection | Total protein <150 mg/24 h | Rarely used first-line; replaced by spot ratios |
| Urine Albumin-Creatinine Ratio (UACR) | CKD Staging:<br>•A1: <30 mg/g (<3 mg/mmol)<br>• A2: 30–300 mg/g (3–30 mg/mmol)<br>• A3: >300 mg/g (>30 mg/mmol) | Preferred for screening, diagnosis, and monitoring (KDIGO guidelines) |

| | | |
|---|---|---|
| Urine Protein-Creatinine Ratio (UPCR) | CKD Staging:<br>• A1: <150 mg/g (<15 mg/mmol)<br>• A2: 150–500 mg/g (15–50 mg/mmol)<br>• A3: >500 mg/g (>50 mg/mmol) | Alternative if UACR unavailable; detects paraproteinuria if UACR gap is high |
| Tubular Protein Markers (e.g., α1-microglobulin) | α1-microglobulin <15 mg/g | Suspected tubulointerstitial disease (e.g., drug-induced nephritis) |

#### 13.3. SLEDAI-2K definition in this study

As shown in Table 9, we adopted the SLEDAI-2K criteria for proteinuria and utilized the spot ***urine protein-to-creatinine ratio*** (UPC or UPCR) as a surrogate measure, as endorsed by the 2019 EULAR/ACR Classification Criteria (10) and SLICC 2012 (11). In the management of lupus nephritis, the UPC has been widely adopted in clinical guidelines as a validated surrogate test for 24-hour urine protein. Although the SLEDAI-2K defines proteinuria as >0.5 g/24 hours, current guidelines do not specify a universally applicable UPC equivalent for this threshold. This omission reflects the fact that the optimal UPC cutoff corresponding to 0.5 g/24 hours may vary depending on laboratory methodologies and patient populations, as supported by previous studies (61, 62). Reported UPC cutoff values for a 24-hour proteinuria of 0.5 g/day include 0.08 g/mmol (800 mg/g) from a Toronto cohort (63), 0.45 mg/mg from Hong Kong (64), and 0.44 mg/mg from Nanjing (65). In the present study, we applied a UPC cutoff of 0.5 mg/mg (equivalent to 50 mg/mmol) equivalent to 24-hour proteinuria of 0.5 g/day, which aligns closely with the Hong Kong data and is also consistent with the proteinuria remission criterion for lupus nephritis recommended by the KDIGO guidelines (66). Additionally, we did not use urine dipstick results solely for diagnosis of proteinuria due to their poor correlation with quantitative 24-hour urine protein measurements (67). For patients with a dipstick reading of 3+ or higher in whom concurrent UPC or 24-hour urine protein data were unavailable, a proteinuria score was assigned to reflect a disease flare if the treating clinician's impression was consistent with lupus nephritis flare.

#### 13.4. Examples present of SLEDAI-2K score

| How to determine. | Examples |
|---|---|
| 2.95g/d is more than 0.5g/d | 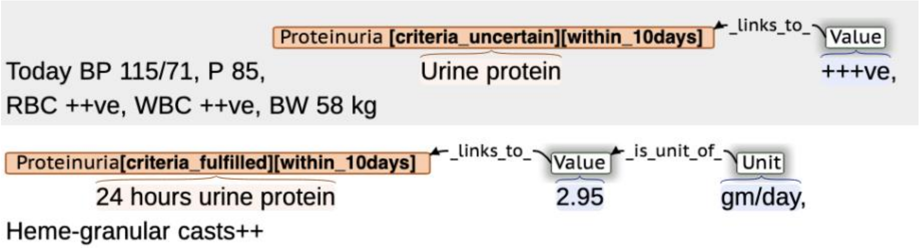 |

| | |
|---|---|
| Given a urinary dipstick result of ≥3+ and renal biopsy confirming active lupus nephritis, treatment with steroids and mycophenolate mofetil was initiated. A corresponding proteinuria score was assigned to quantify the disease flare, in keeping with the clinician's impression. | Note<br>_links_to_<br>Proteinuria [criteria_fulfilled][within_10days]<br>Value<br>BP 133/89 P 121 BW 48.4kg UP++++,<br>RBC+++,WBC ++ve,Heme-granular casts++,<br>Renal biopsy done on showed active class IV nephritis<br>Explained to patient the need to increase steroid and add MMF, and the need to closely monitor LFT after stepping up<br>steroid dosage. Potential GI side effects of MMF explained<br>Management Plan<br>CBP, L/RFT, antidsDNA, C3/4 today<br>Repeat LFT x 1/52<br>Intention_to_treat<br>Increase prednisolone 35mg qd<br>Intention_to_treat<br>Add MMF 1g/750mg bd<br>Add pepcidine 20mg bd<br>Add slow K 600mg qd<br>Lasix 20mg qd prn |
| UPC 64 mg/mmol is equivalent to 24h UP 0.64 g/d. 0.64 > 0.5 g/d so criteria fulfilled. | _links_to_<br>Value<br>Proteinuria[criteria_uncertain][within_10days]<br>urine P+<br>CRP <0.35<br>C3 64 C4 12<br>anti-dsDNA increasing trend 143→188→228<br>Proteinuria[criteria_fulfilled][within_10days] _links_to_ Value<br>urine P/C 64 (previous 40-70) |

#### 13.5. Examples absent of SLEDAI-2K score

| Why is it an exception? | Examples |
|---|---|

| | |
|---|---|
| Increased urine protein was attributed to a urinary tract infection. | Exclusions<br>MSU - significant growth of E coli sensitive to ampicillin<br>Proteinuria[criteria_unfulfilled_diagnostic][within_10days] Value<br>UPC 178<br>Exclusions<br>USG urinary system showed a tiny echogenic focus in L mid pole,<br>Exclusions<br>suspicious of renal stone or vascular calcification<br>No dysuria / haematuria / abdominal pain<br>Still reluctant for repeat biopsy<br>Had very bad experience previously<br>Management Plan<br>FU 12/52<br>Intention_to_treat<br>Augmentin x 1/52<br>Blood tests and UPC before next visit<br>Repeat MSU x R/M and c/st before next visit<br>Early NCCT kidneys |
| UPCR 0.18 is equal to 24hUP 0.18g/d, which is less than 0.5g/d. | Proteinuria[criteria_unfulfilled_diagnostic][within_10days] ←_links_to_ Value<br>UPCR 0.18 |

### 14. Pyuria

#### 14.1. Clinical definition of concepts

***Pyuria*** is defined by the presence of one or more of the following diagnostic findings: a urine sample containing ≥10 white blood cells (WBCs) per cubic millimeter, the identification of ≥3 white blood cells per high-power field in unspun urine, a positive Gram stain result from an unspun urine specimen, or a positive leukocyte esterase reaction on a urinary dipstick test. (68) The quantitative threshold ⩾ $10^5$ CFU/ml (colony-forming units per milliliter) was the most frequently used threshold for a urine culture to be considered bacteriuria.

#### 14.2. Common ways to describe this concept

| | definition | Causes |
|---|---|---|
| ***Sterile Pyuria*** (68) | The persistent finding of white blood cells (pyuria) in the urine without detectable bacteria via standard aerobic culture methods. | Infections: Sexually transmitted infections, viruses, tuberculosis, fungi, parasites<br>Systemic Inflammation: Autoimmune diseases (e.g., **lupus**), interstitial nephritis/cystitis<br>Anatomical Issues: Kidney stones, tumors, foreign bodies, recent catheter use<br>Other: Recent antibiotics, pelvic radiation, inflammation near the urinary tract (e.g., appendicitis) |

| | | |
|---|---|---|
| ***Non-Sterile Pyuria*** (Bacteriuria) | The persistent finding of white blood cells (pyuria) in the urine with detectable bacteria via standard aerobic culture methods. | Asymptomatic Bacteriuria (ASB): the presence of bacteria in the urine without any symptoms of a urinary tract infection.<br>Urinary Tract Infection (UTI): bacteriuria accompanied by clinical signs and/or symptoms (e.g., fever, pain, increased spasticity, cloudy urine, autonomic dysreflexia). |

#### 14.3. SLEDAI-2K definition in this study

The SLEDAI-2K defined ***pyuria*** as ">5 white blood cells/high-power field. Exclude infection." In contrast to the widely accepted clinical definition of pyuria as >3 white blood cells per high-power field (WBCs/HPF), the SLEDAI-2K employs a more stringent threshold of >5 WBCs/HPF. Exclusions to consider were derived from (54). We also allow for qualitative results and provided the reference range for our central lab to determine increased urine RBC, as shown in Table 9.

#### 14.4. Examples present of SLEDAI-2K score

| How to determine. | Examples |
|---|---|
| Elevated urine protein (>0.5 g/d), pyuria (>5 WBCs per hpf), and a negative urine culture collectively indicated a lupus nephritis flare. | 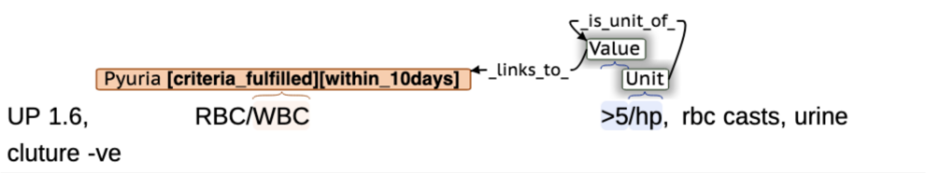 |

#### 14.5. Examples absent of SLEDAI-2K score

| Why is it an exception? | Examples |
|---|---|
| The elevated urine white blood cell count was attributed to a urinary tract infection according to doctors intention to initiate antibiotics. | 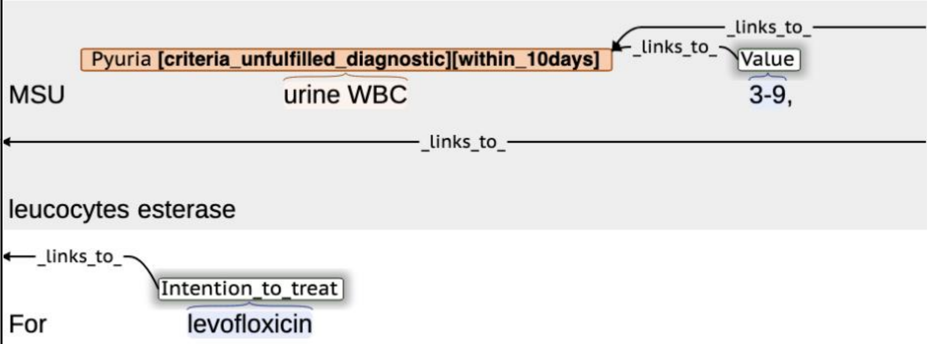 |
| Cannot rule out urinary tract infection according to a positive cultrure. | 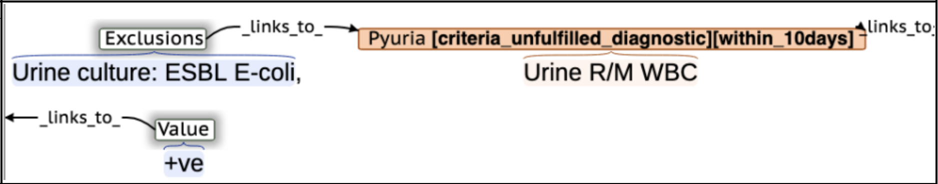 |

### 15. Rash

#### 15.1. Clinical definition of concepts

A rash represents a common clinical manifestation of numerous underlying conditions, spanning a spectrum from benign allergic reactions and localized irritations to severe infections, systemic diseases, and malignancies. In the SLEDAI-2K index, the term 'rash' specifically refers to ***cutaneous lupus erythematosus*** (CLE) occurring with SLE. CLE is categorized into three principal types: (1) ***acute cutaneous lupus***, (2) ***subacute cutaneous lupus***, and (3) ***chronic cutaneous lupus***. (69)

#### 15.2. Common ways to describe this concept

Under the three main categories of CLE, numerous specific disease entities should be considered as diagnostic keywords for rash in SLEDAI-2K (see Table below). Furthermore, the Cutaneous Lupus Disease Area and Severity Index (CLASI) underscores the necessity of distinguishing between disease activity and damage when evaluating lupus-related skin lesions (70).

Table. Classification of cutaneous lupus erythematosus. (71)

| Class | Name | Signs |
|---|---|---|
| **Acute cutaneous lupus erythematosus** | Malar rash (butterfly rash or Butterfly erythema) | fixed erythema, flat or raised, over the malar eminences, tending to spare the nasolabial folds (72) |
| | Bullous lupus erythematosus (BLE). | Solitary small vesicles or groups of vesicles, or larger, firm subepidermal blisters on erythematous or normal skin |
| | Photosensitivity rash | skin rash as a result of unusual reaction to sunlight, by patient history or physician observation.<br>sun-exposed areas (V-shaped area of upper chest, back, extensor surfaces of arms, lateral and posterior neck) |
| | Generalized form | A. Exanthema: morbilliform or maculopapular affecting skin of entire body, palms/soles and interphalangeal extensor aspects of the fingers, erythema of nail fold and telangiectases, red lunula; rarely transformation into toxic epidermal necrolysis(TEN)<br>B. Enanthema:<br>1. Erythema, erosions, superficial ulcerations<br>2. Localization: hard palate > gingiva and buccal mucosa |
| **Subacute cutaneous lupus erythematosus** | Annular form | signs： Ring-like or oval, erythematous plaques with training scale and central clearing |
| | Papulosquamous form | signs :Papulosquamous plaques, possibly transforming into clinical picture resembling psoriasis<br>Combination of both forms is possible |
| **Chronic cutaneous LE** | Discoid lupus erythematosus (DLE) | signs :Erythematous-violaceous cutaneous lesions with secondary changes of atrophic scarring, dyspigmentation, often follicular hyperkeratosis/ plugging (scalp), leading to scarring alopecia on the scalp. |
| | Lupus erythematodes profundus (LEP) | signs :Subcutaneous nodules and plaques, later adhering to overlying skin |

| | | |
|---|---|---|
| | | • Lesional surface: reddened with inflammation, unchanged or concomitant DLE<br>• Predilection sites: face, shoulders, upper arms, chest, buttocks, thighs, hips |
| | Chilblain lupus erythematosus (CHLE) | Signs:Tender, bright red edema and puffy nodules sometimes with central erosion and ulceration<br>• Predilection sites: cold-exposed acral areas (dorsal and marginal areas of the fingers, tips of the toes, heels, ears, nose) |
| | Lupus erythematosus tumidus (LET) | signs :Succulent, urticaria-like, erythematous plaques with a smooth surface and no epidermal involvement<br>• Predilection sites: sun-exposed areas (especially the face, upper trunk, and extensor surfaces of the arms) |

A recent study (73) utilizing a large language model (LLM) to assess CLASI has compiled a vocabulary dictionary to differentiate active from damaged lesions, providing a valuable methodological reference for identifying active rashes in our research (see the Table below).

**Table. Data vocabulary dictionary for relevant terms of the CLASI scoring system.** (73)

| | | |
|---|---|---|
| **Activity** | **Erythema** | pink, faint erythema, pinkish, faint erythema,<br>faintly erythematous, mild erythema, mildly erythematous, mild redness, mildly red<br>red, erythematous, erythema, moderate erythema,<br>moderately erythematous. moderate erythematous,<br>red, redness, reddish<br>dark red, deep red, deeply red, deep reddish, deeply reddish, purple, purplish, violaceous |
| | **Scale/Hypertrophy** | Scale, scaling, scaly, follicular plugging,<br>follicular prominence, carpet tacking,<br>carpet tack sign, perifollicular scale<br>Hypertrophy, hypertrophic, hyperkeratosis,<br>hyperkeratotic |
| **Damage** | **Dyspigmentation** | Hypopigmentation, hypopigmented,<br>hyperpigmentation, hyperpigmented,<br>dyspigmentation, dyspigmented,<br>pigmentary changes, brown, depigment,<br>white |
| | **Scarring/Atrophy/Panniculitis** | Scarring, scar<br>Atrophic scarring, atrophic, panniculitis,<br>lipoatrophy, atrophic scar |

#### 15.3. SLEDAI-2K definition in this study

The SLEDAI-2K defines an active rash broadly as an "***inflammatory-type rash***." To precisely define the disease entities encompassed by this item, we refer to the more detailed subgroup classifications provided by the SLICC 2012 criteria as shown in Table 8 (11). In line with SLEDAI-2K's principle, our protocol scores only active, inflammatory signs to reflect current disease activity. A data vocabulary dictionary distinguishing active features (e.g., erythema) from damage (e.g., scarring) is provided in the table above (73). Therefore, lesions with damage alone, without active signs, will not be considered a rash for scoring. Exclusions to consider were derived from Kuhn's work (71).

#### 15.4. Examples present of SLEDAI-2K score

| How to determine. | Examples |
|---|---|
| A diagnostic keyword | PE:<br>Rash[criteria_fulfilled][within_10days]<br>Malar rash (+) |
| A diagnostic keyword | PE<br>Rash[criteria_fulfilled][within_10days]<br>lupus erythema, no ulceration |
| Rashes appearing in classic locations (face, etc.) with characteristic morphology and distribution (maculopapular, nasolabial sparing). The lesions demonstrated both active (maculopapular) and post-inflammatory (hyperpigmented) morphologies. Moreover, the patient is simultaneously experiencing multi-system lupus activity affecting the kidneys, blood, and nervous system. Therefore, the active rash was very likely lupus related and marked as fulfilling diagnostic criteria. | PE<br>Symptoms_Signs<br>hyperpigmented maculopapular<br>Rash[special_entity_symptoms_signs_occurred][criteria_fulfilled][within_10days]<br>rash<br>(face/neck/back/extremities; spares nasolabial folds).<br>Neuro: Fluctuating orientation (time/place/person), hypervigilance, persecutory delusions, incoherent speech. No focal deficits.<br><br>Labs: Hb 7.6↓, WBC 3.4↓, PLT 83↓, albumin 3.0↓, TSH 16.06↑, ANA ++++,UPCR 501<br>Anti-Sm +++ (not quantified), APLA neg.<br>Imaging: MRI brain (DWI): Tiny diffusion bright foci (bilateral centrum semiovale/parietal gyri) → suggests vasculitis.<br><br>Impression<br>Active SLE flare with Delirium + MRI vasculitis. steroid induce psychosis<br>Renal: Active LN (proteinuria 3+).<br>Hematologic: Anemia, thrombocytopenia. |

#### 15.5. Examples absent of SLEDAI-2K score

| Why is it an exception? | Examples |
|---|---|
| The facial rash was not necessary lupus related. Current lupus stable, tapering steriods. | Minimal<br>Rash[criteria_unfulfilled_diagnostic][within_10days][special_entity_symptoms_signs_occurred]<br>facial rash.<br>Minimal arthralgia<br>No fever<br>No Raynaud's<br>No alopecia<br>No dysuria<br>Discussed with Dr CAREY<br>To reduce Prednisolone to 10mg daily. FU 8/52 |

| | |
|---|---|
| Quiescent rash was not counted. | 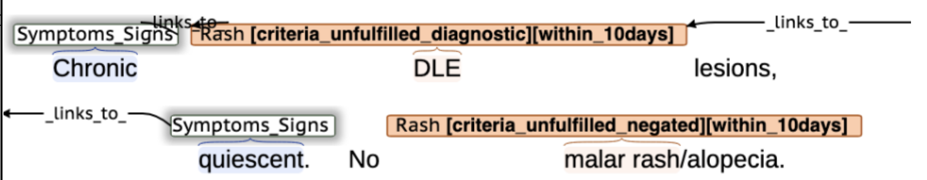 |

### 16. Alopecia

#### 16.1. Clinical definition of concepts

***Alopecia*** is a symptom of partial or complete hair loss from the scalp and other areas of the body. It has various causes, including androgenetic alopecia, alopecia areata, and cicatricial alopecia.

| Types | Causes | Explain |
|---|---|---|
| Nonscarring Alopecia | androgenetic alopecia (pattern baldness) | hereditary and hormonal |
| | alopecia areata(spot baldness) | autoimmune condition where the immune system attacks hair follicles |
| | telogen effluvium | triggered by stress, illness, or childbirth |
| | anagen effluvium | chemotherapy or radiation therapy |
| | traction alopecia | wear pigtails, braids or cornrows, or use tight hair rollers |
| Scarring Alopecia | cicatricial alopecia | destroys hair follicles and causes scarring, leading to permanent hair loss |

#### 16.2. Common ways to describe this concept

| **Symptom** | **Explain** |
|---|---|
| Alopecia Synonyms | Hair shedding/hair loss/alopecia/baldness |
| Patchy hair loss | Small, round, smooth patches on the scalp or other areas of hair growth. |
| Thinning hair | Generalized thinning of hair on the scalp. |

#### 16.3. SLEDAI-2K definition in this study

The SLEDAI-2K defines alopecia as "abnormal, patchy, or diffuse hair loss". The SLICC 2012 (11) and 2019 EULAR/ACR Classification Criteria (10) also specify ***non-scarring alopecia*** to distinguish it from the scarring alopecia characteristic of chronic discoid lupus. To enhance precision in scoring, our study incorporated this specification into the assessment definition of the SLEDAI-2K as shown in Table 8.

#### 16.4. Examples present of SLEDAI-2K score

| How to determine. | Examples |
|---|---|

| | |
|---|---|
| | Alopecia [criteria_fulfilled][within_10days]<br>maculopapular rash, alopecia.<br>PMH: Febrile illness treated w/ antimalarials at seizure onset.<br><br>PE:<br>Mucocutaneous: Malar rash (nasolabial fold sparing), generalized maculopapular<br>Symptoms_Signs<br>rash (trunk/proximal limbs), patchy alopecia. |
| | Symptoms_Signs —links_to— Alopecia [criteria_fulfilled][within_10days]<br>abrupt hair loss, facial/extremity rash. |

#### 16.5. Examples absent of SLEDAI-2K score

| Why is it an exception? | Examples |
|---|---|
| Stress caused alopecia is an exclusion. | 1 Note<br>Alopecia [criteria_unfulfilled_diagnostic][within_10days] —links_to— Exclusions<br>2 c/o alopecia due to stress<br>3 No other symptom<br>4 Management Plan |

### 17. Mucosal ulcers

#### 17.1. Clinical definition of concepts

A ***mucosal ulcer*** is a break in the mucous membrane with loss of surface tissue and the disintegration and necrosis of epithelial tissue. The causes underlying mucosal ulcers are complex, involving local trauma, vitamin deficiency, aphthous stomatitis, infection (e.g., herpes simplex virus, Epstein-Barr virus, HIV), immune dysregulation (SLE, Behçet's disease, inflammatory bowel diseases), drugs, and, in some cases, malignancies.

#### 17.2. Common ways to describe this concept

| | |
|---|---|
| Synonyms of oral ulcer | mouth ulcer<br>mouth aphtha<br>canker sores<br>salt blister<br>aphthous ulcerations<br>aphthous ulcers<br>stomatitis |
| Positions indicating unclers in mouth | oral cavity<br>oral<br>mouth<br>mucosal<br>nasal<br>nasopharyngeal<br>palate/ palatal |

| | |
|---|---|
| | buccal<br>tongue |

##### 17.3. SLEDAI-2K definition in this study

As shown in Table 8, our study applied the mucosal ulcer definition and exclusions established by SLICC 2012 (11), which provided more details than the original SLEDAI-2K definition.

##### 17.4. Examples present of SLEDAI-2K score

| How to determine. | Examples |
|---|---|
| The clinician considered the buccal erosions to be lupus-related; although not an ulcer, they were still scored. | Skin: Hyperpigmentation, erythematous lesions, alopecia,<br>Mucosal_ulcers[criteria_fulfilled][within_10days]<br>buccal erosions. |
| Hard palate ulcer is also oral ulcer. | Mucosal_ulcers _links_to_ Symptoms_Signs<br>Discoid rash, ulcer on the hard palate |

##### 17.5. Examples absent of SLEDAI-2K score

| Why is it an exception? | Examples |
|---|---|
| absent is a negation word | Mucosal_ulcers [criteria_unfulfilled_negated][within_10days]<br>Absent:Synovitis, oral ulcers,<br>dysphagia, renal/pericardial rub. |

#### 18. Pleurisy

##### 18.1. Clinical definition of concepts

***Pleurisy***, also known as ***pleuritis***, refers to the inflammation of the parietal pleura, the membrane lining the chest cavity. This condition commonly manifests as characteristic pleuritic pain, which is a sharp, localized discomfort exacerbated by breathing, coughing, or sneezing. There is a broad spectrum of pleurisy etiologies, including relatively benign conditions like viral infections and more severe causes such as pulmonary embolism, myocardial infarction, or malignancy.

##### 18.2. Common ways to describe this concept

Table. Clinical characteristics of pleurisy. (74)

| | |
|---|---|
| Characteristic Pain | Sharp, localized chest pain<br>Aggravated by deep breathing, coughing, sneezing, or movement<br>May be referred to shoulder/neck (central diaphragm involvement)<br>Onset: Acute, subacute, or chronic |
| Physical Exam | Pleural friction rub (hallmark finding)<br>Rapid, shallow respirations<br>Posturing to limit chest movement |
| Paraclinical tests | Chest X-ray/CT scan/MRI: May show pleural effusion (small/moderate)<br>Pleural Fluid Analysis (to distinguish exudate vs transudate) |

| | |
|---|---|
| | ECG: can be performed to rule out cardiac causes |

#### 18.3. SLEDAI-2K definition in this study

As shown in Table 8, our study utilized the original SLEDAI-2K definition of pleurisy, supplemented with necessary clarifications regarding imaging evidence. We derived exclusions from the SLICC 2012 (11). The SLEDAI-2K criterion mandates the presence of pleuritic pain, which is intended to identify acute pleurisy and not chronic sequelae.

#### 18.4. Examples present of SLEDAI-2K score

| How to determine. | Examples |
|---|---|
| Serosis is a diagnostic keyword for pleurisy. The diagnosis is supported by the presence of acute pleuritic pain, a physical examination revealing a friction rub, and imaging-confirmed effusions. | Vitals:HR ↑, Temp 39.5°C, RR ↑<br>Complaint & HPI:<br>Symptoms_Signs<br>Complaint: Acute pleuritic chest pain, SOB.<br>HPI: Bilateral wrist synovitis at presentation.<br>Prior Tx: Indomethacin 25mg TID + colchicine 0.6mg BID → symptom resolution. Later started prednisone 20mg daily.<br>PE:<br>Symptoms_Signs<br>Pericardial rub.<br>Bilateral wrist synovitis (tender, swelling).<br>Paraclinical:<br>Trop 0.35↑, CRP 13.6↑, ESR 91↑.<br>Paraclinical_tests<br>CT: 12mm PE + bilateral pleural effusions.<br>Serology: ANA 1:1280, dsDNA+, Smith+, SM/RNP+.<br>Impression:<br>Pleurisy [criteria_fulfilled][within_10days]<br>Active SLE: Serositis, synovitis.<br>DDx: Viral pericarditis excluded (neg infectious workup).<br>Plan:<br>Continue colchicine 0.6mg BID ×3mo. |

#### 18.5. Examples absent of SLEDAI-2K score

| Why is it an exception? | Examples |
|---|---|
| Tuberculosis is exclusion | Exclusions —links to— Pleurisy [criteria_unfulfilled_diagnostic][time_uncertain]<br>TB pleuritis<br>anti TB Rx since |

| | |
|---|---|
| No chest pain. Likely a stable, fibrotic scar from a prior episode of pleural inflammation. | Note<br>BP 136/82 P 64<br>UP- rbc -<br>Serology negative<br>Symptoms_Signs<br>no symptoms<br>LFT normalised<br>CXR: Apical<br>Pleurisy[criteria_unfulfilled_diagnostic][within_10days][special_entity_paraclinical_tests_occurred]<br>pleural thickening<br>and LLZ old inflammatory changes<br>Management Plan<br>FU 12 wks |

### 19. Pericarditis

#### 19.1. Clinical definition of concepts

***Pericarditis*** is an inflammatory condition of the pericardium, which may have infectious or non-infectious causes. The disease can manifest as an isolated condition or as a component of another systemic disorder involving the pericardium (e.g., SLE, rheumatoid arthritis, lung cancer, or renal failure). (75)

#### 19.2. Common ways to describe this concept

Table. Classification of pericarditis. (75)

| Terminology | Definition |
|---|---|
| ***Acute Pericarditis*** | First episode of pericarditis with acute symptom onset. |
| ***Incessant Pericarditis*** | Persistent inflammation without a symptom-free interval ≥4-6 weeks after the initial episode. |
| ***Recurrent Pericarditis*** | Reappearance of symptoms after a symptom-free interval ≥4-6 weeks. |
| ***Chronic Pericarditis*** | Inflammation lasting >3 months. |
| ***Constrictive Pericarditis*** | Scarring/stiffening of the pericardium with impaired cardiac filling. |
| ***Effusive-Constrictive Pericarditis*** | Coexisting pericardial effusion (tamponade) with constrictive physiology. |
| ***Myopericarditis*** | Pericarditis with concomitant myocardial involvement |

Clinical characteristics of pericarditis. (75)

| | |
|---|---|
| Characteristic Pain | Chest Pain: Sharp, pleuritic; Improves when sitting/leaning forward |
| Physical Exam | Pericardial Friction Rub:Scratching/squeaking sound; Best heard at left sternal border |
| Paraclinical tests | ECG Changes:Widespread concave ST elevation; PR depression<br>Echocardiography/CT/Cardiac<br>MRI: Pericardial thickening/enhancement/effusion<br>Chest X-ray: Enlarged cardiac silhouette ("water bottle sign") if effusion >200mL<br>Elevated Inflammatory Markers: CRP (most clinically useful), ESR, leukocytosis, Troponin elevation (if myocarditis involved) |

#### 19.3. SLEDAI-2K definition in this study

As shown in Table 8, our study utilized the original SLEDAI-2K definition of pericarditis, supplemented with necessary clarifications regarding imaging evidence. We derived exclusions from the SLICC 2012 (11). The SLEDAI-2K criterion mandates the presence of pericardial pain, which is intended to identify acute pericarditis.

#### 19.4. Examples present of SLEDAI-2K score

| How to determine. | Examples |
|---|---|
| Pain with echo confirmed effusion, criteria fulfilled. | Paraclinical_tests (_links_to_)<br>Recent pericardial chest pain<br>Paraclinical_tests (_links_to_, _links_to_)<br>Echo: Mild pericardial effusion, no tamponade effect.<br>(_links_to_, _links_to_)<br>--> will see cardiac 6/2010<br>(_links_to_, _links_to_)<br>Management Plan<br>(_links_to_, _links_to_) Pericarditis [criteria_fulfilled][within_10days]<br>Admit for workup and start treatment for pericarditis related to SLE |
| Imaging confirmed a pericardial effusion, and physical examination revealed a pericardial rub. Although the patient did not report typical pleuritic pain, the altered mental status may have impaired symptom reporting. Therefore, pericarditis was scored. | Symptoms_Signs<br>Pericardial rub auscultated.<br>No synovitis.<br>Hb 8↓, WBC 29.1k↑, PMNs 96%↑; $Na^+/K^+$/Urea/Cr ↑ (trending up)<br>CSF (07/2015): Protein↑, Glucose↑<br>Paraclinical_tests<br>Imaging: pericardial effusion<br>Brain MRI/EEG nl<br>Serology (historical): dsDNA+, hypocomplementemia (C3↓)<br>Impression<br>Active SLE flare w/ Steroid-induced delirium (Naranjo score 7)<br>Pericarditis[criteria_fulfilled][within_10days]<br>Pericarditis (rub + effusion) |

#### 19.5. Examples absent of SLEDAI-2K score

| Why is it an exception? | Examples |
|---|---|

| | |
|---|---|
| The finding of a suspected pericardial effusion does not constitute strong evidence for pericarditis in the absence of chest pain or other supportive signs; therefore, it was not scored. | Note<br>124/64; P 88; Urine protein3+; RBC 2+, 55 kg<br>recent admission for chest infection → augmentin<br>Cr 172; US: parenchymal disease<br>urine p/c ratio 125, red blood cell casts +ve,<br>WCC 2.71; INR 2.8 (requested to try dabigatran)<br>serology improving though anti-ds DNA still ↑<br>CXR: cardiac shadow suspicious of<br>Pericarditis[criteria_unfulfilled_diagnostic][within_10days][special_entity_paraclinical_tests_occurred] links_t<br>pericardiac effusion<br>_links_to_ History<br>(prev. Hx. of pericarditis)<br>Management Plan<br>step up pred; book echo<br>try dabigatran; off warfarin; repeat bloods@ 4 wks<br>8 wks |

### 20. Low complement

#### 20.1. Clinical definition of concepts

The complement system is a sophisticated proteolytic network comprising over 30 soluble and membrane-bound proteins and glycoproteins present in blood plasma and on cell surfaces. It plays a fundamental role in innate immunity and serves as a bridge to adaptive immune responses.(76) Low serum levels of complement components C3 and C4 typically indicate consumption from immune activation, impaired synthesis, or, less commonly, genetic deficiencies. SLE is characterized by autoantibodies (e.g., anti-dsDNA, anti-histone) that form immune complexes (ICs) with self-antigens. These ICs deposit in tissues (e.g., kidneys, skin, vessels) and activate the classical complement pathway via C1q binding. This triggers a cascade for the consumption of C4, C2, C3, and terminal components (C5-C9). C3 and C4 are consumed fast duringd lupus flare, leading to reduced serum levels. (77) The CH50 (total hemolytic complement) assay measures the functional activity of the entire classical pathway.

#### 20.2. Common ways to describe this concept

***Hypocomplementemia*** refers to a decrease in serum complement levels below the normal reference range, which may involve reduced CH50, C3 or C4. It is critical to recognize that reference intervals are assay-dependent and can vary between laboratories.

#### 20.3. SLEDAI-2K definition in this study

As shown in Table 9, our study utilized the original SLEDAI-2K definition of low complement. If the reference ranges were unspecified, we interpreted the results according to reference ranges from our laboratory.

#### 20.4. Examples present of SLEDAI-2K score

| How to determine. | Examples |
|---|---|

| | |
|---|---|
| before 24/12/2018, the normal references is <76 for C3, <9 for C4. Therefore low C3 fulfilled, low C4 unfilfilled. Should be scored. | 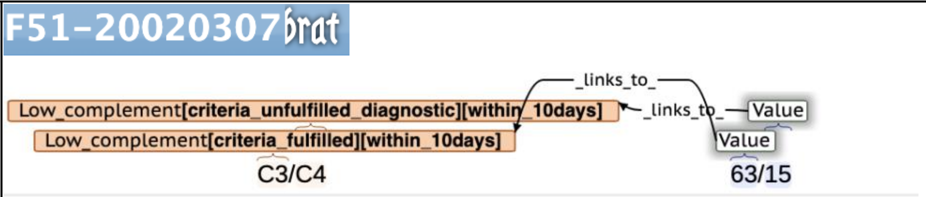  |
| Low indicates lower than reference. Criteria fulfilled. | 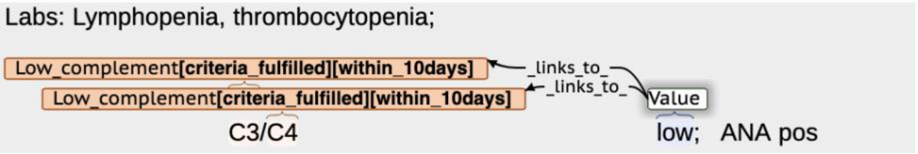  |
| Low indicates lower than reference. Criteria fulfilled. | 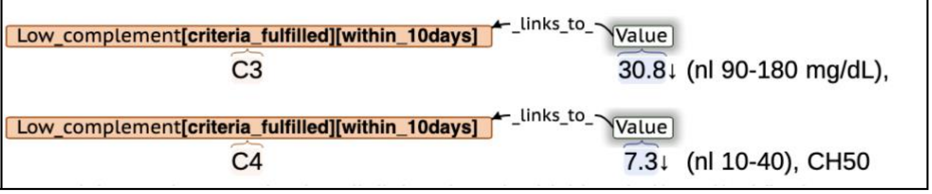  |

#### 20.5. Examples absent of SLEDAI-2K score

| Why is it an exception? | Examples |
|---|---|
| before 24/12/2018, the normal references is <9 for C4. Therefore, low C4 unfilfilled. Should not be scored. | 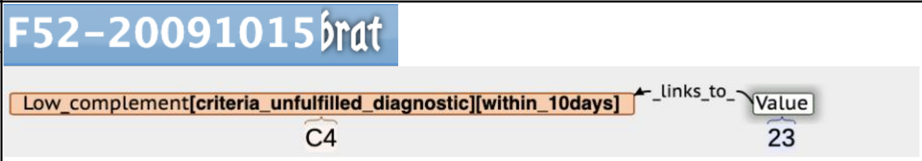  |
| | 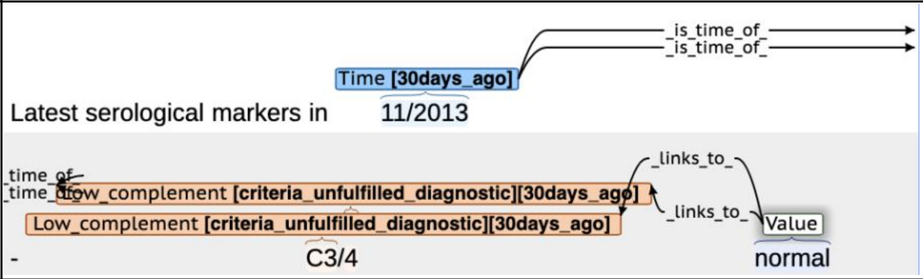  |

### 21. Increased DNA binding

#### 21.1. Clinical definition of concepts

***Increased anti-dsDNA antibodies*** refer to the pathological elevation of autoantibodies targeting endogenous double-stranded DNA (self-dsDNA) in SLE. The accumulation of self-dsDNA in SLE arises from defective clearance of cellular debris and dysregulated cell death, combined with impaired degradation pathways. Increased anti-dsDNA antibodies in SLE arise from defective clearance of self-dsDNA and breakdown of B/T-cell tolerance, amplified by IFN-I-driven inflammation. Testing for anti-dsDNA antibodies is a crucial blood test primarily used to help diagnose SLE, as these antibodies are highly specific to the disease. Furthermore, the test is used to monitor disease activity, as rising levels often predict or coincide with flares, particularly serious complications like kidney inflammation (lupus nephritis).(78)

#### 21.2. Common ways to describe this concept

Table. Several methodologies are available for the detection of anti-dsDNA antibodies.(79)

| **Test Method** | **Test Method** | **Key Characteristics & Notable Limitations** |
|---|---|---|
| FEIA<br>(e.g., EliA dsDNA) | Fluorescence enzyme immunoassay | automated |
| CLIFT<br>(e.g., Euroimmun) | Crithidia luciliae immunofluorescence | visual kinetoplast staining<br>technical expertise required |
| ELISA | Enzyme-linked immunosorbent assay | widely accessible<br>Inconsistent performance |
| CLIA<br>(e.g., QUANTA Flash) | Chemiluminescence immunoassay | high-throughput |
| MIA<br>(e.g., BioPlex) | Multiplex immunoassay | simultaneous autoantibody detection |
| Farr-RIA | Radioimmunoassay | historically "gold standard"<br>radioactive reagents |
| Farr-FIA | Fluorescent immunoassay | non-radioactive alternative to Farr-RIA |

#### 21.3. SLEDAI-2K definition in this study

The original SLEDAI-2K definition only specifies criteria of increased DNA binding via the Farr assay method. However, considering wider types of methods were used in the clinic, we use the definition from SLICC 2012 (11). If the anti-dsDNA assay method or reference ranges were unspecified, we interpreted the results according to ELISA criteria using reference ranges from our laboratory as shown in Table 9.

#### 21.4. Examples present of SLEDAI-2K score

| How to determine. | Examples |
|---|---|
| Anti-dsDNA assay method is immunofluorensence. 1:40 is more than 1:10, criteria fulfilled. | 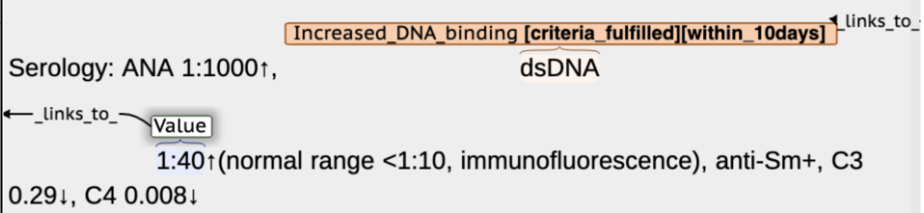 |
| Assume ELISA method. 509 is twice more than the upper reference limit of 100, criteria fulfilled. | 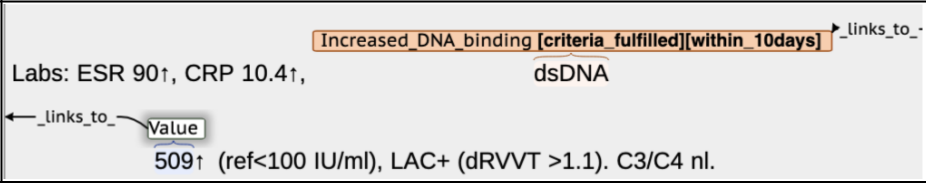 |

#### 21.5. Examples absent of SLEDAI-2K score

| Why is it an exception? | Examples |
|---|---|
| Assume ELISA method. "+" is not necessaryly twice more than the upper reference limit, not fulfilled. | 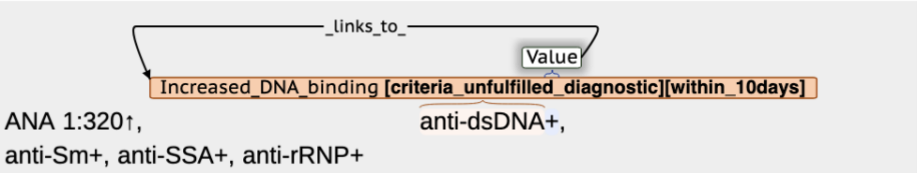 |

| | |
|---|---|
| Assume ELISA method. before 21/1/2013, the laboratory reference range is >67.5 IU/ml. 28 is less than 150, criteria unfulfilled. | 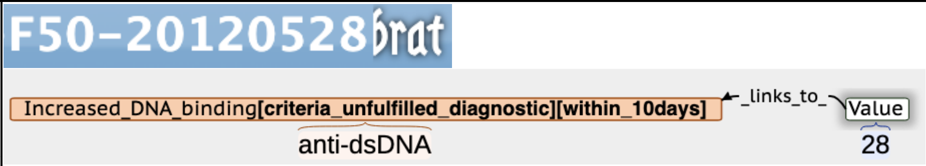  |

### 22. Fever

#### 22.1. Clinical definition of concepts

The Society of Critical Care Medicine and the Infectious Disease Society of America (IDSA) (80) and WHO (80) define rectal temperature of ⩾ 38 ° C (100.4 ° F) or axillary temperatures of ⩾ 37.5 ° C (99.5 ° F) as indicative of ***fever*** in both adults and children.

Many studies showed that disease activity was the most common cause of fever in SLE patients (81, 82). In active lupus without infection, the peak temperature ranges from 38 °C to 40.6 °C in an intermittent pattern. Clinically, differentiating the cause of fever is challenging, whether from disease activity, infection, malignancy, or drug reactions. Therefore, Rovin et al. established specific criteria for attributing fever to SLE: 1) absence of infection despite extensive testing, 2) presence of an illness typical of active SLE accompanying the fever, and 3) no evidence for infection despite escalation of immunosuppression. Clinical clues suggesting infection over a disease flare include, but are not limited to: 1) high white blood cell (WBC), 2) receiving moderate or high doses of glucocorticoids, 3) a poor response to nonsteroidal anti-inflammatory drugs (NSAIDs), acetaminophen, and low to moderate doses of glucocorticoids, and 4) signs and symptoms of commonly seen respiratory, urinary and soft tissue infections (83, 84). Key laboratory findings in lupus-associated fever include leukopenia (not explained by cytotoxic therapy), normal or slightly elevated CRP, low C3 and C4, and elevated anti-dsDNA, ESR/CRP ratio >15.(84) In contrast, infection typically drives a significant CRP increase and an ESR-to-CRP ratio <15. Further localization of infection may require chest imaging, urinalysis, or stool studies.

#### 22.2. Common ways to describe this concept

In medical terminology, "***pyrexia***" is the formal term for fever, and "***febrile***" is the adjective describing a state of having a fever.

| Normal and febrile body temperature ranges (rectal temperatures).(85) | | |
|---|---|---|
| Body temperature | °C | °F |
| Normal | 37–38 | 98.6–100.4 |
| Mild/low grade fever | 38.1–39 | 100.5–102.2 |
| Moderate grade fever | 39.1–40 | 102.2–104.0 |
| High grade fever | 40.1–41.1 | 104.1–106.0 |
| Hyperpyrexia | >41.1 | >106.0 |

#### 22.3. SLEDAI-2K definition in this study

As shown in Table 8, we applied the original SLEDAI-2K criteria: “>38°C. Exclude infectious cause”.

#### 22.4. Examples present of SLEDAI-2K score

| How to determine. | Examples |
|---|---|

<table>
<tr><td>1) fever no improvement despite antibiotics and anti-TB treatment. absence of infection despite extensive testing<br>2) presence of an illness typical of active SLE accompanying the fever</td><td>Fever [criteria_fulfilled][within_10days][special_entity_symptoms_signs_occurred]<br>BP 170/70, HR 116, Temp<br>39.2°C<br>Complaint & HPI/PMH:<br>Acute confusion (GCS 11), functional decline (requiring full care), fever, weight loss<br>Prior Tx response:<br>Treatment_response<br>IV ceftriaxone (for UTI/Enterococcus): No improvement in fever/confusion<br>Treatment_response<br>after 5d. Empiric anti-TB (for presumed meningitis): Persistent fever/confusion; later ruled<br>Exclusions<br>out (neg cultures/smears).<br>PMH: HTN (amlodipine 7.5mg daily). No dementia/psych history.<br><br>PE:<br>Neuro: Confused, disoriented, mild diffuse weakness (power 4+ limbs), no focal deficits/meningism.<br>photosensitive purpuric rash (forearms).<br>No ulcer, joint effusions, or serositis.<br><br>Labs: Hb 7.6↓, WBC 3.74↓, Plt 74↓; CRP 142↑, ESR 106↑; Cr 165↑, urea 8.9↑; UP 1.62g; ANA >1:800 (hom), anti-dsDNA 334↑ (nl<25), C3 0.39↓, C4 0.06↓.<br>TSPOT -ve AFB culture -ve<br>Imaging: CT brain: Chronic microvascular ischemia<br>CT T/A/P: Lymphadenopathy (biopsy neg for malignancy/TB).<br>LP: Protein 1.63↑ (nl 0.1-0.4), WBC 1/μL, no AFB/malignant cells.<br><br>Impression:<br>NPSLE LN</td></tr>
</table>

#### 22.5. Examples absent of SLEDAI-2K score

<table>
<tr><th>Why is it an exception?</th><th>Examples</th></tr>
<tr><td>Cannot exclude infection</td><td>Fever[criteria_unfulfilled_diagnostic][within_10days][special_entity_symptoms_signs_occurred]<br>Temp<br>38.8<br>HS dual no murmur, tachycardia ~ 100bpm, regular<br>Chest clear<br>Abd soft non-tender, no loin tenderness<br>Plan:<br>- admission for work-up for recurrent fever<br>Exclusions<br>? infection vs disease flare<br>Management Plan<br>Admission</td></tr>
<tr><td>Temp lower than 38 °C</td><td>BP 138/85, HR 106,<br>Fever[criteria_unfulfilled_diagnostic][within_10days][special_entity_symptoms_signs_occurred]<br>Temp<br>98.6°F</td></tr>
</table>

| | |
|---|---|
| Cannot exclude upper respiratory infection | 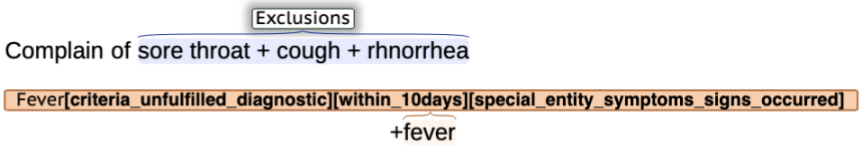  |

### 23. Thrombocytopenia

#### 23.1. Clinical definition of concepts

Generally, ***thrombocytopenia*** is defined as a platelet count below 150 × $10^3$ per μL (150 × $10^9$ per L) (86). Thrombocytopenia arises from three primary mechanisms: decreased production (e.g., bone marrow suppression from chemo/alcohol, nutritional deficiencies, infections like HIV/hepatitis); increased destruction/consumption (e.g., immune-mediated ITP/drug reactions, thrombotic microangiopathies, DIC, sepsis); or sequestration (e.g., hypersplenism in liver disease, gestational thrombocytopenia in pregnancy). (86)

#### 23.2. Common ways to describe this concept

Table. Common etiologies of thrombocytopenia in table below (86).

| **Term** | **Category** | **Definition** | **Key Clinical Features** |
|---|---|---|---|
| Immune Thrombocytopenic Purpura (ITP) | Immune-Mediated destruction | Autoimmune destruction of platelets | Isolated thrombocytopenia; petechiae, mucosal bleeding |
| Heparin-Induced Thrombocytopenia (HIT) | Immune-Mediated destruction | Immune reaction to heparin-platelet factor 4 complexes | Thrombosis despite thrombocytopenia; 50% platelet drop post-heparin |
| Drug-Induced Thrombocytopenia | Immune-Mediated destruction | Antibody-mediated platelet destruction triggered by drugs | Acute drop in platelets 5-14 days after drug exposure |
| Thrombotic Thrombocytopenic Purpura (TTP) | Microangiopathic | ADAMTS13 deficiency casued microthrombi | Pentad: Thrombocytopenia, MAHA, fever, renal/neuro dysfunction |
| Hemolytic Uremic Syndrome (HUS) | Microangiopathic | Shiga toxin-induced endothelial damage | Triad: MAHA, thrombocytopenia, acute renal failure |
| Disseminated Intravascular Coagulation (DIC) | Consumption | Systemic activation of coagulation cascade | Bleeding/thrombosis, low fibrinogen, elevated D-dimer |
| Gestational Thrombocytopenia | Pregnancy-Related | Mild thrombocytopenia in pregnancy | Asymptomatic; resolves postpartum; no fetal risk |
| HELLP Syndrome | Pregnancy-Related | Severe preeclampsia variant | Hemolysis, Elevated LFTs, Low Platelets with hypertension |
| Hypersplenism | Sequestration | Splenic platelet pooling | Associated with liver disease/portal hypertension |
| Myelodysplastic Syndrome (MDS) | Production Defect | Clonal hematopoietic disorder | Cytopenias, dysplastic bone marrow, risk of AML transformation |

| | | | |
|---|---|---|---|
| Pseudothrombocytopenia | Artifact | EDTA-induced platelet clumping in vitro | No clinical bleeding; resolves with alternative anticoagulants |

#### 23.3. SLEDAI-2K definition in this study

The SLEDAI-2K defines thrombocytopenia using a lower platelet threshold than the general clinical concept. As shown in Table 9, we applied the original SLEDAI-2K definition of thrombocytopenia, which is consistent with the SLICC 2012 criteria. Exclusions to consider were derived from the work of Levine et al. (87)

#### 23.4. Examples present of SLEDAI-2K score

| How to determine. | Examples |
|---|---|
| | Immune markers stable<br>Anti-dsDNA 168.4 (rising trend)<br>Thrombocytopenia[criteria_fulfilled][within_10days] ←links_to– Value<br>PLT 67 (decreasing trend)<br>No skin rash / mucositis / arthritis<br>No haematuria<br>No oedema / frothy urine<br>No bruising / PRB / tarry stool / headache<br>D/w Prof ROWLEY, to step up prednisolone and azathioprine<br>Management Plan<br>FU 7/52<br>Intention_to_treat<br>Step up prednisolone and azathioprine<br>Blood tests and UPC before next visit |

#### 23.5. Examples absent of SLEDAI-2K score

| Why is it an exception? | Examples |
|---|---|
| $10^3$/ul is equal to $10^9$/L, 138>100, criteria unfulfilled | Labs: WBC 3.62↓×10³/μL, Hb 10.9↓ g/dL,<br>Thrombocytopenia[criteria_unfulfilled_diagnostic][within_10days] ←links_to– Value; Unit –is_unit_of→ Value<br>Plt 138↓×10³/μL,<br>ESR 88↑ mm/hr, UPCR 8.86↑ (nephrotic), ANA 1:2560↑ (nuclear), anti-dsDNA (+), C3 36↓ (65-190), C4 9↓ (14-40). aPL panel (-). |

### 24. Leukopenia

#### 24.1. Clinical definition of concepts

***Leukopenia***, defined as a reduction in the number of white blood cells (WBCs) in the blood, is a medical condition that can arise from various etiological factors, including medication, systemic diseases, and hematological disorders. However, the threshold to define leukopenia varied widely.

#### 24.2. SLEDAI-2K definition in this study

As shown in Table 9, our study utilized the original SLEDAI-2K cutoff for leukopenia alongside the SLICC 2012 exclusions (11). Recognizing potential ambiguity in the original unit expression ("< 3,000 white blood cells/×$10^9$/L"), we defined the threshold as < 3,000 cells/mm³ (< 3.0 × $10^9$/L) to ensure clarity of unit.

#### 24.3. Examples present of SLEDAI-2K score

| How to determine. | Examples |
|---|---|
| By default the unit for WBC is $10^9$/L. | Time [within_10days] —is_time_of— Leukopenia [criteria_fulfilled][within_10days] ←_links_to_—<br>Labs (Jan 18): WBC<br>←_links_to_ Value<br>2.5↓, Hb 10.6↓; ESR 74↑; C3 61↓; ANA 1:320 (nucleolar);<br>anti-dsDNA 7↑; anti-ribosomal P Ab >480↑ (NL<37)<br>EEG (Jan 19): Diffuse slowing (3-7 Hz theta) → CNS involvement<br>SPECT: Left temporal/occipital hypoperfusion → NPSLE<br>MRI/CSF: Normal<br><br>Impression<br>NPSLE flare: Delirium (DRS 26/32) with EEG/SPECT abnormalities<br>Leukopenia[criteria_fulfilled][within_10days]<br>Leukopenia |

#### 24.4. Examples absent of SLEDAI-2K score

| Why is it an exception? | Examples |
|---|---|
| " ↑ " indicates more than upper reference limit | Leukopenia [criteria_unfulfilled_diagnostic][within_10days] ←_links_to_ Value<br>Hb 8↓, WBC 29.1k↑,<br>PMNs 96%↑; Na⁺/K⁺/Urea/Cr ↑ (trending up) |

*Table 10: Comparative Diagnostic Criteria: SLEDAI-2K definitions alongside supplementary criteria from the ACR, EULAR, and SLICC. Supplementary definitions employed in this study are highlighted.*

| Descriptor | SLEDAI-2K Definition | Definition from ACR 1997 (7, 8) | Definition from 2019 EULAR/ACR Classification Criteria(10) | Definition from SLICC 2012(11) | ACR nomenclature and case definitions for NPSLE in 1999 (9) |
|---|---|---|---|---|---|
| **Seizure** | Recent onset, excluding metabolic, infectious, or drug causes | Seizures-in the absence of offending drugs or known metabolic derangements (e.g., uremia, acidosis, or electrolyte imbalance) | Primary generalized seizure or partial/focal seizure. | / | Criteria:<br>A.Independent description of seizure by a reliable witness<br>B.EEG abnormalities<br>Exclude Seizure-like signs or symptoms OR of other causes. |
| **Psychosis** | Altered ability to function in normal activity due to severe disturbance in the perception of reality. Include hallucinations, incoherence, marked loose associations, impoverished thought content, marked illogical thinking, bizarre, disorganized, or catatonic behavior. Exclude uremia and drug causes. | Psychosis in the absence of offending drugs or known metabolic derangements (e.g., uremia, acidosis, or electrolyte imbalance) | Characterized by (1) delusions and/or hallucinations without insight and (2) absence of delirium. | / | Adopted the terminology of the Diagnostic and Statistical Manual of Mental Disorders (DSM).<br>Lupus psychosis belongs to 'psychosis due to a general medical condition' (DSM-IV and V 293.81/82).<br>Exclude merely reactive psychological disturbances. |
| **Organic brain syndrome** | Altered mental function with impaired orientation, memory, or other intellectual function, with rapid onset, and fluctuating clinical features.<br>Inability to sustain attention to the environment, plus at least two of the following: perceptual disturbance, incoherent speech, insomnia or daytime drowsiness, or increased or decreased psychomotor activity. Exclude metabolic, infectious, or drug causes. | / | / | Acute confusional state in the absence of other causes, including toxic-metabolic, uremia, drugs. | It is equivalent to the term 'delirium' as used in the DSM, or called 'acute confusional state'. |
| **Visual disturbance** | Retinal changes of SLE. Include cytoid bodies, retinal hemorrhages, serous exudate or hemorrhages in the choroid, or optic | / | / | / | / |

| | | | | | |
|---|---|---|---|---|---|
| | neuritis. Exclude hypertension, infection, or drug causes | | | | |
| **Cranial nerve disorder** | New onset of sensory or motor neuropathy involving the cranial nerves | / | / | Cranial neuropathy in the absence of other known causes such as primary vasculitis, infection and diabetes mellitus | Disorder of sensory and/or motor function of a specific cranial nerve(s). Exclude skull fracture, tumor, infection, and Miller-Fischer syndrome. |
| **Lupus headache** | Severe, persistent headache; may be migrainous, but must be nonresponsive to narcotic analgesia | | | | Types of headache including migraine or tension headache (episodic tension-type headache)<br>OR Cluster headache OR Headache from intracranial hypertension (Pseudotumor cerebri, benign intracranial hypertension) |
| **CVA** | New onset of cerebrovascular accident(s). Exclude arteriosclerosis. | | | | Cerebrovascular Disease:<br>1)Stroke syndrome<br>2)Transient ischemic attack (TIA)<br>3) Chronic multifocal disease<br>4)Subarachnoid and intracranial hemorrhage<br>5)Sinus thrombosis |
| **Vasculitis** | Ulceration, gangrene, lender finger nodules, periungual infarction, splinter hemorrhages, or biopsy or angiogram proof of vasculitis | | | | |
| **Arthritis** | ≥2 joints with pain and signs of inflammation (e.g., tenderness, swelling, or effusion) | Nonerosive arthritis involving 2 or more peripheral joints, characterized by tenderness, swelling, or effusion | Joint involvement: EITHER (1) synovitis involving 2 or more joints characterized by swelling or effusion<br>OR (2) tenderness in 2 or more joints and at | Synovitis involving two or more joints, characterized by swelling, effusion OR tenderness in two or more joints, and 30 minutes or | |

| | | | | |
|---|---|---|---|---|
| | | | least 30 minutes of stiffness in the morning. | more of morning stiffness |
| **Myositis** | Proximal muscle aching/weakness, associated with elevated creatine phosphokinase/aldolase or electromyogram changes or a biopsy showing myositis | / | / | / |
| **Urinary casts** | Heme-granular or red blood cell casts. | Cellular casts-may be red blood cell, hemoglobin, granular, tubular, or mixed | / | / |
| **Hematuria** | >5 red blood cells/high-power field. Exclude stone, infection or other cause. | / | / | / |
| **Proteinuria** | >0.5 gram/24 hours. | Persistent proteinuria>0.5 g/day, greater than 3+ if quantitation is not performed | Proteinuria >0.5g/24h by 24-hour urine or equivalent spot urine protein-to-creatinine ratio. | Urine protein/creatinine (or 24-hour urine protein) representing 500 mg of protein/24 hour |
| **Pyuria** | >5 white blood cells/high-power field. Exclude infection. | / | / | / |
| **Rash** | Inflammatory-type rash | 1)Malar rash: Fixed erythema, flat or raised, over the malar eminences, tending to spare the nasolabial folds<br>2)Discoid rash: Erythematous raised patches with adherent keratotic scaling and follicular plugging; atrophic scarring occurs in older lesions | 1)Subacute cutaneous or discoid lupus: Subacute cutaneous lupus erythematosus observed by a clinician*: Annular or papulosquamous (psoriasiform) cutaneous eruption, usually photodistributed. Discoid lupus erythematosus observed by a clinician*: Erythematous-violaceous cutaneous lesions with | 1) Chronic cutaneous lupus: Including classical discoid rash; localized (above the neck); generalized (above and below the neck); hypertrophic (verrucous) lupus; lupus panniculitis (profundus); mucosal lupus; lupus erythematosus tumidus; chillblains lupus; discoid lupus/lichen planus overlap |

| | | | | |
|---|---|---|---|---|
| | | 3)Photosensitivity: Skin rash as a result of unusual reaction to sunlight, by patient history or physician observation | secondary changes of atrophic scarring, dyspigmentation, often follicular hyperkeratosis/ plugging (scalp), leading to scarring alopecia on the scalp. If skin biopsy is performed, typical changes must be present.<br>2) Acute cutaneous lupus:<br>Malar rash or generalized maculopapular rash observed by a clinician*. If skin biopsy is performed, typical changes must be present | 2) Acute cutaneous lupus: Including lupus malar rash (do not count if malar discoid); bullous lupus; toxic epidermal necrolysis variant of SLE; maculopapular lupus rash; photosensitive lupus rash in the absence of dermatomyositis; or 3) subacute cutaneous lupus (nonindurated psoriaform and/or annular polycyclic lesions that resolve without scarring, although occasionally with postinflammatory depigmentation or telangiectasia |
| **Alopecia** | Abnormal, patchy, or diffuse hair loss | / | Non-scarring alopecia observed by a clinician*. | Nonscarring alopecia: Diffuse thinning or hair fragility with visible broken hairs in the absence of other causes such as alopecia areata, drugs, iron deficiency and androgenic alopecia |
| **Mucosal ulcers** | Oral or nasal ulceration | Oral or nasopharyngeal ulceration, usually | Oral ulcers observed by a clinician* | Oral ulcers: Palate, buccal, tongue or nasal ulcers in the absence of other |

| | | | | |
|---|---|---|---|---|
| | | painless, observed by a physician | | causes, such as vasculitis, Behcet' s, infection (herpes), inflammatory bowel disease, reactive arthritis and acidic foods |
| **Pleurisy** | Pleuritic chest pain with pleural rub or effusion, or pleural thickening. | Pleuritis- convincing history of pleuritic pain or rub heard by a physician or evidence of pleural effusion | Imaging evidence (such as ultrasound, x-ray, CT scan, MRI) of pleural effusion. | Typical pleurisy for more than 1 day OR pleural effusions OR pleural rub in the absence of other causes, such as infection, uremia. |
| **Pericarditis** | Pericardial pain with at least one of the following: rub, effusion, or electrocardiogram or echocardiogram confirmation. | Pericarditis- documented by ECG or rub or evidence of pericardial effusion | Imaging evidence (such as ultrasound, x-ray, CT scan, MRI) of pericardial effusion, or both.<br><br>For acute pericarditis: ≥2 of (1) pericardial chest pain (typically sharp, worse with inspiration, improved by leaning forward), (2) pericardial rub, (3) EKG with new widespread ST-elevation or PR depression, (4) new or worsened pericardial effusion on imaging (such as ultrasound, x-ray, CT scan, MRI) | Typical pericardial pain (pain with recumbency improved by sitting forward) for more than 1 day OR pericardial effusion OR pericardial rub OR pericarditis by ECG in the absence of other causes, such as infection, uremia and Dressler's pericarditis |
| **Low complement** | Decrease in CH50, C3, or C4 below the lower limit of normal for testing laboratory | / | C3 OR C4 below the lower limit of normal. | Low C3, low C4, low CH50 |

| | | | | |
|---|---|---|---|---|
| **Increased DNA binding** | Increased DNA binding by Farr assay above normal range for testing laboratory. | Anti-DNA-antibody to native DNA in abnormal titer. | Anti-dsDNA antibodies in an immunoassay with demonstrated ≥ 90% specificity for SLE against relevant disease controls | Above laboratory reference range, except ELISA: twice above laboratory reference range |
| **Fever** | >38°C. Exclude infectious cause | / | Temperature >38.3° Celsius. | / |
| **Thrombo-cytopenia** | <100,000 platelets/×$10^9$/L. Exclude drug causes | <100,000 mm3 in the absence of offending drugs | Platelet count <100,000/mm³ | Thrombocytopenia (<100,000/mm3) at least once: in the absence of other known causes such as drugs, portal hypertension, and thrombotic thrombocytopenic purpura |
| **Leukopenia** | <3,000 white blood cells/×$10^9$/L. Exclude drug causes | Leukopenia-<4,000 mm3; Lymphopenia-<1,500 mm3 | White blood cell count <4,000/mm³. | Leukopenia < 4,000/mm3 at least once (in the absence of other known causes such as Felty's, drugs and portal hypertension) ; Lymphopenia (< 1,000/mm3 at least once) in the absence of other known causes such as corticosteroids, drugs and infection |
